# Psychometric analysis of the Evaluation of Multidimensional Functioning and Risks in Aging (EMFRA) scale: reliability, structural validity, and concurrent validity

**DOI:** 10.64898/2026.11.11.26367642

**Authors:** José Fierro-Marrero, Adrián Gallego-García, Luís Cabrera-Durán, Sergio Martínez-Benito, Alba Paris-Alemany, Roy La Touche

**Author notes:** Corresponding author; Alba Paris-Alemany. José Fierro-Marrero, Adrián Gallego-García Luis Cabrera-Durán Sergio Martínez-Benito, Roy La Touche.

## Abstract

**Background:** EMFRA is a brief scale developed to assess physical, cognitive, emotional, and social functioning in older adults. This study examined its initial psychometric properties.

**Methods:** A cross-sectional study was conducted with 201 older adults aged ≥65 years without a diagnosis of dementia, recruited from the community, day-care and nursing centres from Madrid. Dimensional structure was examined through bootstrap parallel analysis (PA), bootstrap exploratory graph analysis (EGA), and factor analysis (FA), followed by progressive item refinement. Internal consistency, test–retest reliability in a subsample (n = 22), SEM, MDC95, floor and ceiling effects, and associations with SPPB, MoCA, Barthel Index, and HADS were assessed.

**Results:** The 21-item version did not reproduce the four hypothesized domains, yielding a three-dimensional solution with limited stability. A nine-item version comprised two factors, Autonomy and Psychological Distress, although FA fit was limited (TLI = 0.80; RMSEA = 0.13). A low hierarchical omega (0.409) did not support a total score, whereas factor reliability was adequate (ω = 0.840 and 0.785). Test–retest ICCs were 0.82 and 0.78, with wide 95% CIs. No floor or ceiling effects were observed. Autonomy correlated with SPPB (r = 0.751), MoCA (r = 0.697), and Barthel (r = 0.56), and Psychological Distress with HADS (r = 0.625).

**Conclusions:** The nine-item EMFRA shows preliminary evidence of a two-factor structure whose factors should be scored separately. Its temporal reliability and measurement error remain uncertain, and external validation is required before clinical implementation.

## 1. Introduction

Functioning in older adults cannot be adequately characterized by chronological age or the presence of individual health conditions. People of similar ages may differ substantially in their physical and mental capacities and in their ability to perform everyday activities. The World Health Organization’s framework for healthy aging defines functional ability through the interaction between intrinsic capacity and the environment in which a person lives (Beard et al., 2016). Intrinsic capacity encompasses the physical and mental capacities of an individual, with locomotion, vitality, cognition, psychological capacity, and sensory function proposed as its principal domains (Cesari et al., 2018). Functional ability has a broader meaning because it also reflects the environmental conditions that enable or restrict the expression of these capacities. The distinction is relevant to measurement: scores describing impairments in specific capacities cannot be assumed to represent an individual’s functioning across different everyday contexts.

The operationalization of these constructs remains inconsistent. Schwab et al. (2025) documented substantial variation in the definitions, dimensions, and assessment approaches used to examine functional capacity and functional ability in older populations. Such differences complicate comparisons between studies and raise questions about the meaning of summary scores obtained from heterogeneous indicators. Chang et al. (2024) developed a functional ability index incorporating intrinsic capacity and environmental characteristics, illustrating how the World Health Organization’s conceptual framework can inform the selection and organization of measurement indicators. Evidence from the ICOPE-Care program further indicates that declines in intrinsic capacity occur in different combinations rather than following a uniform pattern across older adults (De Souto Barreto et al., 2024). These findings support the clinical relevance of identifying specific areas of difficulty, although the dimensions represented by an assessment instrument and the interpretation of its scores require separate justification.

The International Classification of Functioning, Disability and Health (ICF) provides a framework for examining functioning through body functions and structures, activities, participation, and contextual factors (World Health Organization, 2001). Its conceptual scope extends beyond the identification of impairments and recognizes that functioning is influenced by the circumstances in which activities are performed. The Integrated Care for Older People (ICOPE) guidance applies a related concern to clinical assessment by recommending the identification of declines in intrinsic capacity and the evaluation of social and support needs to inform subsequent care planning (World Health Organization, 2024). These frameworks provide a rationale for multidimensional assessment, but they do not prescribe a single measurement structure. The selection of domains, the relationships expected among their indicators, and the interpretation of the resulting scores remain decisions that must be justified for each assessment instrument.

The Evaluation of Multidimensional Functioning and Risks in Aging (EMFRA) scale was developed as a brief assessment of functioning and potential health-related risks in older adults. Its original conceptual framework comprised four domains: physical function, cognitive function, emotional status, and social situation. Item development was informed by the ICF and by evidence concerning factors associated with adverse health outcomes in older populations. An initial pool of 24 items underwent evaluation by 15 experts, followed by cognitive interviews with 10 clinicians and 10 older adults. The resulting 21-item version incorporated modifications intended to improve the relevance and comprehensibility of its content (Fierro-Marrero, Reina-Varona, Pardo-Montero, et al., 2025). This development process provided preliminary evidence of content validity for the proposed assessment framework. The empirical relationships among the items, the dimensionality of their responses, and the reliability of the resulting scores had not yet been examined.

The distinction between content validity and other sources of measurement evidence is particularly relevant to EMFRA. Content validity concerns the extent to which the content of an instrument is relevant, comprehensive, and comprehensible for its intended construct, population, and purpose (Mokkink et al., 2025; Terwee et al., 2018). The internal structure of item responses addresses a different question: whether the observed associations among indicators support the proposed organization of the scores. These properties must be considered in relation to the instrument’s conceptual development. As discussed by Boateng et al. (2018), decisions concerning item selection and dimensionality can alter the content represented by a scale. This issue deserves attention when an instrument combines indicators of clinically distinct aspects of functioning. Limited correlations between items may reflect difficulties in representing a common latent dimension, but they may also arise because the indicators describe different attributes. Reducing the number of items on statistical grounds may improve the internal consistency of the retained scores while restricting their conceptual coverage. The adequacy of such decisions depends on the intended measurement model and the interpretation assigned to the resulting dimensions.

The evaluation of EMFRA scores therefore requires empirical examination of dimensionality without assuming that the four domains established during content development necessarily correspond to four latent factors. Exploratory factor analysis (FA) and exploratory graph analysis (EGA) can provide complementary information about the organization of item responses, although their findings must be interpreted in relation to the conceptual framework and the characteristics of the data (H. F. Golino & Epskamp, 2017). Reliability estimates address the consistency or stability of scores under specified measurement conditions. Their interpretation depends on the population, the score structure, and the measurement procedure. Evidence concerning the relationships between EMFRA scores and established clinical measures can further inform their interpretation, provided that the expected associations are grounded in the constructs represented by the respective measures. These sources of evidence address distinct measurement questions and should not be treated as interchangeable (Prinsen et al., 2018).

The present study aimed to examine the psychometric properties of scores derived from the 21-item EMFRA scale in older adults. The internal structure of item responses was investigated using complementary exploratory approaches, and internal consistency and test–retest reliability were examined for the resulting scores. Score distributions, including floor and ceiling effects, were also assessed. Relationships with established measures of physical performance, cognitive functioning, emotional symptoms, and functional independence were investigated to obtain evidence relevant to score interpretation. The analyses also examined the correspondence between the empirical dimensional structure and the conceptual framework established during the development of EMFRA.

## 2. Methods

### 2.1. Sample

Participants were recruited from community and healthcare settings in the Community of Madrid, Spain. Recruitment was conducted across settings serving individuals with different health conditions, levels of functional independence, and care needs, with the aim of including a heterogeneous older adult population. Recruitment sites included Clínica Universitaria La Salle; five STIMA day-care centers located in Princesa, Embajadores, Aluche, Moratalaz, and Hortaleza; and the Valdeluz Arroyomolinos nursing home. A total of 204 older adults aged 65 years or older were recruited, of whom 201 were included in the analyses. The remaining three participants were excluded because they did not complete the assessment due to time constraints.

Eligibility was determined using two criteria: (1) age ≥65 years and (2) absence of a medically established diagnosis of dementia. No other eligibility restrictions were applied regarding pain, chronic conditions, mobility (even including subjects with wheelchair use), functional independence, or cognitive functioning. Individuals with cognitive difficulties were therefore eligible if they had no medical diagnosis of dementia.

This study followed the ethical principles of the Declaration of Helsinki and was approved by the Ethics Committee of the Hospital Clínico San Carlos, Madrid, Spain (n° 23/398-E), with all participants being informed about the aim and methods of the study and signing the informed consent.

### 2.2. Assessments

Following written informed consent, participants provided sociodemographic information through verbally administered questions covering age, sex, educational level, years of formal education, marital status, and employment status. Anthropometric information, including height, weight, and body mass index (BMI), was also collected. Clinical history was documented using open-ended questions addressing previous or current pain, surgical procedures, and the presence of musculoskeletal, neurological, psychological, metabolic, or other health conditions. The conditions reported by participants were subsequently coded and grouped into musculoskeletal, neurological, psychological, metabolic, and other categories.

Physical functioning tests were applied including the Short Physical Performance Battery (SPPB) (Guralnik et al., 1994), a global cognitive function measure with the Montreal Cognitive Assessment (MoCA) 8.1 Spanish version (Gómez-Moreno et al., 2022; Nasreddine, 2018; Nasreddine et al., 2005), depression and anxiety traits with the Spanish version of the Hospital Anxiety and Depression Scale (HADS) (Herrero et al., 2003; Herrmann, 1997), and independency in basic activities of daily living with the Spanish version of the Barthel Index (Cabañero-Martínez et al., 2009).

The Evaluation of Multidimensional Functioning and Risks in Aging (EMFRA) scale was also administered. The instrument combines self-reported information with cognitive and physical performance tasks, including assessments of handgrip strength and gait speed (Fierro-Marrero, Reina-Varona, Pardo-Montero, et al., 2025). The time required to complete the EMFRA assessment was recorded.

In addition, a second assessment of the EMFRA scale was provided after 2 to 10 days from the initial evaluation to explore test-retest reliability properties of the scale.

#### 2.2.1. Response distribution of EMFRA items

A descriptive analysis of response distributions of EMFRA items was reported to check on possibly skewed responses. Mean, SD, minimum, maximum values, quantiles of (0-2 point responses) would be reported, along with the skew and kurtosis. In addition, the frequency of response categories would be reported.

### 2.2. Item-level test-retest reliability

Item-level test–retest reliability was evaluated in participants who completed both EMFRA assessments. For each item, contingency tables were constructed to examine the distribution of responses across the two administrations. Agreement was quantified using Gwet’s weighted AC2 coefficient (Gwet, 2008a, 2008b), which is less sensitive than Cohen’s kappa to certain imbalances in response-category prevalence and marginal distributions (Gwet, 2008a).

Radical weighting matrices were applied to all items except Item 2 (gait speed), for which a custom weighting matrix was specified to assign greater penalties to disagreements involving the “unable to walk” category. Agreement coefficients were interpreted using the conventional classification proposed by Landis & Koch (1977), recognizing that these thresholds were originally developed for kappa statistics and were used here as descriptive guidelines.

Items exhibiting very low test–retest agreement were considered for potential exclusion during the subsequent psychometric evaluation. The weighting schemes, item-level contingency tables, and corresponding agreement estimates are reported in the Supplementary Material (SM).

### 2.3. Structural validity

#### 2.3.1. Correlation matrix, and matrix performance

Preliminary analyses examined the correlation structure of the EMFRA items using polychoric correlations, given the ordinal nature of the response categories and the assumption of underlying continuous variables. Under these conditions, polychoric correlations may provide more appropriate estimates of the associations between items than Pearson or Spearman correlations (Ekström, 2011a, 2011b; Holgado–Tello et al., 2010; Pearson & Pearson, 1922). Previous research has also examined their performance in dimensionality analyses involving ordinal variables (Ekström, 2011b; Markos & Tsigilis, 2024).

Pairwise 3 × 3 contingency tables were inspected to identify response-category combinations with zero frequencies. A continuity correction of 0.5 was applied when necessary. The resulting correlation matrix was examined for symmetry and positive semidefiniteness (PSD) through inspection of its eigenvalues (Zhang, 2011).

For descriptive purposes, the polychoric and anti-image matrices were reported both in full and filtered for low correlation values (-0.29 to 0.29) (Akoglu, 2018; Chan, 2003). Kaiser–Meyer– Olkin (KMO) and item-level measures of sampling adequacy (MSA) were reported. In line with Lorenzo-Seva & Ferrando (2021), lower KMO or MSA values were not considered as criteria for rejecting factorability, or elimination of items, as these indices may decrease with low zero-order correlations, low number of items, and with higher number of factors.

#### 2.3.2. Combined dimensional structure analysis

The dimensional structure of EMFRA was analysed using a two-way method employing a customized Horn’s Parallel Analysis (PA) employing bootstrap resamples, based on factors, and the bootstrap Exploratory Graph Analysis (EGA) (Finch & Monahan, 2008; H. F. Golino & Epskamp, 2017; Horn, 1965). Horn’s PA and EGA currently present the best, and similar performance among the different factorial methods for accurately detecting the number of factors in instruments with similar characteristics as EMFRA. The use of the best possible method based on the context of the research (sample size, expected number of items per factor, number of factors, correlation between factors, and type of correlation matrix, among other issues), is suggested in previous research (H. F. Golino & Epskamp, 2017). Based on this, different simulation studies indicate a good performance of the widely employed PA in multidimensional scales, as well as of EGA (H. Golino et al., 2020; H. F. Golino & Epskamp, 2017; Markos & Tsigilis, 2024). Overall, the performance of both methods is similar, and it has not yet been established in which specific context they clearly differ (H. Golino et al., 2020; H. F. Golino & Epskamp, 2017; Markos & Tsigilis, 2024). Additionally, both methods present excellent performance under simulations with asymmetrical response distributions while employing polychoric correlation matrices (Markos & Tsigilis, 2024). Authors such as Garcia-Garzon et al. (2019), suggest the possibility of employing both methods in combination. In addition, both methods would be conducted employing bootstrap resamples to consider the stability of the detected dimensions across other samples.

Both procedures incorporated bootstrap resampling to examine the stability of the estimated dimensional solutions across resamples of the observed data. Given the absence of an established decision framework for integrating the results of bootstrap PA and bootstrap EGA, dimensional retention was evaluated using four considerations: (1) convergence between the dimensional solutions identified by PA and EGA; (2) the performance of the corresponding factor-analytic solution, including global model fit and residual-based indices; (3) the stability of the EGA solution, assessed through the frequency with which the number of communities was recovered across bootstrap resamples and the structural consistency of those communities; and (4) the relative improvement of successive solutions in relation to the preceding considerations. These sources of evidence were interpreted jointly, rather than treating any single global fit index or conventional cutoff as a definitive criterion for dimensional retention.

#### 2.3.3. Bootstrap PA and factor analysis

A customized two-step bootstrap PA procedure was conducted, given the benefits associated with considering population-based parameters (Finch & Monahan, 2008). A null eigenvalue distribution was generated through 1,000 random permutations, and a bootstrap distribution of sample eigenvalues was independently generated through 1,000 bootstrap resamples of the observed data, both computing polychoric matrices and extracting factor-based eigenvalues via the Minimum Residual method. Factor retention required that the lower bound of the bootstrap eigenvalue distribution (P5) exceeded the 95th percentile of the permuted null distribution (P95). The number of retained factors subsequently guided a FA employing the Minimum Residual method with Oblimin rotation and 1000 iterations for parameter estimation.

Dimensional performance was evaluated using multiple indices, including the R^2^, chi-square statistic, TLI, RMSEA, and RMSR, following established cutoff criteria (Hu & Bentler, 1999). Model fit was determined by combining the chi-square test (p ≥ .05 indicating excellent fit) and TLI (≥ .95 indicating excellent fit), yielding either excellent or poor model fit. Error fit was determined by combining RMSEA and RMSR, with RMSEA values ≤ .05 indicating good, ≤ .06 acceptable, and > .06 poor residual fit, and RMSR values ≤ .05 indicating good, ≤ .06 acceptable, ≤ .07 adequate, ≤ .08 marginal, and > .08 poor residual fit. When both indicators within each category converged, a single classification was assigned; when they diverged, a composite range label was reported reflecting the worst-to-best classification across indicators (e.g., poor-to-acceptable residual fit).

Item-level performance was evaluated using communalities, factor loadings, and cross-loadings obtained from the oblimin-rotated factor solution. An item was considered a candidate for elimination if it met at least one of the following criteria: (1) low factor loadings, defined as absolute loadings below 0.30 on all factors; (2) substantial cross-loadings, defined as absolute loadings of at least 0.50 on two or more factors; or (3) low communality, defined as a value below 0.30.

#### 2.3.4. Model performance through Bootstrap and empirical EGA

The network structure in EGA was estimated using the graphical LASSO (glasso) model, which has shown superior performance compared with the alternative TMFG network estimation method (Christensen, Garrido, Guerra-Peña, et al., 2023; H. Golino et al., 2020). Community detection was performed using the walktrap algorithm for multidimensional solutions, while the Louvain algorithm was specified for cases where a unidimensional structure was detected (Christensen, Garrido, Guerra-Peña, et al., 2023; Christensen, 2024).

Bootstrap EGA with 1,000 resampling iterations was employed to evaluate the stability of the dimensional solution. Dimensional performance was assessed through the frequency distribution of the number of communities identified across bootstrap iterations, reporting the modal solution and its difference in frequency relative to the second, third and fourth most frequent solutions. Structural consistency of the empirical EGA solution was also assessed, defined as the proportion of iterations replicating the same community assignment as in the empirical solution.

Item performance was evaluated based on: 1) replicability of each item to its empirically assigned community across bootstrap iterations, with instability defined as a replicability rate ≤0.75; 2) item-level diagnostics identifying minor dimensions (MiD), two-item dimensions (TwD), and multidimensional behaviour (MuD) (Christensen, Garrido, Guerra-Peña, et al., 2023); 3) redundancy assessed through weighted topological overlap (wTO) using the MBR method, with a cut-off of ≥0.25, in addition, to the zero-order and partial correlation between both items (Christensen, Garrido, & Golino, 2023); 4) low community loadings across all communities, defined as an absolute loading <0.30; and 5) high cross-loadings across communities, defined when both primary and secondary absolute community loadings were ≥0.50, computed via the revised net loads method with Oblimin rotation.

#### 2.3.5. Model acceptability and item reduction criteria

The results of FA and EGA were considered jointly to evaluate dimensional solutions and guide item reduction across successive models. Model acceptability was assessed by integrating the four sources of evidence described previously. These indicators were considered collectively, without requiring every global fit index to satisfy a fixed cutoff for dimensional retention. When the available evidence indicated limitations in a given solution, item-level diagnostic indicators were examined to identify candidates for elimination and guide subsequent model refinement.

Item reduction followed a predefined hierarchy based on the identification of unfavourable diagnostic indicators across FA and EGA. Items identified as problematic by both methods were given the highest priority for potential elimination, followed by items identified exclusively through FA and, subsequently, those identified exclusively through EGA. Identification as a candidate did not necessarily imply elimination, as decisions were made within the broader evaluation of successive dimensional solutions.

A summary table provided in SM documented the candidate items identified at each stage of model refinement, their corresponding FA and EGA diagnostic indicators, and the decisions to retain or eliminate each item.

#### 2.3.6. Factor naming and score calculation

The final model fulfilling the dimensional criteria (with FA and EGA) was reported with the previously mentioned measures. Additionally, factor scores for the factors identified through PA and FA were calculated employing the ten Berge method for preserving the correlation structure between latent factors, and therefore preserving the obliquity (Estabrook & Neale, 2013; Ten Berge et al., 1999). Factor scores were then calculated as linear combinations of item responses and their corresponding weights. To ensure interpretability, items with negative regression weights were reverse-coded prior to score computation using the transformation 2, so that higher scores consistently reflected higher levels of the latent construct.

Based on the loadings, a name for each detected factor was proposed based on research team consensus and a literature search.

### 2.4. Factor-level internal consistency

Internal consistency was estimated using McDonald’s omega coefficient, which does not require the assumption of tau-equivalent item contributions (Shaw, 2021). Reliability was examined for the complete item set and for the subsets of items primarily associated with each retained factor.

For the complete item set, a Schmid–Leiman hierarchical decomposition was used to examine the contributions of general and group factors to score reliability. The reported indices included omega total (ωt), representing the proportion of composite-score variance attributable to the common factors collectively, and hierarchical omega (ωh), representing the proportion attributable specifically to the general factor. Additional omega estimates for the general and group factors were reported to characterize their respective contributions to reliable score variance. Explained common variance (ECV) was also calculated to quantify the proportion of common variance accounted for by the general factor and to inform the assessment of essential unidimensionality.

Subscale-level internal consistency was estimated separately for the subsets of items with their primary loadings on each retained factor. Items with negative scoring weights were reverse-coded for the item-subset calculations, as specified in the scoring procedure. Omega coefficients above 0.70 were interpreted as indicative of acceptable internal consistency, using this threshold as a conventional guideline rather than an absolute criterion (McNeish, 2018).

### 2.5. Factor-level test-retest reliability, standard error of measurement, and minimum detectable change

Test–retest reliability was estimated using the intraclass correlation coefficient (ICC 2,1) following the framework of Shrout and Fleiss (1979) and current reporting recommendations (Koo & Li, 2016). ICC was considered adequate when >0.75 (Fleiss, 1999). Standard error of measurement (SEM) was calculated employing formula 5.7 indicated by de Vet et al. (2011), and Minimal Detectable Change at the 95% confidence level (MDC95) was computed with SEM, the square root of the number of measurement time points (2), and the critical 97.5th percentile value of Student’s t distribution with the included sample size (Vet et al., 2011).

### 2.6. Factor-level floor-ceiling effects

Floor and ceiling effects would be analysed checking on whether more than a 15% of subjects reached the minimum and maximal score within the factor (McHorney & Tarlov, 1995; Vet et al., 2011).

Factor floor and ceiling effects were evaluated for each factor score under three independent, non-mutually exclusive criteria. First, exact floor and ceiling effects were considered when the observed factor score equalled exactly the minimum or maximum possible scale value. Second, floor and ceiling effects were assessed by examining whether subjects scored at the minimum (0) or maximum (2) on all items showing the highest loading on each factor. Third, factor scores falling within the lowest or highest 10% of the maximum possible scale value were also considered indicative of floor or ceiling effects. A floor or ceiling effect was considered present when more than 15% of subjects met any of these criteria (McHorney & Tarlov, 1995; Vet et al., 2011). For each criterion, the absolute and relative frequency of subjects meeting the condition was reported. Additionally, the corresponding raw score range and its equivalent percentage of the maximum possible scale score were indicated as a reference for interpreting the magnitude of each criterion.

### 2.7. Factor-level concurrent validity

Pairwise analyses between factor scores and relevant clinical measures would be analysed. These would include physical function with SPPB, global cognitive function with MoCA, anxiety and depression traits separately and combined with HADS, and independency in basic ADL with the Barthel index. The relationship with these predictors and the factor score would be analysed with an Ordinal Regression Model (ORM) employing a logistic distribution. ORM provides a semi-parametric framework that does not require the outcome distribution to be continuous and is robust to extreme values when compared to ordinary least squares method (Harrell, 2015). The relationship would be accounted with each clinical measure considering the Nagelkerke R², and Pearson’s linear correlation coefficient. In addition, regression coefficients (β), and bootstrap 95% percentile confidence intervals (2,000 resamples), and Spearman correlation coefficients would be reported.

All the mentioned analyses were conducted using R Software v.4.6.0 (R Core Team, 2026). R: A language and environment for statistical computing. R Foundation for Statistical Computing, Vienna, Austria. URL https://www.R-project.org/). Packages, syntax, and functions employed are provided in SM.

## 3. Results

### 3.1. Sample characteristics

A total of 204 older adults were recruited, of whom 201 completed the full assessment, with three participants excluded due to time constraints to complete the evaluation. The sample had a mean age of 77.2 ± 9.1 years (range: 65–99), and a mean BMI of 26.57 ± 4.71 (range: 15.81-43.56), indicating a tendency toward overweight, but with high variability. The sample was predominantly composed of women (66%), with most participants being married (41%), or widowed (40%), and retired (84%), see SM. A large proportion had completed university-level education (43%), with a mean of 16.49 ± 5.59 years of formal education (range: 0-31), see Table 1.

**Table 1.**
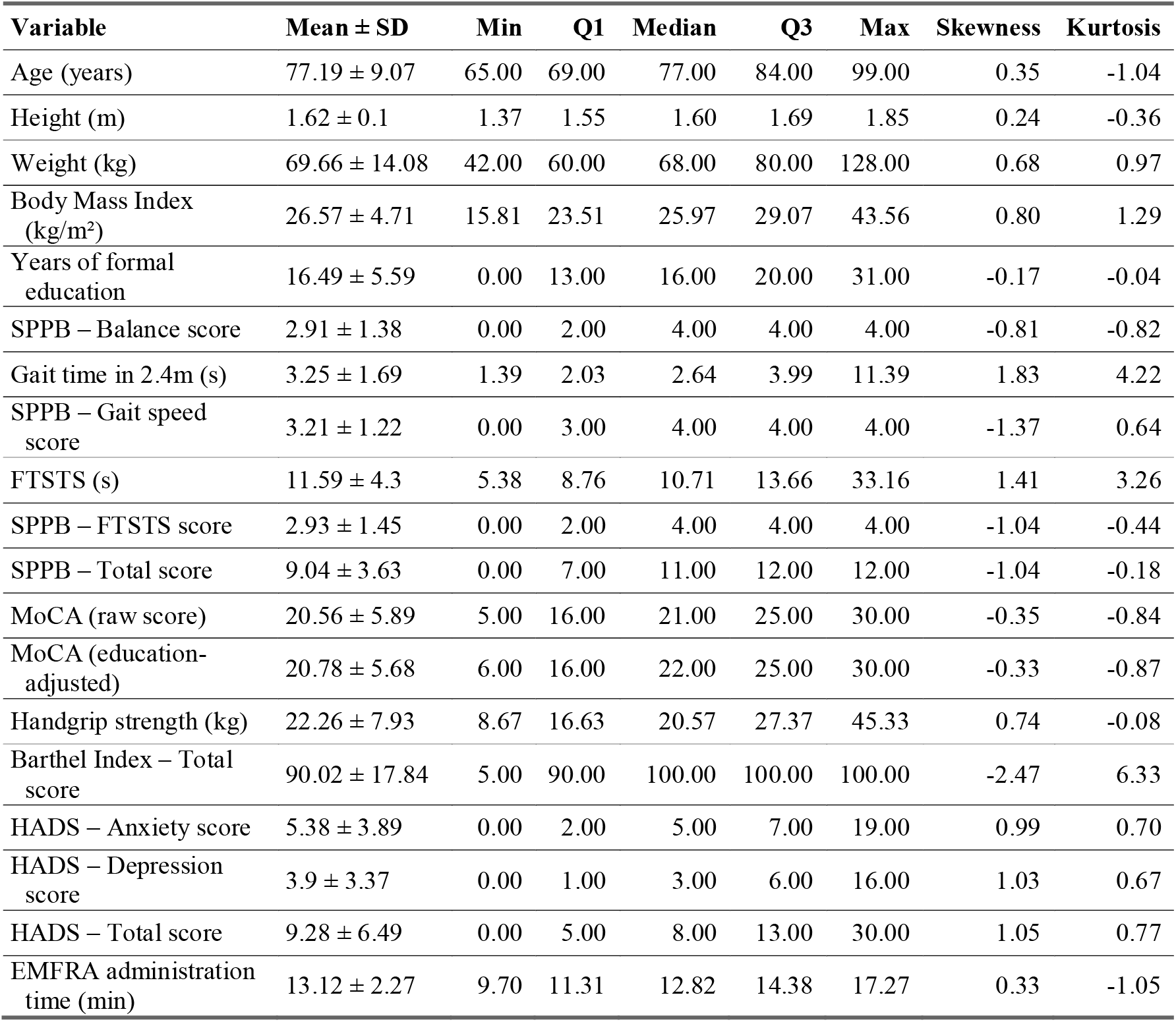
Descriptives.

The majority presented a past or current musculoskeletal condition (69%), with low back and neck the most common locations (23% each). Only a smaller proportion presented neurological (22%) or psychological conditions (8%), with stroke (7%), and depression (6%) being the most prevalent, respectively. Additionally, 52% of the sample had some form of metabolic condition, most commonly hypertension (31%). Other conditions such as cancer, gastrointestinal surgery, or cardiac disorders were relatively uncommon, see SM.

Overall, the sample showed heterogeneous characteristics, with a general tendency toward good physical function (SPPB: 9.04 ± 3.63; range: 0-12), mild cognitive impairment (MoCA: 20.56 ± 5.89, 5-30 range) (Malek-Ahmadi & Nikkhahmanesh, 2024), low levels of anxiety and depression traits (HADS Total: 9.3 ± 6.5, 0-30 range), and almost complete independence in basic ADL (Barthel Index: 90.0 ± 17.8, 5-100 range). Administration time of EMFRA was analysed from 16 subjects, lasting 13.12 ± 2.27 minutes (11.31-17.27 range), see Table 1. Further information is provided in SM.

### 3.2. Item response distributions of EMFRA

All three response categories were represented across the sample for each of the 21 EMFRA items. Items 1 (Grip Strength) and 19 (Socioludic Activity) showed relatively balanced response distributions.

Higher frequencies of the lowest category were observed in Items 3 (Gait Assistance), 4 (Balance Loss), 9 (Concentration Difficulty), 11 (Loneliness), 14 (Self-efficacy Difficulty), 16 (Economic Difficulty), and 20 (Communication Difficulty). In contrast, the highest category was more frequent in Items 2 (Gait Speed), 6 (Money Arithmetic), 7 (Attention–Inhibition), 10 (Cognitive Activities), 15 (Life Satisfaction), 17 (Family Interaction), 18 (Social Interaction), and 21 (Help Availability). The middle category was most common in Items 5 (Fatigue), 8 (Memory Difficulty), 12 (Sadness), and 13 (Irritability).

Overall, although response patterns were heterogeneous, there was a general tendency toward more favorable (better) trait levels across items, see Figure 1, and SM.

**Figure 1.**
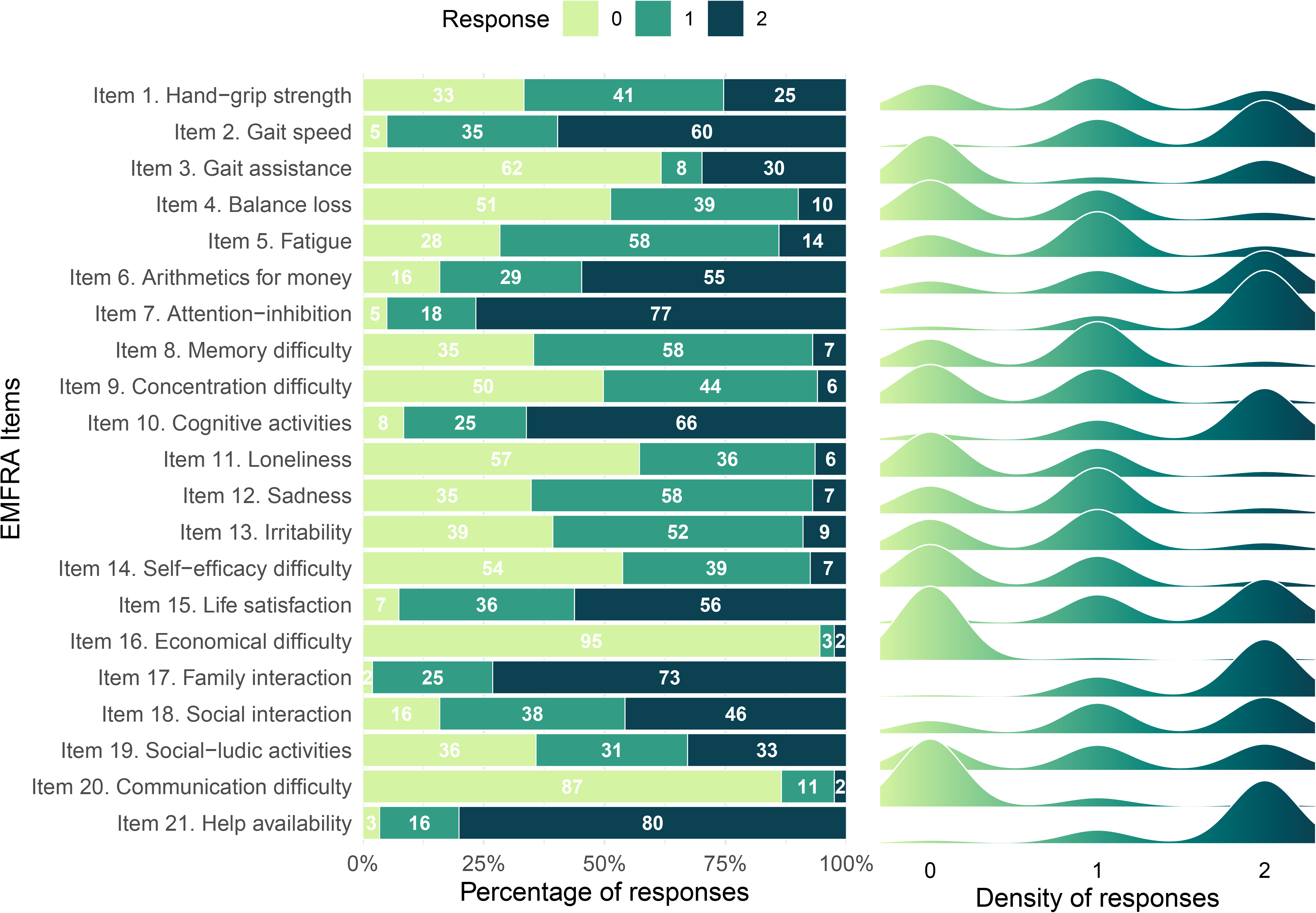
Item response distribution

### 3.3. Item-level test-retest reliability

Due to the low number of subjects analysed in test-retest measures (22 subjects), wide 95%CI prevented drawing clear conclusions about the real test-retest reliability value, and therefore their possible exclusion could not be clearly established.

Most items showed high levels of stability over time. Specifically, five items demonstrated “almost perfect” reliability (Items 2, 3, 4, 10, and 16), while eleven items showed “substantial” reliability (Items 1, 5, 6, 7, 12, 13, 14, 17, 19, 20, and 21). In addition, four items presented “moderate” reliability (Items 8, 9, 11, and 18), and one item showed “fair” reliability (Item 15 – Life satisfaction), see Figure 2. Despite its lower reliability, Item 15 was retained to evaluate its contribution within the final factorial structure and overall reliability of the factors.

**Figure 2.**
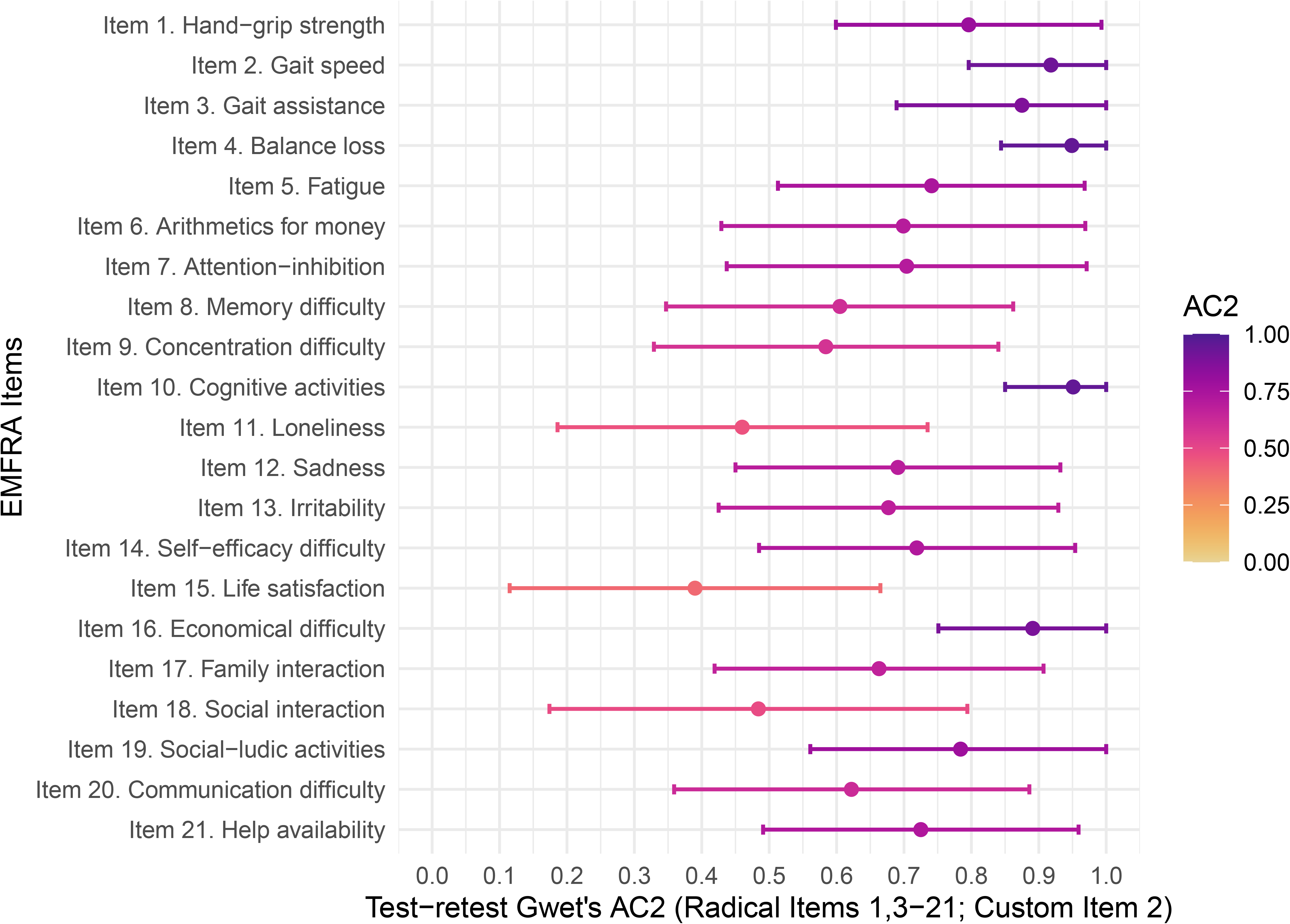
Item level reliability

### 3.3. Polychoric correlation matrix, KMO and MSA

Prior checks revealed a total of 81 pairwise response combinations of zero frequency. This yielded a polychoric matrix with symmetry but not PSD with one negative eigenvalue. PSD was achieved with the continuity correction of 0.5 applied to zero frequency response combinations and employed for the subsequent analyses. Polychoric matrix, and anti-image matrix are reported in SM.

The overall KMO yielded 0.43, with most items presenting low-to-moderate values, with Item 21 (Help availability) being the lowest MSA value (0.09). However, these did not prevent subsequent analyses (Lorenzo-Seva & Ferrando, 2021).

### 3.4. Dimensional analysis

The initial model, comprising all 21 EMFRA items, yielded a three-factor solution in bootstrap PA, while bootstrap EGA identified three communities as the most frequently recovered dimensional solution. However, the FA solution showed poor global fit (proportion of variance: MR1 = 0.199, MR2 = 0.138, and MR3 = 0.075; χ² = 992.86, p < 0.001; TLI = 0.45; RMSEA = 0.18; RMSR = 0.07). Bootstrap EGA also indicated limited dimensional stability. Three- and four-community solutions were recovered in 42% and 34% of bootstrap samples, respectively, while structural consistency was low across the three empirical dimensions (Dim 1: 30.8%; Dim 2: 19.7%; Dim 3: 15.1%).

Three successive reduced models (Reduced Models A–C) were examined through the progressive elimination of items identified as problematic according to the FA and EGA diagnostic criteria. The final retained solution, Reduced Model C, comprised nine items and showed convergence between bootstrap PA and bootstrap EGA, both supporting a two-dimensional structure. The bootstrap PA results and the distribution of dimensional solutions identified through bootstrap EGA for the final model are presented in Figures 3 and 4, respectively.

**Fig 3.**
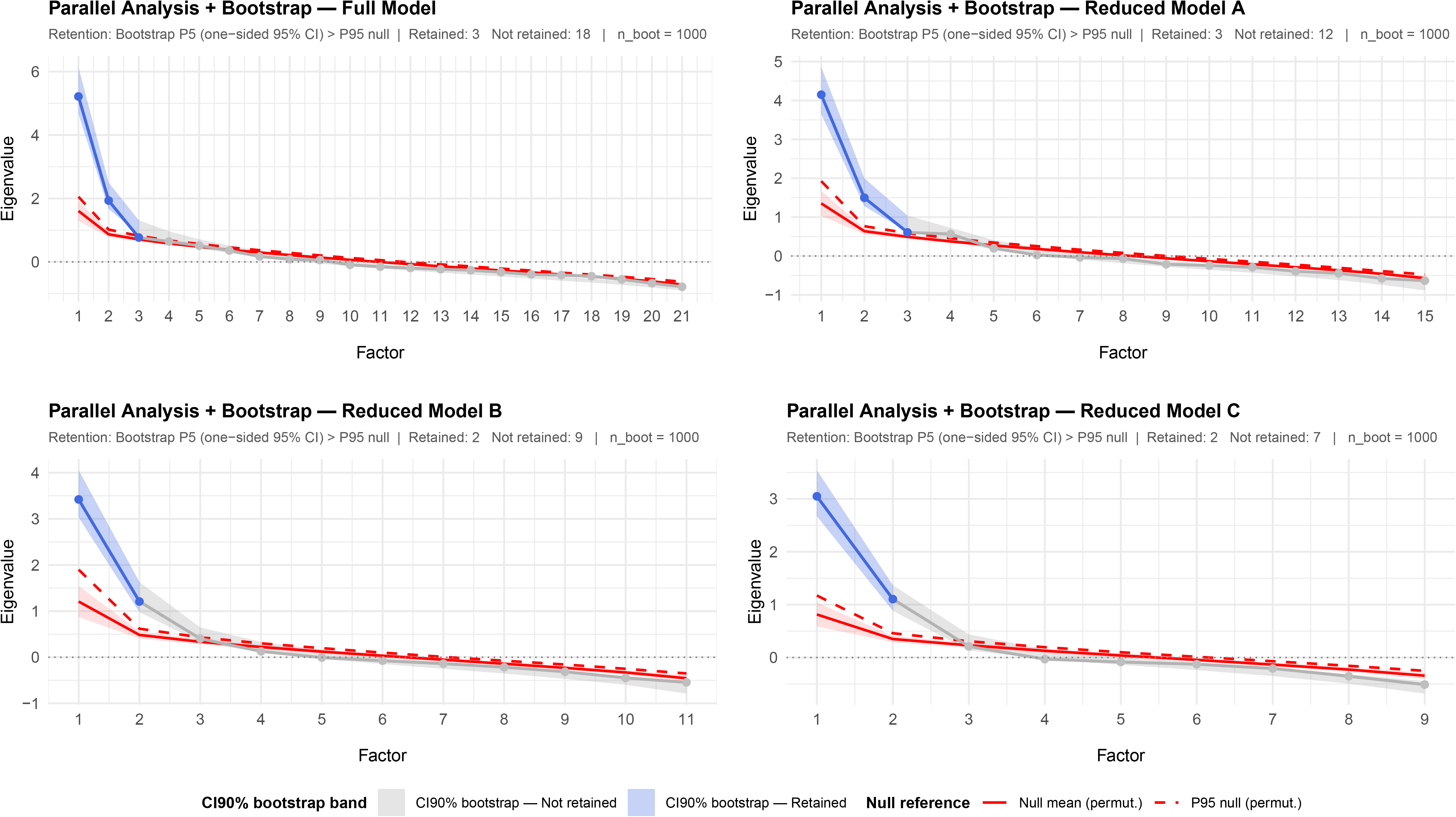
Bootstrap PA Reduced Model C

**Fig 4.**
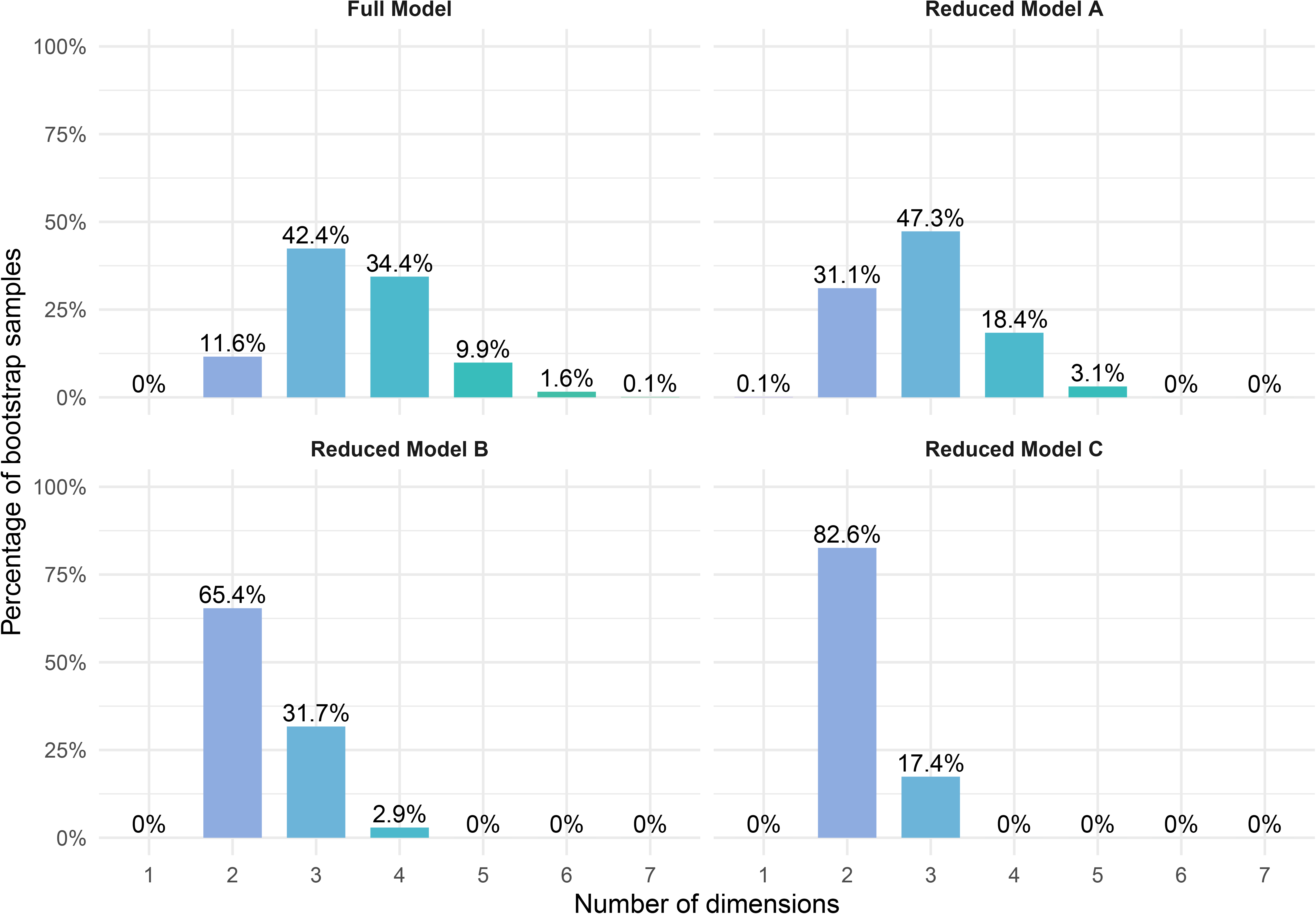
Dimensional solution bootstrap EGA Reduced Model C

Compared with the initial model, Reduced Model C showed improved FA fit indices (proportion of variance: MR1 = 0.297 and MR2 = 0.211; χ² = 86.82, p < 0.001; TLI = 0.80; RMSEA = 0.13; RMSR = 0.05). Nevertheless, the TLI and RMSEA continued to indicate limitations in global model fit. Bootstrap EGA provided stronger evidence of dimensional stability, with two communities recovered in 82.6% of bootstrap samples and structural consistency of 80.3% and 92.4% for Dim 1 and Dim 2, respectively. The empirical EGA network for Reduced Model C is presented in Figure 5, while the corresponding factor loadings and community loadings are shown in Figure 6.

**Fig 5.**
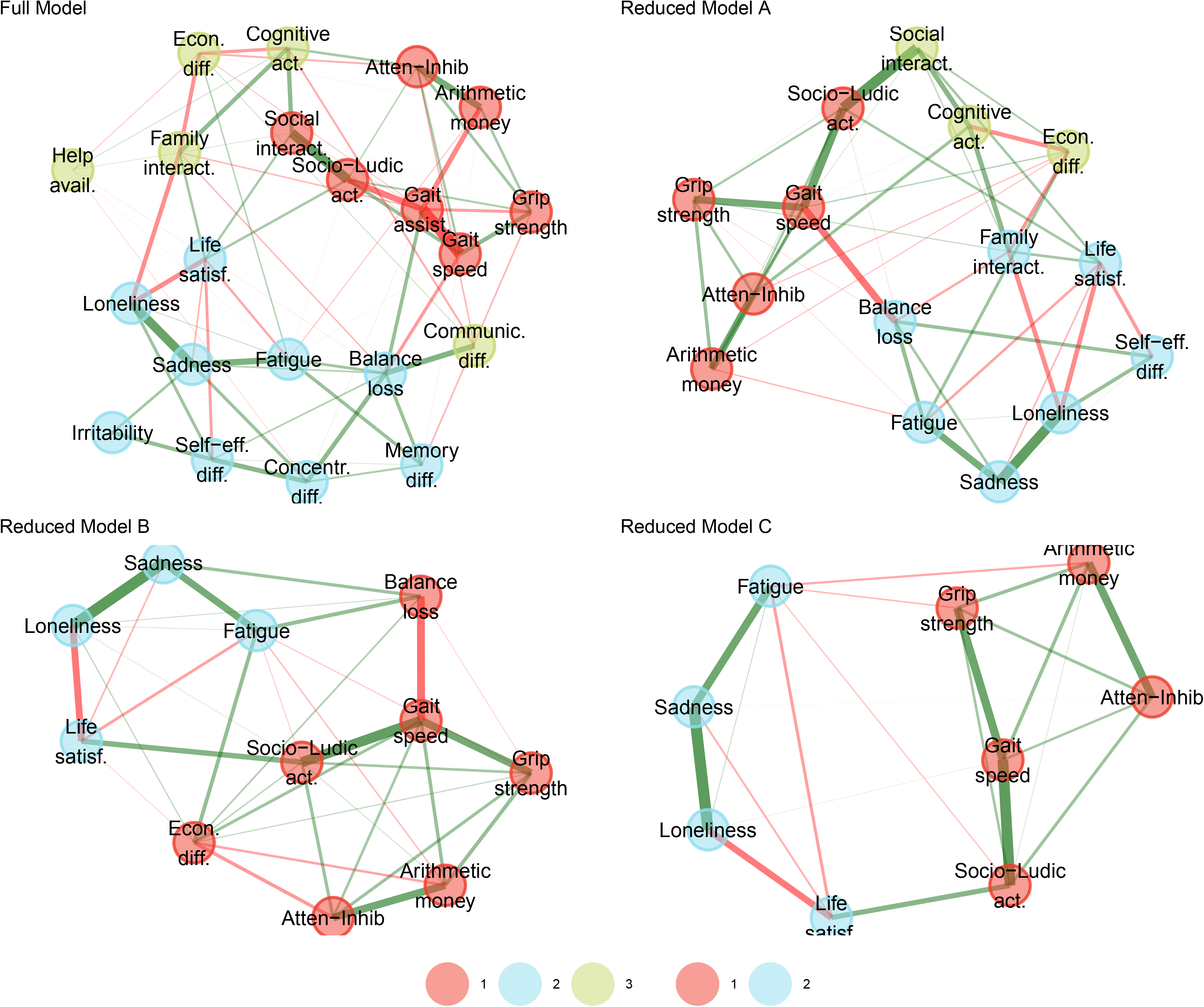
Empirical EGA Network Reduced Model C

**Fig 6.**
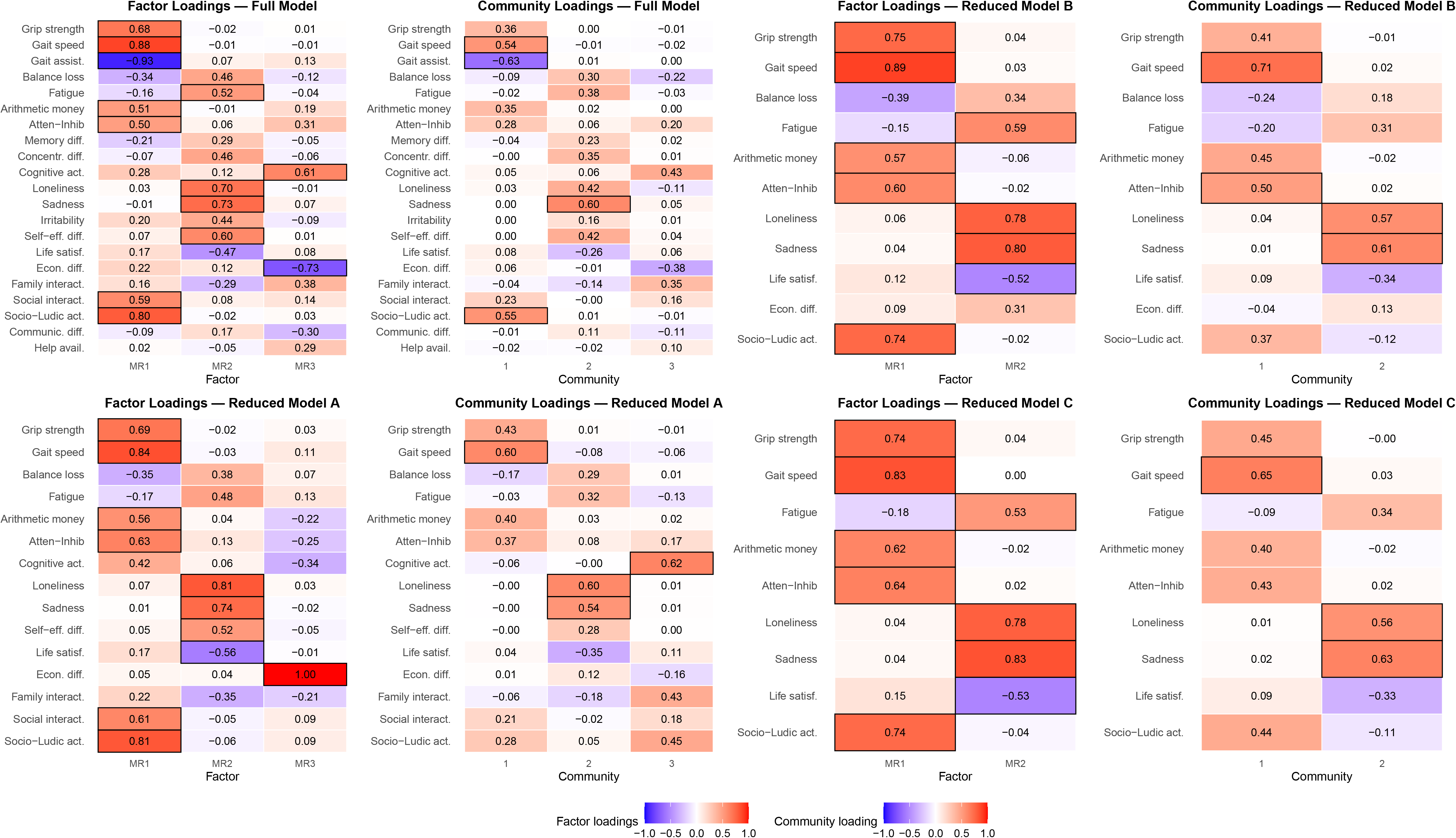
Factor loadings and Community Loadings Reduced Model C

Reduced Model C was retained on the basis of convergence between PA and EGA, improved dimensional stability, and relative improvement in FA fit and residual indices. The remaining discrepancies in global fit were considered when interpreting the retained solution. Detailed results for the successive models, including item-level diagnostic indicators and item retention or elimination decisions, are provided in the SM.

### 3.5. Internal consistency

Internal consistency for the Reduced Model C was good at the total scale level, with an omega total (ωt) of 0.864. However, hierarchical omega (ωh) was low (0.409), and the Explained Common Variance (ECV) was 0.327, indicating that despite the strong reliability of the full scale, only a small-to-moderate proportion of the common variance was attributable to a single general factor, supporting the existence of two distinguishable group factors rather than an essentially unidimensional structure, see Figure 7.

**Fig 7.**
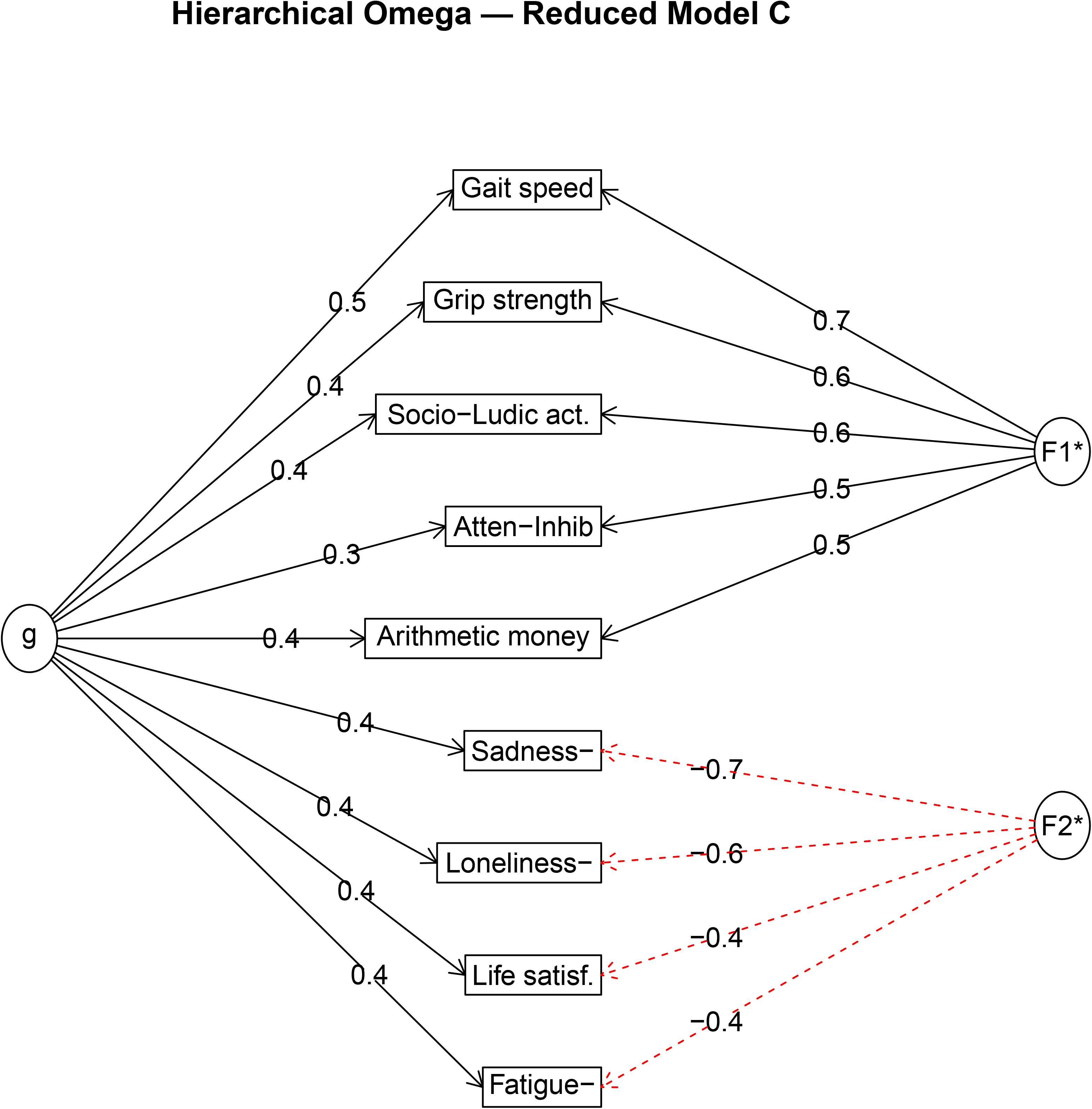
Hierarchical Omega

Subscale-level internal consistency, calculated from the main loading items of each factor, was good for Factor 1 (ω = 0.840, 5 items) and acceptable for Factor 2 (ω = 0.785, 4 items), both exceeding the 0.7 threshold. The omega decomposition by component and the item-level Schmid-Leiman solution are reported in SM.

### 3.6. Factor naming

The two retained factors were interpreted according to the content of their primary contributing items and the pattern of factor loadings.

Factor 1, designated “Autonomy”, comprised five primary indicators: gait speed, handgrip strength, socioludic activities, attention–inhibition, and money arithmetic. These items represented aspects of physical performance, cognitive functioning relevant to everyday activities, and participation in socioludic activities. Their shared contribution was interpreted as reflecting a dimension associated with functional autonomy, although the factor does not encompass all components of autonomy or independence in daily life.

Factor 2, designated “Psychological Distress”, comprised four primary indicators: sadness, loneliness, life satisfaction, and fatigue. These items represented emotional discomfort, subjective social experience, satisfaction with life, and perceived fatigue. Their shared contribution was interpreted as reflecting psychological distress, while recognizing that fatigue may also be influenced by physical health and other non-psychological factors. Collectively, these indicators align with the conceptual attributes of psychological distress described in the literature (Ridner, 2004).

Factor weightings, calculated preserving the obliquity, were reported as follows:

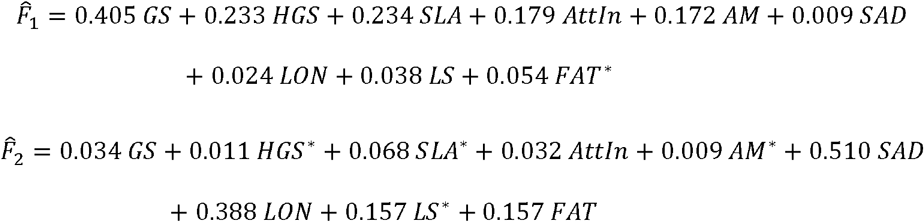

*Where:* GS = Gait speed; HGS = Grip strength; SLA = Socio-Ludic act.; AttIn = Atten-Inhib; AM = Arithmetic money; SAD = Sadness; LON = Loneliness; LS = Life satisf.; FAT = Fatigue. Indicators marked with * denote reverse-coded scores computed as *X_1_* * = 2 − *X* prior to factor score estimation.

### 3.7. Factor descriptive values

The score computation of both factors ranged from 0 points to their respective maximum possible scores (2.696 for Autonomy and 2.732 for Psychological Distress). Each factor yielded 19,683 possible scores, with maximum gaps of 0.009 between consecutive values. The scale can therefore be treated as continuous at two decimals, and scores should be reported with no more than two decimal places, as additional decimals do not provide meaningful precision.

Factor 1 (Autonomy) showed a mean of 1.75 ± 0.62, with observed values ranging from 0.36 (13.4% of the maximum scale value) to 2.69 (99.8%). The distribution was relatively symmetric (skewness = −0.32) and platykurtic (kurtosis = −1.13), with quartiles at Q1 = 1.20 (44.5%), median = 1.86 (69%), and Q3 = 2.33 (86.4%), indicating a tendency toward higher functional levels (see Figure 8).

**Figure 8.**
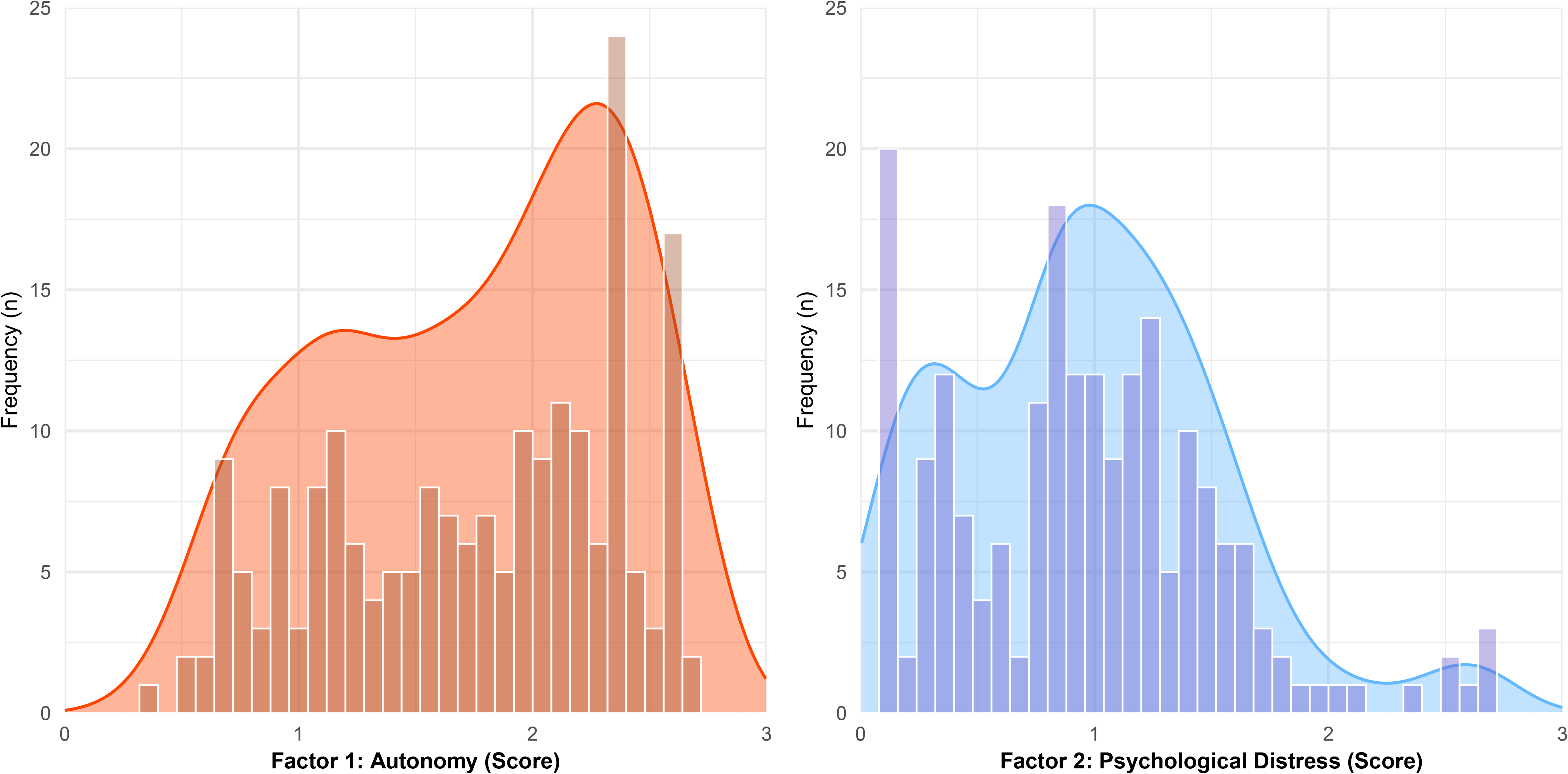
Factor response distribution

Factor 2 (Psychological Distress) presented a lower mean of 0.97 ± 0.57, with a wider observed range (0.09, 3.3%, to 2.69, 98.5%). The distribution showed positive skewness (0.59) and mild leptokurtosis (0.43), with quartiles at Q1 = 0.52 (19%), median = 0.93 (34%), and Q3 = 1.32 (48.3%), suggesting that most participants reported low levels of distress, with fewer cases at higher values, see Figure 8.

### 3.8. Factor-level test-retest reliability, SEM, and MDC

Both factors showed favourable point estimates of test–retest reliability. The intraclass correlation coefficient was 0.82 for Autonomy (95% CI: 0.613–0.923) and 0.78 for Psychological Distress (95% CI: 0.532–0.901). However, the relatively wide confidence intervals indicated uncertainty in the precision of these estimates, particularly given the limited number of participants included in the retest assessment.

The standard error of measurement (SEM) was 0.187 for Autonomy and 0.297 for Psychological Distress, corresponding to approximately 7% and 11% of their respective theoretical score ranges.

The minimum detectable change at the 95% confidence level (MDC95) was 0.551 for Autonomy and 0.872 for Psychological Distress, corresponding to 20.5% and 32.0% of the theoretical score ranges, respectively.

### 3.9. Factor-level floor-ceiling effects

No floor or ceiling effects were observed for either factor under the exact score criterion, as no participants obtained the minimum or maximum possible factor score. Under the primary items criterion, floor effects were negligible for Autonomy (0%) but more frequent for Psychological Distress, with 12.9% of participants scoring at the minimum on all primary items, below but close to the 15% threshold. Ceiling effects under this criterion were minor for both factors, affecting 10.9% of participants for Autonomy and only 2% for Psychological Distress. Similarly, under the within 10% range criterion, results remained below the 15% threshold across both factors and extremes, with values ranging from 3% (Psychological Distress ceiling) to 12.4% (Psychological Distress floor). Overall, no substantial floor or ceiling effects were detected for either factor.

### 3.10. Factor-level concurrent validity

The ordinal regression analyses showed significant associations between both EMFRA factors and all clinical variables, although the strength and direction of these relationships differed notably across predictors.

For Factor 1 (Autonomy), the strongest positive association was observed with physical performance (SPPB) (R² = 0.53, r = 0.75), indicating a strong positive relationship. This suggests that a substantial proportion of the variability in autonomy is explained by lower-extremity physical function. However, this relationship may be partially influenced by mathematical coupling, as gait speed contributes to both SPPB and the Autonomy factor.

A strong positive association was also found with global cognitive function (MoCA) (R² = 0.47, r = 0.70), and a moderate one with independence in basic ADLs (Barthel Index) (R² = 0.32, r = 0.56). In contrast, depressive symptoms showed a moderate negative association (R² = 0.18, r = −0.41), while anxiety (R² = 0.02, r = −0.12) and total HADS (R² = 0.09, r = −0.29) showed negligible-to-small relationships. These findings indicate that Autonomy is primarily related to physical functioning, global cognitive function, and independence in ADL domains, with a limited contribution from emotional factors, see Table 2, and Figure 9.

**Figure 9.**
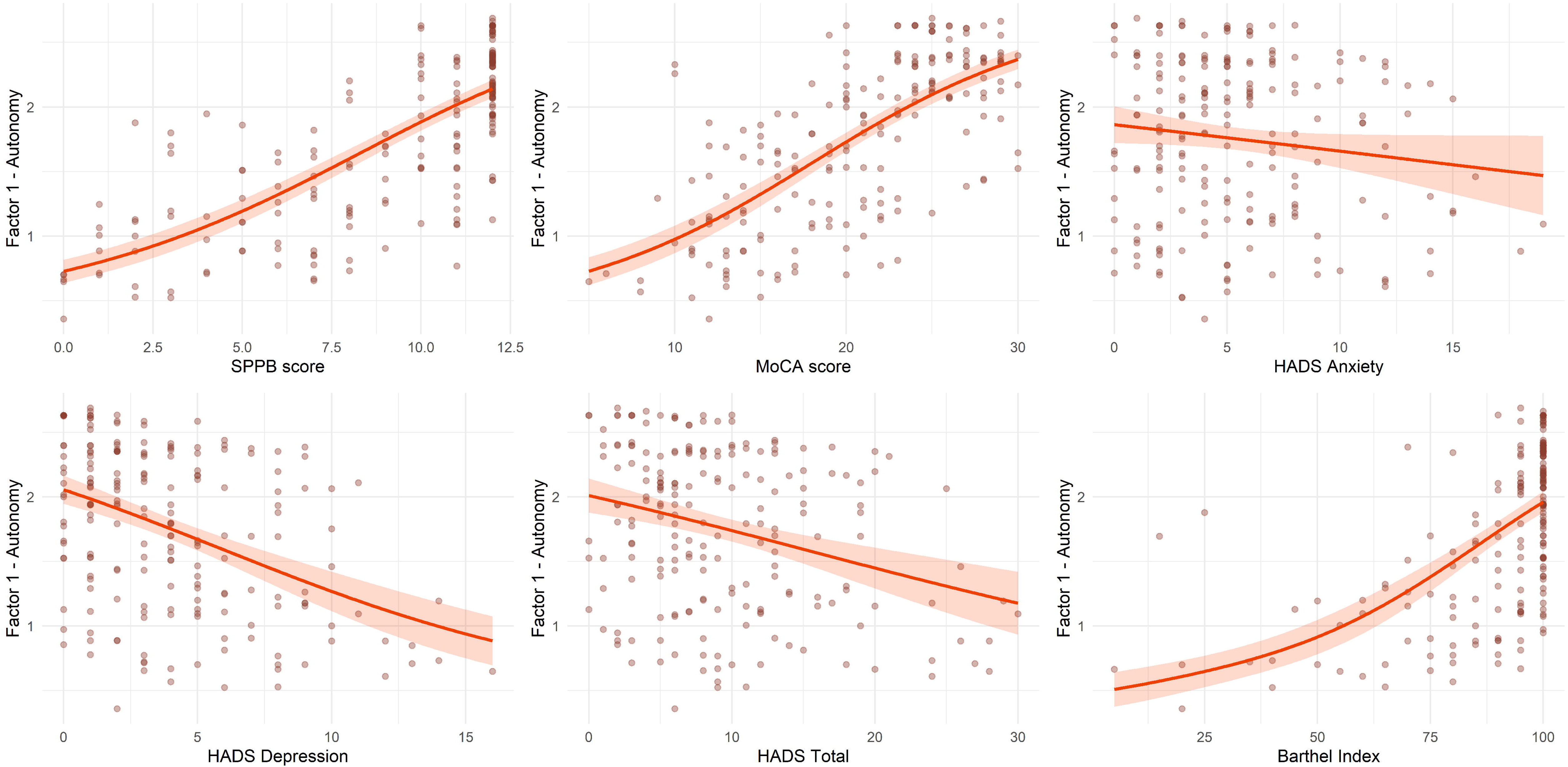
Concurrent plot Autonomy

**Table 2.**
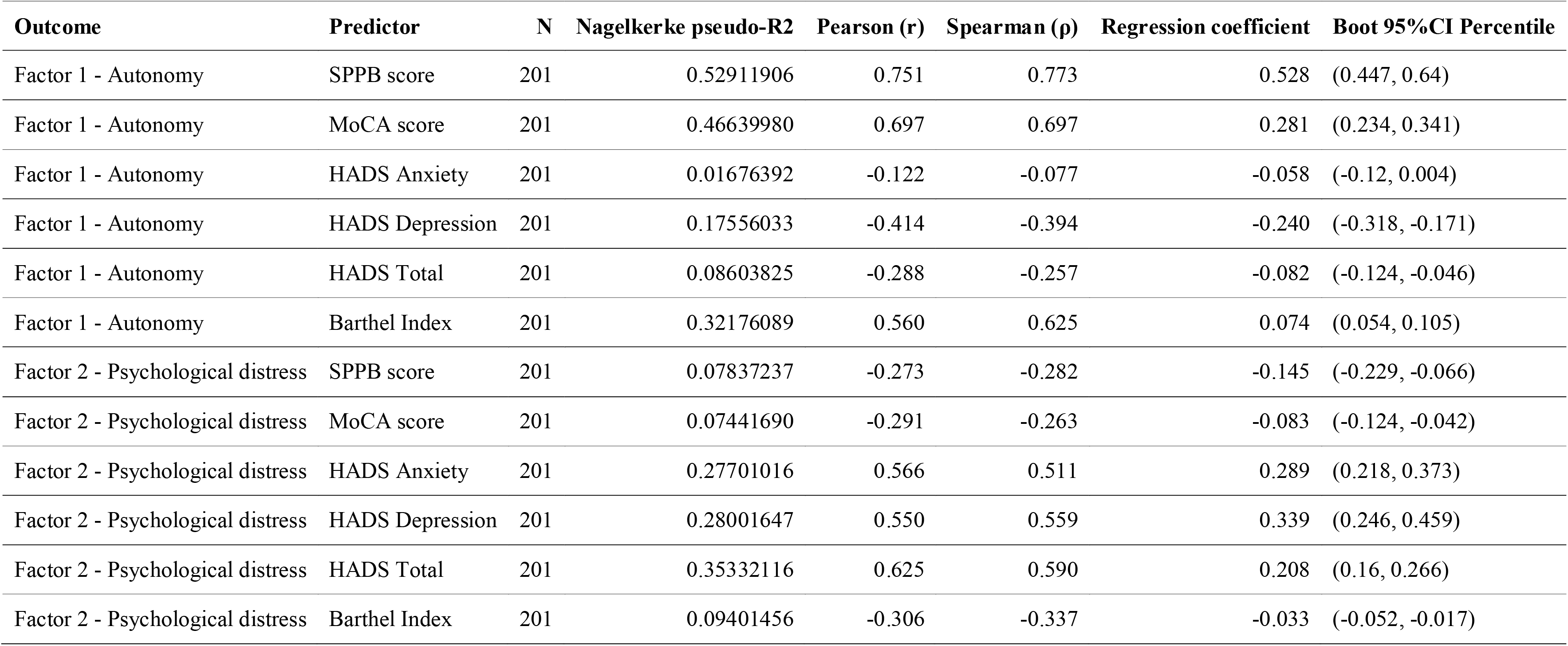
Concurrent validity.

| Outcome | Predictor | N | Nagelkerke pseudo-R2 | Pearson (r) | Spearman (ρ) | Regression coefficient | Boot 95%CI Percentile |
|---|---|---|---|---|---|---|---|
| Factor 1 - Autonomy | SPPB score | 201 | 0.52911906 | 0.751 | 0.773 | 0.528 | (0.447, 0.64) |
| Factor 1 - Autonomy | MoCA score | 201 | 0.46639980 | 0.697 | 0.697 | 0.281 | (0.234, 0.341) |
| Factor 1 - Autonomy | HADS Anxiety | 201 | 0.01676392 | -0.122 | -0.077 | -0.058 | (-0.12, 0.004) |
| Factor 1 - Autonomy | HADS Depression | 201 | 0.17556033 | -0.414 | -0.394 | -0.240 | (-0.318, -0.171) |
| Factor 1 - Autonomy | HADS Total | 201 | 0.08603825 | -0.288 | -0.257 | -0.082 | (-0.124, -0.046) |
| Factor 1 - Autonomy | Barthel Index | 201 | 0.32176089 | 0.560 | 0.625 | 0.074 | (0.054, 0.105) |
| Factor 2 - Psychological distress | SPPB score | 201 | 0.07837237 | -0.273 | -0.282 | -0.145 | (-0.229, -0.066) |
| Factor 2 - Psychological distress | MoCA score | 201 | 0.07441690 | -0.291 | -0.263 | -0.083 | (-0.124, -0.042) |
| Factor 2 - Psychological distress | HADS Anxiety | 201 | 0.27701016 | 0.566 | 0.511 | 0.289 | (0.218, 0.373) |
| Factor 2 - Psychological distress | HADS Depression | 201 | 0.28001647 | 0.550 | 0.559 | 0.339 | (0.246, 0.459) |
| Factor 2 - Psychological distress | HADS Total | 201 | 0.35332116 | 0.625 | 0.590 | 0.208 | (0.16, 0.266) |
| Factor 2 - Psychological distress | Barthel Index | 201 | 0.09401456 | -0.306 | -0.337 | -0.033 | (-0.052, -0.017) |

For Factor 2 (Psychological Distress), the pattern was reversed. Moderate associations were observed with emotional symptoms, particularly HADS total score (R² = 0.35, r = 0.63), followed by depression (R² = 0.28, r = 0.55) and anxiety (R² = 0.28, r = 0.57). In contrast, physical performance (SPPB) (R² = 0.08, r = −0.27), global cognitive function (MoCA) (R² = 0.07, r = −0.29) and independence in basic ADL (Barthel index) (R² = 0.09, r = −0.31) showed negligible-to-small negative associations, explaining only a minimal association. These results indicate that Psychological Distress factor is related as expected with emotional features, and not as related to physical functioning domains, global cognitive performance, or independence in basic ADL, see Table 2, and Figure 10.

**Figure 10.**
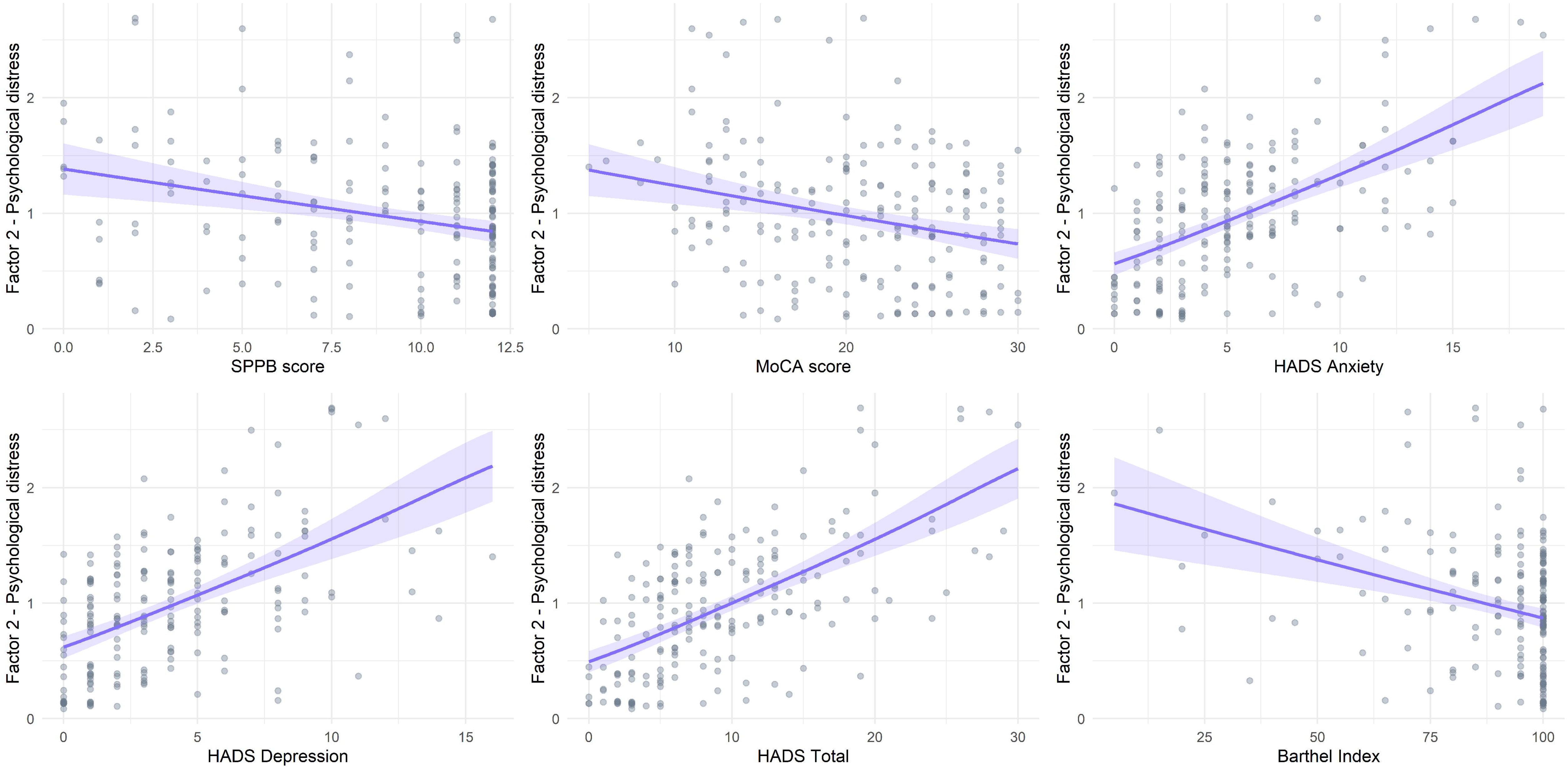
Concurrent plot psychological distress

## 4. Discussion

The present study examined the initial psychometric properties of EMFRA, including its dimensional structure, internal consistency, temporal reliability, measurement error, floor and ceiling effects, and associations with established measures of functioning. The principal finding was a nine-item solution comprising two dimensions, Autonomy and Psychological Distress, which differed from the four domains proposed during instrument development. The retained factors showed internal consistency and associations with external measures consistent with the proposed interpretation. However, limitations of global FA fit, explained variance, and in the precision of test-retest estimates indicate that the psychometric evidence remains preliminary.

EMFRA was originally developed to assess physical, cognitive, emotional, and social aspects of functioning in older adults (Fierro-Marrero, Reina-Varona, Pardo-Montero, et al., 2025). Its development involved an initial pool of 24 items distributed across four domains, subsequently reduced to 21 items before the present psychometric evaluation. The empirical results did not reproduce the proposed four-dimensional organization. Instead, both bootstrap PA and bootstrap EGA converged on a three-dimensional solution with poor fit and limited dimensional stability. This discrepancy may be particularly attributable to the limited item pool per domain, with only five-to-six items per domain, which likely provided an insufficient coverage to represent each intended dimension. Some overlap between domains may also be expected, given the interrelated nature of functioning in later life. A more robust development approach would have involved generating a larger item pool from the outset, enabling the construction of both an extended and a reduced version of the scale while better preserving the intended dimensional coverage.

Following a progressive item refinement process, the Reduced Model C composed of nine items, showed a stable structure. PA and EGA converged on a two-dimensional solution, with two communities recovered in 82.6% of bootstrap samples and structural consistency of 80.3% and 92.4% for the respective dimensions. Nevertheless, FA performance remained limited (TLI = 0.80; RMSEA = 0.13). Global fit indices have shown poor performance as criteria for selecting the number of factors in exploratory FA, where conventional cutoffs are not sufficiently reliable (Montoya & Edwards, 2021). This fact allows the presence of poor model fit with a consistently recovered dimensional structure, as observed in Reduced Model C. In addition, bootstrap methods in both PA and EGA allow to internally validate the model structure across possible samples. These considerations support evaluating PA and EGA convergence, dimensional stability, and FA fit jointly, but they do not justify disregarding the remaining misfit (Garcia-Garzon et al., 2019). Reduced Model C therefore represents the most defensible configuration among those examined, and its performance was considered acceptable given the exploratory stage of scale validation.

Notably, the two main factors remained relatively stable between the full 21-item model and Reduced Model C, suggesting that the detected structure was not an artefact of the item reduction process. Nevertheless, item elimination narrowed the set of indicators available to describe functioning. Indicators grouped within these factors in the full model, such as gait assistance, social interaction, balance loss, memory difficulty, concentration difficulty, and irritability, were not retained, and the four originally proposed domains are no longer represented as separate dimensions. Because the content validity evidence obtained during instrument development (Fierro-Marrero, Reina-Varona, Pardo-Montero, et al., 2025) refers to the original item set, the comprehensiveness of the nine-item version should be additionally analysed through a new content validity process (Terwee et al., 2018).

The Autonomy factor comprised gait speed, handgrip strength, socioludic activity, attention– inhibition, and money arithmetic. These indicators combine aspects of physical performance, cognitive abilities relevant to everyday tasks, and social participation. This cross-domain composition is compatible with a construct related to functional independence, particularly in instrumental ADL, although the few items will only provide a partial coverage of the whole construct assessment. A relevant feature of this factor is the integration of performance-based tests, and self-reported measures addressing everyday functioning. This may be relevant given the discordance reported between self-reported and directly measured functioning in older adults, where older adults may over or even underestimate their physical performance depending on certain individual features (Liu et al., 2026; Schmidle et al., 2023). Therefore, the inclusion of performance-based indicators within Autonomy may reduce such biases.

The Psychological Distress factor comprised sadness, loneliness, life satisfaction, and fatigue. The first three indicators reflect related aspects of emotional experience and subjective well-being, although they do not necessarily represent identical psychological processes. Fatigue introduces a broader component whose interpretation requires particular caution. In older adults, fatigue may be associated with physical health conditions, activity limitations, and psychological symptoms (Yu et al., 2010). In the present sample, fatigue showed its strongest association with sadness (ρ = 0.52), while its association with balance loss was also notable (ρ = 0.41). This pattern suggests that the multidimensional scope of fatigue, including psychological and even physical components may contribute to Psychological Distress. In the full model, this factor additionally included balance loss, memory difficulty, concentration difficulty, and irritability, a grouping compatible with the reported co-occurrence of emotional and cognitive difficulties in older adults (Biringer et al., 2005; Dufouil et al., 1996). However, these indicators were not retained, and the narrower final configuration provides a more specific basis for interpreting the factor.

The internal consistency findings have important implications for score interpretation. Although ωt was 0.864, ωh (0.409), and ECV (0.327) indicated that a substantial proportion of the reliable variance was not attributable to a single general factor (Reise, 2012). The present results consequently provide limited support for calculating a single EMFRA score intended to represent general functioning. This is particularly relevant in geriatric assessment, where domain-specific profiling is essential for targeted assessments (Brown et al., 1988; Fierro-Marrero, Reina-Varona, Paris-Alemany, et al., 2025). Reliability was instead good for Autonomy (ω = 0.840) and acceptable for Psychological Distress (ω = 0.785), both exceeding the recommended threshold of 0.70 (McNeish, 2018), supporting the separate use of both factors. These subscale coefficients also include variance shared with the general factor (Reise, 2012), but given the weakness of this factor, most of their reliable variance is likely specific to each dimension, which is also observed through the modest interfactor correlation (Phi = −0.316) reported in SM. Finally, the proportion of total item variance explained by each factor was modest (29.7% for Autonomy and 21.1% for Psychological Distress). This is not unexpected given the multidimensional nature of both constructs and the limited number of items retained per factor in a brief screening instrument.

Test-retest reliability findings were less conclusive. Although the factor-level ICCs were 0.82 for Autonomy and 0.78 for Psychological Distress, their respective 95% CIs (0.613–0.923 and 0.532–0.901) indicate considerable uncertainty in the underlying reliability estimates. At the item level, Loneliness (AC2 = 0.46, 95%CI 0.19–0.74) and Life Satisfaction (AC2 = 0.39, 95%CI 0.12–0.67) showed the lowest agreements. They were retained because the imprecision of their estimates did not allow taking a definitive decision, and because they presented substantial loadings on Psychological Distress, so their exclusion could have substantially altered the composition of the factor.

Nevertheless, their structural contribution should not obviate their limited temporal reliability, a relevant fact for item exclusion. Regarding SEM and MDC95, Autonomy showed values of 0.187 and 0.551 respectively (approximately 7% and 20.5% of the theoretical score range) and 0.297 and 0.872 respectively for Psychological Distress (approximately 11% and 32%). With the current data, the latter values indicate that considerable changes over time would be required to reliably confirm any change in the factor score. Taken together the current evidence is insufficient to support the use of either factor for monitoring individual changes over time.

Associations with established measures provided initial evidence consistent with the proposed names of the factors. Autonomy showed strong associations with SPPB (r = 0.751) and MoCA (r = 0.697), together with a moderate association with Barthel (r = 0.56). These findings correspond to the physical and cognitive content of the retained indicators. However, the association with SPPB should be interpreted with caution, as gait speed contributes to both measures and this overlap may have inflated the correlation. The moderate association with Barthel is consistent with the partial coverage of basic ADL within Autonomy. Psychological Distress showed a stronger association with HADS (r = 0.625) than with measures of physical performance, global cognitive functioning, or independence in ADL. This pattern is compatible with the emotional content of the factor and provides preliminary evidence of differentiated relationships with external constructs.

No pronounced floor or ceiling effects were observed for either factor. This finding is relevant given the characteristics of the sample, which was predominantly composed of participants with preserved independence in ADL (Barthel score: 90.02 ± 17.84), and low levels of depressive and anxiety traits (HADS depression: 3.9 ± 3.37; HADS anxiety: 5.38 ± 3.89). In such a sample, scores could be expected to cluster near to the ceiling of Autonomy and the floor of the Psychological Distress. The absence of these effects suggests that both factors discriminate adequately among older adults with high autonomy and good psychological well-being. Future studies should examine whether this also holds in sample with a poorer health state.

### 4.1. Clinical implications

The nine-item EMFRA may be useful for the preliminary characterization of functioning in settings where comprehensive geriatric assessment is difficult to implement, such as primary care. Its two-dimensional structure allows clinicians to screen for autonomy, and psychological distress separately, helping to identify which dimensions may require a more thorough assessment, and to orientate referral decisions, for instance toward physiotherapy, occupational therapy, or psychology, without substantially increasing assessment burden. In addition, combining performance-based and self-reported items may reduce the biases associated with relying on self-report alone (Liu et al., 2026; Schmidle et al., 2023). Nonetheless, diagnostic thresholds, predictive accuracy, administration time, and feasibility across settings have not yet been established. EMFRA should therefore not currently be used for individual clinical classification, nor for monitoring changes over time.

### 4.2. Study Limitations and Future Research

The dimensional analyses were conducted in one single sample, without validation in an external sample. Bootstrap methods, allowed to internally validate the structure, but only within the available data. Moreover, participants predominantly showed independence in ADLs and relatively good physical and cognitive functioning, and these sample characteristics may have shaped the observed structure. Both the three-factor structure of the 21-item version and the two-factor structure of the 9-item version, should therefore be examined in external samples that include older adults with greater physical dependency, cognitive impairment, or psychosocial difficulties, across different settings.

A second limitation concerns the naming of both factors. The labels Autonomy and Psychological Distress were assigned based on the contribution of each item to its factor, and on the available literature, but no formal content validation was performed. Future studies should examine whether the retained items adequately represent the proposed dimensions.

A particularly important limitation is the small subsample used to assess test-retest reliability SEM and MDC95. Only 22 participants completed the repeated assessment, which resulted in imprecise estimates. This imprecision limited decisions on whether to retain or exclude items with low reliability, and prevented a confident estimate of the minimum change in factor scores required to be considered a true change.

Finally, other psychometric properties were not examined, such as the predictive validity, responsiveness, or how the floor or ceiling effects of EMFRA compare with those of other instruments assessing the same constructs. Future research should also determine whether EMFRA can be feasibly implemented in routine healthcare settings and whether it improves clinicians’ decisions about further assessment or referral.

## 5. Conclusions

The present study provides the first psychometric evidence of EMFRA. The original 21-item version did not reproduce the four hypothesized domains but yielded a three-dimensional solution. Item refinement provided a nine-item version composed of two dimensions, Autonomy and Psychological Distress, on which bootstrap PA, and bootstrap EGA, converged and showed improved dimensional stability, although with limited fit. Both factors showed adequate internal consistency, no floor or ceiling effects, and associations with established clinical measures consistent with their interpretation. EMFRA should be scored and interpreted through its two factors separately, and not through a single total score. Test-retest reliability, SEM and MDC95 remain uncertain. Future studies should confirm the dimensional structure in an external sample, provide further psychometric properties, and evaluate its feasibility and performance across clinical settings.

## Supporting information

R Quarto Script File

Database - Age Eliminated

## Data Availability

All data produced in the present study are available upon reasonable request to the authors

## 7. Author contributions

JFM: conceptualization, methodology development, research, data curation, formal analysis, funding acquisition, data visualization, writing - original draft, writing - review and editing. AGG, LCD, SMB: research, data curation. APA and RLT: conceptualization, methodology development, project administration, supervision, funding acquisition, project administration, supervision, writing - review and editing.

## 9. Acknowledgments

The authors wish to thank the STIMA day-care centres of Princesa, Embajadores, Aluche, Moratalaz, and Hortaleza, and the Valdeluz Arroyomolinos nursing home for their collaboration and commitment to this project, without which the recruitment of participants would not have been possible.

