## Supplementary material for "Psychometric analysis of the Evaluation of Multidimensional Functioning and Risks in Aging (EMFRA) scale: reliability, structural validity, and concurrent validity": R Quarto Script File

### Analysis Results of EMFRA

José Fierro-Marrero

2026-07-22

**6 Factor descriptives** **208**

**7 Factor-level test-retest reliability, SEM, and MDC.** **213**

**8 Factor-level floor-ceiling effects** **217**

**9 Concurrent validity** **222**

### 1 Descriptive sample data

This section reports the descriptive analysis conducted to examine the distribution of responses across all items of the EMFRA scale. Only participants with complete evaluations (EMFRA and additional assessments) were included in the analysis (n = 201). All items were analysed using their original three-category ordinal response format (0, 1, and 2), where higher values indicate a greater level of the assessed trait.

#### 1.1 Packages and data preparation

The following packages were loaded to support data import, data manipulation, descriptive statistics, and graphical output. The EMFRA database was imported from an Excel file, and only participants with complete evaluations were retained. Additionally, vectors containing the EMFRA item identifiers and their corresponding descriptive labels were defined for subsequent analyses.

```
This analysis was conducted using R version 4.6.0 (2026-04-24 ucrt) running on
 ↪  Windows 10 x64 (platform: x86_64-w64-mingw32/x64). The R session was executed
 ↪  under Windows 11 x64 (build 26200) with locale settings
 ↪  LC_COLLATE=Spanish_Spain.utf8;LC_CTYPE=Spanish_Spain.utf8;LC_MONETARY=Spanish
 ↪  _Spain.utf8;LC_NUMERIC=C;LC_TIME=Spanish_Spain.utf8. The versions of all R
 ↪  packages used in the analysis are reported in Table ref(tbl-r-packages).
```

```
library(readxl)
library(dplyr)
library(tidyr)
library(stringr)
library(tibble)
library(ggplot2)
library(ggthemes)
library(scales)
library(paletteer)
library(ggridges)
library(svglite)
library(grid)
library(gridExtra)
library(patchwork)
library(cowplot)
library(rlang)
library(CHOIRBM)
library(knitr)
library(purrr)
library(irrCAC)
library(matrixcalc)
library(psych)
library(EGAnet)
library(rms)
library(kableExtra)
library(flextable)
```

Table 1: Versions of R packages used in the analysis

| Package | Version |
|---|---|
| CHOIRBM | 0.0.2 |
| EGAnet | 2.3.0 |
| Hmisc | 5.2-5 |
| cowplot | 1.2.0 |
| dplyr | 1.2.1 |
| flextable | 0.9.12 |
| future | 1.70.0 |
| ggplot2 | 4.0.3 |
| ggridges | 0.5.7 |
| ggthemes | 5.2.0 |
| gridExtra | 2.3 |
| irrCAC | 1.4 |
| kableExtra | 1.4.0 |
| knitr | 1.51 |
| lavaan | 0.6-21 |
| matrixcalc | 1.0-6 |
| paletteer | 1.7.0 |
| patchwork | 1.3.2 |
| psych | 2.5.6 |
| purrr | 1.2.2 |
| readxl | 1.5.0 |
| rlang | 1.2.0 |
| rms | 8.1-1 |
| scales | 1.4.0 |
| semPlot | 1.1.8 |
| semTools | 0.5-8 |
| stringr | 1.6.0 |
| svglite | 2.2.2 |
| tibble | 3.3.1 |
| tidyr | 1.3.2 |

```
data_emfra <- read_excel("Database EMFRA Complete.xlsx") %>%
filter(Complete_evaluation == "Yes")

vars_descriptives_raw <- c(
  "Age",
  "Height",
  "Weight",
  "BMI",
  "Years_studied",
  "SPPB_balance_points",
  "SPPB_walk_second_mean",
  "SPPB_walk_final_points",
  "SPPB_FTSS_mean",
```

```r
  "SPPB_FTSS_final_points",
  "SPPB_points",
  "MoCA_without_studies",
  "MOCA_with_studies",
  "Handgrip_mean",
  "Barthel_total",
  "HADS_anxiety_total",
  "HADS_depression_total",
  "HADS_total",
  "EMFRA_time_total_minutes_retest"
)

vars_descriptives_labels <- c(
  Age                             = "Age (years)",
  Height                          = "Height (m)",
  Weight                          = "Weight (kg)",
  BMI                             = "Body Mass Index (kg/m²)",
  Years_studied                   = "Years of formal education",
  SPPB_balance_points             = "SPPB - Balance score",
  SPPB_walk_second_mean           = "Gait time in 2.4m (s)",
  SPPB_walk_final_points          = "SPPB - Gait speed score",
  SPPB_FTSS_mean                  = "FTSTS (s)",
  SPPB_FTSS_final_points          = "SPPB - FTSTS score",
  SPPB_points                     = "SPPB - Total score",
  MoCA_without_studies            = "MoCA (raw score)",
  MOCA_with_studies               = "MoCA (education-adjusted)",
  Handgrip_mean                   = "Handgrip strength (kg)",
  Barthel_total                   = "Barthel Index - Total score",
  HADS_anxiety_total              = "HADS - Anxiety score",
  HADS_depression_total           = "HADS - Depression score",
  HADS_total                      = "HADS - Total score",
  EMFRA_time_total_minutes_retest = "EMFRA administration time (min)"
)

vars_categorical_raw <- c(
  "Sex",
  "Educational_level",
  "Marital_status",
  "Laboral_status"
)

vars_categorical_labels <- c(
  Sex                             = "Sex",
  Educational_level               = "Educational level",
  Marital_status                  = "Marital status",
  Laboral_status                  = "Laboral status"
)
```

```r
emfra_item_ids <- c(
"EMFRA_item1_trait","EMFRA_item2_trait","EMFRA_item3_trait",
"EMFRA_item4_trait","EMFRA_item5_trait","EMFRA_item6_trait",
"EMFRA_item7_trait","EMFRA_item8_trait","EMFRA_item9_trait",
"EMFRA_item10_trait","EMFRA_item11_trait","EMFRA_item12_trait",
"EMFRA_item13_trait","EMFRA_item14_trait","EMFRA_item15_trait",
"EMFRA_item16_trait","EMFRA_item17_trait","EMFRA_item18_trait",
"EMFRA_item19_trait","EMFRA_item20_trait","EMFRA_item21_trait"
)

emfra_item_ids_retest <- paste0(emfra_item_ids, "_retest")

emfra_item_labels <- c(
"Item 1. Hand-grip strength", "Item 2. Gait speed",
"Item 3. Gait assistance", "Item 4. Balance loss",
"Item 5. Fatigue", "Item 6. Arithmetics for money",
"Item 7. Attention-inhibition", "Item 8. Memory difficulty",
"Item 9. Concentration difficulty", "Item 10. Cognitive activities",
"Item 11. Loneliness", "Item 12. Sadness",
"Item 13. Irritability", "Item 14. Self-efficacy difficulty",
"Item 15. Life satisfaction", "Item 16. Economical difficulty",
"Item 17. Family interaction", "Item 18. Social interaction",
"Item 19. Social-ludic activities", "Item 20. Communication difficulty",
"Item 21. Help availability"
)

emfra_item_labels_short <- c(
  "Grip strength", "Gait speed", "Gait assist.", "Balance loss", "Fatigue",
  "Arithmetic money", "Atten-Inhib", "Memory diff.", "Concentr. diff.",
  "Cognitive act.", "Loneliness", "Sadness", "Irritability", "Self-eff. diff.",
  "Life satisf.",  "Econ. diff.", "Family interact.", "Social interact.",
  "Socio-Ludic act.", "Communic. diff.", "Help avail."
)

label_map <- setNames(
  emfra_item_labels,
  emfra_item_labels_short
)
```

#### 1.2 Baseline descriptive analysis

##### 1.2.1 Calculation functions

```r
get_continuous_stats <- function(data, vars) {
```

```r
  psych::describe(
    data %>% select(all_of(vars)),
    na.rm  = TRUE,
    skew   = TRUE,
    ranges = TRUE,
    quant  = c(.25, .50, .75),
    check  = FALSE
  ) %>%
    as.data.frame() %>%
    tibble::rownames_to_column("Variable")
}
```

```r
format_continuous_table <- function(desc, labels = NULL, digits = 2) {

  if (!is.null(labels)) {
    desc <- desc %>%
      mutate(
        Variable = dplyr::recode(Variable, !!!labels)
      )
  }

  desc %>%
    select(
      Variable,
      mean,
      sd,
      Min      = min,
      Q1       = Q0.25,
      Median   = median,
      Q3       = Q0.75,
      Max      = max,
      Skewness = skew,
      Kurtosis = kurtosis
    ) %>%
    mutate(
      across(-Variable, ~ round(.x, digits)),
      `Mean ± SD` = paste0(mean, " ± ", sd)
    ) %>%
    select(
      Variable,
      `Mean ± SD`,
      Min,
      Q1,
      Median,
      Q3,
      Max,
      Skewness,
```

```
      Kurtosis
    )
}
```

```
get_categorical_descriptives <- function(data, vars) {

  data %>%
    select(all_of(vars)) %>%
    pivot_longer(
      cols = everything(),
      names_to = "Variable",
      values_to = "Value"
    ) %>%
    group_by(Variable, Value) %>%
    summarise(
      n = n(),
      .groups = "drop"
    ) %>%
    group_by(Variable) %>%
    mutate(
      percent = 100 * n / sum(n)
    )
}
```

```
format_categorical_table <- function(categorical_df, labels = NULL, digits = 2) {

  df <- categorical_df

  if (!is.null(labels)) {
    df <- df %>%
      mutate(
        Variable = dplyr::recode(Variable, !!!labels)
      )
  }

  df %>%
    mutate(
      percent = round(percent, digits),
      category_label = paste0(
        Value, " (", n, ", ", percent, "%)"
      )
    ) %>%
    arrange(Variable, desc(n)) %>%
    group_by(Variable) %>%
    summarise(
      Categories = paste(category_label, collapse = " | "),
      .groups = "drop"
```

```
    )
}
```

```
plot_donut_thin <- function(
  data,
  var,
  title,
  palette = NULL,
  colors  = NULL,
  digits  = 0
) {

  var_sym <- sym(var)

  df <- data %>%
    filter(!is.na(!!var_sym)) %>%
    count(!!var_sym, name = "n") %>%
    mutate(
      percent = round(100 * n / sum(n), digits),
      label   = paste0(percent, "%")
    )

  p <- ggplot(
    df,
    aes(
      x = 2,
      y = n,
      fill = !!var_sym
    )
  ) +
    geom_col(width = 0.7, color = "white", linewidth = 0.2) +
    coord_polar(theta = "y") +
    geom_text(
      aes(label = label),
      position = position_stack(vjust = 0.5),
      color = "white",
      face  = "bold",
      size  = 2.8
    ) +
    xlim(0.5, 2.5) +
    labs(
      title = title,
      fill  = NULL
    ) +
    theme_void() +
    theme(
      plot.title = element_text(
```

```
        hjust = 0.5,
        face  = "bold",
        size  = 11
      ),
      legend.position = "right",
      legend.text = element_text(size = 9)
    )

  if (!is.null(palette)) {

    p <- p + scale_fill_paletteer_d(palette)

  } else if (!is.null(colors)) {

    p <- p + scale_fill_manual(values = colors)

  }

  p
}
```

```
compute_affection_and_types <- function(
  data,
  var_raw,
  affection_name,
  recode_map = NULL,
  cancer_terms = NULL
) {

  n_total <- nrow(data)

  data <- data %>%
    mutate(
      !!affection_name := ifelse(
        .data[[var_raw]] == "No" |
          is.na(.data[[var_raw]]) |
          .data[[var_raw]] == "",
        "No",
        "Yes"
      )
    )

  affection_summary <- data %>%
    count(.data[[affection_name]], name = "n") %>%
    mutate(
      percent = round(n / n_total * 100, 1),
```

```
      label    = paste0(n, " (", percent, "%)")
    )

  type_summary <- data %>%
    mutate(subject_id = row_number()) %>%
    select(subject_id, !!sym(var_raw)) %>%
    filter(
      !is.na(.data[[var_raw]]),
      .data[[var_raw]] != "",
      .data[[var_raw]] != "No"
    ) %>%
    separate_rows(!!sym(var_raw), sep = ",") %>%
    mutate(type_raw = str_trim(.data[[var_raw]]))

  if (!is.null(recode_map)) {
    type_summary <- type_summary %>%
      mutate(
        type = recode(type_raw, !!!recode_map, .default = type_raw)
      )
  } else {
    type_summary <- type_summary %>% mutate(type = type_raw)
  }

  if (!is.null(cancer_terms)) {
    type_summary <- type_summary %>%
      mutate(
        type = ifelse(type_raw %in% cancer_terms, "Cancer", type)
      )
  }

  type_summary <- type_summary %>%
    distinct(subject_id, type) %>%
    count(type, name = "n_subjects") %>%
    mutate(
      percent    = round(n_subjects / n_total * 100, 1),
      label      = paste0(n_subjects, " (", percent, "%)"),
      fill_value = n_subjects
    ) %>%
    arrange(n_subjects)

  list(
    data = data,
    affection_summary = affection_summary,
    type_summary = type_summary,
    n_total = n_total
  )
}
```

```r
plot_affection_yes_no <- function(
  summary_df,
  n_total,
  title,
  yes_color,
  no_color = "grey60"
) {

  ggplot(
    summary_df,
    aes(x = n, y = !!sym(names(summary_df)[1]), fill =
      ↪  !!sym(names(summary_df)[1]))
  ) +
    geom_col(
      width = 0.7,
      alpha = 0.85,
      color = "white",
      linewidth = 0.5
    ) +
    geom_text(
      aes(label = label),
      hjust = -0.2,
      size  = 2.8
    ) +
    scale_fill_manual(
      values = c("No" = no_color, "Yes" = yes_color)
    ) +
    scale_x_continuous(
      limits = c(0, n_total),
      expand = expansion(mult = c(0, 0.05))
    ) +
    labs(
      title = title,
      x = "Number of subjects",
      y = NULL
    ) +
    theme_minimal() +
    theme(
      plot.title = element_text(hjust = 0.5, face = "bold", size = 10),
      legend.position = "none",
      axis.text.y = element_text(face = "bold", size = 8),
      axis.text.x = element_text(size = 8),
      axis.title.x = element_text(size = 9)
    )
}

plot_type_histogram <- function(
  type_df,
```

```r
  n_total,
  title,
  fill_color = NULL,
  palette = NULL
) {

  p <- ggplot(
    type_df,
    aes(
      x = n_subjects,
      y = reorder(type, n_subjects)
    )
  )

  if (!is.null(palette)) {

    p <- p +
      geom_col(
        aes(fill = fill_value),
        alpha = 0.9,
        color = "white",
        linewidth = 0.5
      ) +
      scale_fill_gradientn(
        colours = palette,
        limits  = c(0, n_total),
        oob     = scales::squish
      )

  } else {

    p <- p +
      geom_col(
        fill  = fill_color,
        alpha = 0.9,
        color = "white",
        linewidth = 0.5
      )
  }

  p +
    geom_text(
      aes(label = label),
      hjust = -0.05,
      size  = 2.6
    ) +
    scale_x_continuous(
      limits = c(0, 150),
```

```
      expand = expansion(mult = c(0, 0.05))
    ) +
    labs(
      title = title,
      x = "Number of subjects",
      y = NULL
    ) +
    theme_minimal() +
    theme(
      plot.title = element_text(hjust = 0.5, face = "bold", size = 10),
      legend.position = "none",
      axis.text.y = element_text(face = "bold", size = 8),
      axis.text.x = element_text(size = 8),
      axis.title.x = element_text(size = 9)
    )
}

plot_distribution <- function(
    data,
    var_raw,
    var_label,
    # Density
    fill_density       = "firebrick1",
    color_density      = "firebrick1",
    density_top_shrink = 0.9,
    # Histogram
    fill_hist          = "royalblue1",
    binwidth           = NULL,
    # Axes
    xmin               = NULL,
    xmax               = NULL,
    ymax               = NULL,
    # Percentile lines
    show_percentiles   = FALSE,
    color_percentiles  = "gray30"
) {

  x <- data[[var_raw]]
  x <- as.numeric(x)
  x <- x[!is.na(x)]

  if (length(x) < 2) {
    return(ggplot() + theme_void() + labs(title = var_label))
  }

  # Axis limits
  xmin <- if (!is.null(xmin)) xmin else min(x)
  xmax <- if (!is.null(xmax)) xmax else max(x)
```

```r
# Binwidth
bw <- if (!is.null(binwidth)) binwidth else get_binwidth(var_raw)

# Histogram scaling for density overlay
brks       <- seq(xmin, xmax + bw, by = bw)
hist_max   <- max(hist(x, breaks = brks, plot = FALSE)$counts)
target_max <- hist_max * density_top_shrink

# KDE
kde          <- density(x, bw = stats::bw.nrd0(x))
scale_factor <- ifelse(max(kde$y) > 0, target_max / max(kde$y), 0)

dens_df <- tibble(x = kde$x, y = kde$y * scale_factor) %>%
  dplyr::filter(x >= xmin, x <= xmax)

# Y limit
y_upper <- if (!is.null(ymax)) ymax else hist_max * 1.05

# Base plot
p <- ggplot() +
  geom_area(
    data      = dens_df,
    aes(x = x, y = y),
    fill      = fill_density,
    color     = color_density,
    alpha     = 0.4,
    linewidth = 0.5
  ) +
  geom_histogram(
    data      = tibble(Value = x),
    aes(x = Value, y = after_stat(count)),
    binwidth  = bw,
    fill      = fill_hist,
    color     = "white",
    linewidth = 0.3,
    alpha     = 0.4,
    boundary  = xmin
  ) +
  coord_cartesian(
    xlim   = c(xmin, xmax),
    ylim   = c(0, y_upper),
    expand = FALSE
  ) +
  labs(
    title = NULL,
    x     = var_label,
    y     = "Frequency (n)"
```

```r
    ) +
    theme_minimal() +
    theme(
      plot.title   = element_blank(),
      axis.title.y = element_text(size = 8),
      axis.title.x = element_text(face = "bold", size = 8),
      axis.text.y  = element_text(size = 7),
      axis.text.x  = element_text(size = 7)
    )

  # Optional percentile lines
  if (show_percentiles) {
    percentile_df <- tibble(
      xintercept = quantile(x, c(0.25, 0.50, 0.75), na.rm = TRUE),
      label      = c("Q1", "Mdn", "Q3")
    )
    p <- p +
      geom_vline(
        data      = percentile_df,
        aes(xintercept = xintercept),
        color     = color_percentiles,
        linetype  = "dashed",
        linewidth = 0.5
      ) +
      geom_text(
        data  = percentile_df,
        aes(x = xintercept, y = y_upper * 0.95, label = label),
        color = color_percentiles,
        size  = 2.5,
        hjust = -0.2
      )
  }

  p
}
```

```r
data_emfra <- read_excel("Database EMFRA Complete.xlsx") %>%
  filter(Complete_evaluation == "Yes")

data_emfra_numeric <- data_emfra %>%
  mutate(
    across(all_of(vars_descriptives_raw), ~ as.numeric(.x))
  )
```

##### 1.2.2 Continuous variables

Descriptive results would be provided for continuous variables, including mean, sd, minimum, q1, median, q3, maximum, skewness and kurtosis values. In addition, histogram and density plots would be provided for representing the distribution of frequencies.

```r
continuous_stats <- get_continuous_stats(
  data = data_emfra_numeric,
  vars = vars_descriptives_raw
)

categorical_stats <- get_categorical_descriptives(
  data = data_emfra,
  vars = vars_categorical_raw
)

continuous_descriptives_table <- format_continuous_table(
  desc   = continuous_stats,
  labels = vars_descriptives_labels,
  digits = 2
)

categorical_descriptives_table <- format_categorical_table(
  categorical_df = categorical_stats,
  labels = vars_categorical_labels,
  digits = 2
)
```

Table 2: Descriptive continuous demographic variables

| Variable | Mean ± SD | Min | Q1 | Median | Q3 | Max | Skewness | Kurtosis |
|---|---|---|---|---|---|---|---|---|
| Age (years) | 77.19 ± 9.07 | 65.00 | 69.00 | 77.00 | 84.00 | 99.00 | 0.35 | -1.04 |
| Height (m) | 1.62 ± 0.1 | 1.37 | 1.55 | 1.60 | 1.69 | 1.85 | 0.24 | -0.36 |
| Weight (kg) | 69.66 ± 14.08 | 42.00 | 60.00 | 68.00 | 80.00 | 128.00 | 0.68 | 0.97 |
| Body Mass Index (kg/m²) | 26.57 ± 4.71 | 15.81 | 23.51 | 25.97 | 29.07 | 43.56 | 0.80 | 1.29 |
| Years of formal education | 16.49 ± 5.59 | 0.00 | 13.00 | 16.00 | 20.00 | 31.00 | -0.17 | -0.04 |
| SPPB – Balance score | 2.91 ± 1.38 | 0.00 | 2.00 | 4.00 | 4.00 | 4.00 | -0.81 | -0.82 |
| Gait time in 2.4m (s) | 3.25 ± 1.69 | 1.39 | 2.03 | 2.64 | 3.99 | 11.39 | 1.83 | 4.22 |
| SPPB – Gait speed score | 3.21 ± 1.22 | 0.00 | 3.00 | 4.00 | 4.00 | 4.00 | -1.37 | 0.64 |
| FTSTS (s) | 11.59 ± 4.3 | 5.38 | 8.76 | 10.71 | 13.66 | 33.16 | 1.41 | 3.26 |
| SPPB – FTSTS score | 2.93 ± 1.45 | 0.00 | 2.00 | 4.00 | 4.00 | 4.00 | -1.04 | -0.44 |
| SPPB – Total score | 9.04 ± 3.63 | 0.00 | 7.00 | 11.00 | 12.00 | 12.00 | -1.04 | -0.18 |
| MoCA (raw score) | 20.56 ± 5.89 | 5.00 | 16.00 | 21.00 | 25.00 | 30.00 | -0.35 | -0.84 |
| MoCA (education-adjusted) | 20.78 ± 5.68 | 6.00 | 16.00 | 22.00 | 25.00 | 30.00 | -0.33 | -0.87 |
| Handgrip strength (kg) | 22.26 ± 7.93 | 8.67 | 16.63 | 20.57 | 27.37 | 45.33 | 0.74 | -0.08 |
| Barthel Index – Total score | 90.02 ± 17.84 | 5.00 | 90.00 | 100.00 | 100.00 | 100.00 | -2.47 | 6.33 |
| HADS – Anxiety score | 5.38 ± 3.89 | 0.00 | 2.00 | 5.00 | 7.00 | 19.00 | 0.99 | 0.70 |
| HADS – Depression score | 3.9 ± 3.37 | 0.00 | 1.00 | 3.00 | 6.00 | 16.00 | 1.03 | 0.67 |
| HADS – Total score | 9.28 ± 6.49 | 0.00 | 5.00 | 8.00 | 13.00 | 30.00 | 1.05 | 0.77 |

Table 2: Descriptive continuous demographic variables

| Variable | Mean ± SD | Min | Q1 | Median | Q3 | Max | Skewness | Kurtosis |
|---|---|---|---|---|---|---|---|---|
| EMFRA administration time (min) | 13.12 ± 2.27 | 9.70 | 11.31 | 12.82 | 14.38 | 17.27 | 0.33 | -1.05 |

```
vars_plot_order <- c(
  "Age",
  "Height",
  "Weight",
  "BMI",
  "Years_studied",
  "SPPB_balance_points",
  "SPPB_walk_final_points",
  "SPPB_FTSS_final_points",
  "SPPB_points",
  "SPPB_walk_second_mean",
  "SPPB_FTSS_mean",
  "Handgrip_mean",
  "MoCA_without_studies",
  "MOCA_with_studies",
  "Barthel_total",
  "HADS_anxiety_total",
  "HADS_depression_total",
  "HADS_total"
)

binwidth_default_descriptives <- 1

binwidth_map_descriptives <- c(
  Height                 = 0.02,
  Weight                 = 2,
  SPPB_walk_second_mean  = 0.2,
  SPPB_balance_points    = 0.8,
  SPPB_walk_final_points = 0.8,
  SPPB_FTSS_final_points = 0.8,
  Barthel_total          = 5
)

get_binwidth <- function(var_raw) {
  if (var_raw %in% names(binwidth_map_descriptives)) {
    return(unname(binwidth_map_descriptives[var_raw]))
  }
  binwidth_default_descriptives
}

plots_descriprives_continuous <- lapply(vars_plot_order, function(v) {
  plot_distribution(
```

```r
    data      = data_emfra_numeric,
    var_raw   = v,
    var_label = unname(vars_descriptives_labels[v])
  )
})

gridExtra::grid.arrange(grobs = plots_descriprives_continuous, ncol = 3)
```

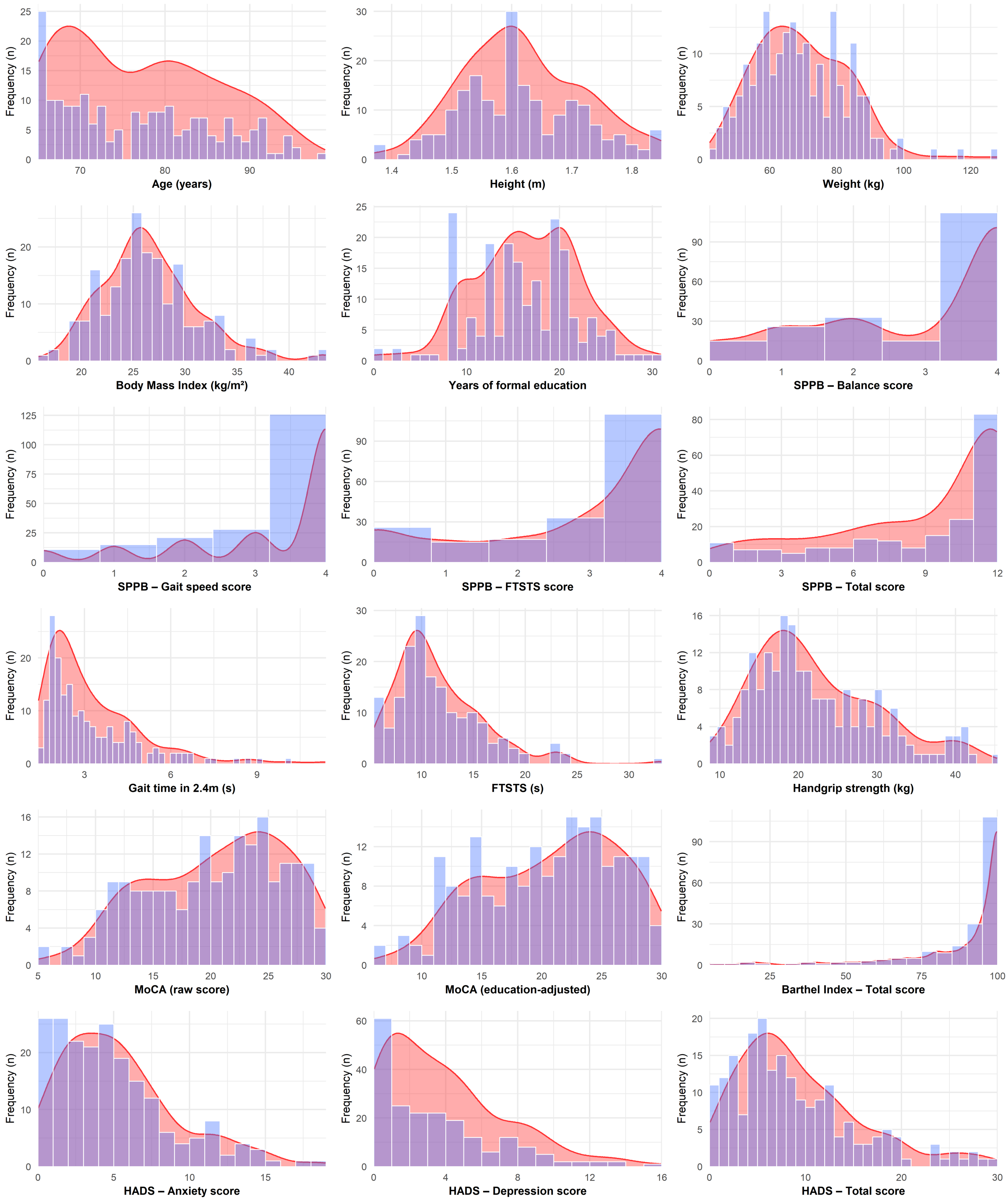

Figure 1: Histogram and density plot of baseline continuous variables

##### 1.2.3 Categorical variables

Descriptive results would be provided for continuous variables, including mean, sd, minimum, q1, median, q3, maximum, skewness and kurtosis values.

Table 3: Descriptive categorical demographic variables

| Variable | Categories |
|---|---|
| Educational level | University (86, 42.79%) \| Elementary school (35, 17.41%) \| High school (28, 13.93%) \| Secondary school (25, 12.44%) \| Professional training (21, 10.45%) \| Preschool education (4, 1.99%) \| Without studies (2, 1%) |
| Laboral status | Retired (169, 84.08%) \| Active (22, 10.95%) \| Unpaid domestic worker (6, 2.99%) \| Unemployed (4, 1.99%) |
| Marital status | Married (83, 41.29%) \| Widowed (80, 39.8%) \| Divorced (19, 9.45%) \| Single (18, 8.96%) \| Common-law partnership (1, 0.5%) |
| Sex | Female (132, 65.67%) \| Male (69, 34.33%) |

```r
p_sex <- plot_donut_thin(
  data  = data_emfra,
  var   = "Sex",
  title = "Sex",
  colors = c(
    "Male"   = "slateblue3",
    "Female" = "darkseagreen3"
  )
)

p_education <- plot_donut_thin(
  data  = data_emfra,
  var   = "Educational_level",
  title = "Educational level",
  palette = "ggthemes::Tableau_20"
)

p_marital <- plot_donut_thin(
  data  = data_emfra,
  var   = "Marital_status",
  title = "Marital status",
  palette = "ggthemes::Tableau_20"
)

p_laboral <- plot_donut_thin(
  data  = data_emfra,,
  var   = "Laboral_status",
  title = "Laboral status",
  palette = "ggthemes::Tableau_20"
)

grid.arrange(
```

```
  grobs = list(
    p_sex,
    p_education,
    nullGrob(),
    p_marital,
    p_laboral,
    nullGrob()
  ),
  ncols = 3,
  nrows = 2,
  widths  = c(1, 1, 0.1),
  heights = c(1, 1)
)
```

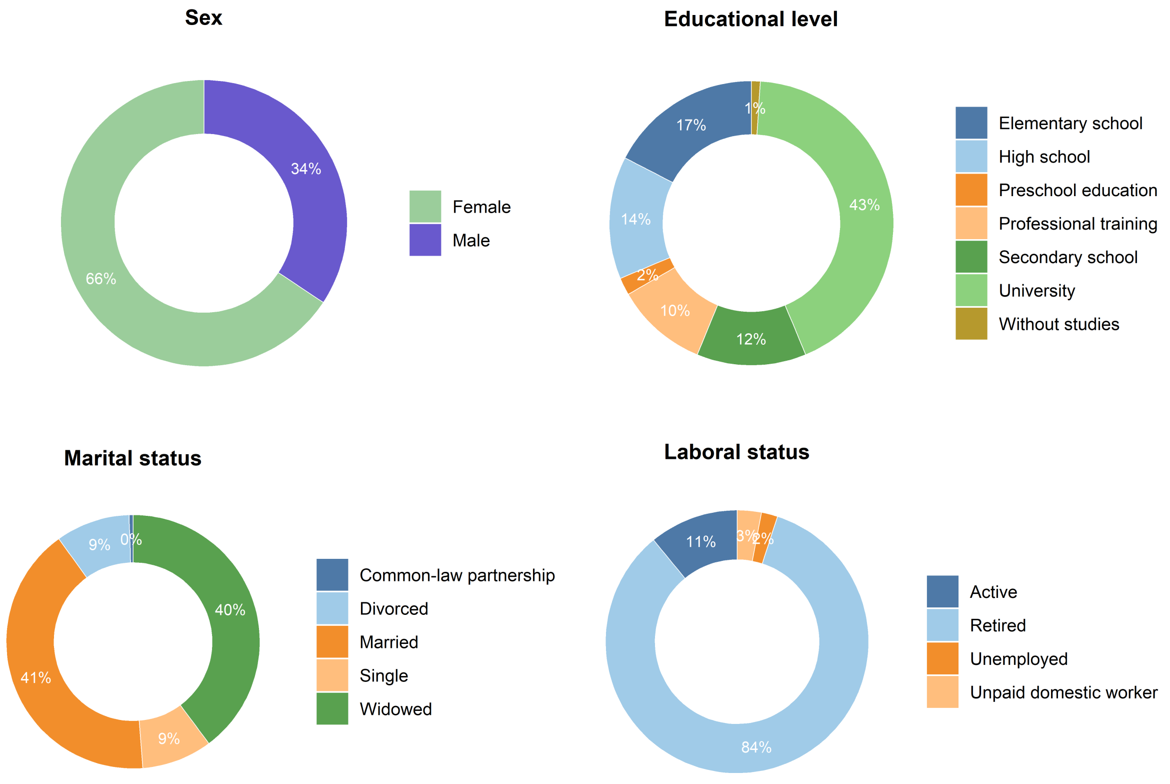

Figure 2: Distribution of categorical descriptive variables

```
msk_location_map <- c(

  # --- Systemic ---
  "Osteoporosis"                     = "Systemic",
  "Osteoarthritis (unspecified)"     = "Systemic",
  "Rheumatoid arthritis"             = "Systemic",
```

```
"Rheumatic polymyalgia"           = "Systemic",

# --- Low back ---
"Low back pain"                   = "Low back",
"Lumbar arthrodesis"              = "Low back",
"Lumbar stenosis"                 = "Low back",
"Lumbar osteoarthritis"           = "Low back",
"Lumbar radiculopathy"            = "Low back",
"Lumbar surgery"                  = "Low back",
"Lumbar arthrodesis lumbar"       = "Low back",
"Lumbar disc herniation"           = "Low back",
"Lumbar fracture"                 = "Low back",
"Lumbar radicular pain"            = "Low back",
"Lumbar disc surgery"              = "Low back",
"Lumbar discopathy"               = "Low back",
"Lumbar discoplasty"              = "Low back",
"Lumbar epifisitis"               = "Low back",
"Lumbar laminectomy"              = "Low back",
"Lumbar osteoporosis"             = "Low back",
"Lumbar osteosynthesis"            = "Low back",
"Tarlov cyst"                     = "Low back",

# --- Neck ---
"Neck pain"                       = "Neck",
"Cervical arthrodesis"             = "Neck",
"Cervical surgery"                 = "Neck",
"Cervical osteoarthritis"          = "Neck",

# --- Thoracic ---
"Dorsal pain"                      = "Thoracic",
"Severe cervicothoracic kyphosis"  = "Thoracic",
"Scoliosis"                        = "Thoracic",

# --- Shoulder ---
"Shoulder pain"                    = "Shoulder",
"Rotator cuff tear"                = "Shoulder",
"Rotator cuff massive tear"        = "Shoulder",
"Rotator cuff suture"              = "Shoulder",
"Rotator cuff surgery"             = "Shoulder",
"Shoulder tendinopathy"             = "Shoulder",
"Adhesive capsulitis"               = "Shoulder",
"Shoulder arthroplasty"             = "Shoulder",
"Shoulder surgery"                  = "Shoulder",
"Shoulder instability surgery"     = "Shoulder",
"Humeral fracture"                  = "Shoulder",
"Humeral osteosynthesis"            = "Shoulder",
"Clavicule fracture"                = "Shoulder",
"Clavicule osteosynthesis"          = "Shoulder",
```

```
# --- Hip ---
"Hip pain"                          = "Hip",
"Hip osteoarthritis"                = "Hip",
"Hip arthroplasty"                  = "Hip",
"Hip fracture"                      = "Hip",
"Femoral neck osteosynthesis"       = "Hip",
"Hip osteoporosis"                  = "Hip",

# --- Lumbo-pelvic ---
"Coxis fracture"                    = "Lumbo-pelvic",
"Gluteal pain"                      = "Lumbo-pelvic",
"Pelvic osteoarthritis"             = "Lumbo-pelvic",

# --- Knee ---
"Knee osteoarthritis"               = "Knee",
"Knee pain"                         = "Knee",
"Knee arthroplasty"                 = "Knee",
"Meniscal tear"                     = "Knee",
"Meniscal surgery"                  = "Knee",
"Meniscectomy"                      = "Knee",
"Meniscal suture"                   = "Knee",
"Knee sprain"                       = "Knee",
"Knee surgery"                      = "Knee",
"Patellofemoral contropathy"        = "Knee",

# --- Elbow ---
"Elbow fracture"                    = "Elbow",

# --- Wrist ---
"Wrist pain"                        = "Wrist",
"Wrist fracture"                    = "Wrist",
"Wrist sprain"                      = "Wrist",

# --- Hand ---
"Hand osteoarthritis"               = "Hand",
"Finger osteoarthritis"             = "Hand",
"Finger arthrosis"                  = "Hand",
"Trapeziometacarpal osteoarthritis" = "Hand",
"Hand edema"                        = "Hand",
"Carpal tunnel syndrome"            = "Hand",
"Dupuytren disease"                 = "Hand",

# --- Thigh ---
"Thigh pain"                        = "Thigh",
"Leg pain"                          = "Thigh",

# --- Calf ---
```

```r
  "Calf atrophy"                       = "Calf",
  "Tibial fracture"                    = "Calf",
  "Peroneal fracture"                  = "Calf",

  # --- Ankle ---
  "Ankle arthrodesis"                  = "Ankle",
  "Ankle osteoarthritis"               = "Ankle",

  # --- Foot ---
  "Bunionectomy"                       = "Foot",
  "Bunion"                             = "Foot",
  "Hammertoe surgery"                  = "Foot",
  "Hammertoes"                         = "Foot",
  "Plantar fascitis"                   = "Foot",
  "Aquiles tendon surgery"             = "Foot",

  # --- Face ---
  "Facial fracture"                    = "Facial",
  "Parietal fracture"                  = "Facial",
  "Headache"                           = "Facial"
)

msk_res <- compute_affection_and_types(
  data = data_emfra,
  var_raw = "Msk_pathology_renamed",
  affection_name = "Msk_pathology_affection",
  recode_map = msk_location_map
)

p_msk_affection <- plot_affection_yes_no(
  msk_res$affection_summary,
  msk_res$n_total,
  "Musculoskeletal affection",
  yes_color = "darkgoldenrod1"
)

p_msk_location <- plot_type_histogram(
  type_df = msk_res$type_summary,
  n_total = msk_res$n_total,
  title   = "Musculoskeletal location",
  palette = rev(paletteer::paletteer_c("grDevices::Inferno", 100))
)
```

```r
choir_region_ids <- list(
  Knee = c("231","232"),
  `Low back` = c("218","219"),
  Shoulder = c("207","210"),
```

```r
  Hip = c("222","225"),
  Neck = c("205","206"),
  Thoracic = c("208","209","212","213"),
  Elbow = c("215","216"),
  Wrist = c("221","226"),
  Hand = c("227","230"),
  Thigh = c("228","229"),
  Calf = c("233","234"),
  Ankle = c("235","236"),
  Foot = c("237","238"),
  Facial = c("201","202","203","204"),
  `Lumbo-pelvic` = c("223","224")
  )

base_map <- gen_example_data() %>%
  filter(as.numeric(id) >= 200) %>%
  select(id) %>%
  distinct() %>%
  mutate(
    prevalence = 0,
    group = "Posterior"
  )

prevalence_values <- msk_res$type_summary %>%
  filter(!type %in% c("Systemic", "No")) %>%
  mutate(
    prevalence = percent / 100,
    id = choir_region_ids[type]
  ) %>%
  unnest(id) %>%
  mutate(id = as.character(id)) %>%
  select(id, prevalence)

plot_df <- base_map %>%
  left_join(prevalence_values, by = "id") %>%
  mutate(
    prevalence = ifelse(is.na(prevalence.y), prevalence.x, prevalence.y)
  ) %>%
  select(id, prevalence, group)

msk_choirbm_plot <- plot_male_choirbm(
  df    = plot_df,
  value = "prevalence"
) +
  scale_fill_gradientn(
```

```r
    colours = rev(paletteer_c("grDevices::Inferno", 100)),
    limits  = c(0, 1),
    name    = "Prevalence"
  ) +
  geom_polygon(
    aes(group = id),
    fill   = NA,
    colour = "grey85",
    linewidth = 1
  ) +
  facet_wrap(
    ~ group,
    labeller = labeller(
      group = c(Posterior = "Musculoskeletal location")
    )
  ) +
  theme_minimal() +
  theme(
    axis.title = element_blank(),
    axis.text  = element_blank(),
    axis.ticks = element_blank(),
    legend.position = "right",
    strip.text.x.top = element_text(face = "bold", size = 10)
  )

layout_mat <- rbind(
  c(1, 3),
  c(2, 3)
)

grid.arrange(
  grobs = list(
    p_msk_affection,
    p_msk_location,
    msk_choirbm_plot
  ),
  layout_matrix = layout_mat,
  widths  = c(1, 0.7),
  heights = c(0.4, 1)
)
```

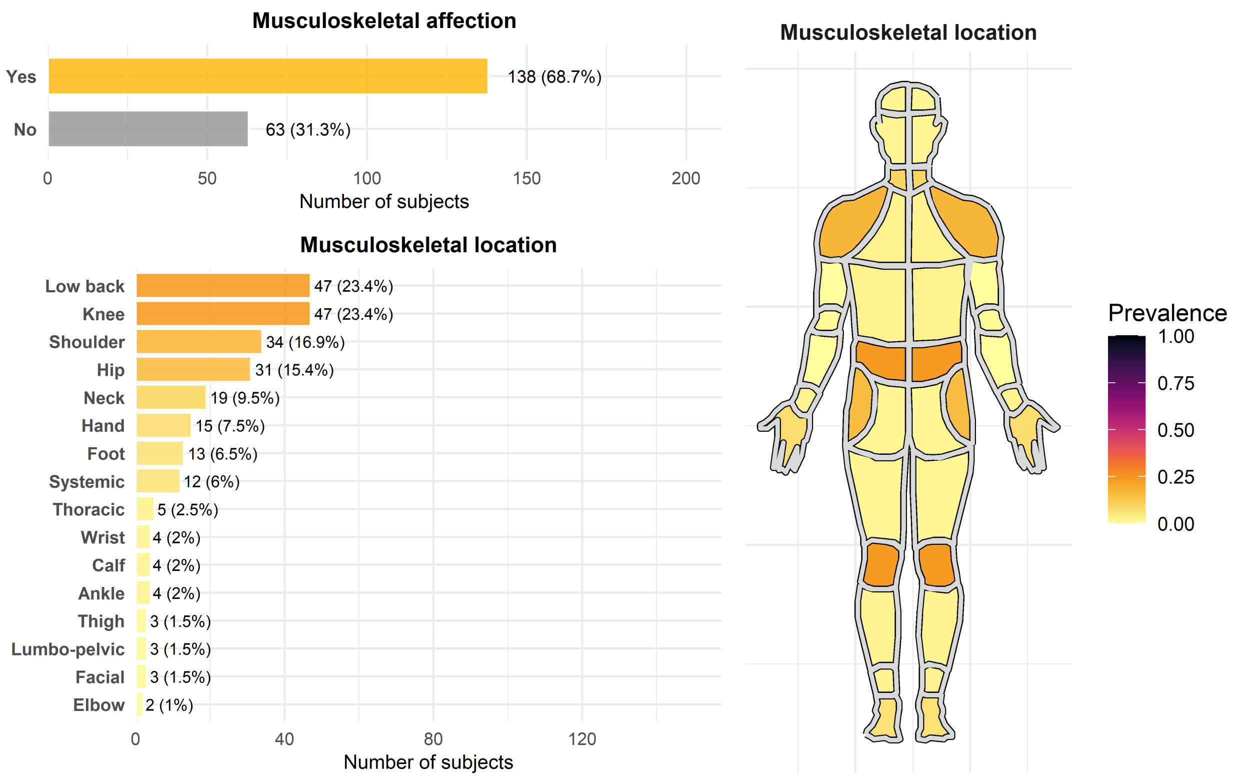

Figure 3: Distribution of Musculoskeletal Conditions

```r
neuro_res <- compute_affection_and_types(
  data = data_emfra,
  var_raw = "Neurological_pathology_renamed",
  affection_name = "Neurological_pathology_affection"
)

p_neuro_affection <- plot_affection_yes_no(
  neuro_res$affection_summary,
  neuro_res$n_total,
  "Neurological affection",
  yes_color = "steelblue2"
)

p_neuro_type <- plot_type_histogram(
  neuro_res$type_summary,
  neuro_res$n_total,
  "Neurological pathology type",
  fill_color = "steelblue2"
)

grid.arrange(
  grobs = list(
    p_neuro_affection,
```

```r
    p_neuro_type
    ),
  nrows = 2,
  heights = c(0.4, 1)
)
```

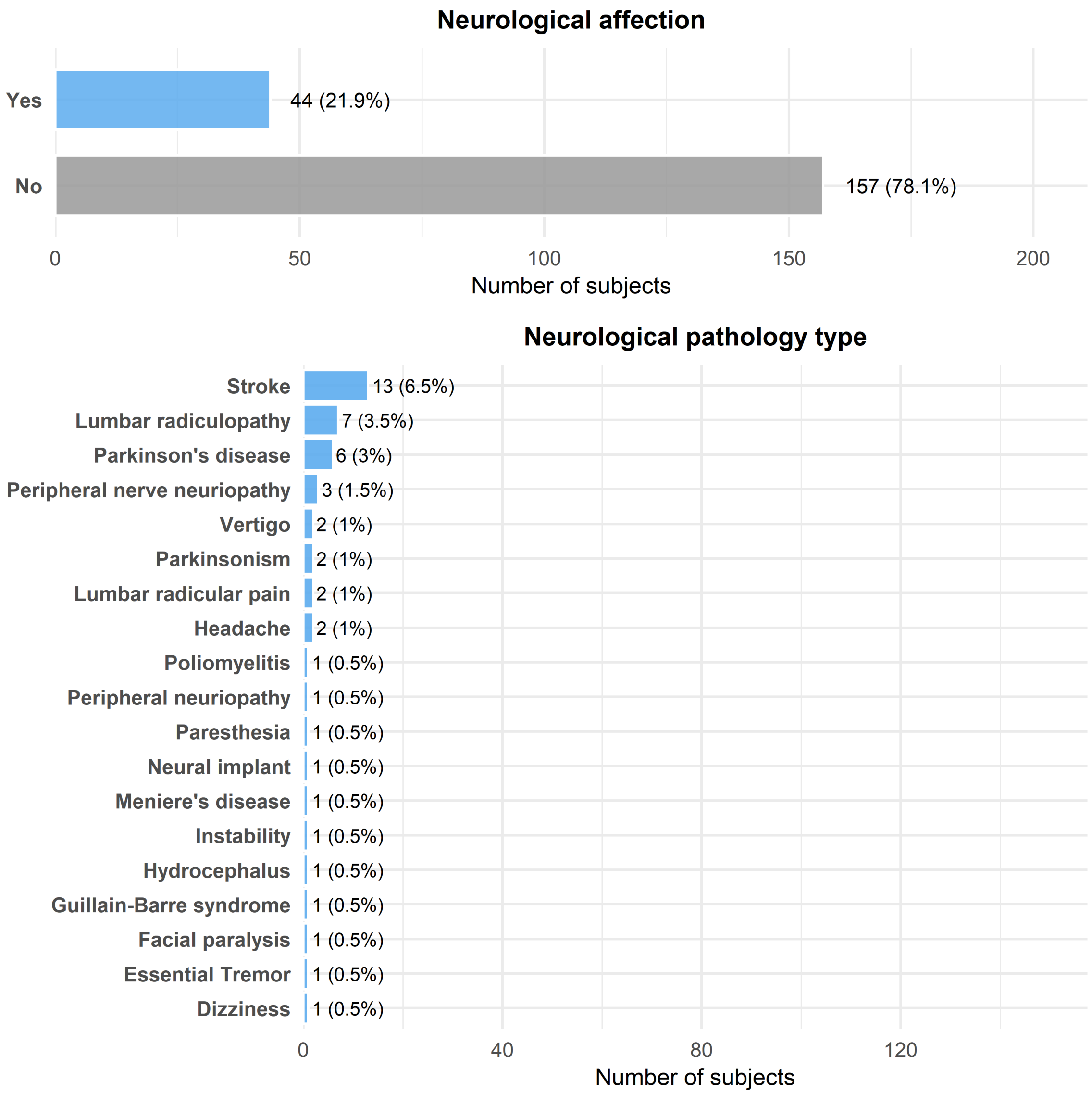

Figure 4: Distribution of Neurological Pathologies

```r
psych_res <- compute_affection_and_types(
  data = data_emfra,
  var_raw = "Psych_pathology_renamed",
  affection_name = "Psych_pathology_affection"
)
```

```r
p_psych_affection <- plot_affection_yes_no(
  psych_res$affection_summary,
  psych_res$n_total,
  "Psychological affection",
  yes_color = "mediumorchid1"
)

p_psych_type <- plot_type_histogram(
  psych_res$type_summary,
  psych_res$n_total,
  "Psychological pathology type",
  fill_color = "mediumorchid1"
)

grid.arrange(
  grobs = list(
    p_psych_affection,
    p_psych_type
    ),
  nrows = 2,
  heights = c(0.8, 1)
)
```

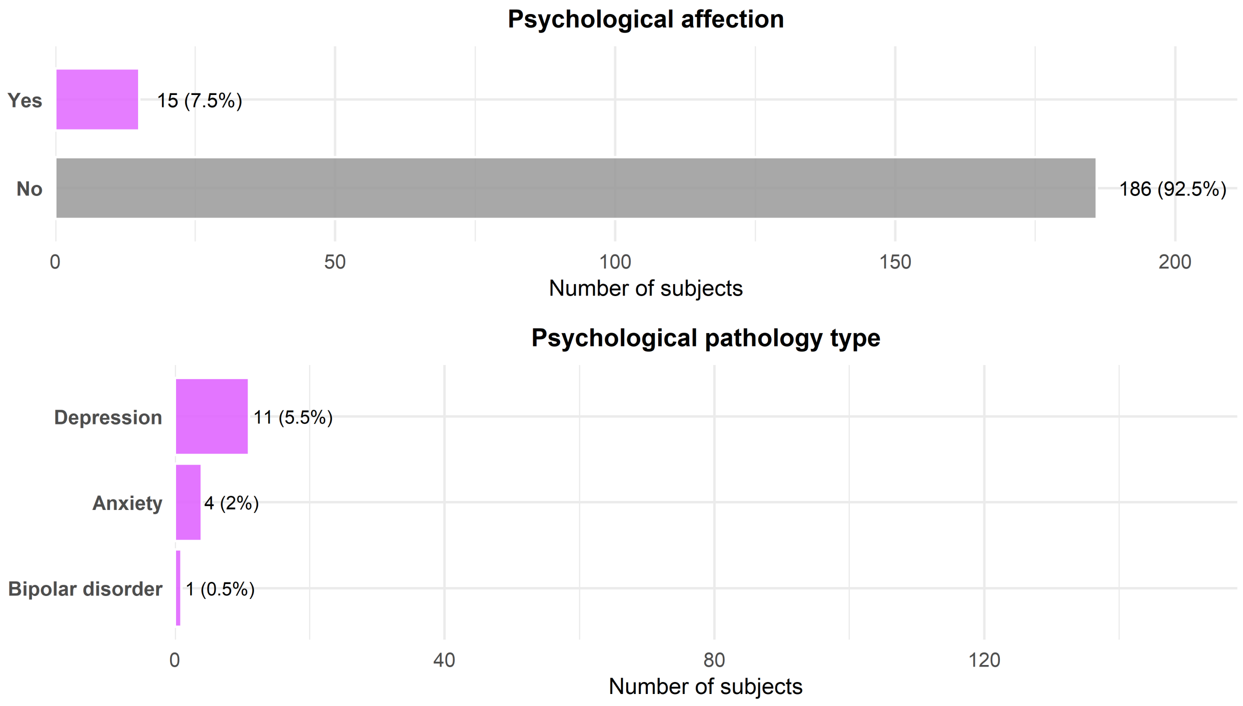

Figure 5: Distribution of Psychological Affections

```r
metabo_res <- compute_affection_and_types(
  data = data_emfra,
  var_raw = "Metabo_pathology_renamed",
  affection_name = "Metabo_pathology_affection"
)

p_metabo_affection <- plot_affection_yes_no(
  metabo_res$affection_summary,
  metabo_res$n_total,
  "Metabolic affection",
  yes_color = "palegreen3"
)

p_metabo_type <- plot_type_histogram(
  metabo_res$type_summary,
  metabo_res$n_total,
  "Metabolic pathology type",
  fill_color = "palegreen3"
)

grid.arrange(
  grobs = list(
    p_metabo_affection,
    p_metabo_type
    ),
  nrows = 2,
  heights = c(0.6, 1)
)
```

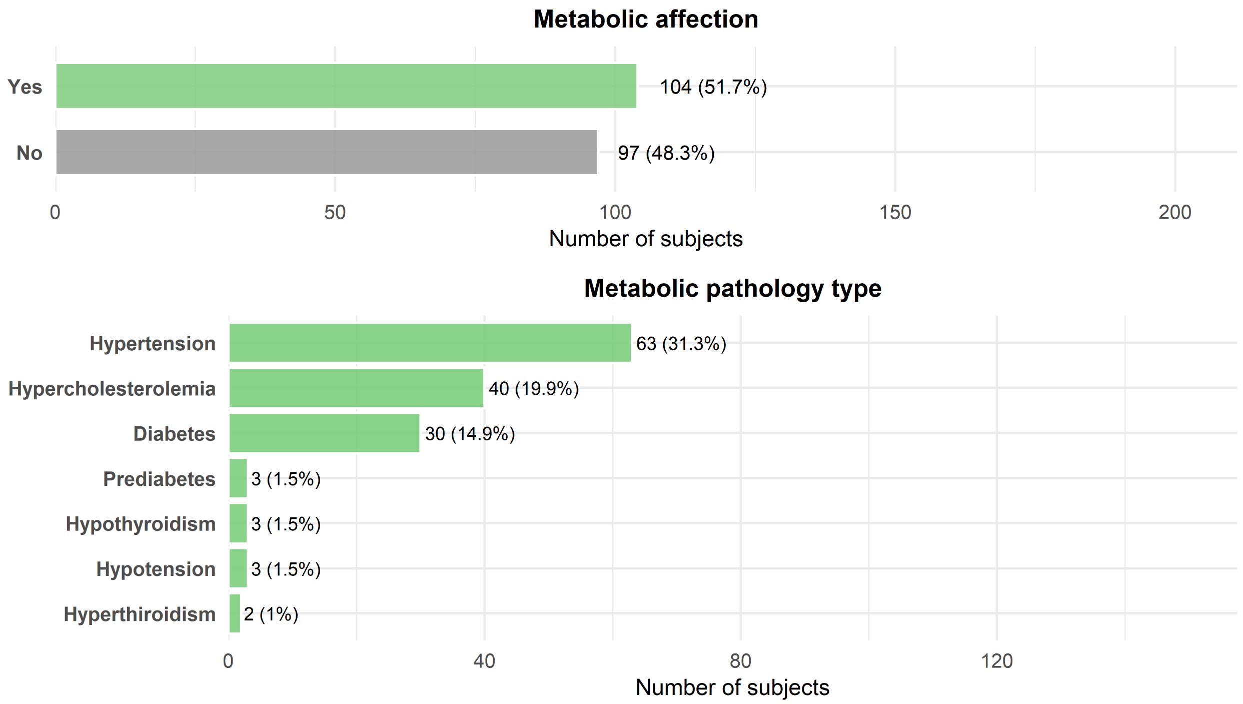

Figure 6: Distribution of Metabolic Affections

```
cancer_terms <- c(
  "Colon cancer",
  "Stomach cancer",
  "Kidney cancer",
  "Prostate cancer",
  "Ovarian cancer",
  "Uterine cancer",
  "Mamarian cancer",
  "Skin cancer",
  "Meningioma"
)

other_group_map <- c(

  # --- Cardiac disorders ---
  "Angina pectoris"             = "Cardiac disorders",
  "Atrial fibrilation"          = "Cardiac disorders",
  "Cardiac arrhythmia"          = "Cardiac disorders",
  "Cardiac arrythmia"           = "Cardiac disorders",
  "Cardiac valve disease"       = "Cardiac disorders",
  "Heart disease"               = "Cardiac disorders",
  "Heart failure"               = "Cardiac disorders",
  "Hypertensive cardiomyopathy"= "Cardiac disorders",
  "Ischemic cardiopathy"        = "Cardiac disorders",
  "Ishcemic cardiopathy"        = "Cardiac disorders",
```

```
"Myocardial infarction"      = "Cardiac disorders",
"STEM"                       = "Cardiac disorders",

# --- Cardiac surgery ---
"Cardiac surgery"            = "Cardiac surgery",
"Cardiac valve surgery"      = "Cardiac surgery",

 # --- Vascular disorders ---
"Anemia"                     = "Vascular disorders",

# --- Vascular surgery ---
"Carotid bypass"             = "Vascular surgery",
"Varicose vein surgery"      = "Vascular surgery",

# --- Respiratory disorders ---
"Asthma"                     = "Respiratory disorders",
"Bronchiectasis"             = "Respiratory disorders",
"COPD"                       = "Respiratory disorders",
"Pneumonia"                  = "Respiratory disorders",
"Pulmonary edema"            = "Respiratory disorders",

# --- Respiratory surgery ---
"Pulmonary surgery"          = "Respiratory surgery",

# --- Gastrointestinal disorders ---
"Gastritis"                  = "Gastrointestinal disorders",
"Hiatal hernia"              = "Gastrointestinal disorders",
"Diverticulos"               = "Gastrointestinal disorders",
"Irritable bowel"            = "Gastrointestinal disorders",
"Pancreatitis"               = "Gastrointestinal disorders",

# --- Gastrointestinal surgery ---
"Appendix surgery"           = "Gastrointestinal surgery",
"Gallbladder surgery"        = "Gastrointestinal surgery",
"Diverticulitis surgery"     = "Gastrointestinal surgery",
"Anal fistula surgery"       = "Gastrointestinal surgery",
"Pilonidal fistula surgery"  = "Gastrointestinal surgery",
"Intesitine surgery"         = "Gastrointestinal surgery",
"Inguinal hernia surgery"    = "Gastrointestinal surgery",

# --- Urinary disorders ---
"Kidney stones"              = "Urinary disorders",
"Chronic renal disease"      = "Urinary disorders",
"Kidney failure"             = "Urinary disorders",
"Bladder diverticulum"       = "Urinary disorders",
"Bladder prolapse"           = "Urinary disorders",

# --- Urinary surgery ---
```

```r
  "Bladder surgery"             = "Urinary surgery",

  # --- Urologic surgery ---
  "Prostate surgery"            = "Urologic surgery",
  "Hydrocele surgery"           = "Urologic surgery",

  # --- Gynecological disorders ---
  "Polycystic ovary"             = "Ovarian disorders",
  "Uterine prolapse"             = "Uterine disorders",

  # --- Gynecological surgery ---
  "Uterine surgery"              = "Uterine surgery",
  "Caesarean surgery"            = "Uterine surgery",
  "Ovarian removal"              = "Ovarian surgery",
  "Ovarian surgery"              = "Ovarian surgery",

  # --- Endocrine surgery ---
  "Thyroid surgery"              = "Endocrine surgery",
  "Thyroglossal duct surgery"    = "Endocrine surgery",

  # --- Sensory disorders ---
  "Blindness"                     = "Sensory disorders",
  "Visual loss"                   = "Sensory disorders",
  "Eye stroke"                    = "Sensory disorders",
  "Hearing loss"                  = "Sensory disorders",
  "Otosclerosis"                  = "Sensory disorders",
  "Cochlear implant"              = "Sensory disorders",

  # --- Autoimmune / systemic ---
  "Lupus erythematosus"           = "Autoimmune disorders",
  "Psoriasic arthritis"           = "Autoimmune disorders",
  "Autoimmune lymphoproliferative syndrome" = "Autoimmune disorders",

  # --- Other surgery ---
  "Inguinal ganglion surgery"     = "Other surgery"
)

other_res <- compute_affection_and_types(
  data = data_emfra,
  var_raw = "Other_pathology_renamed",
  affection_name = "Other_pathology_affection",
  recode_map = other_group_map,
  cancer_terms = cancer_terms
)

p_other_affection <- plot_affection_yes_no(
  other_res$affection_summary,
  other_res$n_total,
```

```r
  "Other pathology affection",
  yes_color = "thistle3"
)

p_other_type <- plot_type_histogram(
  other_res$type_summary,
  other_res$n_total,
  "Other pathology groups",
  fill_color = "thistle3"
)

grid.arrange(
  grobs = list(
    p_other_affection,
    p_other_type
    ),
  nrows = 2,
  heights = c(0.25, 1)
)
```

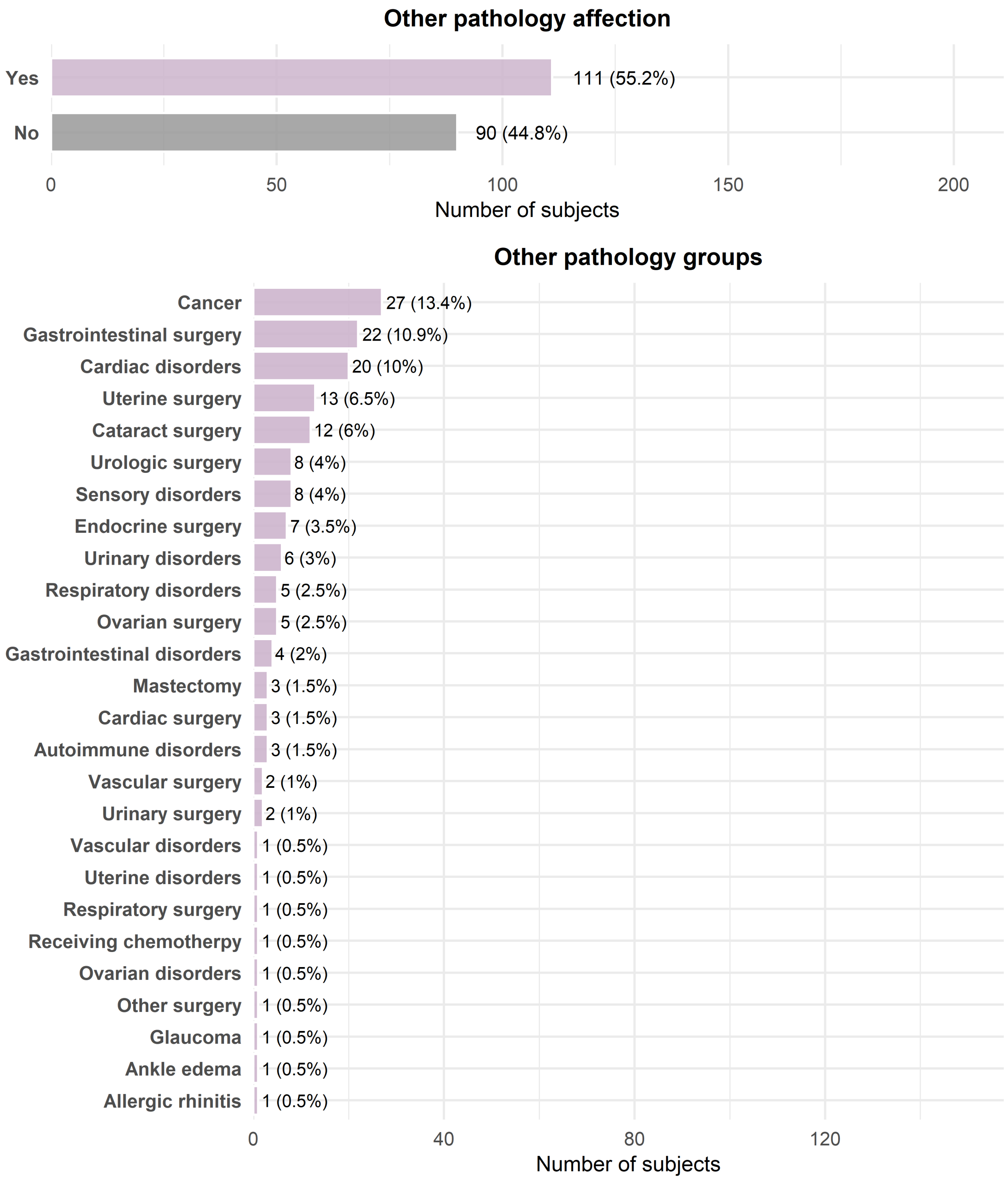

Figure 7: Distribution of Other Affections

### 2 EMFRA item-level descriptive statistics

The absolute (n) and relative frequencies (%) were calculated for the three ordinal response categories (0, 1 and 2), alongside the mean and SD, minimum, Q1, median, Q3, maximum, skewness and kurtosis.

```r
emfra_items <- data_emfra %>%
  select(all_of(emfra_item_ids))

desc_stats <- psych::describe(
  emfra_items,
  na.rm  = TRUE,
  skew   = TRUE,
  ranges = TRUE,
  quant  = c(.25, .50, .75)
) %>%
  as.data.frame() %>%
  tibble::rownames_to_column("item_id")

freq_long <- emfra_items %>%
  pivot_longer(
    cols      = everything(),
    names_to  = "item_id",
    values_to = "response"
  ) %>%
  mutate(response = factor(response, levels = c(0, 1, 2))) %>%
  group_by(item_id, response) %>%
  summarise(n = n(), .groups = "drop_last") %>%
  mutate(
    total_n = sum(n),
    pct     = round(n / total_n * 100, 2),
    n_pct   = paste0(n, " (", pct, "%)")
  ) %>%
  ungroup()

freq_wide <- freq_long %>%
  select(item_id, response, n_pct) %>%
  pivot_wider(
    names_from   = response,
    values_from = n_pct,
    names_prefix = "Response "
  )

EMFRA_descriptives_df <- desc_stats %>%
  left_join(freq_wide, by = "item_id") %>%
  mutate(
    Item = factor(
      item_id,
      levels = emfra_item_ids,
      labels = emfra_item_labels
    ),
    `Mean ± SD` = paste0(
      round(mean, 2), " ± ", round(sd, 2)
    )
```

```r
) %>%
select(
  Item,
  `Response 0`,
  `Response 1`,
  `Response 2`,
  `Mean ± SD`,
  Min      = min,
  Q1       = Q0.25,
  Median   = median,
  Q3       = Q0.75,
  Max      = max,
  Skewness = skew,
  Kurtosis = kurtosis
)
```

Table 4: Descriptive statistics and response distribution for EMFRA items

| Item | Response 0 | Response 1 | Response 2 | Mean ± SD | Min | Q1 | Median | Q3 | Max | Skewness | Kurtosis |
|---|---|---|---|---|---|---|---|---|---|---|---|
| Item 1. Hand-grip strength | 67 (33.33%) | 83 (41.29%) | 51 (25.37%) | 0.92 ± 0.76 | 0 | 0 | 1 | 2 | 2 | 0.13 | -1.29 |
| Item 2. Gait speed | 10 (4.98%) | 71 (35.32%) | 120 (59.7%) | 1.55 ± 0.59 | 0 | 1 | 2 | 2 | 2 | -0.91 | -0.19 |
| Item 3. Gait assistance | 124 (61.69%) | 17 (8.46%) | 60 (29.85%) | 0.68 ± 0.9 | 0 | 0 | 0 | 2 | 2 | 0.66 | -1.45 |
| Item 4. Balance loss | 103 (51.24%) | 78 (38.81%) | 20 (9.95%) | 0.59 ± 0.67 | 0 | 0 | 0 | 1 | 2 | 0.69 | -0.62 |
| Item 5. Fatigue | 57 (28.36%) | 116 (57.71%) | 28 (13.93%) | 0.86 ± 0.64 | 0 | 0 | 1 | 1 | 2 | 0.13 | -0.60 |
| Item 6. Arithmetics for money | 32 (15.92%) | 59 (29.35%) | 110 (54.73%) | 1.39 ± 0.75 | 0 | 1 | 2 | 2 | 2 | -0.76 | -0.84 |
| Item 7. Attention-inhibition | 10 (4.98%) | 37 (18.41%) | 154 (76.62%) | 1.72 ± 0.55 | 0 | 2 | 2 | 2 | 2 | -1.80 | 2.24 |
| Item 8. Memory difficulty | 71 (35.32%) | 116 (57.71%) | 14 (6.97%) | 0.72 ± 0.59 | 0 | 0 | 1 | 1 | 2 | 0.15 | -0.59 |
| Item 9. Concentration difficulty | 100 (49.75%) | 89 (44.28%) | 12 (5.97%) | 0.56 ± 0.61 | 0 | 0 | 1 | 1 | 2 | 0.57 | -0.62 |
| Item 10. Cognitive activities | 17 (8.46%) | 51 (25.37%) | 133 (66.17%) | 1.58 ± 0.64 | 0 | 1 | 2 | 2 | 2 | -1.24 | 0.32 |
| Item 11. Loneliness | 115 (57.21%) | 73 (36.32%) | 13 (6.47%) | 0.49 ± 0.62 | 0 | 0 | 0 | 1 | 2 | 0.85 | -0.30 |
| Item 12. Sadness | 70 (34.83%) | 117 (58.21%) | 14 (6.97%) | 0.72 ± 0.58 | 0 | 0 | 1 | 1 | 2 | 0.14 | -0.57 |
| Item 13. Irritability | 79 (39.3%) | 104 (51.74%) | 18 (8.96%) | 0.7 ± 0.63 | 0 | 0 | 1 | 1 | 2 | 0.33 | -0.69 |
| Item 14. Self-efficacy difficulty | 108 (53.73%) | 78 (38.81%) | 15 (7.46%) | 0.54 ± 0.63 | 0 | 0 | 0 | 1 | 2 | 0.75 | -0.47 |
| Item 15. Life satisfaction | 15 (7.46%) | 73 (36.32%) | 113 (56.22%) | 1.49 ± 0.63 | 0 | 1 | 2 | 2 | 2 | -0.83 | -0.36 |
| Item 16. Economical difficulty | 190 (94.53%) | 6 (2.99%) | 5 (2.49%) | 0.08 ± 0.35 | 0 | 0 | 0 | 0 | 2 | 4.58 | 20.54 |
| Item 17. Family interaction | 4 (1.99%) | 50 (24.88%) | 147 (73.13%) | 1.71 ± 0.5 | 0 | 1 | 2 | 2 | 2 | -1.41 | 0.95 |
| Item 18. Social interaction | 32 (15.92%) | 77 (38.31%) | 92 (45.77%) | 1.3 ± 0.73 | 0 | 1 | 1 | 2 | 2 | -0.52 | -0.99 |
| Item 19. Social-ludic activities | 72 (35.82%) | 63 (31.34%) | 66 (32.84%) | 0.97 ± 0.83 | 0 | 0 | 1 | 2 | 2 | 0.06 | -1.55 |
| Item 20. Communication difficulty | 174 (86.57%) | 22 (10.95%) | 5 (2.49%) | 0.16 ± 0.43 | 0 | 0 | 0 | 0 | 2 | 2.73 | 7.01 |
| Item 21. Help availability | 7 (3.48%) | 33 (16.42%) | 161 (80.1%) | 1.77 ± 0.5 | 0 | 2 | 2 | 2 | 2 | -2.04 | 3.36 |

```
EMFRA_long2 <- data_emfra %>%
  select(all_of(emfra_item_ids)) %>%
  pivot_longer(
    cols = everything(),
    names_to = "item_id",
    values_to = "response"
  )

EMFRA_freq2 <- EMFRA_long2 %>%
  group_by(item_id, response) %>%
  summarise(n = n(), .groups = "drop") %>%
  group_by(item_id) %>%
  mutate(percent = n / sum(n) * 100) %>%
  ungroup()

EMFRA_freq2$item_label <- factor(
  EMFRA_freq2$item_id,
  levels = emfra_item_ids,
  labels = emfra_item_labels
)

EMFRA_freq2$response <- factor(
  EMFRA_freq2$response,
  levels = c(0, 1, 2)
)

EMFRA_freq2 <- EMFRA_freq2 %>%
  arrange(item_label, response)

p_bars_EMFRA <- ggplot(
  EMFRA_freq2,
  aes(x = percent, y = item_label, fill = response)
) +
  geom_bar(
    stat = "identity",
    position = position_fill(reverse = TRUE),
    color = "white",
    linewidth = 0.2
  ) +
  geom_text(
    aes(label = paste0(round(percent, 0))),
    color = "white",
    size = 3,
    fontface = "bold",
    position = position_fill(vjust = 0.5, reverse = TRUE)
  ) +
  scale_fill_manual(
    values = rev(paletteer::paletteer_c("grDevices::Emrld",3)),
```

```
    name = "Response"
  ) +
  scale_x_continuous(
    labels = scales::percent_format(scale = 100),
    expand = c(0, 0)
  ) +
  scale_y_discrete(
    limits = rev(levels(EMFRA_freq2$item_label))
  ) +
  labs(
    x = "Percentage of responses",
    y = "EMFRA Items",
  ) +
  theme_minimal() +
  theme(
    legend.position = "top",
    axis.title = element_text(size = 12),
    axis.text  = element_text(size = 10),
  )

EMFRA_long2 <- EMFRA_long2 %>%
  filter(!is.na(response)) %>%
  mutate(
    response_num = as.numeric(response),
    item_label = factor(
      item_id,
      levels = emfra_item_ids,
      labels = emfra_item_labels
    )
  )

p_ridges_EMFRA <- ggplot(
  EMFRA_long2,
  aes(x = response_num, y = item_label)
) +
  geom_density_ridges_gradient(
    aes(fill = after_stat(x)),
    scale = 2,
    rel_min_height = 0.001,
    color = "white",
    linewidth = 0.3
  ) +
  scale_x_continuous(
    breaks = c(0, 1, 2),
    limits = c(-0.3, 2.3),
    expand = c(0, 0)
  ) +
  scale_fill_gradientn(
```

```r
    colours = rev(paletteer::paletteer_c("grDevices::Emrld",100)),
    limits = c(-0.3, 2.3)
  ) +
  scale_y_discrete(
    limits = rev(levels(EMFRA_long2$item_label))
  ) +
  labs(
    x = "Density of responses",
    y = NULL
  ) +
  theme_void() +
  theme(
    legend.position = "none",
    axis.text.x = element_text(size = 10, vjust = 1.5),
    axis.title.x = element_text(size = 12),
    plot.margin = margin(t = 47, r = 0, b = 4, l = 0)
  )

gridExtra::grid.arrange(
  p_bars_EMFRA,
  grid::nullGrob(),
  p_ridges_EMFRA,
  grid::nullGrob(),
  ncol = 4,
  widths = c(2, 0.05, 1, 0.2)
)
```

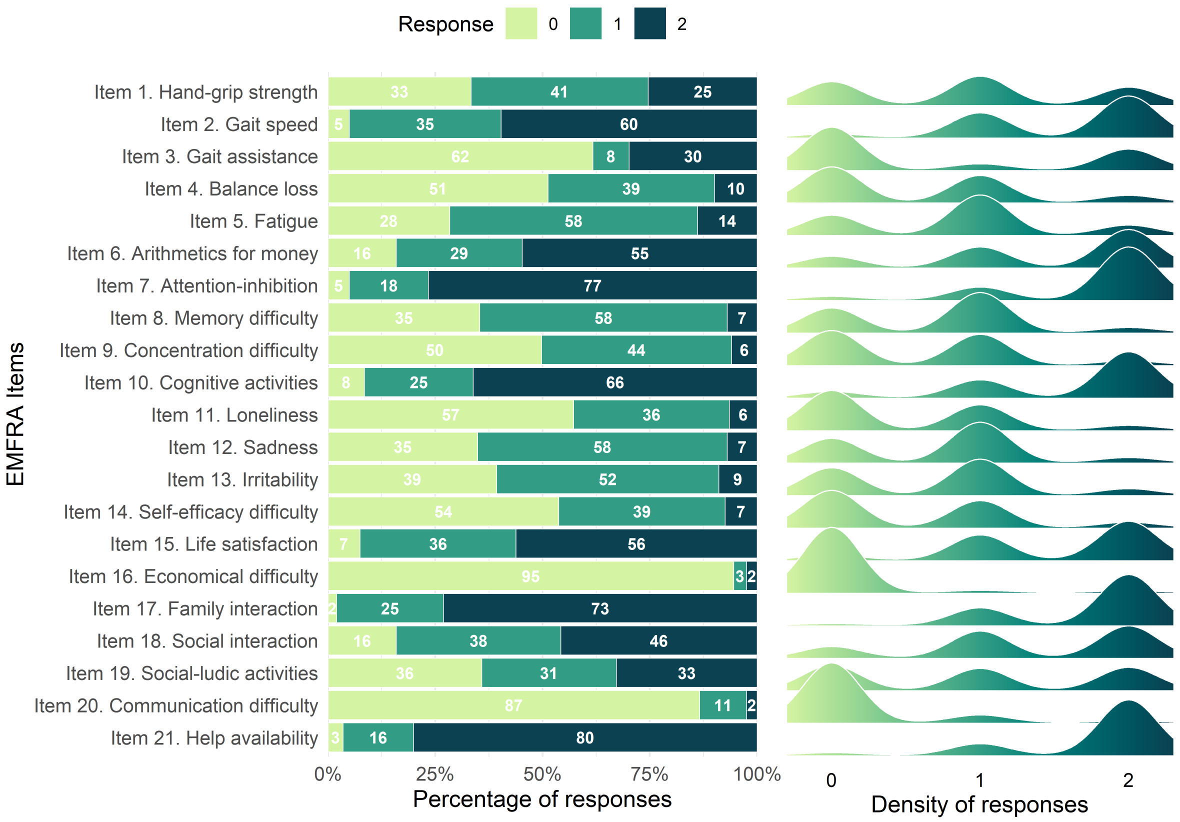

Figure 8: Distribution and density of EMFRA item responses

### 3 Item-level test–retest reliability

```
Dataset_retest <- data_emfra %>%
  filter(Assessed_retest == "Yes")
```

#### 3.1 Contingency tables

For each EMFRA item, contingency tables were computed to describe the distribution of test and retest responses. Rows correspond to test scores and columns correspond to retest scores. A supplementary file with all contingency tables was generated. Tables would be displayed as follows:

```
compute_contingency_emfra <- function(test_col, retest_col) {

  data_pair <- Dataset_retest[, c(test_col, retest_col)] %>%
    as.data.frame()

  data_pair[] <- lapply(data_pair, as.numeric)
```

```r
  data_pair <- data_pair[complete.cases(data_pair), ]

  table(
    Test   = data_pair[[1]],
    Retest = data_pair[[2]]
  )
}

contingency_tables_emfra <- map2(
  emfra_item_ids,
  emfra_item_ids_retest,
  compute_contingency_emfra
)

names(contingency_tables_emfra) <- emfra_item_labels

contingency_tables_emfra[["Item 1. Hand-grip strength"]]
```

```
    Retest
Test  0  1  2
   0 10  1  0
   1  3  7  0
   2  0  0  1
```

#### 3.2 Type of weightings

Radical weightings were applied to all items, except for Item 2, for which a custom weighting matrix was specified to reflect any additional discrepancy for disagreements involving the "0" category ("unable to walk") with any of the others ("0" and "1"), which would indicate the ability to walk.

```r
radical.weights(0:2)
```

```
          [,1]      [,2]      [,3]
[1,] 1.0000000 0.2928932 0.0000000
[2,] 0.2928932 1.0000000 0.2928932
[3,] 0.0000000 0.2928932 1.0000000
```

```r
custom_weight_matrix <- matrix(
  c(
    1, 0,         0,
    0, 1, 0.2928932,
    0, 0.2928932, 1
  ),
  nrow = 3,
  byrow = TRUE
)
```

```r
colnames(custom_weight_matrix) <- c("Category 0", "Category 1", "Category 2")
rownames(custom_weight_matrix) <- c("Category 0", "Category 1", "Category 2")

custom_weight_matrix
```

```
           Category 0 Category 1 Category 2
Category 0          1  0.0000000  0.0000000
Category 1          0  1.0000000  0.2928932
Category 2          0  0.2928932  1.0000000
```

##### 3.3 AC2 calculation and results

The AC2 coefficient was employed for test-retest reliability, reporting the following values: the AC2 coefficient, observed agreement (pa), expected agreement (pe), standard error, 95% CI, and p-value.

```r
compute_ac2_one_item <- function(test_col, retest_col, allowed_categories) {

  data_pair <- Dataset_retest[, c(test_col, retest_col)] %>%
    as.data.frame()

  data_pair[] <- lapply(data_pair, as.numeric)
  data_pair <- data_pair[complete.cases(data_pair), ]

  N_retest <- nrow(data_pair)

  if (N_retest < 2) {
    return(data.frame(
      item_id    = test_col,
      N          = N_retest,
      AC2        = NA,
      pa         = NA,
      pe         = NA,
      se         = NA,
      CI_low     = NA,
      CI_high    = NA,
      p_value    = NA,
      Weighting = "insufficient data",
      stringsAsFactors = FALSE
    ))
  }

  weight_matrix <- radical.weights(allowed_categories)
  weight_used   <- "radical"

  if (test_col == "EMFRA_item2_trait") {
```

```
    weight_matrix <- matrix(
      c(
        1, 0,         0,
        0, 1, 0.2928932,
        0, 0.2928932, 1
      ),
      nrow = 3, byrow = TRUE
    )
    colnames(weight_matrix) <- allowed_categories
    rownames(weight_matrix) <- allowed_categories
    weight_used <- "custom"
  }

  ac2_res <- gwet.ac1.raw(
    ratings      = data_pair,
    weights      = weight_matrix,
    categ.labels = allowed_categories
  )

  est <- ac2_res$est

  ci_raw   <- est$conf.int
  ci_clean <- gsub("[()]", "", ci_raw)

  CI_low  <- as.numeric(sub(",.*", "", ci_clean))
  CI_high <- as.numeric(sub(".*,", "", ci_clean))

  data.frame(
    item_id   = test_col,
    N         = N_retest,
    AC2       = est$coeff.val,
    pa        = est$pa,
    pe        = est$pe,
    se        = est$coeff.se,
    CI_low    = CI_low,
    CI_high   = CI_high,
    p_value   = est$p.value,
    Weighting = weight_used,
    stringsAsFactors = FALSE
  )
}
```

Producing the following results:

```
Reliability_AC2_EMFRA <- map2_df(
  emfra_item_ids,
  paste0(emfra_item_ids, "_retest"),
```

```
  ~ compute_ac2_one_item(.x, .y, allowed_categories = 0:2)
) %>%
  mutate(
    Item = factor(item_id, levels = emfra_item_ids, labels = emfra_item_labels),
    `95% CI` = paste0("(", round(CI_low, 3), ", ", round(CI_high, 3), ")"),
    AC2 = round(AC2, 3),
    pa  = round(pa, 3),
    pe  = round(pe, 3),
    se  = round(se, 3),
    p_value = format.pval(p_value, digits = 3, eps = .001)
  ) %>%
  select(
    Item,
    N,
    AC2,
    se,
    `95% CI`,
    p_value,
    pa,
    pe,
    Weighting
  )
```

Table 5: Test-retest Reliability (AC2)

| Item | N | AC2 | se | 95% CI | p_value | pa | pe | Weighting |
|---|---|---|---|---|---|---|---|---|
| Item 1. Hand-grip strength | 22 | 0.796 | 0.095 | (0.599, 0.993) | <0.001 | 0.871 | 0.371 | radical |
| Item 2. Gait speed | 22 | 0.918 | 0.059 | (0.796, 1) | <0.001 | 0.936 | 0.220 | custom |
| Item 3. Gait assistance | 22 | 0.875 | 0.089 | (0.689, 1) | <0.001 | 0.922 | 0.381 | radical |
| Item 4. Balance loss | 22 | 0.949 | 0.051 | (0.844, 1) | <0.001 | 0.968 | 0.366 | radical |
| Item 5. Fatigue | 22 | 0.741 | 0.109 | (0.513, 0.968) | <0.001 | 0.839 | 0.380 | radical |
| Item 6. Arithmetics for money | 22 | 0.699 | 0.130 | (0.429, 0.969) | <0.001 | 0.826 | 0.422 | radical |
| Item 7. Attention-inhibition | 22 | 0.704 | 0.128 | (0.437, 0.971) | <0.001 | 0.813 | 0.367 | radical |
| Item 8. Memory difficulty | 22 | 0.605 | 0.124 | (0.347, 0.862) | <0.001 | 0.743 | 0.350 | radical |
| Item 9. Concentration difficulty | 22 | 0.584 | 0.123 | (0.329, 0.84) | <0.001 | 0.743 | 0.381 | radical |
| Item 10. Cognitive activities | 22 | 0.951 | 0.049 | (0.85, 1) | <0.001 | 0.968 | 0.341 | radical |
| Item 11. Loneliness | 22 | 0.460 | 0.132 | (0.186, 0.735) | 0.001 | 0.679 | 0.404 | radical |
| Item 12. Sadness | 22 | 0.691 | 0.116 | (0.45, 0.932) | <0.001 | 0.807 | 0.376 | radical |
| Item 13. Irritability | 22 | 0.677 | 0.121 | (0.425, 0.929) | <0.001 | 0.794 | 0.361 | radical |
| Item 14. Self-efficacy difficulty | 22 | 0.719 | 0.113 | (0.485, 0.954) | <0.001 | 0.839 | 0.427 | radical |
| Item 15. Life satisfaction | 22 | 0.390 | 0.132 | (0.115, 0.665) | 0.004 | 0.646 | 0.420 | radical |
| Item 16. Economical difficulty | 22 | 0.891 | 0.067 | (0.751, 1) | <0.001 | 0.904 | 0.117 | radical |
| Item 17. Family interaction | 22 | 0.663 | 0.117 | (0.419, 0.907) | <0.001 | 0.775 | 0.333 | radical |
| Item 18. Social interaction | 22 | 0.484 | 0.149 | (0.174, 0.794) | 0.002 | 0.697 | 0.414 | radical |
| Item 19. Social-ludic activities | 22 | 0.784 | 0.107 | (0.561, 1) | <0.001 | 0.871 | 0.404 | radical |
| Item 20. Communication difficulty | 22 | 0.622 | 0.127 | (0.359, 0.886) | <0.001 | 0.730 | 0.284 | radical |
| Item 21. Help availability | 22 | 0.725 | 0.112 | (0.491, 0.959) | <0.001 | 0.794 | 0.251 | radical |

```r
ggplot(Reliability_AC2_EMFRA,
       aes(x = AC2, y = Item, color = AC2)) +

  geom_errorbarh(
    aes(xmin = as.numeric(sub(".*, ", "", gsub("[()]", "", `95% CI`))),
        xmax = as.numeric(sub(",.*", "", gsub("[()]", "", `95% CI`)))),
    height = 0.25,
    linewidth = 0.7
  ) +

  geom_point(size = 2) +

  scale_color_gradientn(
    colours = rev(paletteer_c("grDevices::ag_Sunset", 100)),
    limits = c(0, 1)
  ) +

  scale_x_continuous(
    limits = c(0, 1),
    breaks = seq(0, 1, 0.1)
  ) +

  scale_y_discrete(
    limits = rev(levels(Reliability_AC2_EMFRA$Item))
  ) +

  labs(
    x = "Test-retest Gwet's AC2 (Radical Items 1,3-21; Custom Item 2)",
    y = "EMFRA Items",
    color = "AC2"
  ) +
  theme_minimal()
```

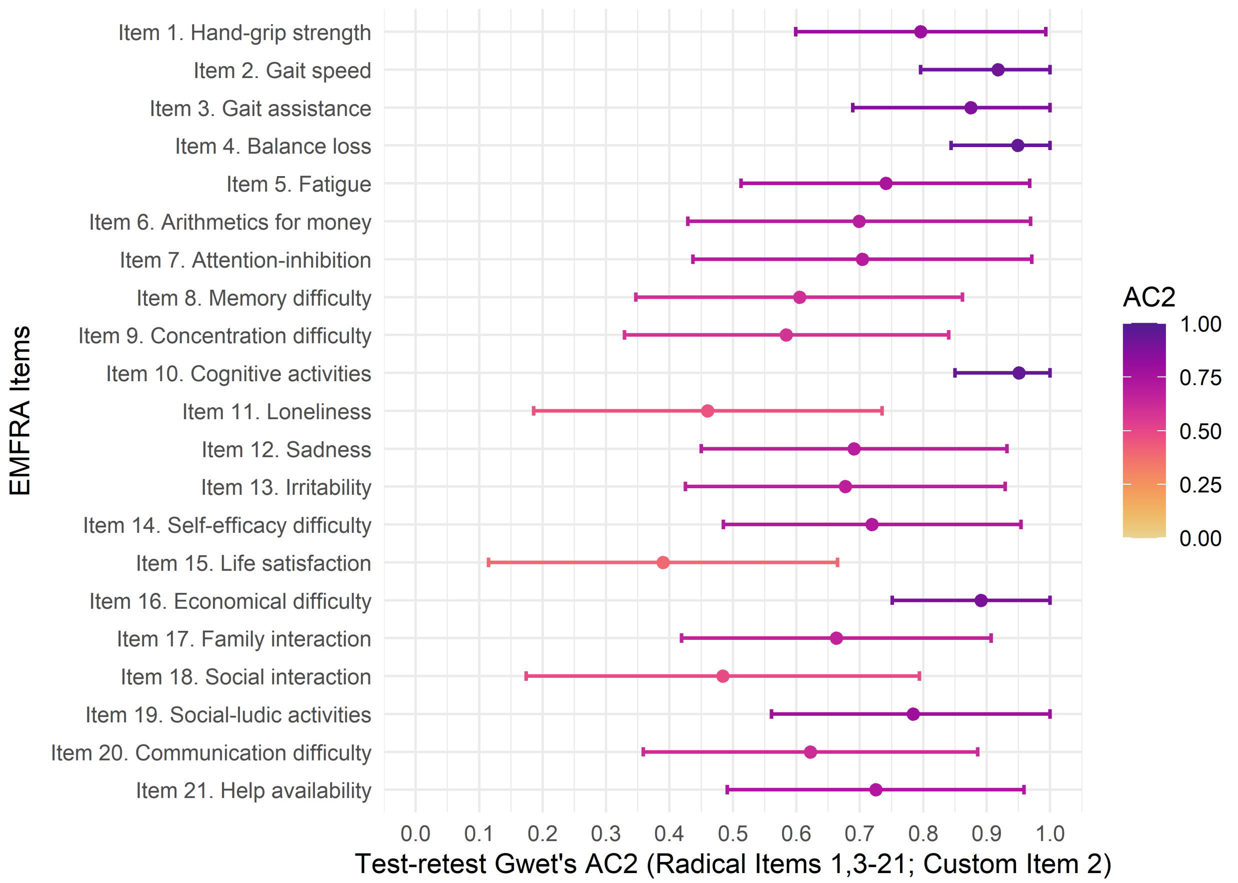

Figure 9: Test-Retest reliability of EMFRA items

### 4 Structural validity

#### 4.1 Response category combinations between EMFRA items

To evaluate the empirical support of the correlation structure and to detect potential sources of numerical instability, all pairwise combinations of EMFRA response categories were examined. For each pair of items, full 3×3 contingency tables were computed, explicitly retaining empty cells. The presence of structurally empty response combinations was inspected, as these patterns may indicate sparse contingency structures and potential violations of positive semi-definiteness (PSD) in subsequent correlation matrices.

```
emfra_items_data <- data_emfra %>%
  select(all_of(emfra_item_ids)) %>%
  mutate(across(everything(), as.numeric))

label_lookup <- tibble(
  item_id    = emfra_item_ids,
  item_label = emfra_item_labels
```

```
)

cell_report <- list()

for (i in 1:(length(emfra_item_ids) - 1)) {
  for (j in (i + 1):length(emfra_item_ids)) {

    `Item A` <- emfra_item_ids[i]
    `Item B` <- emfra_item_ids[j]

    tab <- table(
      factor(emfra_items_data[[`Item A`]], levels = c(0, 1, 2)),
      factor(emfra_items_data[[`Item B`]], levels = c(0, 1, 2))
    )

    cell_report[[length(cell_report) + 1]] <- as.data.frame(tab) %>%
      mutate(
        `Item A` = label_lookup$item_label[match(`Item A`, label_lookup$item_id)],
        `Item B` = label_lookup$item_label[match(`Item B`, label_lookup$item_id)]
      ) %>%
      rename(
        `Category A` = Var1,
        `Category B` = Var2,
        Count      = Freq
      )
  }
}

emfra_combination_cells <- bind_rows(cell_report)

emfra_combination_cells <- emfra_combination_cells %>%
  mutate(
    `Item A` = factor(`Item A`, levels = emfra_item_labels),
    `Item B` = factor(`Item B`, levels = emfra_item_labels)
  ) %>%
  arrange(
    `Item A`,
    `Category A`,
    `Item B`,
    `Category B`
  ) %>%
  select(
    `Item A`,
    `Category A`,
    `Item B`,
    `Category B`,
    Count
  )
```

Table 6: EMFRA response combinations

| Item A | Category A | Item B | Category B | Count |
|---|---|---|---|---|
| Item 1. Hand-grip strength | 0 | Item 2. Gait speed | 0 | 9 |
| Item 1. Hand-grip strength | 0 | Item 2. Gait speed | 1 | 38 |
| Item 1. Hand-grip strength | 0 | Item 2. Gait speed | 2 | 20 |
| Item 1. Hand-grip strength | 0 | Item 3. Gait assistance | 0 | 21 |
| Item 1. Hand-grip strength | 0 | Item 3. Gait assistance | 1 | 9 |
| Item 1. Hand-grip strength | 0 | Item 3. Gait assistance | 2 | 37 |
| Item 1. Hand-grip strength | 0 | Item 4. Balance loss | 0 | 20 |
| Item 1. Hand-grip strength | 0 | Item 4. Balance loss | 1 | 34 |
| Item 1. Hand-grip strength | 0 | Item 4. Balance loss | 2 | 13 |
| Item 1. Hand-grip strength | 0 | Item 5. Fatigue | 0 | 13 |

```
Total combinations between items and response categories: 1890
```

Certain combinations were not present in the dataset:

```r
emfra_combination_empty_cells <- emfra_combination_cells %>%
  filter(Count == 0)
```

Table 7: EMFRA missing combinations

| Item A | Category A | Item B | Category B | Count |
|---|---|---|---|---|
| Item 1. Hand-grip strength | 0 | Item 20. Communication difficulty | 2 | 0 |
| Item 1. Hand-grip strength | 1 | Item 17. Family interaction | 0 | 0 |
| Item 1. Hand-grip strength | 2 | Item 2. Gait speed | 0 | 0 |
| Item 1. Hand-grip strength | 2 | Item 20. Communication difficulty | 2 | 0 |
| Item 2. Gait speed | 0 | Item 3. Gait assistance | 0 | 0 |
| Item 2. Gait speed | 0 | Item 3. Gait assistance | 1 | 0 |
| Item 2. Gait speed | 0 | Item 4. Balance loss | 0 | 0 |
| Item 2. Gait speed | 0 | Item 9. Concentration difficulty | 2 | 0 |
| Item 2. Gait speed | 0 | Item 11. Loneliness | 2 | 0 |
| Item 2. Gait speed | 0 | Item 14. Self-efficacy difficulty | 2 | 0 |

```
Total empty combinations: 81
```

#### 4.2 Polychoric correlation matrix

##### 4.2.1 Matrix diagnostics and selection functions

```r
check_correlation_matrix <- function(R) {
  eig <- eigen(R, symmetric = TRUE, only.values = TRUE)$values
  neg <- eig[eig < 0]

  tibble::tibble(
    Symmetric = is.symmetric.matrix(R),
    PSD = matrixcalc::is.positive.semi.definite(R, tol = 0),
    `Total eigenvalues (n)` = length(eig),
    `Negative eigenvalues (n)` = length(neg),
    `Minimum eigenvalue` = min(eig),
    `Negative eigenvalues` = if (length(neg) > 0) {
      paste(round(neg, 8), collapse = ", ")
    } else {
      NA_character_
    }
  )
}

compute_polychoric_psych <- function(data, correct = 0) {
  psych::polychoric(
    x = data,
    correct = correct,
    smooth = FALSE,
    global = FALSE,
    weight = NULL,
    na.rm = TRUE,
    delete = TRUE,
    max.cat = 8
  )$rho
}

compute_polychoric_eganet <- function(data, correction = c("zero", "point_five"))
 ↪  {
  correction <- match.arg(correction)

  empty.value <- if (correction == "zero") "none" else "point_five"

  EGAnet::polychoric.matrix(
    data = data,
    na.data = "pairwise",
    empty.method = "zero",
```

```r
    empty.value = empty.value
  )
}

select_polychoric_matrices <- function(data) {

  R_psych_0  <- compute_polychoric_psych(data, correct = 0)
  R_eganet_0 <- compute_polychoric_eganet(data, correction = "zero")

  diag_psych_0  <- check_correlation_matrix(R_psych_0)
  diag_eganet_0 <- check_correlation_matrix(R_eganet_0)

  ok_psych_0  <- isTRUE(diag_psych_0$Symmetric)  && isTRUE(diag_psych_0$PSD)
  ok_eganet_0 <- isTRUE(diag_eganet_0$Symmetric) && isTRUE(diag_eganet_0$PSD)

  if (ok_psych_0 && ok_eganet_0) {
    return(list(
      correction_used = 0,
      psych = list(
        Rpoly = R_psych_0,
        diagnostics = diag_psych_0,
        method = list(
          package = "psych",
          correct = 0,
          smooth = FALSE,
          global = FALSE,
          na.rm = TRUE,
          delete = TRUE,
          max.cat = 8
        )
      ),
      eganet = list(
        Rpoly = R_eganet_0,
        diagnostics = diag_eganet_0,
        method = list(
          package = "EGAnet",
          na.data = "pairwise",
          empty.method = "zero",
          empty.value = "none"
        )
      ),
      diagnostics_initial = list(
        psych_0 = diag_psych_0,
        eganet_0 = diag_eganet_0
      ),
      diagnostics_final = list(
        psych = diag_psych_0,
```

```
      eganet = diag_eganet_0
    )
  ))
}

R_psych_05  <- compute_polychoric_psych(data, correct = 0.5)
R_eganet_05 <- compute_polychoric_eganet(data, correction = "point_five")

diag_psych_05  <- check_correlation_matrix(R_psych_05)
diag_eganet_05 <- check_correlation_matrix(R_eganet_05)

ok_psych_05  <- isTRUE(diag_psych_05$Symmetric)  && isTRUE(diag_psych_05$PSD)
ok_eganet_05 <- isTRUE(diag_eganet_05$Symmetric) && isTRUE(diag_eganet_05$PSD)

if (ok_psych_05 && ok_eganet_05) {
  return(list(
    correction_used = 0.5,
    psych = list(
      Rpoly = R_psych_05,
      diagnostics = diag_psych_05,
      method = list(
        package = "psych",
        correct = 0.5,
        smooth = FALSE,
        global = FALSE,
        na.rm = TRUE,
        delete = TRUE,
        max.cat = 8
      )
    ),
    eganet = list(
      Rpoly = R_eganet_05,
      diagnostics = diag_eganet_05,
      method = list(
        package = "EGAnet",
        na.data = "pairwise",
        empty.method = "zero",
        empty.value = "point_five"
      )
    ),
    diagnostics_initial = list(
      psych_0 = diag_psych_0,
      eganet_0 = diag_eganet_0,
      psych_05 = diag_psych_05,
      eganet_05 = diag_eganet_05
    ),
    diagnostics_final = list(
```

```r
        psych = diag_psych_05,
        eganet = diag_eganet_05
      )
    ))
  }

  stop(
    paste0(
      "No valid polychoric matrix scenario was obtained. ",
      "At least one of the psych or EGAnet matrices failed symmetry and/or PSD ",
      "with correction = 0, and at least one of them also failed with correction =
       ↪  0.5."
    )
  )
}

build_matrix_diagnostics_table <- function(selected_obj, model_name) {

  diagnostics_tbl <- dplyr::bind_rows(
    selected_obj$diagnostics_initial$psych_0 %>%
      dplyr::mutate(
        Model = model_name,
        Package = "psych",
        Correction = "0",
        `Selected for dim. analysis` = ifelse(
          selected_obj$correction_used == 0,
          "Yes",
          "No"
        )
      ),

    selected_obj$diagnostics_initial$eganet_0 %>%
      dplyr::mutate(
        Model = model_name,
        Package = "EGAnet",
        Correction = "0",
        `Selected for dim. analysis` = ifelse(
          selected_obj$correction_used == 0,
          "Yes",
          "No"
        )
      )
  )

  if (!is.null(selected_obj$diagnostics_initial$psych_05) &&
      !is.null(selected_obj$diagnostics_initial$eganet_05)) {
```

```r
    diagnostics_tbl <- dplyr::bind_rows(
      diagnostics_tbl,

      selected_obj$diagnostics_initial$psych_05 %>%
        dplyr::mutate(
          Model = model_name,
          Package = "psych",
          Correction = "0.5",
          `Selected for dim. analysis` = ifelse(
            selected_obj$correction_used == 0.5,
            "Yes",
            "No"
          )
        ),

      selected_obj$diagnostics_initial$eganet_05 %>%
        dplyr::mutate(
          Model = model_name,
          Package = "EGAnet",
          Correction = "0.5",
          `Selected for dim. analysis` = ifelse(
            selected_obj$correction_used == 0.5,
            "Yes",
            "No"
          )
        )
    )
  }

  diagnostics_tbl %>%
    dplyr::relocate(
      Model,
      Package,
      Correction,
      `Selected for dim. analysis`
    )
}
```

##### 4.2.2 Matrix diagnostics

Symmetry and PSD (zero or positive eigenvalues) features of the matrix should be present to perform the structural analyses.

The polychoric matrix was calculated using the package *EGAnet* with the function polychoric.matrix. The adequacy of the resulting correlation matrices was evaluated using eigenvalue inspection and formal PSD checks implemented in the *matrixcalc* package.

The raw polychoric matrix is generated (poly_raw), and checked for symmetry and PSD.

```r
selected_poly <- select_polychoric_matrices(emfra_items_data)

Rpoly <- selected_poly$eganet$Rpoly

emfra_poly_diagnostics_tbl <- build_matrix_diagnostics_table(
  selected_obj = selected_poly,
  model_name   = "EMFRA"
)
```

Table 8: Diagnosis and correction features of Polychoric Matrix in EMFRA (21 items)

| Model | Package | Correction | Selected for dim. analysis | Symmetric | PSD | Total eigenvalues (n) | Negative eigenvalues (n) | Minimum eigenvalue | Negative eigenvalues |
|---|---|---|---|---|---|---|---|---|---|
| EMFRA | psych | 0 | No | TRUE | FALSE | 21 | 1 | -0.035 | -0.03508537 |
| EMFRA | EGAnet | 0 | No | TRUE | FALSE | 21 | 1 | -0.014 | -0.0139064 |
| EMFRA | psych | 0.5 | Yes | TRUE | TRUE | 21 | 0 | 0.018 | NA |
| EMFRA | EGAnet | 0.5 | Yes | TRUE | TRUE | 21 | 0 | 0.025 | NA |

```r
emfra_corr_df <- Rpoly %>%
  as.data.frame() %>%
  rownames_to_column("Item A") %>%
  pivot_longer(
    cols      = -`Item A`,
    names_to  = "Item B",
    values_to = "Correlation"
  ) %>%
  mutate(
    `Item A` = factor(`Item A`, levels = rownames(Rpoly)),
    `Item B` = factor(`Item B`, levels = colnames(Rpoly))
  ) %>%
  filter(as.integer(`Item A`) >= as.integer(`Item B`)) %>%
  mutate(
    Item_A_label = factor(`Item A`, levels = emfra_item_ids, labels =
     ↪  emfra_item_labels),
    Item_B_label = factor(`Item B`, levels = emfra_item_ids, labels =
     ↪  emfra_item_labels)
  )
```

Graphical representation of Rpoly applying gradient colors to all values.

```r
ggplot(emfra_corr_df,
       aes(x = Item_B_label, y = Item_A_label, fill = Correlation)) +
  geom_tile(color = "white", linewidth = 0.3) +
  geom_text(
    aes(label = sprintf("%.2f", Correlation)),
    size  = 3,
```

```
    color = "black"
  ) +
  scale_fill_gradientn(
    colours = c(
      paletteer_c("ggthemes::Orange-Blue-White Diverging", 1000)[1000],
      paletteer_c("ggthemes::Orange-Blue-White Diverging", 1000)[500],
      paletteer_c("ggthemes::Orange-Blue-White Diverging", 1000)[1]
    ),
    values = scales::rescale(c(-1, 0, 1)),
    limits = c(-1, 1),
    oob    = scales::squish,
    name   = expression(rho ~ polychoric)
  ) +
  scale_x_discrete(position = "top") +
  scale_y_discrete(limits = rev(levels(emfra_corr_df$Item_A_label))) +
  theme_minimal(base_size = 12) +
  theme(
    panel.grid.major.x = element_line(color = "grey80"),
    panel.grid.major.y = element_blank(),
    panel.grid.minor   = element_blank(),
    axis.title         = element_blank(),
    axis.text.x        = element_text(size = 10, angle = 45, vjust = 0, hjust =
     ↪  0),
    axis.text.y        = element_text(size = 10),
    legend.position    = "right"
  )
```

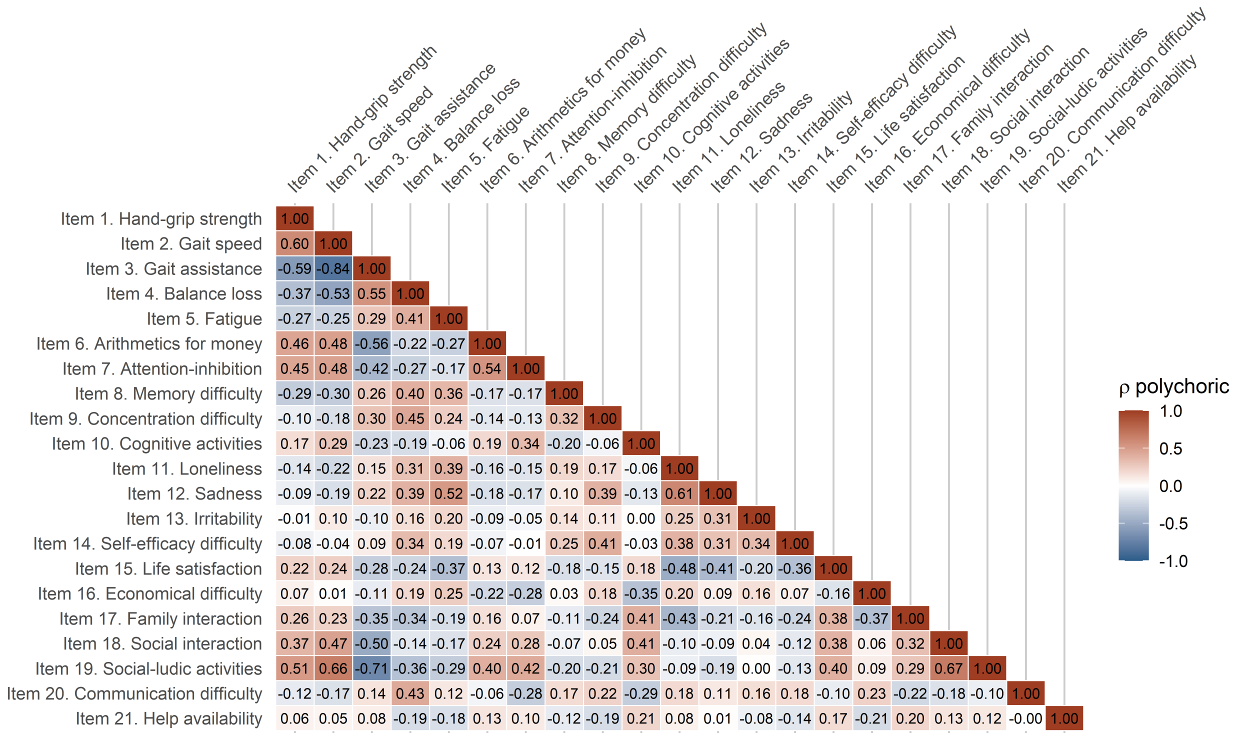

Figure 10: Polychoric correlation matrix of EMFRA items

In addition, Rpoly is presented with poor correlation values (-0.29 to 0.29) filled in white.

```
threshold_0_29 <- 0.29

emfra_corr_df <- emfra_corr_df %>%
  mutate(
    Correlation_fill = ifelse(abs(Correlation) < threshold_0_29, NA, Correlation)
  )

p_main_corr <- ggplot(
  emfra_corr_df,
  aes(x = Item_B_label, y = Item_A_label, fill = Correlation_fill)
) +
  geom_tile(color = "white", linewidth = 0.3) +
  geom_tile(
    data      = subset(emfra_corr_df, abs(Correlation) >= threshold_0_29),
    fill      = NA,
    color     = "grey40",
    linewidth = 0.1
  ) +
  geom_text(
    aes(label = sprintf("%.2f", Correlation)),
    color = "black",
    size  = 3
```

```r
  ) +
  scale_fill_gradientn(
    colours = c(
      paletteer_c("ggthemes::Orange-Blue-White Diverging", 1000)[1000],
      "white",
      paletteer_c("ggthemes::Orange-Blue-White Diverging", 1000)[1]
    ),
    values   = scales::rescale(c(-1, 0, 1)),
    limits   = c(-1, 1),
    na.value = "white",
    guide    = "none"
  ) +
  scale_x_discrete(position = "top") +
  scale_y_discrete(limits = rev(levels(emfra_corr_df$Item_A_label))) +
  theme_minimal(base_size = 12) +
  theme(
    panel.grid.major.x = element_line(color = "grey80"),
    panel.grid.major.y = element_blank(),
    panel.grid.minor   = element_blank(),
    axis.title         = element_blank(),
    axis.text.x        = element_text(size = 10, angle = 45, vjust = 0, hjust =
    ↪  0),
    axis.text.y        = element_text(size = 10),
    legend.position    = "none"
  )

legend_df_corr <- data.frame(
  x     = 1,
  y     = seq(-1, 1, length.out = 1000),
  value = seq(-1, 1, length.out = 1000)
)

p_legend_corr <- ggplot(legend_df_corr, aes(x, y, fill = value)) +
  geom_tile() +
  scale_fill_gradientn(
    colours = c(
      paletteer_c("ggthemes::Orange-Blue-White Diverging", 1000)[1000],
      paletteer_c("ggthemes::Orange-Blue-White Diverging", 1000)[500],
      "white",
      "white",
      paletteer_c("ggthemes::Orange-Blue-White Diverging", 1000)[500],
      paletteer_c("ggthemes::Orange-Blue-White Diverging", 1000)[1]
    ),
    values = scales::rescale(c(-1, -0.30, -0.29, 0.29, 0.30, 1)),
    limits = c(-1, 1),
    name   = expression(rho ~ polychoric)
  ) +
  theme_void() +
```

```r
  theme(legend.position = "right")

legend_grob_corr <- cowplot::get_legend(p_legend_corr)

grid.arrange(
  p_main_corr,
  legend_grob_corr,
  ncol    = 2,
  widths = c(3.5, 0.5)
)
```

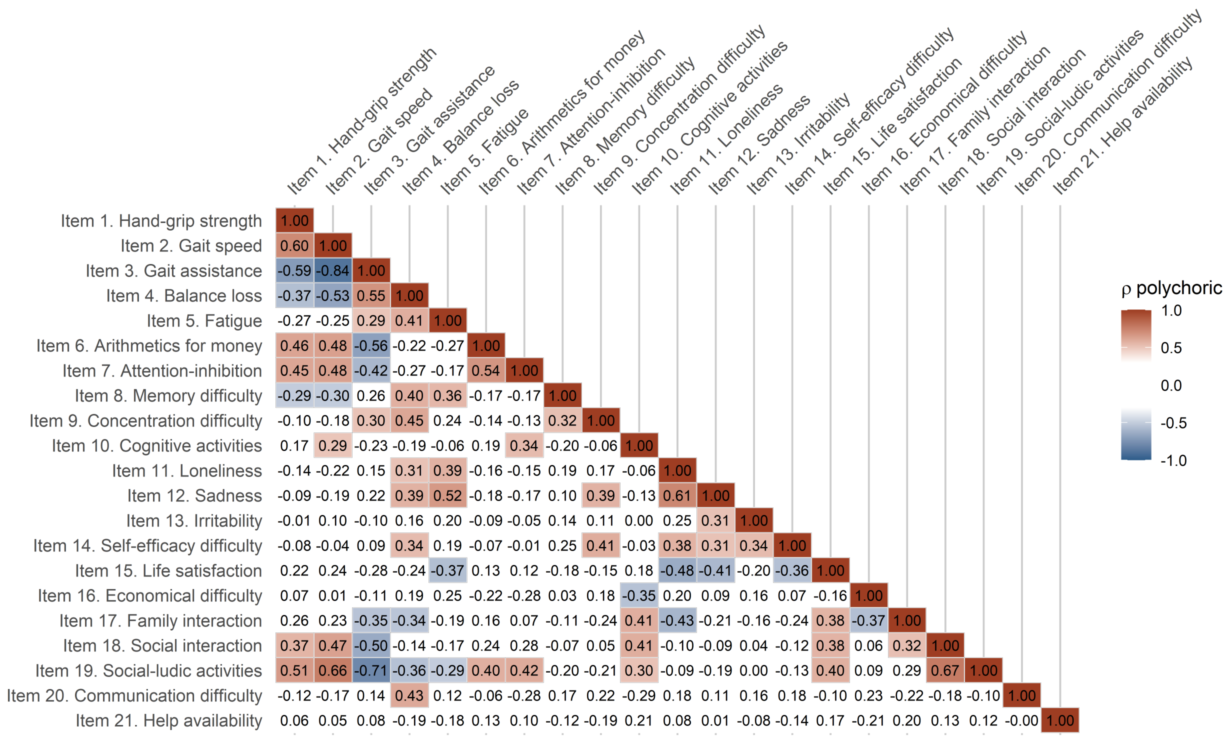

Figure 11: Polychoric correlation matrix of EMFRA items - Filtered

#### 4.3 Kaiser Meyer Olking (KMO) Measure of Sampling Adequacy (MSA)

##### 4.3.1 Anti-image correlation matrix

```r
emfra_KMO        <- KMO(Rpoly)
emfra_anti_image <- emfra_KMO$Image

emfra_anti_df <- emfra_anti_image %>%
  as.data.frame() %>%
  rownames_to_column("Item A") %>%
```

```r
  pivot_longer(
    cols      = -`Item A`,
    names_to  = "Item B",
    values_to = "Partial_correlation"
  ) %>%
  mutate(
    `Item A` = factor(`Item A`, levels = rownames(emfra_anti_image)),
    `Item B` = factor(`Item B`, levels = colnames(emfra_anti_image))
  ) %>%
  filter(as.integer(`Item A`) >= as.integer(`Item B`)) %>%
  mutate(
    Item_A_label = factor(`Item A`, levels = emfra_item_ids, labels =
     ↪  emfra_item_labels),
    Item_B_label = factor(`Item B`, levels = emfra_item_ids, labels =
     ↪  emfra_item_labels)
  )

ggplot(emfra_anti_df,
       aes(x = Item_B_label, y = Item_A_label, fill = Partial_correlation)) +
  geom_tile(color = "white") +
  geom_text(
    aes(label = sprintf("%.2f", Partial_correlation)),
    size  = 3,
    color = "black"
  ) +
  scale_fill_gradientn(
    colours = c(
      paletteer_c("ggthemes::Orange-Blue-White Diverging", 1000)[1000],
      paletteer_c("ggthemes::Orange-Blue-White Diverging", 1000)[500],
      paletteer_c("ggthemes::Orange-Blue-White Diverging", 1000)[1]
    ),
    values = scales::rescale(c(-1, 0, 1)),
    limits = c(-1, 1),
    oob    = scales::squish,
    name   = "Partial correlation\n(anti-image)"
  ) +
  scale_x_discrete(position = "top") +
  scale_y_discrete(limits = rev(levels(emfra_anti_df$Item_A_label))) +
  theme_minimal(base_size = 12) +
  theme(
    panel.grid.major.x = element_line(color = "grey80"),
    panel.grid.major.y = element_blank(),
    panel.grid.minor   = element_blank(),
    axis.text.x        = element_text(angle = 45, vjust = 0, hjust = 0),
    axis.title         = element_blank()
  )
```

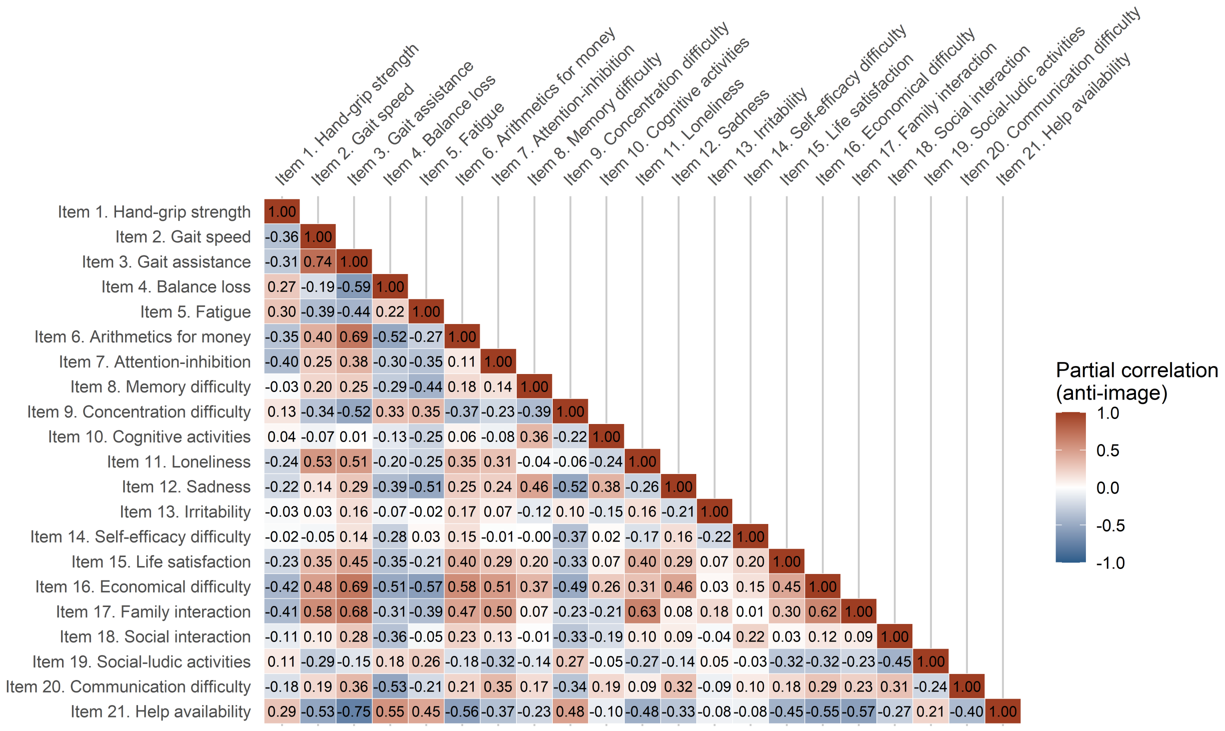

Figure 12: Polychoric Anti-image matrix of EMFRA items

##### 4.3.2 KMO MSA analysis

```
emfra_KMO_plot <- data.frame(
  item = names(emfra_KMO$MSAi),
  MSA  = as.numeric(emfra_KMO$MSAi)
) %>%
  mutate(
    item_label = factor(item, levels = emfra_item_ids, labels = emfra_item_labels)
  ) %>%
  bind_rows(
    data.frame(
      item       = "Overall_KMO",
      MSA        = emfra_KMO$MSA,
      item_label = "Overall KMO"
    )
  ) %>%
  mutate(
    item_label = factor(item_label, levels = c(emfra_item_labels, "Overall KMO")),
    is_overall = item == "Overall_KMO"
  )

ggplot(emfra_KMO_plot, aes(x = MSA, y = item_label)) +
```

```r
geom_point(
  data  = subset(emfra_KMO_plot, !is_overall),
  aes(color = MSA),
  size  = 3
) +
geom_point(
  data  = subset(emfra_KMO_plot, is_overall),
  color = "black",
  shape = 15,
  size  = 3
) +
scale_color_gradientn(
  colours = rev(paletteer_c("grDevices::Emrld", 100)),
  limits  = c(0, 1)
) +
scale_x_continuous(limits = c(0, 1), breaks = seq(0, 1, 0.1)) +
scale_y_discrete(limits = rev(levels(emfra_KMO_plot$item_label))) +
labs(
  x     = "Kaiser-Meyer-Olkin (MSA)",
  y     = "EMFRA Items",
  color = "MSA value"
) +
theme_minimal() +
theme(
  axis.title      = element_text(size = 12),
  axis.text       = element_text(size = 10),
  legend.position = "right"
)
```

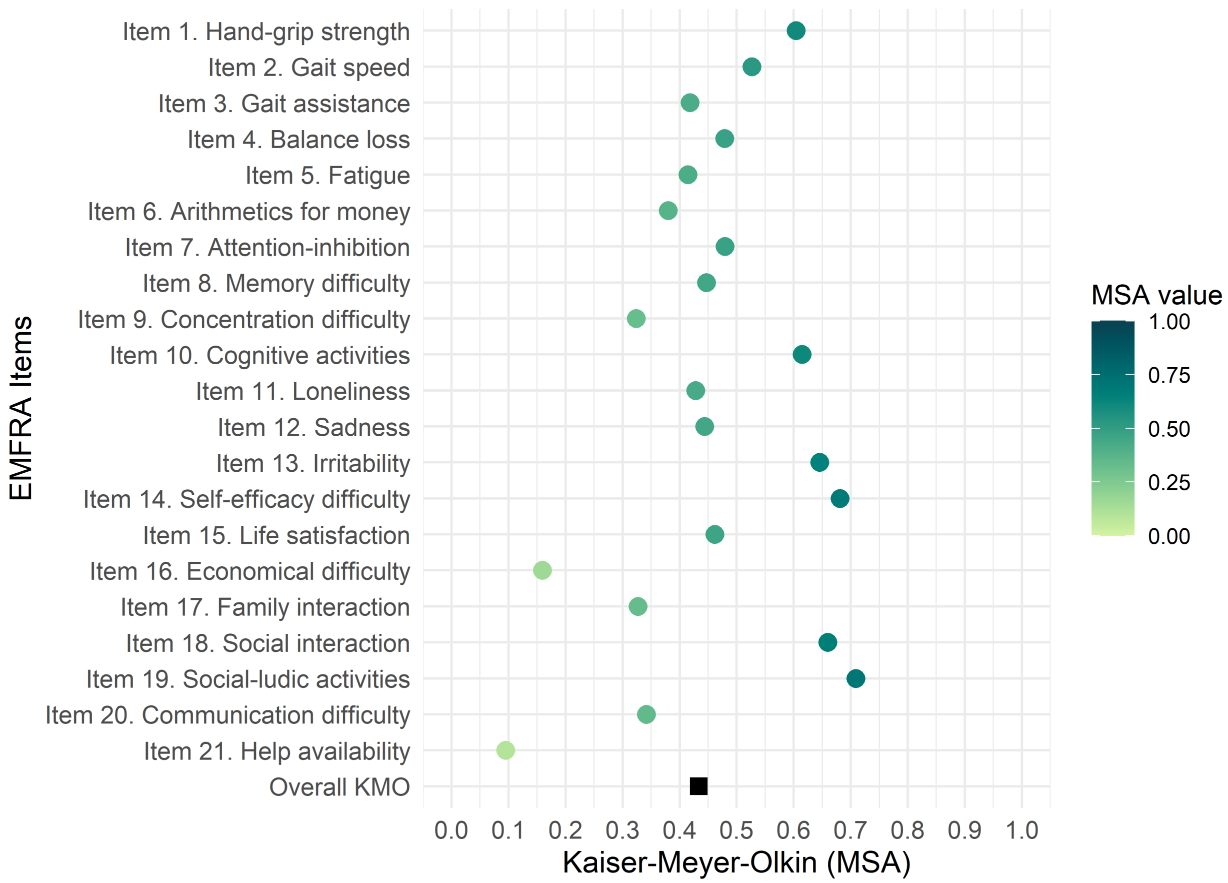

Figure 13: Kaiser-Meyer-Olkin and Measure of Sampling Adequacy of EMFRA items

#### 4.4 Exploratory detection of latent factors with EGA

#### 4.5 Functions for Dimensionality analysis

##### 4.5.1 Bootstrap PA and FA dimensionality functions

```
.compute_polychoric_safe <- function(data,
                                     correct_override = NULL,
                                     psd_tol          = -0.10) {

  .try_poly <- function(correct) {
    tryCatch(
      suppressWarnings(suppressMessages(
        psych::polychoric(
          x       = data,
          correct = correct,
          smooth  = FALSE,
          global  = FALSE,
```

```
          na.rm    = TRUE,
          delete   = TRUE,
          max.cat = 8
        )$rho
      )),
      error = function(e) NULL
    )
  }

  .has_bad_values <- function(R) {
    is.null(R) || anyNA(R) || any(!is.finite(R))
  }

  if (!is.null(correct_override)) {
    R <- .try_poly(correct_override)
  } else {
    R <- .try_poly(0)
    if (.has_bad_values(R)) R <- .try_poly(0.5)
  }

  if (.has_bad_values(R)) return(NULL)

  R <- (R + t(R)) / 2
  diag(R) <- 1

  eig_min <- min(eigen(R, symmetric = TRUE, only.values = TRUE)$values)

  if (eig_min < psd_tol) {
    R_near <- tryCatch(
      as.matrix(Matrix::nearPD(
        R,
        corr            = TRUE,
        ensureSymmetry = TRUE,
        keepDiag        = TRUE
      )$mat),
      error = function(e) NULL
    )
    if (is.null(R_near) || anyNA(R_near)) return(NULL)
    R <- R_near
  }

  R
}

run_bootstrap_eigen <- function(data,
                                n_boot            = 1000,
                                probs             = c(0.025, 0.975),
```

```r
                              correct_override = NULL,
                              psd_tol          = -0.10,
                              fm               = "minres",
                              nfactors_fa      = 1,
                              report_every     = 50,
                              parallel         = FALSE,
                              ncores           = max(1L, parallel::detectCores()
                               ↪  - 1L),
                              seed             = 42,
                              verbose          = TRUE) {

  if (!is.data.frame(data) && !is.matrix(data))
    stop("`data` must be a data.frame or a matrix.")
  if (n_boot < 10)
    stop("`n_boot` must be >= 10.")
  if (length(probs) != 2 || any(probs < 0) || any(probs > 1) || probs[1] >=
   ↪  probs[2])
    stop("`probs` must be a vector of two increasing probabilities in [0, 1].")
  if (!is.null(correct_override) && !correct_override %in% c(0, 0.5))
    stop("`correct_override` must be NULL, 0, or 0.5.")

  n_vars       <- ncol(data)
  use_parallel <- parallel && .Platform$OS.type != "windows"

  counter_env        <- new.env(parent = emptyenv())
  counter_env$i      <- 0L
  counter_env$active <- verbose && !use_parallel

  .boot_stat <- function(d, idx) {

    if (counter_env$active) {
      counter_env$i <- counter_env$i + 1L
      if (counter_env$i %% report_every == 0L ||
          counter_env$i == 1L ||
          counter_env$i == n_boot) {
        message(sprintf("  Iteration %d / %d  (%.0f%%)",
                        counter_env$i, n_boot,
                        100 * counter_env$i / n_boot))
      }
    }

    d_boot <- d[idx, , drop = FALSE]

    R <- .compute_polychoric_safe(
      data             = d_boot,
      correct_override = correct_override,
      psd_tol          = psd_tol
    )
```

```r
  if (is.null(R)) return(rep(NA_real_, n_vars))

  eig_vals <- tryCatch(
    suppressWarnings(suppressMessages(
      psych::fa(
        r         = R,
        nfactors = nfactors_fa,
        n.obs     = nrow(d_boot),
        fm        = fm,
        rotate    = "none",
        warnings = FALSE
      )$values
    )),
    error = function(e) rep(NA_real_, n_vars)
  )

  length(eig_vals) <- n_vars
  eig_vals
}

set.seed(seed)

if (verbose) {
  corr_label <- if (is.null(correct_override)) {
    "automatic (0 -> 0.5 if fails)"
  } else {
    as.character(correct_override)
  }
  message(sprintf(
    paste0(
      "Bootstrap Parallel Analysis\n",
      "  Iterations    : %d\n",
      "  Items (vars)  : %d\n",
      "  Correction    : %s\n",
      "  PSD threshold : %.2f\n",
      "  Eigenvalues   : fa()$values (fm = %s, nfactors = %d)\n",
      "  CI            : P%.4g \u2013 P%.4g\n",
      "  Mode          : %s\n",
      "Starting...\n"
    ),
    n_boot, n_vars, corr_label, psd_tol, fm, nfactors_fa,
    probs[1] * 100, probs[2] * 100,
    if (use_parallel) paste0("parallel (", ncores, " cores)") else "sequential"
  ))
  if (use_parallel)
    message("  (progress reporting disabled in parallel mode)")
}
```

```r
boot_result <- boot::boot(
  data      = data,
  statistic = .boot_stat,
  R         = n_boot,
  sim       = "ordinary",
  parallel  = if (use_parallel) "multicore" else "no",
  ncpus     = if (use_parallel) ncores else 1L
)

boot_matrix <- boot_result$t

failed_rows <- which(apply(boot_matrix, 1, anyNA))
n_failed    <- length(failed_rows)

if (n_failed > 0) {
  fail_pct <- 100 * n_failed / n_boot
  if (fail_pct > 10) {
    warning(sprintf(
      paste0(
        "%d/%d iterations (%.1f%%) discarded due to convergence failure.\n",
        "  If the rate is high, try:\n",
        "  - correct_override = 0.5\n",
        "  - a more negative psd_tol (current: %.2f), e.g. -0.15\n",
        "  - check for items with very few categories or low variance."
      ),
      n_failed, n_boot, fail_pct, psd_tol
    ))
  } else if (verbose) {
    message(sprintf(
      "  %d iterations discarded (%.1f%%) \u2014 acceptable rate.",
      n_failed, fail_pct
    ))
  }
  boot_matrix <- boot_matrix[-failed_rows, , drop = FALSE]
}

n_ok <- nrow(boot_matrix)

if (n_ok < 50)
  stop(sprintf(
    paste0(
      "Only %d/%d valid iterations. Bootstrap is not reliable.\n",
      "Try: correct_override = 0.5, psd_tol = -0.15, or both."
    ),
    n_ok, n_boot
  ))
```

```r
  boot_summary <- data.frame(
    Factor = seq_len(n_vars),
    Mean   = apply(boot_matrix, 2, mean,                          na.rm = TRUE),
    SD     = apply(boot_matrix, 2, sd,                            na.rm = TRUE),
    P_low  = apply(boot_matrix, 2, quantile, probs = probs[1], na.rm = TRUE),
    P_high = apply(boot_matrix, 2, quantile, probs = probs[2], na.rm = TRUE)
  )

  if (verbose)
    message(sprintf(
      "\nBootstrap completed: %d/%d valid iterations (%.1f%%).",
      n_ok, n_boot, 100 * n_ok / n_boot
    ))

  structure(
    list(
      boot_matrix      = boot_matrix,
      summary          = boot_summary,
      n_boot           = n_boot,
      n_ok             = n_ok,
      n_failed         = n_failed,
      probs            = probs,
      correct_override = correct_override,
      psd_tol          = psd_tol,
      fm               = fm,
      nfactors_fa      = nfactors_fa
    ),
    class = "bootstrap_eigen"
  )
}

print.bootstrap_eigen <- function(x, digits = 3, ...) {
  corr_label <- if (is.null(x$correct_override)) {
    "automatic (0 -> 0.5 if fails)"
  } else {
    as.character(x$correct_override)
  }
  cat("Bootstrap Parallel Analysis \u2014 Polychoric matrix eigenvalues\n")
  cat(sprintf("  Iterations requested     : %d\n",       x$n_boot))
  cat(sprintf("  Valid iterations         : %d (%.1f%%)\n",
              x$n_ok, 100 * x$n_ok / x$n_boot))
  cat(sprintf("  Discarded iterations     : %d\n",       x$n_failed))
  cat(sprintf("  Continuity correction    : %s\n",       corr_label))
  cat(sprintf("  PSD threshold (psd_tol)  : %.2f\n",     x$psd_tol))
  cat(sprintf("  Eigenvalue method        : fa()$values (fm = %s, nfactors =
  ↪  %d)\n",
              x$fm, x$nfactors_fa))
```

```r
  cat(sprintf("  CI used                  : P%.4g \u2013 P%.4g\n",
              x$probs[1] * 100, x$probs[2] * 100))
  cat("\nEigenvalue summary:\n")
  print(round(x$summary, digits))
  invisible(x)
}

run_parallel_bootstrap <- function(selected_obj,
                                   data,
                                   model_name    = "",
                                   n_iter_null   = 1000,
                                   n_boot        = 1000,
                                   probs         = c(0.05, 0.95),
                                   quant         = 0.95,
                                   fm            = "minres",
                                   nfactors_fa   = 1,
                                   psd_tol       = -0.10,
                                   report_every  = 50,
                                   parallel_boot = FALSE,
                                   ncores        = max(1L,
                                   ↪  parallel::detectCores() - 1L),
                                   seed          = 42,
                                   verbose       = TRUE) {

  if (!is.list(selected_obj) || is.null(selected_obj$correction_used))
    stop("`selected_obj` must be an object returned by
    ↪  select_polychoric_matrices().")
  if (!is.data.frame(data) && !is.matrix(data))
    stop("`data` must be a data.frame or matrix of raw ordinal responses.")
  if (ncol(data) != ncol(selected_obj$psych$Rpoly))
    stop("`data` columns must match the items in `selected_obj`.")

  correct <- selected_obj$correction_used

  if (verbose)
    message(sprintf(
      paste0(
        "\n--- Step 1: Null distribution (permutations) ---\n",
        "  Model          : %s\n",
        "  Permutations   : %d\n",
        "  Correction     : %s\n",
        "  FA method      : %s  (nfactors = %d)\n",
        "  Null quantile : P%.0f\n"
      ),
      model_name, n_iter_null,
      as.character(correct),
      fm, nfactors_fa,
```

```
    quant * 100
  ))

utils::capture.output(
  parallel_result <- psych::fa.parallel(
    x           = data,
    n.obs       = NULL,
    fm          = fm,
    fa          = "fa",
    nfactors    = nfactors_fa,
    n.iter      = n_iter_null,
    cor         = "poly",
    correct     = correct,
    sim         = FALSE,
    error.bars  = FALSE,
    se.bars     = FALSE,
    show.legend = FALSE,
    quant       = quant,
    SMC         = FALSE,
    plot        = FALSE,
    main        = paste0("Parallel Analysis (", model_name, ")")
  ),
  type = "output"
)

real_eigen <- parallel_result$fa.values
all_cols   <- colnames(parallel_result$values)
sim_cols   <- all_cols[grepl("^F", all_cols)]
sim_matrix <- parallel_result$values[, sim_cols, drop = FALSE]
n_compare  <- min(length(real_eigen), ncol(sim_matrix))
real_eigen <- real_eigen[1:n_compare]
sim_matrix <- sim_matrix[, 1:n_compare, drop = FALSE]
sim_mean   <- apply(sim_matrix, 2, mean)
sim_quant  <- apply(sim_matrix, 2, quantile, probs = quant)

empirical_quantile <- sapply(seq_len(n_compare), function(i) {
  mean(sim_matrix[, i] < real_eigen[i])
})
n_ret <- sum(empirical_quantile >= quant)
n_mar <- sum(real_eigen > sim_mean & empirical_quantile < quant)
n_rej <- sum(real_eigen <= sim_mean)

if (verbose)
  message(sprintf(
    paste0(
      "  Factors retained (PA classic) : %d  (empirical quantile >= P%.0f)\n",
      "  Factors marginal              : %d  (above null mean, below P%.0f)\n",
      "  Factors rejected              : %d  (at or below null mean)\n"
```

```r
    ),
    n_ret, quant * 100,
    n_mar, quant * 100,
    n_rej
  ))

if (verbose)
  message(sprintf(
    paste0(
      "\n--- Step 2: Bootstrap CI (real eigenvalues) ---\n",
      "  Resamples      : %d\n",
      "  CI (one-sided): P%.4g\n",
      "  PSD threshold : %.2f\n"
    ),
    n_boot,
    probs[1] * 100,
    psd_tol
  ))

boot_obj <- run_bootstrap_eigen(
  data                = data,
  n_boot              = n_boot,
  probs               = probs,
  correct_override = correct,
  psd_tol             = psd_tol,
  fm                  = fm,
  nfactors_fa         = nfactors_fa,
  report_every        = report_every,
  parallel            = parallel_boot,
  ncores              = ncores,
  seed                = seed,
  verbose             = verbose
)

boot_sum  <- boot_obj$summary[seq_len(n_compare), , drop = FALSE]
boot_plow <- boot_sum$P_low   # P5 - one-sided 95% CI lower bound

# Retention criterion: one-sided 95% CI lower bound (P5) > null P95
boot_retained <- boot_plow > sim_quant

# % of bootstrap resamples whose eigenvalue exceeds the null P(quant)
boot_support_pct <- sapply(seq_len(n_compare), function(i) {
  round(mean(boot_obj$boot_matrix[, i] > sim_quant[i]) * 100, 1)
})

pa_retained <- real_eigen > sim_quant

ic_low_label <- paste0("Boot_P", probs[1] * 100)
```

```r
  ic_hi_label  <- paste0("Boot_P", probs[2] * 100)

  summary_tbl <- tibble::tibble(
    Factor                                          = seq_len(n_compare),
    Eigenvalue                                      = round(real_eigen,     3),
    Null_mean                                       = round(sim_mean,       3),
    Null_P95                                        = round(sim_quant,      3),
    Boot_mean                                       = round(boot_sum$Mean,  3),
    !!ic_low_label                                 := round(boot_plow,      3),
    !!ic_hi_label                                  := round(boot_sum$P_high, 3),
    `Retained - Horn's PA (P95)`                    = pa_retained,
    `Retained - Bootstrap PA (90% CI, one-sided)` = boot_retained,
    `Non-overlap with permuted eigenvalues (%)`     = boot_support_pct
  )

  n_boot_ret <- sum(boot_retained)

  if (verbose)
    message(sprintf(
      paste0(
        "\n--- Retention summary ---\n",
        "  Horn's PA retained (P%.0f)                : %d\n",
        "  Bootstrap PA retained (90%% CI, one-sided): %d  (%s > P%.0f null)\n"
      ),
      quant * 100, n_ret,
      n_boot_ret, ic_low_label, quant * 100
    ))

  structure(
    list(
      parallel_result = parallel_result,
      boot_obj        = boot_obj,
      model_name      = model_name,
      quant           = quant,
      correction_used = correct,
      summary_tbl     = summary_tbl
    ),
    class = "parallel_bootstrap"
  )
}

print.parallel_bootstrap <- function(x, digits = 3, ...) {
  cat(sprintf(
    "Bootstrap Parallel Analysis \u2014 %s\n",
    x$model_name
  ))
```

```
  cat(sprintf("  Continuity correction : %s\n", as.character(x$correction_used)))
  cat(sprintf("  Null quantile         : P%.0f\n", x$quant * 100))
  ic_cols <- grep("^Boot_P|^P[0-9]", names(x$summary_tbl), value = TRUE)
  cat(sprintf("  Bootstrap CI          : %s \u2013 %s\n", ic_cols[1], ic_cols[2]))
  cat(sprintf("  Bootstrap resamples   : %d valid / %d requested\n",
              x$boot_obj$n_ok, x$boot_obj$n_boot))
  cat("\nRetention table:\n")
  print(x$summary_tbl, row.names = FALSE, digits = digits)
  invisible(x)
}

get_nfact_empirical <- function(parallel_result, quant = 0.95) {

  if (inherits(parallel_result, "parallel_bootstrap"))
    parallel_result <- parallel_result$parallel_result

  real_eigen <- parallel_result$fa.values
  all_cols   <- colnames(parallel_result$values)
  sim_cols   <- all_cols[grepl("^F", all_cols)]
  sim_matrix <- parallel_result$values[, sim_cols, drop = FALSE]

  n_compare  <- min(length(real_eigen), ncol(sim_matrix))
  real_eigen <- real_eigen[1:n_compare]
  sim_matrix <- sim_matrix[, 1:n_compare, drop = FALSE]

  empirical_quantile <- sapply(seq_len(n_compare), function(i) {
    mean(sim_matrix[, i] < real_eigen[i])
  })

  sum(empirical_quantile >= quant)
}

plot_parallel_bootstrap_ggplot <- function(parallel_result,
                                           boot_obj           = NULL,
                                           model_name         = "",
                                           quant              = 0.95,
                                           real_color         = "royalblue",
                                           sim_color          = "red",
                                           not_retained_color = "grey60",
                                           show_boot_mean     = FALSE) {

  if (inherits(parallel_result, "parallel_bootstrap")) {
    pb_obj          <- parallel_result
    parallel_result <- pb_obj$parallel_result
    boot_obj        <- pb_obj$boot_obj
```

```
    if (model_name == "") model_name <- pb_obj$model_name
    quant <- pb_obj$quant
  }

  if (is.null(boot_obj) || !inherits(boot_obj, "bootstrap_eigen"))
    stop(paste0(
      "`boot_obj` must be a 'bootstrap_eigen' object, or pass a ",
      "'parallel_bootstrap' object as the first argument."
    ))

  real_eigen <- parallel_result$fa.values
  n_vars     <- length(real_eigen)

  all_cols   <- colnames(parallel_result$values)
  sim_cols   <- all_cols[grepl("^F", all_cols)]
  sim_matrix <- parallel_result$values[, sim_cols, drop = FALSE]

  n_compare  <- min(n_vars, ncol(sim_matrix))
  real_eigen <- real_eigen[1:n_compare]
  sim_matrix <- sim_matrix[, 1:n_compare, drop = FALSE]

  sim_mean  <- apply(sim_matrix, 2, mean)
  sim_sd    <- apply(sim_matrix, 2, sd)
  sim_quant <- apply(sim_matrix, 2, quantile, probs = quant)

  boot_sum  <- boot_obj$summary[seq_len(n_compare), , drop = FALSE]
  boot_plow <- boot_sum$P_low    # P5 one-sided lower bound

  # Retention: P5 bootstrap > P95 null
  boot_retained <- boot_plow > sim_quant

  n_retained     <- sum( boot_retained)
  n_not_retained <- sum(!boot_retained)

  ic_pct       <- round((boot_obj$probs[2] - boot_obj$probs[1]) * 100)
  ic_low_label <- paste0("P", boot_obj$probs[1] * 100)
  ic_hi_label  <- paste0("P", boot_obj$probs[2] * 100)

  label_null_mean <- "Null mean (permut.)"
  label_null_q    <- paste0("P", quant * 100, " null (permut.)")
  label_band_ret  <- paste0("CI", ic_pct, "% bootstrap \u2014 Retained")
  label_band_not  <- paste0("CI", ic_pct, "% bootstrap \u2014 Not retained")

  df_sim <- data.frame(
    Factor       = seq_len(n_compare),
    Mean         = sim_mean,
    SD_upper     = sim_mean + sim_sd,
    SD_lower     = sim_mean - sim_sd,
```

```r
  Quant_upper = sim_quant,
  Line_mean   = label_null_mean,
  Line_quant  = label_null_q
)

df_real <- data.frame(
  Factor     = seq_len(n_compare),
  Eigenvalue = real_eigen,
  Boot_mean  = boot_sum$Mean,
  Boot_low   = boot_plow,
  Boot_high  = boot_sum$P_high,
  Retained   = boot_retained,
  Band_label = ifelse(boot_retained, label_band_ret, label_band_not)
)

df_ret     <- df_real[df_real$Retained,  , drop = FALSE]
df_not_ret <- df_real[!df_real$Retained, , drop = FALSE]

if (nrow(df_ret) > 0 && nrow(df_not_ret) > 0) {
  transition_row            <- df_ret[nrow(df_ret), , drop = FALSE]
  transition_row$Retained   <- FALSE
  transition_row$Band_label <- label_band_not
  df_not_ret <- rbind(transition_row, df_not_ret)
}

linetype_vals        <- c("solid", "dashed")
names(linetype_vals) <- c(label_null_mean, label_null_q)

fill_vals        <- c(real_color, not_retained_color)
names(fill_vals) <- c(label_band_ret, label_band_not)

fill_vals_used <- fill_vals[names(fill_vals) %in% unique(df_real$Band_label)]

p <- ggplot2::ggplot() +

  # Null distribution +/-1SD band
  ggplot2::geom_ribbon(
    data = df_sim,
    ggplot2::aes(x = Factor, ymin = SD_lower, ymax = SD_upper),
    fill = sim_color, alpha = 0.12
  ) +

  # Bootstrap CI band - not retained (grey)
  {
    if (nrow(df_not_ret) > 0)
      ggplot2::geom_ribbon(
        data = df_not_ret,
        ggplot2::aes(x = Factor, ymin = Boot_low, ymax = Boot_high,
```

```r
                    fill = Band_label),
      alpha = 0.25
    )
  else NULL
} +

# Bootstrap CI band - retained (blue)
{
  if (nrow(df_ret) > 0)
    ggplot2::geom_ribbon(
      data = df_ret,
      ggplot2::aes(x = Factor, ymin = Boot_low, ymax = Boot_high,
                   fill = Band_label),
      alpha = 0.30
    )
  else NULL
} +

# Null mean line
ggplot2::geom_line(
  data = df_sim,
  ggplot2::aes(x = Factor, y = Mean, linetype = Line_mean),
  color = sim_color, linewidth = 0.7
) +

# Null P95 threshold line
ggplot2::geom_line(
  data = df_sim,
  ggplot2::aes(x = Factor, y = Quant_upper, linetype = Line_quant),
  color = sim_color, linewidth = 0.7
) +

# Eigenvalue line - not retained (light grey)
{
  if (nrow(df_not_ret) > 0)
    ggplot2::geom_line(
      data      = df_not_ret,
      ggplot2::aes(x = Factor, y = Eigenvalue),
      color     = "grey70",
      linewidth = 0.8
    )
  else NULL
} +

# Eigenvalue line - retained (blue)
{
  if (nrow(df_ret) > 0)
    ggplot2::geom_line(
```

```r
      data      = df_ret,
      ggplot2::aes(x = Factor, y = Eigenvalue),
      color     = real_color,
      linewidth = 0.8
    )
  else NULL
} +

{
  if (show_boot_mean)
    ggplot2::geom_line(
      data = df_real,
      ggplot2::aes(x = Factor, y = Boot_mean),
      color     = "grey40",
      linewidth = 0.4,
      linetype  = "dotdash"
    )
  else NULL
} +

# Points - retained (blue), not retained (light grey)
ggplot2::geom_point(
  data  = df_real,
  ggplot2::aes(x = Factor, y = Eigenvalue),
  color = ifelse(df_real$Retained, real_color, "grey75"),
  fill  = ifelse(df_real$Retained, real_color, "grey75"),
  shape = 16,
  size  = 2
) +

ggplot2::geom_hline(
  yintercept = 0, linetype = "dotted", color = "grey50", linewidth = 0.4
) +

ggplot2::scale_linetype_manual(
  name   = "Null reference",
  values = linetype_vals
) +

ggplot2::scale_fill_manual(
  name   = paste0("CI", ic_pct, "% bootstrap band"),
  values = fill_vals_used
) +

ggplot2::scale_x_continuous(breaks = seq_len(n_compare)) +

ggplot2::labs(
  title    = paste0("Parallel Analysis + Bootstrap \u2014 ", model_name),
```

```
      subtitle = paste0(
        "Retention: Bootstrap P", boot_obj$probs[1] * 100,
        " (one-sided 95% CI) > P", quant * 100, " null  |  ",
        "Retained: ", n_retained,
        "   Not retained: ", n_not_retained,
        "   |   n_boot = ", boot_obj$n_ok
      ),
      x = "Factor",
      y = "Eigenvalue"
    ) +

    ggplot2::theme_minimal(base_size = 11) +
    ggplot2::theme(
      plot.title       = ggplot2::element_text(face   = "bold",
                                               size   = 11,
                                               margin = ggplot2::margin(b = 6)),
      plot.subtitle    = ggplot2::element_text(color      = "grey35",
                                               size       = 8,
                                               lineheight = 1.4,
                                               margin     = ggplot2::margin(b =
                                                ↪  8)),
      axis.text        = ggplot2::element_text(size = 9),
      axis.title       = ggplot2::element_text(size = 10),
      axis.title.x     = ggplot2::element_text(margin = ggplot2::margin(t = 12)),
      legend.position  = "bottom",
      legend.box       = "vertical",
      legend.text      = ggplot2::element_text(size = 8),
      legend.title     = ggplot2::element_text(size = 8.5, face = "bold"),
      legend.key.size  = ggplot2::unit(0.6, "cm"),
      legend.spacing.y = ggplot2::unit(0.1, "cm"),
      legend.margin    = ggplot2::margin(t = 2, b = 0),
      plot.margin      = ggplot2::margin(t = 20, r = 10, b = 4, l = 8)
    )

  return(p)
}

run_fa_psych_selected <- function(selected_obj,
                                  data,
                                  model_name,
                                  nfactors   = NULL,
                                  pa_obj     = NULL,
                                  quant      = 0.95,
                                  fm         = "minres",
                                  rotate     = "oblimin",
                                  n.iter     = 1000,
```

```r
                                           SMC        = TRUE,
                                           smooth     = FALSE,
                                           warnings   = FALSE,
                                           residuals = FALSE,
                                           scores     = "tenBerge") {

  if (is.null(nfactors)) {
    if (!is.null(pa_obj)) {
      if (inherits(pa_obj, "parallel_bootstrap")) {
        if ("Retained - Bootstrap PA (90% CI, one-sided)" %in%
          ↪  names(pa_obj$summary_tbl)) {
          nfactors <- sum(pa_obj$summary_tbl[["Retained - Bootstrap PA (90% CI,
↪  one-sided)"]])
        } else if ("Combined_retained" %in% names(pa_obj$summary_tbl)) {
          nfactors <- sum(pa_obj$summary_tbl$Combined_retained)
        } else {
          stop("Cannot determine number of factors from `pa_obj$summary_tbl`.
            ↪  Check column names.")
        }
      } else {
        nfactors <- get_nfact_empirical(pa_obj, quant = quant)
      }
    } else {
      stop("Please provide either `nfactors` or a valid `pa_obj`.")
    }
  }

  fa_fit <- psych::fa(
    r          = selected_obj$psych$Rpoly,
    nfactors   = nfactors,
    n.obs      = nrow(data),
    n.iter     = n.iter,
    fm         = fm,
    rotate     = rotate,
    SMC        = SMC,
    smooth     = smooth,
    warnings   = warnings,
    residuals = residuals,
    scores     = scores
  )

  list(
    model_name = model_name,
    nfactors   = nfactors,
    fa         = fa_fit
  )
}
```

```r
extract_fa_fit_table <- function(fa_obj, digits = 3) {

  fa_fit <- if (!is.null(fa_obj$fa)) fa_obj$fa else fa_obj

  model_label <- if (!is.null(fa_obj$model_name)) {
    fa_obj$model_name
  } else {
    NA_character_
  }

  n_factors <- if (!is.null(fa_obj$nfactors)) {
    fa_obj$nfactors
  } else if (!is.null(fa_fit$factors)) {
    fa_fit$factors
  } else {
    NA_integer_
  }

  chi_square <- unname(fa_fit$STATISTIC)
  p_value <- unname(fa_fit$PVAL)
  tli <- unname(fa_fit$TLI)
  rmsea <- unname(fa_fit$RMSEA[1])
  rmsr <- unname(fa_fit$rms)

  prop_var_text <- if (!is.null(fa_fit$Vaccounted)) {
    vaccounted   <- fa_fit$Vaccounted
    prop_var     <- vaccounted["Proportion Var", ]
    factor_names <- colnames(vaccounted)

    paste(
      paste0(factor_names, ": ", format(round(prop_var, digits), nsmall =
       ↪  digits)),
      collapse = "; "
    )
  } else {
    NA_character_
  }

  classify_chisq <- function(p) {
    if (is.na(p)) return(NA_character_)
    if (p >= 0.05) return("Excellent model fit")
    "Poor model fit"
  }

  classify_tli <- function(x) {
    if (is.na(x)) return(NA_character_)
    if (x >= 0.95) return("Excellent model fit")
```

```r
  "Poor model fit"
}

classify_rmsea <- function(x) {
  if (is.na(x)) return(NA_character_)
  if (x <= 0.05) return("Good residual fit")
  if (x <= 0.06) return("Acceptable residual fit")
  "Poor residual fit"
}

classify_rmsr <- function(x) {
  if (is.na(x)) return(NA_character_)
  if (x <= 0.05) return("Good residual fit")
  if (x <= 0.06) return("Acceptable residual fit")
  if (x <= 0.07) return("Adequate residual fit")
  if (x <= 0.08) return("Marginal residual fit")
  "Poor residual fit"
}

model_fit_rank <- c(
  "Excellent model fit" = 1,
  "Poor model fit" = 2
)

error_fit_rank <- c(
  "Good residual fit" = 1,
  "Acceptable residual fit" = 2,
  "Adequate residual fit" = 3,
  "Marginal residual fit" = 4,
  "Poor residual fit" = 5
)

combine_fit_range <- function(categories, rank_map, suffix_label) {
  categories <- categories[!is.na(categories)]

  if (length(categories) == 0) {
    return(NA_character_)
  }

  if (length(unique(categories)) == 1) {
    return(tolower(categories[1]))
  }

  ranks <- rank_map[categories]
  best_cat <- categories[which.min(ranks)]
  worst_cat <- categories[which.max(ranks)]

  best_prefix <- tolower(sub(paste0(" ", suffix_label, "$"), "", best_cat))
```

```
    worst_prefix <- tolower(sub(paste0(" ", suffix_label, "$"), "", worst_cat))

    paste0(worst_prefix, "-to-", best_prefix, " ", tolower(suffix_label))
  }

  chisq_fit <- classify_chisq(p_value)
  tli_fit <- classify_tli(tli)
  rmsea_fit <- classify_rmsea(rmsea)
  rmsr_fit <- classify_rmsr(rmsr)

  overall_model_fit <- combine_fit_range(
    categories = c(chisq_fit, tli_fit),
    rank_map = model_fit_rank,
    suffix_label = "model fit"
  )

  overall_error_fit <- combine_fit_range(
    categories = c(rmsea_fit, rmsr_fit),
    rank_map = error_fit_rank,
    suffix_label = "residual fit"
  )

  tibble::tibble(
    Model = model_label,
    `No. factors` = n_factors,
    `Proportion Var` = prop_var_text,
    `Chi-square` = chi_square,
    `p-value` = p_value,
    TLI = tli,
    RMSEA = rmsea,
    RMSR = rmsr,
    `Model fit` = overall_model_fit,
    `Error fit` = overall_error_fit
  )
}
```

##### 4.5.2 EGA dimensionality functions

```
run_bootega_selected <- function(data,
                                 selected_obj,
                                 model_name = NULL,
                                 iter = 1000,
                                 seed = 123,
                                 ncores = 1,
                                 plot.itemStability = FALSE,
                                 typicalStructure = FALSE,
```

```
                                 plot.typicalStructure = FALSE,
                                 plot.EGA = FALSE,
                                 verbose = TRUE) {

  boot <- EGAnet::bootEGA(
    data = data,
    iter = iter,
    type = "resampling",
    corr = "auto",
    na.data = selected_obj$eganet$method$na.data,
    empty.method = selected_obj$eganet$method$empty.method,
    empty.value = selected_obj$eganet$method$empty.value,
    model = "glasso",
    algorithm = "walktrap",
    uni.method = "louvain",
    plot.itemStability = plot.itemStability,
    typicalStructure = typicalStructure,
    plot.typicalStructure = plot.typicalStructure,
    ncores = ncores,
    seed = seed,
    verbose = verbose
  )

  ega_plot <- NULL

  if (plot.EGA) {
    ega_plot <- plot(boot$EGA)
    print(ega_plot)
  }

  list(
    model_name = model_name,
    boot       = boot,
    ega_plot   = ega_plot
  )
}

extract_ndim_frequency <- function(boot, model) {

  freq_df <- boot$frequency %>%
    stats::setNames(c("n_factors", "frequency"))

  freq_df %>%
    dplyr::mutate(
      percentage = frequency * 100,
      Model = model
    ) %>%
```

```r
    dplyr::select(Model, n_factors, frequency, percentage) %>%
    dplyr::arrange(n_factors)
}

build_ndim_frequency_table <- function(boot_list,
                                       n_factors_range = NULL) {

  freq_tbl <- purrr::imap_dfr(
    boot_list,
    ~ extract_ndim_frequency(
      boot = .x,
      model = .y
    )
  )

  if (is.null(n_factors_range)) {
    n_factors_range <- seq_len(max(freq_tbl$n_factors, na.rm = TRUE))
  }

  freq_tbl %>%
    tidyr::complete(
      Model,
      n_factors = n_factors_range,
      fill = list(
        frequency = 0,
        percentage = 0
      )
    ) %>%
    dplyr::arrange(Model, n_factors)
}

plot_ndim_frequency <- function(freq_tbl,
                                palette = NULL,
                                y_limit = NULL,
                                facet = TRUE,
                                base_size = 10,
                                label_size = 3) {

  n_levels <- length(sort(unique(freq_tbl$n_factors)))

  if (is.null(palette)) {
    palette <- paletteer::paletteer_c("grDevices::Cold", n_levels)
  }

  p <- ggplot(
    freq_tbl,
```

```r
    aes(
      x = factor(n_factors),
      y = percentage,
      fill = factor(n_factors)
    )
  ) +
    geom_col(width = 0.7) +
    geom_text(
      aes(label = paste0(round(percentage, 1), "%")),
      vjust = -0.3,
      size = label_size
    ) +
    scale_fill_manual(
      values = palette,
      name = "Number of dimensions"
    ) +
    labs(
      x = "Number of dimensions",
      y = "Percentage of bootstrap samples"
    ) +
    theme_minimal(base_size = base_size) +
    theme(
      legend.position = "none"
    )

  if (!is.null(y_limit)) {
    p <- p +
      scale_y_continuous(
        labels = scales::percent_format(scale = 1),
        limits = c(0, y_limit)
      )
  } else {
    p <- p +
      scale_y_continuous(
        labels = scales::percent_format(scale = 1)
      )
  }

  if (facet && dplyr::n_distinct(freq_tbl$Model) > 1) {
    p <- p +
      facet_wrap(~ Model) +
      theme(
        strip.text = ggplot2::element_text(face = "bold")
      )
  }

  p
}
```

```r
summarise_ndim_frequency <- function(freq_tbl,
                                     model_name = NULL,
                                     top_n = 4,
                                     digits = 1) {
  model_label <- if (!is.null(model_name)) {
    model_name
  } else {
    unique(freq_tbl$Model)[1]
  }

  ordered_freq <- freq_tbl %>%
    dplyr::filter(Model == model_label) %>%
    dplyr::filter(frequency > 0) %>%
    dplyr::arrange(dplyr::desc(frequency), n_factors)

  top_freq <- ordered_freq %>%
    dplyr::slice_head(n = top_n)

  mode_solution   <- top_freq$n_factors[1]
  mode_frequency  <- top_freq$frequency[1]
  mode_percentage <- top_freq$percentage[1]

  ranked_solutions <- top_freq %>%
    dplyr::mutate(
      Solution = paste0(
        n_factors,
        " dimensions: ",
        round(percentage, digits),
        "%"
      )
    ) %>%
    dplyr::pull(Solution) %>%
    paste(collapse = "; ")

  get_difference <- function(rank_position) {
    if (nrow(top_freq) >= rank_position) {
      diff_value <- mode_percentage - top_freq$percentage[rank_position]
      paste0(round(diff_value, digits), "%")
    } else {
      NA_character_
    }
  }

  tibble::tibble(
    Model = model_label,
    `Modal solution` = paste0(
```

```
      mode_solution,
      " dimensions (",
      round(mode_percentage, digits),
      "%)"
    ),
    `Ranked dimensional solutions` = ranked_solutions,
    `Difference 1st-2nd` = get_difference(2),
    `Difference 1st-3rd` = get_difference(3),
    `Difference 1st-4th` = get_difference(4)
  )
}
```

```
extract_structural_consistency <- function(boot,
                                           model,
                                           accuracy = 0.1) {

  sc <- EGAnet::dimensionStability(
    boot,
    IS.plot = FALSE
  )$dimension.stability$structural.consistency

  sc_tbl <- tibble::tibble(
    Model = model,
    Dimension = paste0("Dim ", seq_along(sc)),
    Structural_consistency_pct = scales::percent(sc, accuracy = accuracy)
  ) %>%
    tidyr::pivot_wider(
      names_from = Dimension,
      values_from = Structural_consistency_pct
    )

  sc_tbl
}
```

##### 4.5.3 Dimensionality performance criteria functions

```
build_dimensional_decision_table <- function(model_name,
                                             pa_obj                     = NULL,
                                             pa_n_factors               = NULL,
                                             quant                      = 0.95,
                                             fa_fit_table,
                                             ndim_summary,
```

```
                                                structural_consistency,
                                                previous_model_comparison = NULL,
                                                researcher_judgement      = rep("",
                                                 ↪  5)) {

  if (is.null(pa_n_factors)) {
    if (!is.null(pa_obj)) {
      if (!inherits(pa_obj, "parallel_bootstrap"))
        stop("`pa_obj` must be a 'parallel_bootstrap' object from
         ↪  run_parallel_bootstrap().")
      if ("Retained - Bootstrap PA (90% CI, one-sided)" %in%
       ↪  names(pa_obj$summary_tbl)) {
        pa_n_factors <- sum(pa_obj$summary_tbl[["Retained - Bootstrap PA (90% CI,
↪  one-sided)"]])
      } else if ("Combined_retained" %in% names(pa_obj$summary_tbl)) {
        pa_n_factors <- sum(pa_obj$summary_tbl$Combined_retained)
      } else {
        stop("Cannot determine number of factors from `pa_obj$summary_tbl`. Check
         ↪  column names.")
      }
    } else {
      pa_n_factors <- NA_integer_
    }
  }

  fa_row <- fa_fit_table %>%
    dplyr::filter(Model == model_name) %>%
    dplyr::slice(1)

  ndim_row <- ndim_summary %>%
    dplyr::filter(Model == model_name) %>%
    dplyr::slice(1)

  sc_text <- structural_consistency %>%
    dplyr::filter(Model == model_name) %>%
    dplyr::select(-Model) %>%
    tidyr::pivot_longer(
      cols      = dplyr::everything(),
      names_to  = "Dimension",
      values_to = "Structural consistency"
    ) %>%
    dplyr::mutate(Text = paste0(Dimension, ": ", `Structural consistency`)) %>%
    dplyr::pull(Text) %>%
    paste(collapse = "; ")

  if (length(researcher_judgement) != 5) {
    researcher_judgement <- rep("", 5)
  }
```

```r
  pa_label <- paste0("Bootstrap PA suggested ", pa_n_factors, " factors")

  tibble::tibble(
    Model     = model_name,
    Criterion = c(
      "1. Bootstrap PA-EGA dimensional convergence",
      "2. FA dimensional performance",
      "3. EGA dimensional stability",
      "4. Improvement over previous model",
      "5. Model retention"
    ),
    `Objective evidence` = c(
      paste0(
        pa_label,
        "; EGA modal solution was ", ndim_row$`Modal solution`, "."
      ),
      paste0(
        "Proportion Var. = ", fa_row$`Proportion Var`,
        "; model fit = ", fa_row$`Model fit`,
        "; error fit = ", fa_row$`Error fit`, "."
      ),
      paste0(
        "Modal solution = ", ndim_row$`Modal solution`,
        "; differences: 1st-2nd = ", ndim_row$`Difference 1st-2nd`,
        ", 1st-3rd = ", ndim_row$`Difference 1st-3rd`,
        ", 1st-4th = ", ndim_row$`Difference 1st-4th`,
        "; structural consistency = ", sc_text, "."
      ),
      ifelse(
        is.null(previous_model_comparison),
        "Not applicable for the first evaluated model.",
        previous_model_comparison
      ),
      "Final model decision based on the previous dimensional criteria (1-4)"
    ),
    `Researcher judgement` = researcher_judgement
  )
}
```

##### 4.5.4 FA item diagnostic functions

```r
build_fa_item_table <- function(fa_obj,
                                model_name = NULL,
                                item_ids = emfra_item_ids,
                                item_labels = emfra_item_labels,
```

```r
                                item_labels_short = emfra_item_labels_short,
                                loading_cutoff = 0.30,
                                primary_loading_cutoff = 0.50,
                                communality_cutoff = 0.30,
                                digits = 3) {

  fa_fit <- if (!is.null(fa_obj$fa)) fa_obj$fa else fa_obj
  model_label <- if (!is.null(model_name)) {
    model_name
  } else if (!is.null(fa_obj$model_name)) {
    fa_obj$model_name
  } else {
    NA_character_
  }

  fa_row_names <- rownames(fa_fit$loadings)
  lookup_key <- if (all(fa_row_names %in% item_ids)) item_ids else
↪  item_labels_short

  item_lookup <- tibble::tibble(
    Item = lookup_key,
    `Item number` = stringr::str_extract(item_labels, "^Item\\s+\\d+"),
    `Item name` = item_labels_short
    )

  loadings_mat <- as.data.frame(unclass(fa_fit$loadings))
  loadings_mat <- tibble::rownames_to_column(loadings_mat, var = "Item")

  communality_values <- fa_fit$communality

  item_tbl <- loadings_mat %>%
    dplyr::mutate(
      Communality = communality_values
    ) %>%
    dplyr::left_join(
      item_lookup,
      by = "Item"
    )

  loading_cols <- setdiff(
    colnames(item_tbl),
    c("Item", "Item number", "Item name", "Communality")
  )

  item_tbl <- item_tbl %>%
    dplyr::rowwise() %>%
    dplyr::mutate(
```

```r
    .abs_loadings = list(abs(c_across(dplyr::all_of(loading_cols)))),

    .primary = max(unlist(.abs_loadings), na.rm = TRUE),

    .secondary = {
      tmp <- sort(unlist(.abs_loadings), decreasing = TRUE)
      if (length(tmp) >= 2) tmp[2] else NA_real_
    },

    `Low loadings` = ifelse(
      all(unlist(.abs_loadings) < loading_cutoff),
      "Yes",
      "No"
    ),

    `High cross-loadings` = ifelse(
      !is.na(.secondary) &&
        .primary >= primary_loading_cutoff &&
        .secondary >= primary_loading_cutoff,
      "Yes",
      "No"
    ),

    `Low communalities` = ifelse(
      !is.na(Communality) && Communality < communality_cutoff,
      "Yes",
      "No"
    )
  ) %>%
  dplyr::ungroup() %>%
  dplyr::select(
    `Item number`,
    `Item name`,
    Communality,
    dplyr::all_of(loading_cols),
    `Low loadings`,
    `High cross-loadings`,
    `Low communalities`
  ) %>%
  dplyr::mutate(
    dplyr::across(
      c(Communality, dplyr::all_of(loading_cols)),
      ~ round(.x, digits)
    ),
    Model = model_label,
    .before = 1
  )
```

```
  item_tbl
}

plot_fa_loadings <- function(fa_item_table,
                             model_title    = NULL,
                             primary_cutoff = 0.30,
                             text_size      = 3.0,
                             base_size      = 11) {

  non_loading_cols <- c("Model", "Item number", "Item name", "Communality",
                        "Low loadings", "High cross-loadings", "Low
                         ↪  communalities")
  loading_cols <- setdiff(colnames(fa_item_table), non_loading_cols)

  title_str <- if (!is.null(model_title)) {
    model_title
  } else if ("Model" %in% colnames(fa_item_table)) {
    paste0("Factor Loadings - ", unique(fa_item_table$Model)[1])
  } else {
    "Factor Loadings"
  }

  plot_data <- fa_item_table %>%
    dplyr::select(`Item name`, dplyr::all_of(loading_cols)) %>%
    tidyr::pivot_longer(
      cols      = dplyr::all_of(loading_cols),
      names_to  = "Factor",
      values_to = "Loading"
    ) %>%
    dplyr::mutate(
      `Item name` = factor(`Item name`, levels = rev(fa_item_table$`Item name`)),
      is_primary  = abs(Loading) >= primary_cutoff
    )

  ggplot(plot_data, aes(x = Factor, y = `Item name`, fill = Loading)) +
    geom_tile(color = "white", linewidth = 0.3) +
    geom_tile(
      data  = dplyr::filter(plot_data, is_primary),
      color = "black", linewidth = 0.4, fill = NA
    ) +
    geom_text(aes(label = sprintf("%.2f", Loading)), size = text_size) +
    scale_fill_gradient2(
      low      = "blue",
      mid      = "white",
      high     = "red",
      midpoint = 0,
      limits   = c(-1, 1),
      name     = "Loading"
```

```
    ) +
    labs(
      x     = "Factor",
      y     = NULL,
      title = title_str
    ) +
    theme_minimal(base_size = base_size) +
    theme(
      panel.grid  = element_blank(),
      axis.text.y = element_text(size = base_size - 2),
      plot.title  = element_text(hjust = 0.5, face = "bold", size = base_size)
    )
}

build_fa_candidate_items_table <- function(fa_item_table,
                                           digits = 3) {

  loading_cols <- setdiff(
    colnames(fa_item_table),
    c(
      "Model",
      "Item",
      "Item number",
      "Item name",
      "Communality",
      "Low loadings",
      "High cross-loadings",
      "Low communalities"
    )
  )

  candidate_tbl <- fa_item_table %>%
    dplyr::filter(
      `Low loadings` == "Yes" |
        `High cross-loadings` == "Yes" |
        `Low communalities` == "Yes"
    )

  if (nrow(candidate_tbl) == 0) {
    return(
      tibble::tibble(
        Model = character(),
        `Item number` = character(),
        `Item name` = character(),
        `Low factor loadings reason` = character(),
        `High cross-loadings reason` = character(),
        `Low communality reason` = character()
```

```r
    )
  )
}

loading_matrix <- as.matrix(candidate_tbl[, loading_cols, drop = FALSE])
abs_loading_matrix <- abs(loading_matrix)

primary_idx <- apply(
  abs_loading_matrix,
  1,
  function(x) order(x, decreasing = TRUE)[1]
)

secondary_idx <- apply(
  abs_loading_matrix,
  1,
  function(x) {
    ord <- order(x, decreasing = TRUE)
    if (length(ord) >= 2) ord[2] else NA_integer_
  }
)

row_idx <- seq_len(nrow(candidate_tbl))

primary_factor <- loading_cols[primary_idx]
primary_loading <- loading_matrix[cbind(row_idx, primary_idx)]

secondary_factor <- ifelse(
  !is.na(secondary_idx),
  loading_cols[secondary_idx],
  NA_character_
)

secondary_loading <- ifelse(
  !is.na(secondary_idx),
  loading_matrix[cbind(row_idx, secondary_idx)],
  NA_real_
)

candidate_tbl %>%
  dplyr::mutate(
    .primary_factor = primary_factor,
    .primary_loading = primary_loading,
    .secondary_factor = secondary_factor,
    .secondary_loading = secondary_loading,

    `Low factor loadings reason` = ifelse(
      `Low loadings` == "Yes",
```

```r
        paste0(
          "Highest abs. loading = ",
          .primary_factor,
          ": ",
          format(round(.primary_loading, digits), nsmall = digits)
        ),
        "No"
      ),

      `High cross-loadings reason` = ifelse(
        `High cross-loadings` == "Yes",
        paste0(
          "Primary abs. loading = ",
          .primary_factor,
          ": ",
          format(round(.primary_loading, digits), nsmall = digits),
          "; secondary abs. loading = ",
          .secondary_factor,
          ": ",
          format(round(.secondary_loading, digits), nsmall = digits)
        ),
        "No"
      ),

      `Low communality reason` = ifelse(
        `Low communalities` == "Yes",
        paste0(
          "Communality = ",
          format(round(Communality, digits), nsmall = digits)
        ),
        "No"
      )
    ) %>%
    dplyr::select(
      Model,
      `Item number`,
      `Item name`,
      `Low factor loadings reason`,
      `High cross-loadings reason`,
      `Low communality reason`
    )
}
```

##### 4.5.5 EGA item diagnostic functions

```r
extract_item_stability <- function(boot,
                                   model,
                                   item_ids = emfra_item_ids,
                                   item_labels = emfra_item_labels,
                                   item_labels_short = emfra_item_labels_short,
                                   stability_cutoff = 0.75,
                                   accuracy = 0.1) {

  item_lookup <- dplyr::bind_rows(
    tibble::tibble(
      Item = item_ids,
      `Item number` = stringr::str_extract(item_labels, "^Item\\s+\\d+"),
      `Item name` = item_labels_short
    ),
    tibble::tibble(
      Item = item_labels_short,
      `Item number` = stringr::str_extract(item_labels, "^Item\\s+\\d+"),
      `Item name` = item_labels_short
    ),
    tibble::tibble(
      Item = item_labels,
      `Item number` = stringr::str_extract(item_labels, "^Item\\s+\\d+"),
      `Item name` = item_labels_short
    )
  ) %>%
    dplyr::distinct(Item, .keep_all = TRUE)

  stability_obj <- EGAnet::dimensionStability(
    boot,
    IS.plot = FALSE
  )

  emp_dims <- stability_obj$item.stability$item.stability$empirical.dimensions
  community_membership <- boot$EGA$wc

  stability_df <- tibble::tibble(
    Item          = names(emp_dims),
    Replicability = as.numeric(emp_dims)
  ) %>%
    dplyr::mutate(
      `Assigned community` = paste0("Dim ", community_membership[Item])
    ) %>%
    dplyr::arrange(
      dplyr::desc(`Assigned community`),
      Replicability
```

```r
  ) %>%
  dplyr::mutate(
    Item = factor(Item, levels = Item)
  )

  fixed_colors <- c(
  "Dim 1" = "indianred1",
  "Dim 2" = "lightskyblue",
  "Dim 3" = "darkolivegreen2"
)

comm_levels <- unique(stability_df$`Assigned community`)
extra_comm  <- setdiff(comm_levels, names(fixed_colors))

if (length(extra_comm) > 0) {
  extra_colors <- scales::hue_pal()(length(extra_comm))
  names(extra_colors) <- extra_comm
  fixed_colors <- c(fixed_colors, extra_colors)
}

base_colors  <- fixed_colors[comm_levels]
light_colors <- scales::alpha(base_colors, 0.35)

stability_plot <- ggplot2::ggplot(
  stability_df,
  ggplot2::aes(
    x      = Item,
    y      = Replicability,
    colour = `Assigned community`,
    fill   = `Assigned community`
  )
) +
  ggplot2::geom_segment(
    ggplot2::aes(xend = Item, y = 0, yend = Replicability),
    linewidth = 0.6
  ) +
  ggplot2::geom_point(
    shape  = 21,
    size   = 3.5,
    stroke = 1
  ) +
  ggplot2::geom_text(
    ggplot2::aes(label = sprintf("%.2f", Replicability)),
    hjust  = -0.4,
    size   = 2.8,
    colour = "black"
  ) +
  ggplot2::scale_colour_manual(values = base_colors) +
```

```
    ggplot2::scale_fill_manual(values = light_colors) +
    ggplot2::coord_flip(clip = "off") +
    ggplot2::scale_y_continuous(expand = ggplot2::expansion(mult = c(0, 0.12))) +
    ggplot2::labs(
      x      = NULL,
      y      = "Replicability",
      colour = "Community",
      fill   = "Community"
    ) +
    ggplot2::theme_minimal()

  item_tbl <- stability_df %>%
    dplyr::mutate(
      `Replicability (%)` = scales::percent(
        Replicability,
        accuracy = accuracy
      ),
      `Below replicability threshold` = ifelse(
        Replicability <= stability_cutoff,
        "Yes",
        "No"
      ),
      Model = model,
      .before = 1
    ) %>%
    dplyr::left_join(
      item_lookup,
      by = "Item"
    ) %>%
    dplyr::arrange(
      `Assigned community`,
      dplyr::desc(Replicability)
    ) %>%
    dplyr::select(
      Model,
      `Item number`,
      `Item name`,
      `Assigned community`,
      `Replicability (%)`,
      `Below replicability threshold`
    )

  list(
    table = item_tbl,
    plot  = stability_plot
  )
}
```

```r
build_ega_redundancy_table <- function(data,
                                       selected_obj,
                                       model_name,
                                       item_ids = emfra_item_ids,
                                       item_labels = emfra_item_labels,
                                       item_labels_short =
                                        ↪  emfra_item_labels_short,
                                       uva.method = "MBR",
                                       cut.off = 0.25,
                                       reduce = FALSE,
                                       reduce.method = "latent",
                                       auto = TRUE,
                                       report_cutoff = 0.20,
                                       moderate_cutoff = 0.25,
                                       large_cutoff = 0.30,
                                       digits = 3,
                                       verbose = FALSE) {

  item_lookup <- dplyr::bind_rows(
    tibble::tibble(
      Item = item_ids,
      `Item label` = item_labels
    ),
    tibble::tibble(
      Item = item_labels_short,
      `Item label` = item_labels
    ),
    tibble::tibble(
      Item = item_labels,
      `Item label` = item_labels
    )
  ) %>%
    dplyr::distinct(Item, .keep_all = TRUE)

  R_eganet <- selected_obj$eganet$Rpoly

  key_items <- colnames(R_eganet)

  if (is.null(key_items)) {
    key_items <- colnames(data)
  }

  uva_obj <- EGAnet::UVA(
    data = R_eganet,
    n = nrow(data),
    key = key_items,
    uva.method = uva.method,
```

```r
    cut.off = cut.off,
    reduce = reduce,
    reduce.method = reduce.method,
    auto = auto,
    verbose = verbose
  )

  empty_tbl <- tibble::tibble(
    Model = character(),
    `Item 1` = character(),
    `Item 2` = character(),
    `wTO value` = numeric(),
    `wTO cut-off` = character(),
    `Redundancy level` = character()
  )

  if (is.null(uva_obj$wto$pairwise) || nrow(uva_obj$wto$pairwise) == 0) {
    return(
      list(
        model_name = model_name,
        correction_used = selected_obj$correction_used,
        uva = uva_obj,
        table = empty_tbl
      )
    )
  }

  redundancy_tbl <- uva_obj$wto$pairwise %>%
    dplyr::filter(wto >= report_cutoff) %>%
    dplyr::left_join(
      item_lookup %>%
        dplyr::rename(
          node_i = Item,
          `Item 1` = `Item label`
        ),
      by = "node_i"
    ) %>%
    dplyr::left_join(
      item_lookup %>%
        dplyr::rename(
          node_j = Item,
          `Item 2` = `Item label`
        ),
      by = "node_j"
    ) %>%
    dplyr::mutate(
      `wTO value` = round(wto, digits),
```

```r
      `Redundancy level` = dplyr::case_when(
        wto >= large_cutoff    ~ "Large-to-very large",
        wto >= moderate_cutoff ~ "Moderate-to-large",
        TRUE                   ~ "Small-to-moderate"
      ),

      `wTO cut-off` = dplyr::case_when(
        wto >= large_cutoff    ~ paste0("  ", large_cutoff),
        wto >= moderate_cutoff ~ paste0("  ", moderate_cutoff),
        TRUE                   ~ paste0("  ", report_cutoff)
      ),

      Model = model_name
    ) %>%
    dplyr::select(
      Model,
      `Item 1`,
      `Item 2`,
      `wTO value`,
      `wTO cut-off`,
      `Redundancy level`
    ) %>%
    dplyr::arrange(dplyr::desc(`wTO value`))

  list(
    model_name = model_name,
    correction_used = selected_obj$correction_used,
    uva = uva_obj,
    table = redundancy_tbl
  )
}

run_item_diagnostics_selected <- function(data,
                                          selected_obj,
                                          model_name,
                                          item_ids          = emfra_item_ids,
                                          item_labels       = emfra_item_labels,
                                          item_labels_short =
                                           ↪  emfra_item_labels_short,
                                          iter              = 1000,
                                          corr              = "auto",
                                          model             = "glasso",
                                          algorithm         = "walktrap",
                                          uni.method        = "louvain",
                                          smooth            = FALSE,
                                          ncores            = 1,
                                          seed              = 123) {
```

```
empty_tbl <- tibble::tibble(
  Model         = character(),
  `Item number` = character(),
  `Item name`   = character(),
  Diagnostic    = character()
)

item_lookup <- dplyr::bind_rows(
  tibble::tibble(
    Item          = item_ids,
    `Item number` = stringr::str_extract(item_labels, "^Item\\s+\\d+"),
    `Item name`   = item_labels_short
  ),
  tibble::tibble(
    Item          = item_labels_short,
    `Item number` = stringr::str_extract(item_labels, "^Item\\s+\\d+"),
    `Item name`   = item_labels_short
  ),
  tibble::tibble(
    Item          = item_labels,
    `Item number` = stringr::str_extract(item_labels, "^Item\\s+\\d+"),
    `Item name`   = item_labels_short
  )
) %>%
  dplyr::distinct(Item, .keep_all = TRUE)

grDevices::pdf(NULL)
item_diag <- tryCatch(
  EGAnet::itemDiagnostics(
    data         = data,
    iter         = iter,
    type         = "resampling",
    corr         = corr,
    na.data      = selected_obj$eganet$method$na.data,
    model        = model,
    algorithm    = algorithm,
    uni.method   = uni.method,
    empty.method = selected_obj$eganet$method$empty.method,
    empty.value  = selected_obj$eganet$method$empty.value,
    smooth       = smooth,
    ncores       = ncores,
    seed         = seed
  ),
  error = function(e) {
    message("itemDiagnostics skipped for ", model_name, ": ",
     ↪  conditionMessage(e))
    NULL
```

```
  }
)
grDevices::dev.off()

if (is.null(item_diag)) {
  return(list(
    model_name = model_name,
    item_diag  = NULL,
    skipped    = TRUE,
    table      = empty_tbl
  ))
}

if (!is.data.frame(item_diag$diagnostics) ||
    nrow(item_diag$diagnostics) == 0) {
  message("No item diagnostics issues detected for ", model_name,
          ". Returning empty table.")
  return(list(
    model_name = model_name,
    item_diag  = item_diag,
    skipped    = FALSE,
    table      = empty_tbl
  ))
}

target_flags <- c("MiD", "TwD", "MuD")
flag_labels  <- c(
  "MiD" = "Minor dimension",
  "TwD" = "Two-item dimension",
  "MuD" = "Multidimensional"
)

diagnostics_tbl <- item_diag$diagnostics %>%
  tibble::rownames_to_column("Item") %>%
  tibble::as_tibble() %>%
  dplyr::mutate(
    .flags      = stringr::str_split(stringr::str_trim(diagnostic), "\\s+"),
    .target     = purrr::map(.flags, ~ intersect(.x, target_flags)),
    .has_target = purrr::map_lgl(.target, ~ length(.x) > 0)
  ) %>%
  dplyr::filter(.has_target) %>%
  dplyr::mutate(
    Diagnostic = purrr::map_chr(
      .target,
      ~ paste(flag_labels[.x], collapse = ", ")
    )
  ) %>%
  dplyr::left_join(item_lookup, by = "Item") %>%
```

```r
    dplyr::mutate(
      Model        = model_name,
      .item_order = as.numeric(stringr::str_extract(`Item number`, "\\d+"))
    ) %>%
    dplyr::arrange(.item_order) %>%
    dplyr::select(Model, `Item number`, `Item name`, Diagnostic)

  if (nrow(diagnostics_tbl) == 0) {
    diagnostics_tbl <- empty_tbl
  }

  list(
    model_name = model_name,
    item_diag  = item_diag,
    skipped    = FALSE,
    table      = diagnostics_tbl
  )
}

extract_ega_community_loadings <- function(boot,
                                           model,
                                           item_ids = emfra_item_ids,
                                           item_labels = emfra_item_labels,
                                           item_labels_short =
                                            ↪  emfra_item_labels_short,
                                           loading.method = "revised",
                                           scaling = 2,
                                           rotation = "oblimin",
                                           loading_cutoff = 0.30,
                                           primary_loading_cutoff = 0.50,
                                           digits = 3) {

  item_lookup <- dplyr::bind_rows(
    tibble::tibble(
      Item = item_ids,
      `Item number` = stringr::str_extract(item_labels, "^Item\\s+\\d+"),
      `Item name` = item_labels_short
    ),
    tibble::tibble(
      Item = item_labels_short,
      `Item number` = stringr::str_extract(item_labels, "^Item\\s+\\d+"),
      `Item name` = item_labels_short
    ),
    tibble::tibble(
      Item = item_labels,
      `Item number` = stringr::str_extract(item_labels, "^Item\\s+\\d+"),
      `Item name` = item_labels_short
```

```r
  )
) %>%
  dplyr::distinct(Item, .keep_all = TRUE)

net_loads_obj <- EGAnet::net.loads(
  boot$EGA,
  loading.method = loading.method,
  scaling = scaling,
  rotation = rotation
)

loading_mat <- net_loads_obj$rotated$loadings

loading_tbl <- as.data.frame(loading_mat) %>%
  tibble::rownames_to_column("Item") %>%
  tibble::as_tibble()

loading_cols <- setdiff(
  colnames(loading_tbl),
  "Item"
)

loading_tbl <- loading_tbl %>%
  dplyr::left_join(
    item_lookup,
    by = "Item"
  ) %>%
  dplyr::rowwise() %>%
  dplyr::mutate(
    .abs_loadings = list(
      abs(c_across(dplyr::all_of(loading_cols)))
    ),

    .primary = max(
      unlist(.abs_loadings),
      na.rm = TRUE
    ),

    .secondary = {
      tmp <- sort(
        unlist(.abs_loadings),
        decreasing = TRUE
      )

      if (length(tmp) >= 2) {
        tmp[2]
      } else {
        NA_real_
```

```r
        }
      },

      `Low loadings` = ifelse(
        all(unlist(.abs_loadings) < loading_cutoff),
        "Yes",
        "No"
      ),

      `High cross-loadings` = ifelse(
        !is.na(.secondary) &&
          .primary >= primary_loading_cutoff &&
          .secondary >= primary_loading_cutoff,
        "Yes",
        "No"
      )
    ) %>%
    dplyr::ungroup() %>%
    dplyr::select(
      `Item number`,
      `Item name`,
      dplyr::all_of(loading_cols),
      `Low loadings`,
      `High cross-loadings`
    ) %>%
    dplyr::mutate(
      dplyr::across(
        dplyr::all_of(loading_cols),
        ~ round(.x, digits)
      ),
      Model = model,
      .before = 1
    )

  list(
    model_name = model,
    net_loads = net_loads_obj,
    table = loading_tbl
  )
}

plot_ega_loadings <- function(ega_loadings_table,
                              model_title    = NULL,
                              primary_cutoff = 0.30,
                              text_size      = 3.0,
                              base_size      = 11) {

  non_loading_cols <- c("Model", "Item number", "Item name",
```

```
                      "Low loadings", "High cross-loadings")
loading_cols <- setdiff(colnames(ega_loadings_table), non_loading_cols)

title_str <- if (!is.null(model_title)) {
  model_title
} else if ("Model" %in% colnames(ega_loadings_table)) {
  paste0("Community Loadings - ", unique(ega_loadings_table$Model)[1])
} else {
  "Community Loadings"
}

plot_data <- ega_loadings_table %>%
  dplyr::select(`Item name`, dplyr::all_of(loading_cols)) %>%
  tidyr::pivot_longer(
    cols      = dplyr::all_of(loading_cols),
    names_to  = "Community",
    values_to = "Loading"
  ) %>%
  dplyr::mutate(
    `Item name` = factor(`Item name`, levels = rev(ega_loadings_table$`Item
     ↪  name`)),
    is_primary  = abs(Loading) >= primary_cutoff
  )

ggplot(plot_data, aes(x = Community, y = `Item name`, fill = Loading)) +
  geom_tile(color = "white", linewidth = 0.3) +
  geom_tile(
    data  = dplyr::filter(plot_data, is_primary),
    color = "black", linewidth = 0.4, fill = NA
  ) +
  geom_text(aes(label = sprintf("%.2f", Loading)), size = text_size) +
  scale_fill_gradient2(
    low      = "blue",
    mid      = "white",
    high     = "red",
    midpoint = 0,
    limits   = c(-1, 1),
    name     = "Loading"
  ) +
  labs(
    x     = "Community",
    y     = NULL,
    title = title_str
  ) +
  theme_minimal(base_size = base_size) +
  theme(
    panel.grid  = element_blank(),
    axis.text.y = element_text(size = base_size - 2),
```

```
      plot.title  = element_text(hjust = 0.5, face = "bold", size = base_size)
    )
}

build_ega_candidate_items_table <- function(item_stability_table,
                                            ega_redundancy_table,
                                            ega_loadings_table = NULL,
                                            ega_item_diagnostics_table = NULL,
                                            item_ids = emfra_item_ids,
                                            item_labels = emfra_item_labels,
                                            item_labels_short =
                                             ↪  emfra_item_labels_short,
                                            stability_cutoff = 0.75,
                                            redundancy_cutoff = 0.25,
                                            loading_cutoff = 0.30,
                                            primary_loading_cutoff = 0.50,
                                            digits = 3,
                                            accuracy = 0.1) {

  item_lookup <- dplyr::bind_rows(
    tibble::tibble(
      Item = item_ids,
      `Item number` = stringr::str_extract(item_labels, "^Item\\s+\\d+"),
      `Item name` = item_labels_short,
      `Item label` = item_labels
    ),
    tibble::tibble(
      Item = item_labels_short,
      `Item number` = stringr::str_extract(item_labels, "^Item\\s+\\d+"),
      `Item name` = item_labels_short,
      `Item label` = item_labels
    ),
    tibble::tibble(
      Item = item_labels,
      `Item number` = stringr::str_extract(item_labels, "^Item\\s+\\d+"),
      `Item name` = item_labels_short,
      `Item label` = item_labels
    )
  ) %>%
    dplyr::distinct(Item, .keep_all = TRUE)

  stability_candidates <- item_stability_table %>%
    dplyr::filter(
      `Below replicability threshold` == "Yes"
    ) %>%
    dplyr::mutate(
      `Low item stability reason` = paste0(
```

```r
      "Replicability = ",
      `Replicability (%)`,
      " (",
      `Assigned community`,
      ")"
    )
  ) %>%
  dplyr::select(
    Model,
    `Item number`,
    `Item name`,
    `Low item stability reason`
  )

if (is.null(ega_redundancy_table) || nrow(ega_redundancy_table) == 0) {

  redundancy_candidates <- tibble::tibble(
    Model = character(),
    `Item number` = character(),
    `Item name` = character(),
    `Redundancy reason` = character()
  )

} else {

  redundancy_candidates <- dplyr::bind_rows(
    ega_redundancy_table %>%
      dplyr::filter(`wTO value` >= redundancy_cutoff) %>%
      dplyr::transmute(
        Model,
        Item = `Item 1`,
        Partner = `Item 2`,
        `wTO value`,
        `wTO cut-off`,
        `Redundancy level`
      ),

    ega_redundancy_table %>%
      dplyr::filter(`wTO value` >= redundancy_cutoff) %>%
      dplyr::transmute(
        Model,
        Item = `Item 2`,
        Partner = `Item 1`,
        `wTO value`,
        `wTO cut-off`,
        `Redundancy level`
      )
  ) %>%
```

```r
    dplyr::left_join(
      item_lookup %>%
        dplyr::select(
          Item,
          `Item number`,
          `Item name`
        ),
      by = "Item"
    ) %>%
    dplyr::mutate(
      `Redundancy detail` = paste0(
        "Redundant with ",
        Partner,
        " (wTO = ",
        format(round(`wTO value`, digits), nsmall = digits),
        "; ",
        `wTO cut-off`,
        "; ",
        `Redundancy level`,
        ")"
      )
    ) %>%
    dplyr::group_by(
      Model,
      `Item number`,
      `Item name`
    ) %>%
    dplyr::summarise(
      `Redundancy reason` = paste(
        `Redundancy detail`,
        collapse = "; "
      ),
      .groups = "drop"
    )
}

if (is.null(ega_loadings_table) || nrow(ega_loadings_table) == 0) {

  loading_candidates <- tibble::tibble(
    Model = character(),
    `Item number` = character(),
    `Item name` = character(),
    `Low community loading reason` = character(),
    `High community cross-loading reason` = character()
  )

} else {
```

```r
  loading_cols <- setdiff(
    colnames(ega_loadings_table),
    c(
      "Model",
      "Item",
      "Item number",
      "Item name",
      "Low loadings",
      "High cross-loadings"
    )
  )

  loading_candidates <- ega_loadings_table %>%
    dplyr::filter(
      `Low loadings` == "Yes" |
        `High cross-loadings` == "Yes"
    )

  if (nrow(loading_candidates) == 0) {

    loading_candidates <- tibble::tibble(
      Model = character(),
      `Item number` = character(),
      `Item name` = character(),
      `Low community loading reason` = character(),
      `High community cross-loading reason` = character()
    )

  } else {

    loading_matrix <- as.matrix(
      loading_candidates[, loading_cols, drop = FALSE]
    )

    abs_loading_matrix <- abs(loading_matrix)

    primary_idx <- apply(
      abs_loading_matrix,
      1,
      function(x) order(x, decreasing = TRUE)[1]
    )

    secondary_idx <- apply(
      abs_loading_matrix,
      1,
      function(x) {
        ord <- order(x, decreasing = TRUE)
        if (length(ord) >= 2) ord[2] else NA_integer_
```

```r
    }
  )

  row_idx <- seq_len(nrow(loading_candidates))

  primary_community <- loading_cols[primary_idx]
  primary_loading <- loading_matrix[cbind(row_idx, primary_idx)]

  secondary_community <- ifelse(
    !is.na(secondary_idx),
    loading_cols[secondary_idx],
    NA_character_
  )

  secondary_loading <- ifelse(
    !is.na(secondary_idx),
    loading_matrix[cbind(row_idx, secondary_idx)],
    NA_real_
  )

  primary_community_label <- ifelse(
    stringr::str_detect(primary_community, "^Dim\\s+"),
    primary_community,
    paste0("Dim ", primary_community)
  )

  secondary_community_label <- ifelse(
    !is.na(secondary_community) &
      stringr::str_detect(secondary_community, "^Dim\\s+"),
    secondary_community,
    ifelse(
      !is.na(secondary_community),
      paste0("Dim ", secondary_community),
      NA_character_
    )
  )

  loading_candidates <- loading_candidates %>%
    dplyr::mutate(
      .primary_community = primary_community_label,
      .primary_loading = primary_loading,
      .secondary_community = secondary_community_label,
      .secondary_loading = secondary_loading,

      `Low community loading reason` = ifelse(
        `Low loadings` == "Yes",
        paste0(
          "Highest abs. community loading = ",
```

```
            format(round(abs(.primary_loading), digits), nsmall = digits),
            " (",
            .primary_community,
            ")"
          ),
          "No"
        ),

        `High community cross-loading reason` = ifelse(
          `High cross-loadings` == "Yes",
          paste0(
            "Primary abs. community loading = ",
            format(round(abs(.primary_loading), digits), nsmall = digits),
            " (",
            .primary_community,
            "); secondary abs. community loading = ",
            format(round(abs(.secondary_loading), digits), nsmall = digits),
            " (",
            .secondary_community,
            ")"
          ),
          "No"
        )
      ) %>%
      dplyr::select(
        Model,
        `Item number`,
        `Item name`,
        `Low community loading reason`,
        `High community cross-loading reason`
      )
  }
}

if (is.null(ega_item_diagnostics_table) || nrow(ega_item_diagnostics_table) ==
 ↪  0) {

  diagnostics_candidates <- tibble::tibble(
    Model                        = character(),
    `Item number`                = character(),
    `Item name`                  = character(),
    `Item diagnostics reason` = character()
  )

} else {

  diagnostics_candidates <- ega_item_diagnostics_table %>%
    dplyr::rename(`Item diagnostics reason` = Diagnostic) %>%
```

```
    dplyr::select(
      Model,
      `Item number`,
      `Item name`,
      `Item diagnostics reason`
    )

}

candidate_tbl <- stability_candidates %>%
  dplyr::full_join(
    redundancy_candidates,
    by = c(
      "Model",
      "Item number",
      "Item name"
    )
  ) %>%
  dplyr::full_join(
    loading_candidates,
    by = c(
      "Model",
      "Item number",
      "Item name"
    )
  ) %>%
  dplyr::full_join(
    diagnostics_candidates,
    by = c(
      "Model",
      "Item number",
      "Item name"
    )
  ) %>%
  dplyr::mutate(
    `Low item stability reason` = dplyr::if_else(
      is.na(`Low item stability reason`),
      "No",
      `Low item stability reason`
    ),

    `Redundancy reason` = dplyr::if_else(
      is.na(`Redundancy reason`),
      "No",
      `Redundancy reason`
    ),

    `Low community loading reason` = dplyr::if_else(
```

```r
      is.na(`Low community loading reason`),
      "No",
      `Low community loading reason`
    ),

    `High community cross-loading reason` = dplyr::if_else(
      is.na(`High community cross-loading reason`),
      "No",
      `High community cross-loading reason`
    ),

    `Item diagnostics reason` = dplyr::if_else(
      is.na(`Item diagnostics reason`),
      "No",
      `Item diagnostics reason`
    ),

    `EGA candidate criterion` = purrr::pmap_chr(
      list(
        `Low item stability reason`,
        `Redundancy reason`,
        `Low community loading reason`,
        `High community cross-loading reason`,
        `Item diagnostics reason`
      ),
      function(stability, redundancy, low_loading, cross_loading, item_diag) {

        criteria <- c(
          if (stability != "No") "Low replicability" else NULL,
          if (redundancy != "No") "Redundancy" else NULL,
          if (low_loading != "No") "Low community loading" else NULL,
          if (cross_loading != "No") "High community cross-loading" else NULL,
          if (item_diag != "No") paste0("Item diagnostic (", item_diag, ")")
           ↪  else NULL
        )

        if (length(criteria) == 0) {
          "No EGA issue"
        } else {
          paste(criteria, collapse = " + ")
        }
      }
    ),

    .item_order = as.numeric(
      stringr::str_extract(`Item number`, "\\d+")
    )
  ) %>%
```

```r
    dplyr::filter(
      `EGA candidate criterion` != "No EGA issue"
    ) %>%
    dplyr::arrange(
      Model,
      .item_order
    ) %>%
    dplyr::select(
      Model,
      `Item number`,
      `Item name`,
      `EGA candidate criterion`,
      `Low item stability reason`,
      `Redundancy reason`,
      `Item diagnostics reason`,
      `Low community loading reason`,
      `High community cross-loading reason`
    )

  if (nrow(candidate_tbl) == 0) {
    return(
      tibble::tibble(
        Model = character(),
        `Item number` = character(),
        `Item name` = character(),
        `EGA candidate criterion` = character(),
        `Low item stability reason` = character(),
        `Redundancy reason` = character(),
        `Item diagnostics reason` = character(),
        `Low community loading reason` = character(),
        `High community cross-loading reason` = character()
      )
    )
  }

  candidate_tbl
}
```

##### 4.5.6 Item retention and elimiation criteria functions

```r
build_item_reduction_summary_table <- function(model_name,
                                               fa_candidate_items_table,
                                               ega_candidate_items_table,
                                               all_item_labels,
                                               all_item_labels_short) {
```

```r
clean_fa_low_loading <- function(x) {
  if (is.na(x) || x == "No") return(x)

  x <- stringr::str_replace(
    x,
    "^Highest abs\\. loading = ",
    ""
  )

  x
}

clean_fa_cross_loading <- function(x) {
  if (is.na(x) || x == "No") return(x)

  x <- stringr::str_replace(
    x,
    "^Primary abs\\. loading = ",
    ""
  )

  x <- stringr::str_replace(
    x,
    "; secondary abs\\. loading = ",
    ", "
  )

  x
}

clean_fa_communality <- function(x) {
  if (is.na(x) || x == "No") return(x)

  x <- stringr::str_replace(
    x,
    "^Communality = ",
    "h2 = "
  )

  x
}

clean_ega_stability <- function(x) {
  if (is.na(x) || x == "No") return(x)

  x <- stringr::str_replace(
    x,
    "^Replicability = ",
```

```
    ""
  )

  x
}

clean_ega_low_loading <- function(x) {
  if (is.na(x) || x == "No") return(x)

  x <- stringr::str_replace(
    x,
    "^Highest abs\\. community loading = ",
    ""
  )

  x
}

clean_ega_cross_loading <- function(x) {
  if (is.na(x) || x == "No") return(x)

  x <- stringr::str_replace(
    x,
    "^Primary abs\\. community loading = ",
    "Primary = "
  )

  x <- stringr::str_replace(
    x,
    "; secondary abs\\. community loading = ",
    ", secondary = "
  )

  x
}

clean_ega_redundancy <- function(x) {
  if (is.na(x) || x == "No") return(x)

  x <- stringr::str_replace_all(
    x,
    "Redundant with ",
    "Redundancy with "
  )

  x <- stringr::str_replace_all(
    x,
    "\\(wTO\\s*=\\s*([0-9.]+);[^\\)]*\\)",
```

```r
    "(wTO = \\1)"
  )

  x
}

empty_fa <- tibble::tibble(
  Model = character(),
  `Item number` = character(),
  `Item name` = character(),
  `Low factor loadings reason` = character(),
  `High cross-loadings reason` = character(),
  `Low communality reason` = character()
)

empty_ega <- tibble::tibble(
  Model = character(),
  `Item number` = character(),
  `Item name` = character(),
  `EGA candidate criterion` = character(),
  `Low item stability reason` = character(),
  `Redundancy reason` = character(),
  `Item diagnostics reason` = character(),
  `Low community loading reason` = character(),
  `High community cross-loading reason` = character()
)

fa_tbl <- if (is.null(fa_candidate_items_table) ||
              nrow(fa_candidate_items_table) == 0) {
  empty_fa
} else {
  fa_candidate_items_table
}

ega_tbl <- if (is.null(ega_candidate_items_table) ||
               nrow(ega_candidate_items_table) == 0) {
  empty_ega
} else {
  ega_candidate_items_table
}

fa_summary <- fa_tbl %>%
  dplyr::filter(Model == model_name) %>%
  dplyr::mutate(
    `FA diagnostic summary` = purrr::pmap_chr(
      list(
        `Low factor loadings reason`,
        `High cross-loadings reason`,
```

```
        `Low communality reason`
      ),
      function(low_loading, cross_loading, communality) {

        low_loading_clean <- clean_fa_low_loading(low_loading)
        cross_loading_clean <- clean_fa_cross_loading(cross_loading)
        communality_clean <- clean_fa_communality(communality)

        reasons <- c(
          if (!is.na(low_loading_clean) && low_loading_clean != "No") {
            paste0("Low factor loading: ", low_loading_clean)
          } else {
            NULL
          },

          if (!is.na(cross_loading_clean) && cross_loading_clean != "No") {
            paste0("High cross-loading: ", cross_loading_clean)
          } else {
            NULL
          },

          if (!is.na(communality_clean) && communality_clean != "No") {
            paste0("Low communality: ", communality_clean)
          } else {
            NULL
          }
        )

        if (length(reasons) == 0) {
          "No FA diagnostic issue detected."
        } else {
          paste(reasons, collapse = "; ")
        }
      }
    )
  ) %>%
  dplyr::select(
    Model,
    `Item number`,
    `Item name`,
    `FA diagnostic summary`
  )

ega_summary <- ega_tbl %>%
  dplyr::filter(Model == model_name) %>%
  dplyr::mutate(
    `Item diagnostics reason` = if (
      "Item diagnostics reason" %in% colnames(ega_tbl)
```

```r
    ) {
      `Item diagnostics reason`
    } else {
      NA_character_
    },
    `EGA diagnostic summary` = purrr::pmap_chr(
      list(
        `Low item stability reason`,
        `Redundancy reason`,
        `Item diagnostics reason`,
        `Low community loading reason`,
        `High community cross-loading reason`
      ),
      function(stability, redundancy, item_diag, low_loading, cross_loading) {

        stability_clean    <- clean_ega_stability(stability)
        redundancy_clean   <- clean_ega_redundancy(redundancy)
        low_loading_clean  <- clean_ega_low_loading(low_loading)
        cross_loading_clean <- clean_ega_cross_loading(cross_loading)

        reasons <- c(
          if (!is.na(stability_clean) && stability_clean != "No") {
            paste0("Low item stability: ", stability_clean)
          } else {
            NULL
          },

          if (!is.na(redundancy_clean) && redundancy_clean != "No") {
            redundancy_clean
          } else {
            NULL
          },

          if (!is.na(item_diag) && item_diag != "No") {
            paste0("Item diagnostic: ", item_diag)
          } else {
            NULL
          },

          if (!is.na(low_loading_clean) && low_loading_clean != "No") {
            paste0("Low community loading: ", low_loading_clean)
          } else {
            NULL
          },

          if (!is.na(cross_loading_clean) && cross_loading_clean != "No") {
            paste0("High community cross-loading: ", cross_loading_clean)
          } else {
```

```
              NULL
            }
          )

          if (length(reasons) == 0) {
            "No EGA diagnostic issue detected."
          } else {
            paste(reasons, collapse = "; ")
          }
        }
      )
    ) %>%
    dplyr::select(
      Model,
      `Item number`,
      `Item name`,
      `EGA diagnostic summary`
    )

  all_items <- tibble::tibble(
    Model          = model_name,
    `Item number` = stringr::str_extract(all_item_labels, "^Item\\s+\\d+"),
    `Item name`    = all_item_labels_short
  )

  all_items %>%
    dplyr::left_join(
      fa_summary %>% dplyr::select(`Item number`, `Item name`, `FA diagnostic
↪   summary`),
      by = c("Item number", "Item name")
    ) %>%
    dplyr::left_join(
      ega_summary %>% dplyr::select(`Item number`, `Item name`, `EGA diagnostic
↪   summary`),
      by = c("Item number", "Item name")
    ) %>%
    dplyr::mutate(
      `FA diagnostic summary` = dplyr::if_else(
        is.na(`FA diagnostic summary`),
        "No FA diagnostic issue detected.",
        `FA diagnostic summary`
      ),

      `EGA diagnostic summary` = dplyr::if_else(
        is.na(`EGA diagnostic summary`),
        "No EGA diagnostic issue detected.",
        `EGA diagnostic summary`
      ),
```

```
      .item_order = as.numeric(
        stringr::str_extract(`Item number`, "\\d+")
      )
    ) %>%
    dplyr::arrange(
      Model,
      .item_order
    ) %>%
    dplyr::select(
      Model,
      `Item number`,
      `Item name`,
      `FA diagnostic summary`,
      `EGA diagnostic summary`
    )
}
```

#### 4.6 Full Model

##### 4.6.1 Matrix diagnosis and selection

```
emfra_items_full <- data_emfra %>%
  dplyr::select(all_of(emfra_item_ids))

full_selected_matrix <- select_polychoric_matrices(emfra_items_full)

matrix_diagnostics_full <- build_matrix_diagnostics_table(
  selected_obj = full_selected_matrix,
  model_name = "Full Model"
)
```

Table 9: Selected matrix for the Full Model

| Model | Package | Correction | Selected for dim. analysis | Symmetric | PSD | Total eigenvalues (n) | Negative eigenvalues (n) | Minimum eigenvalue | Negative eigenvalues |
|---|---|---|---|---|---|---|---|---|---|
| Full Model | psych | 0 | No | TRUE | FALSE | 21 | 1 | -0.035 | -0.03508537 |
| Full Model | EGAnet | 0 | No | TRUE | FALSE | 21 | 1 | -0.014 | -0.0139064 |
| Full Model | psych | 0.5 | Yes | TRUE | TRUE | 21 | 0 | 0.018 | NA |
| Full Model | EGAnet | 0.5 | Yes | TRUE | TRUE | 21 | 0 | 0.025 | NA |

##### 4.6.2 Dimensional diagnosis - Bootstrap PA and FA

```r
if (!exists("boot_pa_full")) {

  boot_pa_full <- run_parallel_bootstrap(
    selected_obj = full_selected_matrix,
    data         = emfra_items_full,
    model_name   = "Full Model"
  )

  dir.create("cache", showWarnings = FALSE)
  saveRDS(boot_pa_full, ruta_cache_pa_full)
}
```

Table 10: Bootstrap Parallel Analysis — Full Model

| Factor | Eigenvalue | Null_mean | Null_P95 | Boot_mean | Boot_P5 | Boot_P95 | Retained - Horn's PA (P95) | Retained - Bootstrap PA (90% CI, one-sided) | Non-overlap with permuted eigenvalues (%) |
|---|---|---|---|---|---|---|---|---|---|
| 1 | 5.217 | 1.606 | 2.054 | 5.399 | 4.671 | 6.144 | Yes | Yes | 100.0 |
| 2 | 1.933 | 0.870 | 1.020 | 2.054 | 1.672 | 2.500 | Yes | Yes | 100.0 |
| 3 | 0.768 | 0.709 | 0.821 | 1.067 | 0.858 | 1.311 | No | Yes | 98.1 |
| 4 | 0.641 | 0.581 | 0.674 | 0.791 | 0.629 | 0.967 | No | No | 88.2 |
| 5 | 0.512 | 0.478 | 0.565 | 0.566 | 0.427 | 0.713 | No | No | 50.4 |
| 6 | 0.358 | 0.380 | 0.457 | 0.392 | 0.275 | 0.513 | No | No | 16.8 |
| 7 | 0.171 | 0.296 | 0.368 | 0.242 | 0.148 | 0.351 | No | No | 3.0 |
| 8 | 0.084 | 0.216 | 0.284 | 0.123 | 0.041 | 0.211 | No | No | 0.2 |
| 9 | 0.059 | 0.137 | 0.204 | 0.021 | -0.055 | 0.102 | No | No | 0.0 |
| 10 | -0.094 | 0.065 | 0.124 | -0.067 | -0.138 | 0.007 | No | No | 0.0 |
| 11 | -0.152 | -0.006 | 0.054 | -0.145 | -0.211 | -0.075 | No | No | 0.0 |
| 12 | -0.193 | -0.074 | -0.017 | -0.217 | -0.279 | -0.158 | No | No | 0.0 |
| 13 | -0.231 | -0.139 | -0.079 | -0.282 | -0.339 | -0.225 | No | No | 0.0 |
| 14 | -0.263 | -0.205 | -0.144 | -0.342 | -0.401 | -0.282 | No | No | 0.0 |
| 15 | -0.325 | -0.270 | -0.212 | -0.402 | -0.460 | -0.343 | No | No | 0.0 |
| 16 | -0.396 | -0.332 | -0.274 | -0.462 | -0.524 | -0.403 | No | No | 0.0 |
| 17 | -0.415 | -0.398 | -0.341 | -0.526 | -0.594 | -0.464 | No | No | 0.0 |
| 18 | -0.451 | -0.465 | -0.402 | -0.590 | -0.653 | -0.524 | No | No | 0.0 |
| 19 | -0.551 | -0.534 | -0.469 | -0.662 | -0.727 | -0.601 | No | No | 0.0 |
| 20 | -0.670 | -0.612 | -0.542 | -0.739 | -0.810 | -0.670 | No | No | 0.0 |
| 21 | -0.785 | -0.720 | -0.627 | -0.823 | -0.902 | -0.745 | No | No | 0.0 |

```r
plot_parallel_bootstrap_ggplot(boot_pa_full)
```

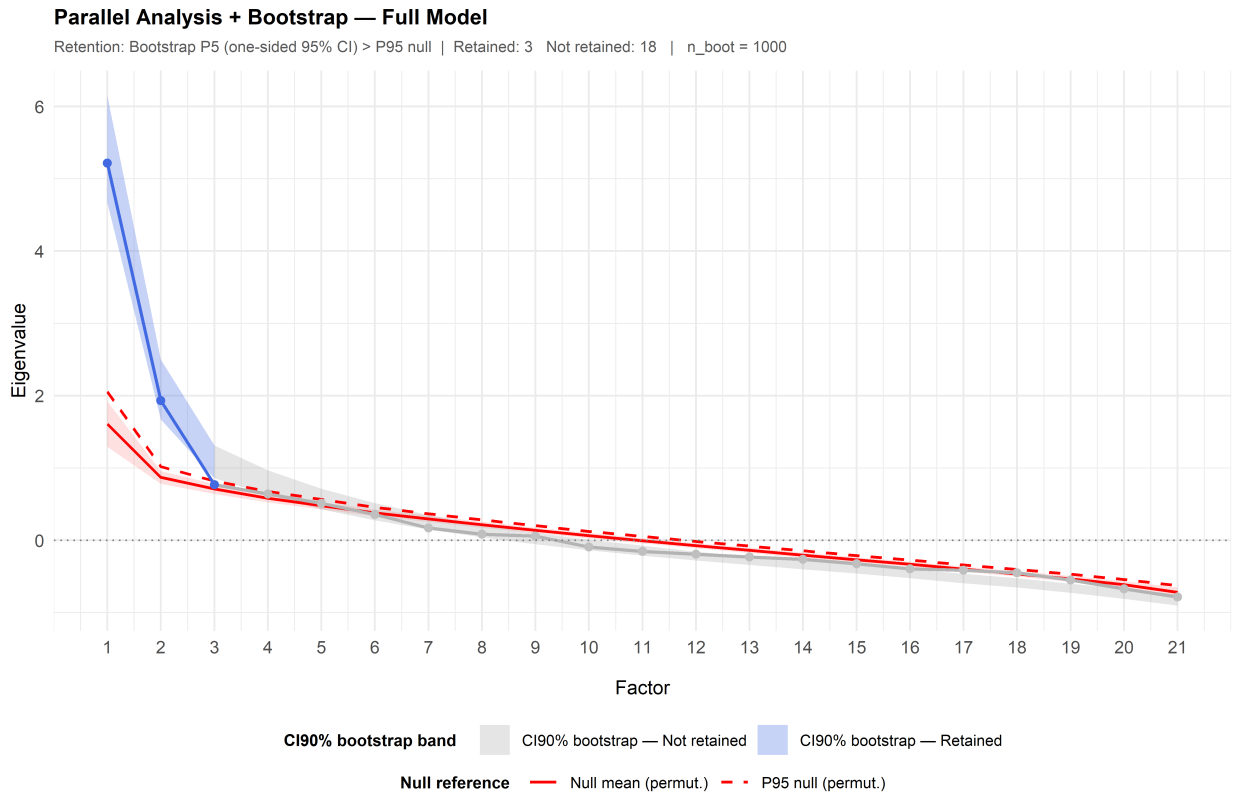

```r
fa_full <- run_fa_psych_selected(
  selected_obj = full_selected_matrix,
  data = emfra_items_full,
  model_name = "Full Model",
  pa_obj = boot_pa_full,
  fm = "minres",
  rotate = "oblimin",
  n.iter = 1000,
  SMC = TRUE,
  smooth = FALSE,
  warnings = FALSE,
  residuals = FALSE,
  scores = "tenBerge"
)

fa_fit_full <- extract_fa_fit_table(
  fa_obj = fa_full
)
```

Table 11: Exploratory Factor Analysis of Full Model

| Model | No. factors | Proportion Var | Chi-square | p-value | TLI | RMSEA | RMSR | Model fit | Error fit |
|---|---|---|---|---|---|---|---|---|---|
| Full Model | 3 | MR1: 0.199; MR2: 0.138; MR3: 0.075 | 992.864 | 0 | 0.446 | 0.167 | 0.074 | poor model fit | poor-to-marginal residual fit |

##### 4.6.3 Dimensional diagnosis - EGA

```
emfra_items_full_ega <- emfra_items_full

colnames(emfra_items_full_ega) <- emfra_item_labels_short[
  match(colnames(emfra_items_full), emfra_item_ids)
]

full_selected_matrix_ega <- select_polychoric_matrices(
  data = emfra_items_full_ega
)

bootega_full <- run_bootega_selected(
  data         = emfra_items_full_ega,
  selected_obj = full_selected_matrix_ega,
  model_name   = "Full Model",
  iter         = 1000,
  seed         = 123,
  ncores       = 1,
  plot.EGA     = TRUE,
  verbose      = TRUE
)
```

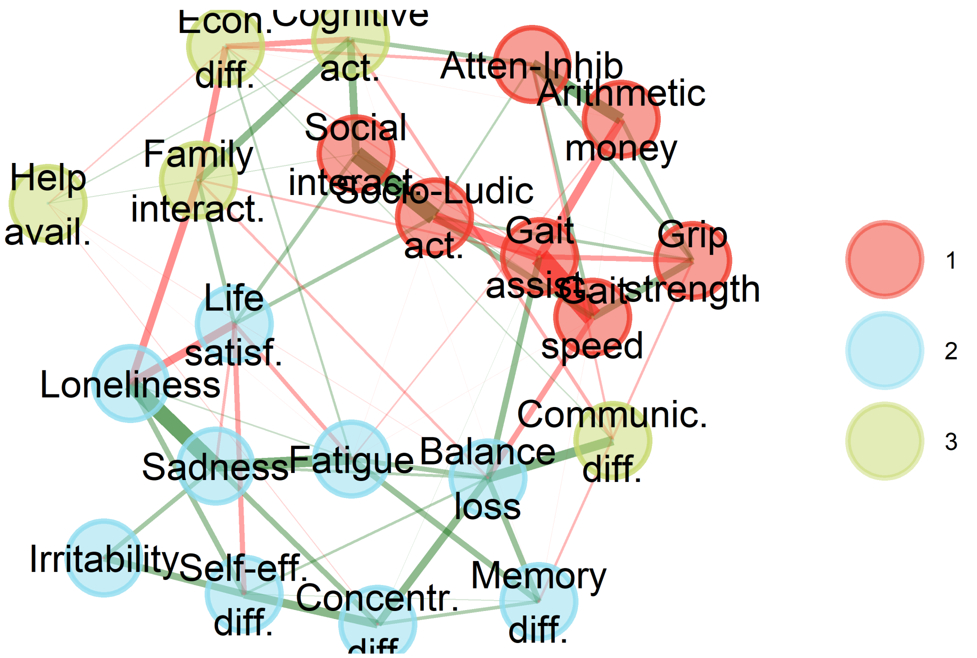

```r
ndim_freq_full <- build_ndim_frequency_table(
  boot_list = list(
    "Full Model" = bootega_full$boot
  ),
  n_factors_range = 1:7
)

plot_ndim_frequency(
  freq_tbl = ndim_freq_full,
  y_limit = 70,
  facet = FALSE
)
```

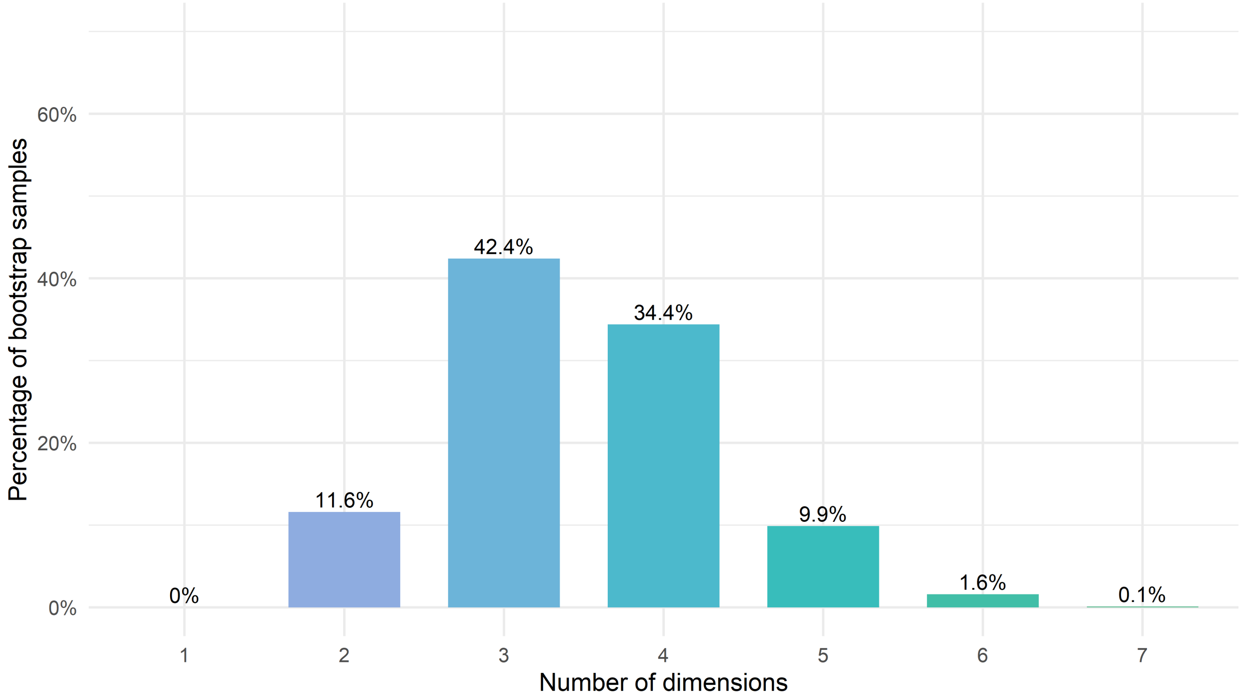

Figure 14: Frequency of the number of communities across bootstrap samples in the Full Model

```
ndim_summary_full <- summarise_ndim_frequency(
  freq_tbl = ndim_freq_full,
  model_name = "Full Model",
  top_n = 4,
  digits = 1
)
```

Table 12: Dimensional summary of EGA dimensions in the Full Model

| Model | Modal solution | Ranked dimensional solutions | Difference 1st-2nd | Difference 1st-3rd | Difference 1st-4th |
|---|---|---|---|---|---|
| Full Model | 3 dimensions (42.4%) | 3 dimensions: 42.4%; 4 dimensions: 34.4%; 2 dimensions: 11.6%; 5 dimensions: 9.9% | 8% | 30.8% | 32.5% |

```
structural_consistency_full <- extract_structural_consistency(
  boot = bootega_full$boot,
  model = "Full Model"
)
```

Table 13: Structural consistency of EGA dimensions in the Full Model

| Model | Dim 1 | Dim 2 | Dim 3 |
|---|---|---|---|
| Full Model | 30.8% | 19.7% | 15.1% |

##### 4.6.4 Dimensional diagnosis - Model acceptance criteria

```
dimensional_decision_full <- build_dimensional_decision_table(
  model_name = "Full Model",
  pa_obj = boot_pa_full,
  fa_fit_table = fa_fit_full,
  ndim_summary = ndim_summary_full,
  structural_consistency = structural_consistency_full,
  previous_model_comparison = NULL,
  researcher_judgement = c(
    "Convergence between Bootstrap PA and modal Bootstrap EGA",
    "Poor model performance",
    "Poor model performance",
    "First evaluated model. No previous comparison available.",
    "Do not retain. Proceed with item reduction for better model performance."
  )
)
```

Table 14: Dimensional acceptability criteria of the Full Model

| Model | Criterion | Objective evidence | Researcher judgement |
|---|---|---|---|
| Full Model | 1. Bootstrap PA-EGA dimensional convergence | Bootstrap PA suggested 3 factors; EGA modal solution was 3 dimensions (42.4%). | Convergence between Bootstrap PA and modal Bootstrap EGA |
| Full Model | 2. FA dimensional performance | Proportion Var. = MR1: 0.199; MR2: 0.138; MR3: 0.075; model fit = poor model fit; error fit = poor-to-marginal residual fit. | Poor model performance |
| Full Model | 3. EGA dimensional stability | Modal solution = 3 dimensions (42.4%); differences: 1st-2nd = 8%, 1st-3rd = 30.8%, 1st-4th = 32.5%; structural consistency = Dim 1: 30.8%; Dim 2: 19.7%; Dim 3: 15.1%. | Poor model performance |
| Full Model | 4. Improvement over previous model | Not applicable for the first evaluated model. | First evaluated model. No previous comparison available. |
| Full Model | 5. Model retention | Final model decision based on the previous dimensional criteria (1-4) | Do not retain. Proceed with item reduction for better model performance. |

##### 4.6.5 Item diagnosis - PA and FA

```r
fa_item_table_full <- build_fa_item_table(
  fa_obj = fa_full,
  model_name = "Full Model",
  item_ids = emfra_item_ids,
  item_labels = emfra_item_labels,
  item_labels_short = emfra_item_labels_short,
  loading_cutoff = 0.30,
  primary_loading_cutoff = 0.50,
  communality_cutoff = 0.30,
  digits = 3
)
```

Table 15: Factor analysis item diagnostics for the Full Model

| Model | Item number | Item name | Communality | MR1 | MR2 | MR3 | Low loadings | High cross-loadings | Low communalities |
|---|---|---|---|---|---|---|---|---|---|
| Full Model | Item 1 | Grip strength | 0.474 | 0.681 | -0.021 | 0.006 | No | No | No |
| Full Model | Item 2 | Gait speed | 0.776 | 0.881 | -0.007 | -0.013 | No | No | No |
| Full Model | Item 3 | Gait assist. | 0.877 | -0.928 | 0.070 | 0.126 | No | No | No |
| Full Model | Item 4 | Balance loss | 0.475 | -0.340 | 0.462 | -0.117 | No | No | No |
| Full Model | Item 5 | Fatigue | 0.358 | -0.160 | 0.519 | -0.040 | No | No | No |
| Full Model | Item 6 | Arithmetic money | 0.329 | 0.508 | -0.013 | 0.187 | No | No | No |
| Full Model | Item 7 | Atten-Inhib | 0.373 | 0.501 | 0.064 | 0.310 | No | No | No |
| Full Model | Item 8 | Memory diff. | 0.179 | -0.214 | 0.288 | -0.053 | Yes | No | Yes |
| Full Model | Item 9 | Concentr. diff. | 0.252 | -0.067 | 0.459 | -0.060 | No | No | Yes |
| Full Model | Item 10 | Cognitive act. | 0.465 | 0.275 | 0.124 | 0.614 | No | No | No |
| Full Model | Item 11 | Loneliness | 0.480 | 0.027 | 0.696 | -0.015 | No | No | No |
| Full Model | Item 12 | Sadness | 0.518 | -0.009 | 0.731 | 0.066 | No | No | No |
| Full Model | Item 13 | Irritability | 0.205 | 0.198 | 0.442 | -0.088 | No | No | Yes |
| Full Model | Item 14 | Self-eff. diff. | 0.337 | 0.075 | 0.600 | 0.007 | No | No | No |
| Full Model | Item 15 | Life satisf. | 0.331 | 0.174 | -0.472 | 0.081 | No | No | No |
| Full Model | Item 16 | Econ. diff. | 0.582 | 0.220 | 0.125 | -0.735 | No | No | No |
| Full Model | Item 17 | Family interact. | 0.362 | 0.163 | -0.294 | 0.381 | No | No | No |
| Full Model | Item 18 | Social interact. | 0.370 | 0.591 | 0.078 | 0.143 | No | No | No |
| Full Model | Item 19 | Socio-Ludic act. | 0.653 | 0.798 | -0.018 | 0.026 | No | No | No |

| Model | Item number | Item name | Communality | MR1 | MR2 | MR3 | Low loadings | High cross-loadings | Low communalities |
|---|---|---|---|---|---|---|---|---|---|
| Full Model | Item 20 | Communic. diff. | 0.168 | -0.086 | 0.174 | -0.296 | Yes | No | Yes |
| Full Model | Item 21 | Help avail. | 0.097 | 0.020 | -0.051 | 0.290 | Yes | No | Yes |

```r
loading_cols_fa_full <- setdiff(
  colnames(fa_item_table_full),
  c("Model", "Item number", "Item name", "Communality",
    "Low loadings", "High cross-loadings", "Low communalities")
)

plot_loadings_fa_full <- fa_item_table_full %>%
  dplyr::select(`Item name`, dplyr::all_of(loading_cols_fa_full)) %>%
  tidyr::pivot_longer(
    cols = dplyr::all_of(loading_cols_fa_full),
    names_to = "Factor",
    values_to = "Loading"
  ) %>%
  dplyr::mutate(
    `Item name` = factor(`Item name`, levels = rev(fa_item_table_full$`Item
     ↪  name`)),
    is_primary = abs(Loading) >= 0.50
  )

ggplot(plot_loadings_fa_full, aes(x = Factor, y = `Item name`, fill = Loading)) +
  geom_tile(color = "white", linewidth = 0.3) +
  geom_tile(
    data = dplyr::filter(plot_loadings_fa_full, is_primary),
    color = "black", linewidth = 0.4, fill = NA
  ) +
  geom_text(aes(label = sprintf("%.2f", Loading)), size = 2.5) +
  scale_fill_gradient2(
    low      = "blue",
    mid      = "white",
    high     = "red",
    midpoint = 0,
    limits   = c(-1, 1),
    name     = "Factor loadings"
  ) +
  labs(
    x     = "Factor",
    y     = NULL,
    title = "Factor Loadings - Full Model"
  ) +
  theme_minimal(base_size = 10) +
  theme(
```

```
  panel.grid   = element_blank(),
  axis.text.y  = element_text(size = 8),
  plot.title   = element_text(hjust = 0.5, face = "bold", size = 10),
  legend.title = element_text(size = 9),
  legend.text  = element_text(size = 8),
)
```

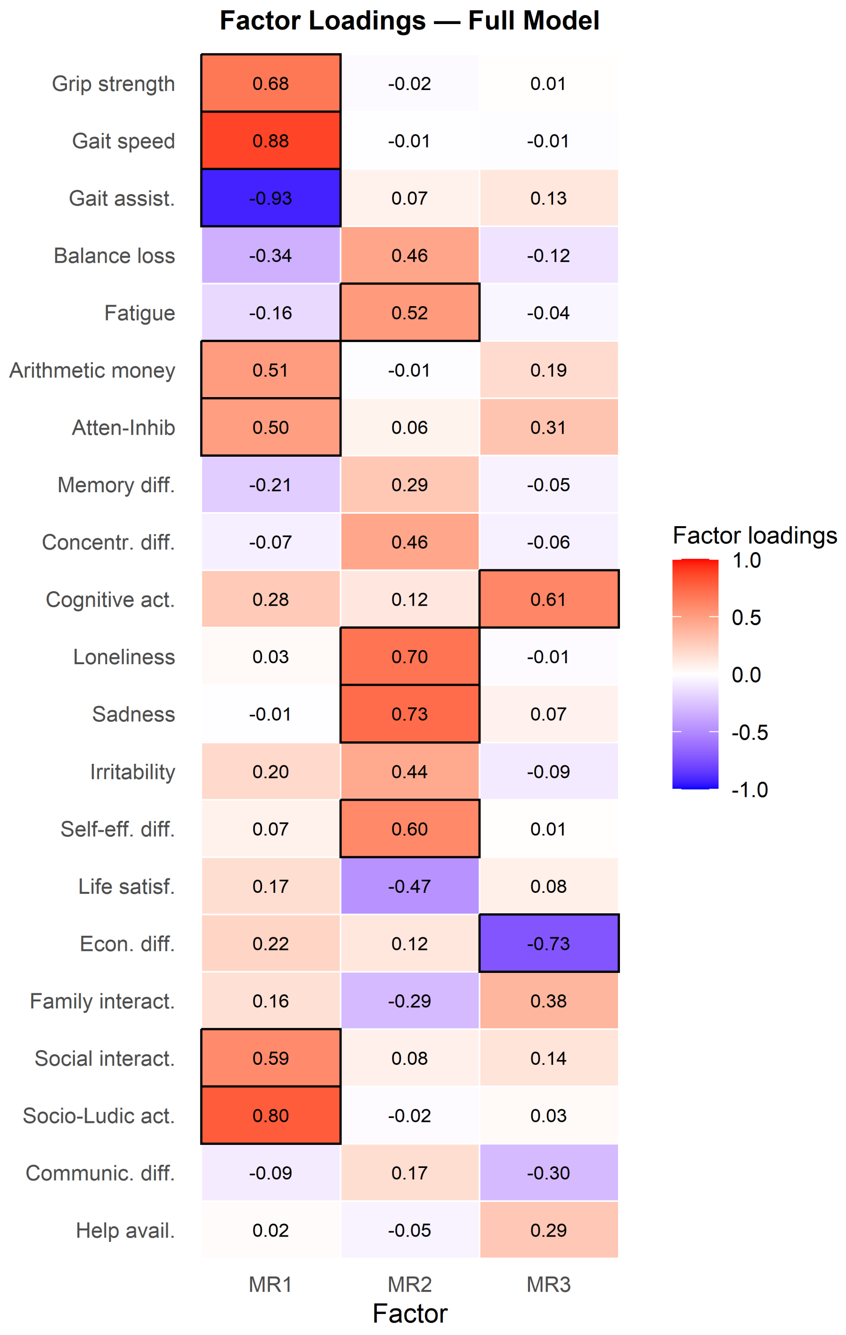

Figure 15: Factor loading heatmap for Reduced Model C. Black borders mark primary loadings ($|\lambda| \geq 0.50$).

```r
fa_candidate_items_full <- build_fa_candidate_items_table(
  fa_item_table = fa_item_table_full,
  digits = 3
)
```

Table 16: Candidate items for elimination in FA from Full Model

| Model | Item number | Item name | Low factor loadings reason | High cross-loadings reason | Low communality reason |
|---|---|---|---|---|---|
| Full Model | Item 8 | Memory diff. | Highest abs. loading = MR2: 0.288 | No | Communality = 0.179 |
| Full Model | Item 9 | Concentr. diff. | No | No | Communality = 0.252 |
| Full Model | Item 13 | Irritability | No | No | Communality = 0.205 |
| Full Model | Item 20 | Communic. diff. | Highest abs. loading = MR3: -0.296 | No | Communality = 0.168 |
| Full Model | Item 21 | Help avail. | Highest abs. loading = MR3: 0.290 | No | Communality = 0.097 |

##### 4.6.6 Item diagnosis - EGA

```r
item_stability_full <- extract_item_stability(
  boot = bootega_full$boot,
  model = "Full Model",
  item_ids = emfra_item_ids,
  item_labels = emfra_item_labels,
  item_labels_short = emfra_item_labels_short,
  stability_cutoff = 0.75
)

item_stability_table_full <- item_stability_full$table
item_stability_plot_full <- item_stability_full$plot
```

```r
item_stability_plot_full
```

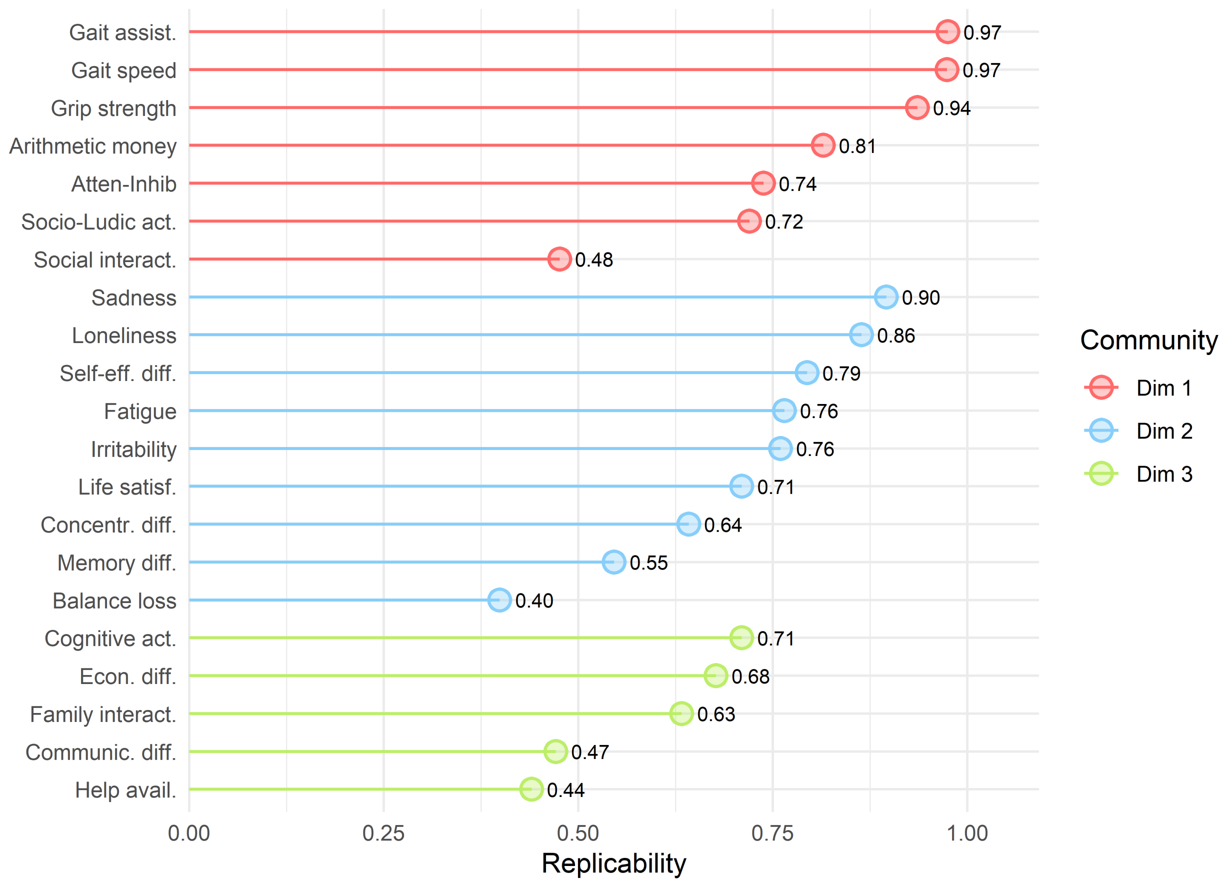

Figure 16: Item replicability from Full Model

```r
ega_redundancy_full <- build_ega_redundancy_table(
  data = emfra_items_full,
  selected_obj = full_selected_matrix,
  model_name = "Full Model",
  uva.method = "MBR",
  cut.off = 0.25,
  reduce = FALSE,
  report_cutoff = 0.20
)

ega_redundancy_table_full <- ega_redundancy_full$table
```

Table 17: Item redundancy from Full Model

| Model | Item 1 | Item 2 | wTO value | wTO cut-off | Redundancy level |
|---|---|---|---|---|---|
| Full Model | Item 2. Gait speed | Item 3. Gait assistance | 0.331 | 0.3 | Large-to-very large |

| Model | Item 1 | Item 2 | wTO value | wTO cut-off | Redundancy level |
|---|---|---|---|---|---|
| Full Model | Item 18. Social interaction | Item 19. Social-ludic activities | 0.295 | 0.25 | Moderate-to-large |
| Full Model | Item 11. Loneliness | Item 12. Sadness | 0.247 | 0.2 | Small-to-moderate |
| Full Model | Item 6. Arithmetics for money | Item 7. Attention-inhibition | 0.206 | 0.2 | Small-to-moderate |

```
ega_item_diagnostics_full <- run_item_diagnostics_selected(
  data         = emfra_items_full_ega,
  selected_obj = full_selected_matrix_ega,
  model_name   = "Full Model",
  iter         = 1000,
  ncores       = 1,
  seed         = 123
)

ega_item_diagnostics_table_full <- ega_item_diagnostics_full$table
```

Table 18: Item diagnostics (Minor dimension, Two-item dimension, Multidimensional) from Full Model

| Model | Item number | Item name | Diagnostic |
|---|---|---|---|

```
ega_loadings_full <- extract_ega_community_loadings(
  boot = bootega_full$boot,
  model = "Full Model",
  item_ids = emfra_item_ids,
  item_labels = emfra_item_labels,
  item_labels_short = emfra_item_labels_short,
  loading.method = "revised",
  scaling = 2,
  rotation = "oblimin",
  loading_cutoff = 0.30,
  primary_loading_cutoff = 0.50,
  digits = 3
)

ega_loadings_table_full <- ega_loadings_full$table
```

Table 19: Item loadings in EGA from Full Model

| Model | Item number | Item name | 1 | 2 | 3 | Low loadings | High cross-loadings |
|---|---|---|---|---|---|---|---|
| Full Model | Item 19 | Socio-Ludic act. | 0.550 | 0.006 | -0.015 | No | No |
| Full Model | Item 2 | Gait speed | 0.542 | -0.012 | -0.016 | No | No |
| Full Model | Item 1 | Grip strength | 0.359 | 0.002 | -0.010 | No | No |
| Full Model | Item 6 | Arithmetic money | 0.347 | 0.020 | 0.001 | No | No |
| Full Model | Item 7 | Atten-Inhib | 0.279 | 0.060 | 0.196 | Yes | No |
| Full Model | Item 18 | Social interact. | 0.233 | -0.002 | 0.158 | Yes | No |
| Full Model | Item 3 | Gait assist. | -0.627 | 0.011 | 0.001 | No | No |
| Full Model | Item 12 | Sadness | 0.001 | 0.599 | 0.051 | No | No |
| Full Model | Item 11 | Loneliness | 0.033 | 0.416 | -0.109 | No | No |
| Full Model | Item 14 | Self-eff. diff. | 0.001 | 0.419 | 0.035 | No | No |
| Full Model | Item 5 | Fatigue | -0.018 | 0.377 | -0.027 | No | No |
| Full Model | Item 9 | Concentr. diff. | -0.003 | 0.352 | 0.009 | No | No |
| Full Model | Item 4 | Balance loss | -0.086 | 0.302 | -0.217 | No | No |
| Full Model | Item 8 | Memory diff. | -0.038 | 0.227 | 0.017 | Yes | No |
| Full Model | Item 13 | Irritability | 0.000 | 0.163 | 0.014 | Yes | No |
| Full Model | Item 15 | Life satisf. | 0.081 | -0.264 | 0.059 | Yes | No |
| Full Model | Item 10 | Cognitive act. | 0.047 | 0.061 | 0.434 | No | No |
| Full Model | Item 17 | Family interact. | -0.037 | -0.140 | 0.353 | No | No |
| Full Model | Item 21 | Help avail. | -0.021 | -0.018 | 0.095 | Yes | No |
| Full Model | Item 20 | Communic. diff. | -0.006 | 0.106 | -0.108 | Yes | No |
| Full Model | Item 16 | Econ. diff. | 0.058 | -0.010 | -0.375 | No | No |

```r
loading_cols_ega_full <- setdiff(
  colnames(ega_loadings_table_full),
  c("Model", "Item number", "Item name", "Low loadings", "High cross-loadings")
)

plot_loadings_ega_full <- ega_loadings_table_full %>%
  dplyr::select(`Item name`, dplyr::all_of(loading_cols_ega_full)) %>%
  tidyr::pivot_longer(
    cols = dplyr::all_of(loading_cols_ega_full),
    names_to = "Community",
    values_to = "Loading"
  ) %>%
  dplyr::mutate(
    `Item name` = factor(`Item name`, levels = rev(ega_loadings_table_full$`Item
    ↪ name`)),
    is_primary = abs(Loading) >= 0.50
  )

ggplot(plot_loadings_ega_full, aes(x = Community, y = `Item name`, fill =
↪ Loading)) +
  geom_tile(color = "white", linewidth = 0.3) +
  geom_tile(
    data = dplyr::filter(plot_loadings_ega_full, is_primary),
    color = "black", linewidth = 0.4, fill = NA
  ) +
  geom_text(aes(label = sprintf("%.2f", Loading)), size = 2.5) +
  scale_fill_gradient2(
    low      = "blue",
    mid      = "white",
    high     = "red",
    midpoint = 0,
    limits   = c(-1, 1),
    name     = "Community loading"
  ) +
  labs(
    x     = "Community",
    y     = NULL,
    title = "Community Loadings - Full Model"
  ) +
  theme_minimal(base_size = 10) +
  theme(
    panel.grid   = element_blank(),
    axis.text.y  = element_text(size = 8.5),
    plot.title   = element_text(hjust = 0.5, face = "bold", size = 10),
    legend.title = element_text(size = 9),
    legend.text  = element_text(size = 8),
  )
```

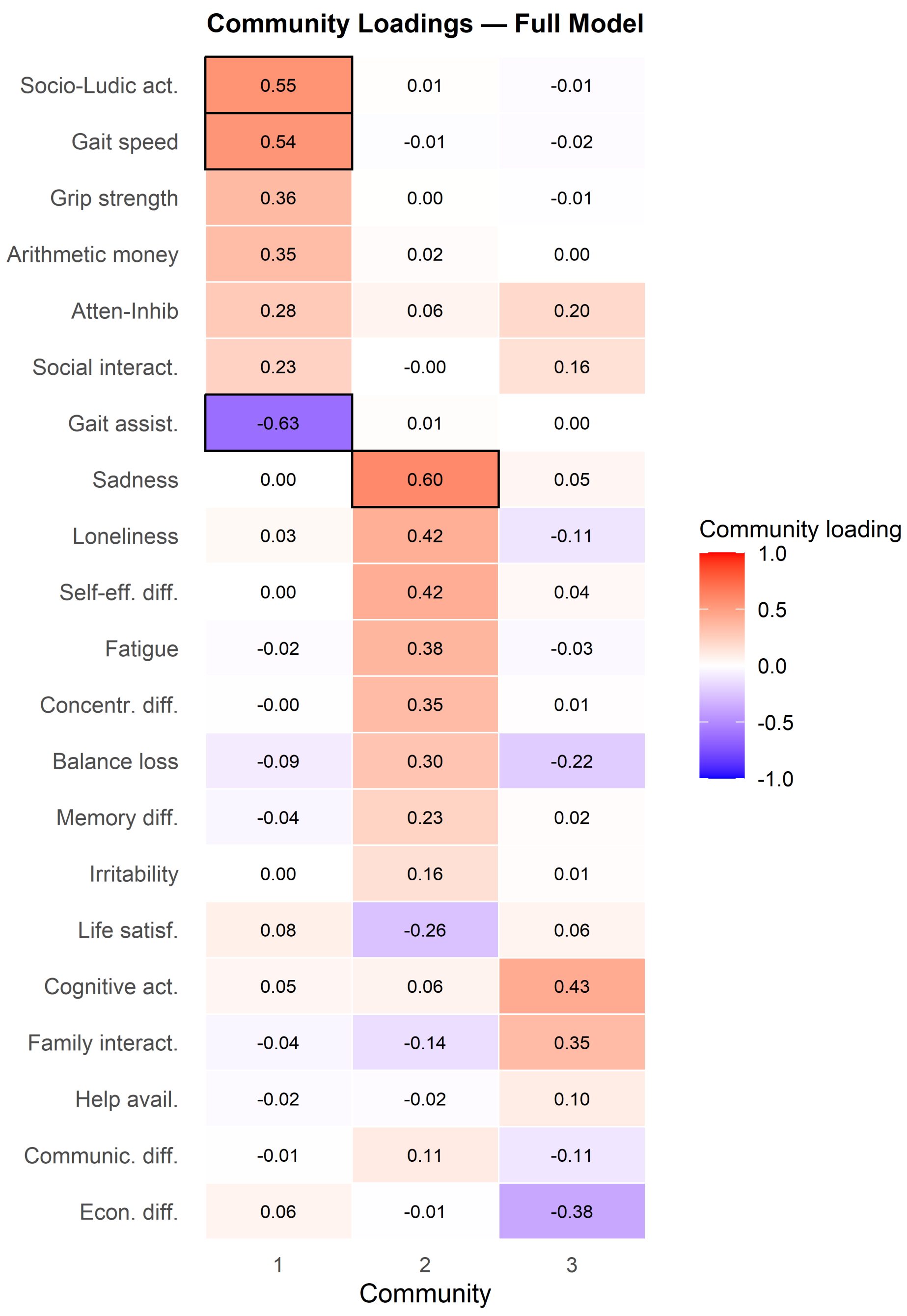

Figure 17: Community loading (CL) heatmap for Full Model. Black borders mark primary loadings ($CL \geq 0.50$).

```
ega_candidate_items_full <- build_ega_candidate_items_table(
  item_stability_table        = item_stability_table_full,
  ega_redundancy_table        = ega_redundancy_table_full,
  ega_loadings_table          = ega_loadings_table_full,
  ega_item_diagnostics_table = ega_item_diagnostics_table_full,
  stability_cutoff            = 0.75,
```

```
  redundancy_cutoff           = 0.25,
  loading_cutoff              = 0.30,
  primary_loading_cutoff      = 0.50,
  digits                      = 3
)
```

Table 20: Candidate items for elimination in EGA from Full Model

| Model | Item number | Item name | EGA candidate criterion | Low item stability reason | Redundancy reason | Item diagnostics reason | Low community loading reason | High community cross-loading reason |
|---|---|---|---|---|---|---|---|---|
| Full Model | Item 2 | Gait speed | Redundancy | No | Redundant with Item 3. Gait assistance (wTO = 0.331; 0.3; Large-to-very large) | No | No | No |
| Full Model | Item 3 | Gait assist. | Redundancy | No | Redundant with Item 2. Gait speed (wTO = 0.331; 0.3; Large-to-very large) | No | No | No |
| Full Model | Item 4 | Balance loss | Low replicability | Replicability = 39.9% (Dim 2) | No | No | No | No |
| Full Model | Item 7 | Atten-Inhib | Low replicability + Low community loading | Replicability = 73.8% (Dim 1) | No | No | Highest abs. community loading = 0.279 (Dim 1) | No |
| Full Model | Item 8 | Memory diff. | Low replicability + Low community loading | Replicability = 54.6% (Dim 2) | No | No | Highest abs. community loading = 0.227 (Dim 2) | No |
| Full Model | Item 9 | Concentr. diff. | Low replicability | Replicability = 64.2% (Dim 2) | No | No | No | No |
| Full Model | Item 10 | Cognitive act. | Low replicability | Replicability = 71.0% (Dim 3) | No | No | No | No |
| Full Model | Item 13 | Irritability | Low community loading | No | No | No | Highest abs. community loading = 0.163 (Dim 2) | No |
| Full Model | Item 15 | Life satisf. | Low replicability + Low community loading | Replicability = 71.0% (Dim 2) | No | No | Highest abs. community loading = 0.264 (Dim 2) | No |
| Full Model | Item 16 | Econ. diff. | Low replicability | Replicability = 67.7% (Dim 3) | No | No | No | No |
| Full Model | Item 17 | Family inter-act. | Low replicability | Replicability = 63.3% (Dim 3) | No | No | No | No |
| Full Model | Item 18 | Social inter-act. | Low replicability + Redundancy + Low community loading | Replicability = 47.6% (Dim 1) | Redundant with Item 19. Social-ludic activities (wTO = 0.295; 0.25; Moderate-to-large) | No | Highest abs. community loading = 0.233 (Dim 1) | No |

| Model | Item number | Item name | EGA candidate criterion | Low item stability reason | Redundancy reason | Item diagnostics reason | Low community loading reason | High community cross-loading reason |
|---|---|---|---|---|---|---|---|---|
| Full Model | Item 19 | Socio-Ludic act. | Low replicability + Redundancy | Replicability = 72.0% (Dim 1) | Redundant with Item 18. Social interaction (wTO = 0.295; 0.25; Moderate-to-large) | No | No | No |
| Full Model | Item 20 | Communic diff. | Low replicability + Low community loading | Replicability = 47.1% (Dim 3) | No | No | Highest abs. community loading = 0.108 (Dim 3) | No |
| Full Model | Item 21 | Help avail. | Low replicability + Low community loading | Replicability = 44.0% (Dim 3) | No | No | Highest abs. community loading = 0.095 (Dim 3) | No |

##### 4.6.7 Item diagnosis - Retention and elimination of items

```r
item_reduction_summary_full <- build_item_reduction_summary_table(
  model_name = "Full Model",
  fa_candidate_items_table = fa_candidate_items_full,
  ega_candidate_items_table = ega_candidate_items_full,
  all_item_labels = emfra_item_labels,
  all_item_labels_short = emfra_item_labels_short
)
```

Table 21: Item summary diagnostics from FA and EGA in Full Model

| Model | Item number | Item name | FA diagnostic summary | EGA diagnostic summary |
|---|---|---|---|---|
| Full Model | Item 1 | Grip strength | No FA diagnostic issue detected. | No EGA diagnostic issue detected. |
| Full Model | Item 2 | Gait speed | No FA diagnostic issue detected. | Redundancy with Item 3. Gait assistance (wTO = 0.331) |
| Full Model | Item 3 | Gait assist. | No FA diagnostic issue detected. | Redundancy with Item 2. Gait speed (wTO = 0.331) |
| Full Model | Item 4 | Balance loss | No FA diagnostic issue detected. | Low item stability: 39.9% (Dim 2) |
| Full Model | Item 5 | Fatigue | No FA diagnostic issue detected. | No EGA diagnostic issue detected. |
| Full Model | Item 6 | Arithmetic money | No FA diagnostic issue detected. | No EGA diagnostic issue detected. |
| Full Model | Item 7 | Atten-Inhib | No FA diagnostic issue detected. | Low item stability: 73.8% (Dim 1); Low community loading: 0.279 (Dim 1) |
| Full Model | Item 8 | Memory diff. | Low factor loading: MR2: 0.288; Low communality: h2 = 0.179 | Low item stability: 54.6% (Dim 2); Low community loading: 0.227 (Dim 2) |
| Full Model | Item 9 | Concentr. diff. | Low communality: h2 = 0.252 | Low item stability: 64.2% (Dim 2) |
| Full Model | Item 10 | Cognitive act. | No FA diagnostic issue detected. | Low item stability: 71.0% (Dim 3) |
| Full Model | Item 11 | Loneliness | No FA diagnostic issue detected. | No EGA diagnostic issue detected. |
| Full Model | Item 12 | Sadness | No FA diagnostic issue detected. | No EGA diagnostic issue detected. |
| Full Model | Item 13 | Irritability | Low communality: h2 = 0.205 | Low community loading: 0.163 (Dim 2) |
| Full Model | Item 14 | Self-eff. diff. | No FA diagnostic issue detected. | No EGA diagnostic issue detected. |
| Full Model | Item 15 | Life satisf. | No FA diagnostic issue detected. | Low item stability: 71.0% (Dim 2); Low community loading: 0.264 (Dim 2) |
| Full Model | Item 16 | Econ. diff. | No FA diagnostic issue detected. | Low item stability: 67.7% (Dim 3) |
| Full Model | Item 17 | Family interact. | No FA diagnostic issue detected. | Low item stability: 63.3% (Dim 3) |
| Full Model | Item 18 | Social interact. | No FA diagnostic issue detected. | Low item stability: 47.6% (Dim 1); Redundancy with Item 19. Social-ludic activities (wTO = 0.295); Low community loading: 0.233 (Dim 1) |
| Full Model | Item 19 | Socio-Ludic act. | No FA diagnostic issue detected. | Low item stability: 72.0% (Dim 1); Redundancy with Item 18. Social interaction (wTO = 0.295) |

| Model | Item number | Item name | FA diagnostic summary | EGA diagnostic summary |
|---|---|---|---|---|
| Full Model | Item 20 | Communic. diff. | Low factor loading: MR3: -0.296; Low communality: h2 = 0.168 | Low item stability: 47.1% (Dim 3); Low community loading: 0.108 (Dim 3) |
| Full Model | Item 21 | Help avail. | Low factor loading: MR3: 0.290; Low communality: h2 = 0.097 | Low item stability: 44.0% (Dim 3); Low community loading: 0.095 (Dim 3) |

Five items were detected as conflictive through both FA and EGA. The following items would be eliminated, including Item 8 (Memory difficulty), Item 9 (Concentration Difficulty), Item 13 (Irritability), Item 20 (Communication diffiulty), and Item 21 (Help availability). In addition, item 3 (Gait assistance) would be eliminated, due to its large redundancy with item 2, in addition to its high zero-order and partial correlations (above 0.7).

#### 4.7 Reduced Model A

##### 4.7.1 Matrix diagnosis and selection

```
excluded_item_ids_rma <- c(
  "EMFRA_item3_trait",   # Item 3. Gait assistance
  "EMFRA_item8_trait",   # Item 8. Memory difficulty
  "EMFRA_item9_trait",   # Item 9. Concentration difficulty.
  "EMFRA_item13_trait",  # Item 13. Irritability
  "EMFRA_item20_trait",  # Item 20. Communication difficulty
  "EMFRA_item21_trait"  # Item 21. Help availability
)

excluded_labels_rma <- emfra_item_labels_short[
  match(excluded_item_ids_rma, emfra_item_ids)
]

emfra_item_ids_reducedA <- emfra_item_ids[
  !emfra_item_ids %in% excluded_item_ids_rma
]

emfra_item_labels_reducedA <- emfra_item_labels[
  match(emfra_item_ids_reducedA, emfra_item_ids)
]

emfra_item_labels_short_reducedA <- emfra_item_labels_short[
  match(emfra_item_ids_reducedA, emfra_item_ids)
]

label_map_reducedA <- setNames(
  emfra_item_labels_reducedA,
  emfra_item_labels_short_reducedA
)

emfra_items_reducedA <- data_emfra %>%
  dplyr::select(all_of(emfra_item_ids_reducedA))

colnames(emfra_items_reducedA) <- emfra_item_labels_short_reducedA
```

```r
reduced_selected_matrix_A <- select_polychoric_matrices(emfra_items_reducedA)

matrix_diagnostics_reducedA <- build_matrix_diagnostics_table(
  selected_obj = reduced_selected_matrix_A,
  model_name   = "Reduced Model A"
)
```

Table 22: Selected matrix for the Reduced Model A

| Model | Package | Correction | Selected for dim. analysis | Symmetric | PSD | Total eigenvalues (n) | Negative eigenvalues (n) | Minimum eigenvalue | Negative eigenvalues |
|---|---|---|---|---|---|---|---|---|---|
| Reduced Model A | psych | 0 | Yes | TRUE | TRUE | 15 | 0 | 0.099 | NA |
| Reduced Model A | EGAnet | 0 | Yes | TRUE | TRUE | 15 | 0 | 0.115 | NA |

##### 4.7.2 Dimensional diagnosis - Bootstrap PA and FA

```r
if (!exists("boot_pa_reducedA")) {

  boot_pa_reducedA <- run_parallel_bootstrap(
    selected_obj = reduced_selected_matrix_A,
    data         = emfra_items_reducedA,
    model_name   = "Reduced Model A"
  )

  dir.create("cache", showWarnings = FALSE)
  saveRDS(boot_pa_reducedA, ruta_cache_pa_reducedA)
}
```

Table 23: Bootstrap Parallel Analysis — Reduced Model A

| Factor | Eigenvalue | Null_mean | Null_P95 | Boot_mean | Boot_P5 | Boot_P95 | Retained - Horn's PA (P95) | Retained - Bootstrap PA (90% CI, one-sided) | Non-overlap with permuted eigenvalues (%) |
|---|---|---|---|---|---|---|---|---|---|
| 1 | 4.148 | 1.336 | 1.921 | 4.263 | 3.649 | 4.859 | Yes | Yes | 100.0 |
| 2 | 1.498 | 0.636 | 0.778 | 1.617 | 1.280 | 1.998 | Yes | Yes | 100.0 |
| 3 | 0.607 | 0.488 | 0.586 | 0.808 | 0.618 | 1.038 | Yes | Yes | 97.3 |
| 4 | 0.564 | 0.375 | 0.457 | 0.551 | 0.387 | 0.725 | Yes | No | 82.0 |
| 5 | 0.198 | 0.274 | 0.358 | 0.280 | 0.159 | 0.409 | No | No | 14.0 |
| 6 | 0.026 | 0.182 | 0.252 | 0.118 | 0.024 | 0.222 | No | No | 1.7 |
| 7 | -0.035 | 0.097 | 0.162 | -0.001 | -0.091 | 0.088 | No | No | 0.2 |
| 8 | -0.067 | 0.017 | 0.078 | -0.109 | -0.189 | -0.030 | No | No | 0.0 |
| 9 | -0.213 | -0.060 | -0.003 | -0.203 | -0.277 | -0.126 | No | No | 0.0 |
| 10 | -0.243 | -0.135 | -0.072 | -0.287 | -0.357 | -0.214 | No | No | 0.0 |
| 11 | -0.289 | -0.211 | -0.148 | -0.367 | -0.432 | -0.299 | No | No | 0.0 |
| 12 | -0.393 | -0.288 | -0.228 | -0.448 | -0.524 | -0.376 | No | No | 0.0 |
| 13 | -0.443 | -0.371 | -0.299 | -0.536 | -0.620 | -0.464 | No | No | 0.0 |
| 14 | -0.576 | -0.457 | -0.383 | -0.643 | -0.740 | -0.559 | No | No | 0.0 |
| 15 | -0.635 | -0.564 | -0.473 | -0.780 | -0.886 | -0.667 | No | No | 0.0 |

```r
plot_parallel_bootstrap_ggplot(boot_pa_reducedA)
```

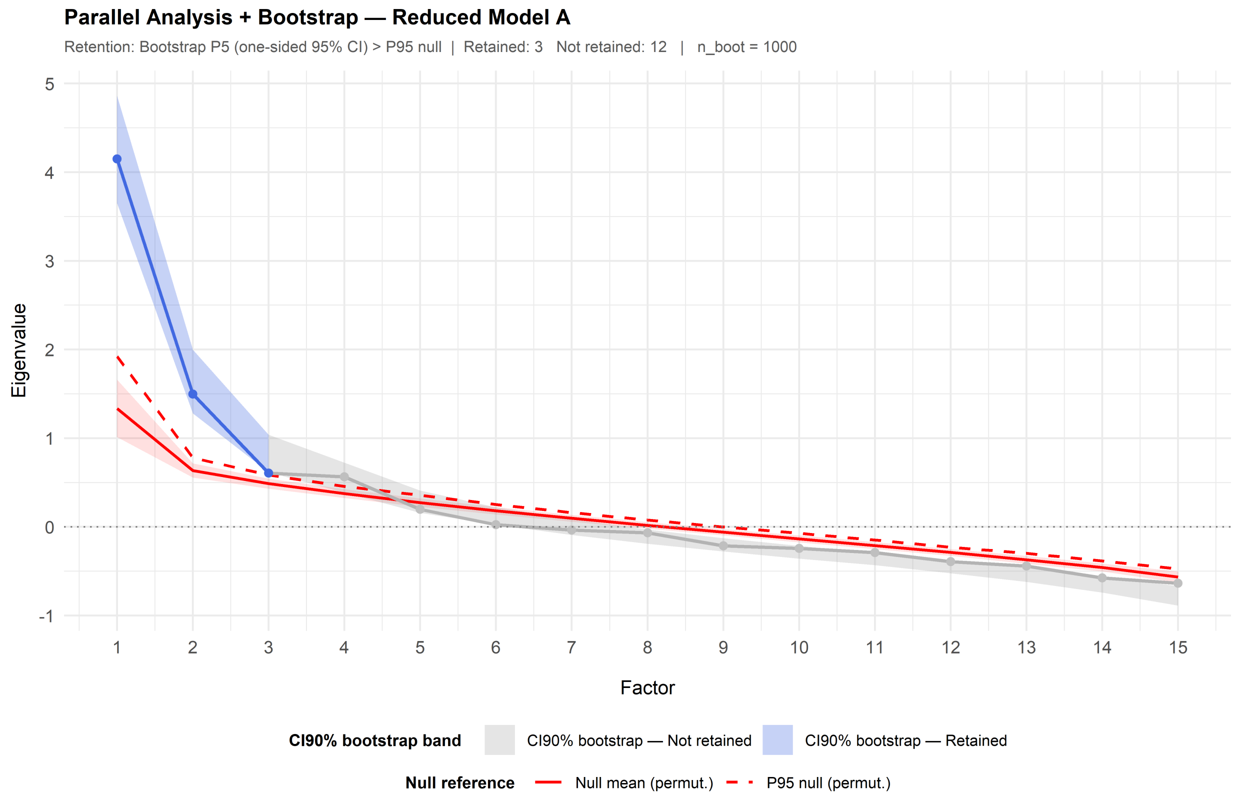

```r
fa_reducedA <- run_fa_psych_selected(
  selected_obj = reduced_selected_matrix_A,
  data = emfra_items_reducedA,
  model_name = "Reduced Model A",
  pa_obj = boot_pa_reducedA,
  fm = "minres",
  rotate = "oblimin",
  n.iter = 1000,
  SMC = TRUE,
  smooth = FALSE,
  warnings = FALSE,
  residuals = FALSE,
  scores = "tenBerge"
)

fa_fit_reducedA <- extract_fa_fit_table(
  fa_obj = fa_reducedA
)
```

```r
fa_fit_full_rma <- dplyr::bind_rows(
  fa_fit_full,
  fa_fit_reducedA
)
```

Table 24: Exploratory Factor Analysis of Reduced Model A

| Model | No. factors | Proportion Var | Chi-square | p-value | TLI | RMSEA | RMSR | Model fit | Error fit |
|---|---|---|---|---|---|---|---|---|---|
| Full Model | 3 | MR1: 0.199; MR2: 0.138; MR3: 0.075 | 992.864 | 0 | 0.446 | 0.167 | 0.074 | poor model fit | poor-to-marginal residual fit |
| Reduced Model A | 3 | MR1: 0.230; MR2: 0.161; MR3: 0.090 | 405.660 | 0 | 0.551 | 0.164 | 0.073 | poor model fit | poor-to-marginal residual fit |

##### 4.7.3 Dimensional diagnosis - EGA

```r
emfra_items_reducedA_ega <- emfra_items_reducedA
colnames(emfra_items_reducedA_ega) <- emfra_item_labels_short[
  !emfra_item_ids %in% excluded_item_ids_rma
]

reduced_selected_matrix_A_ega <- select_polychoric_matrices(
  data = emfra_items_reducedA_ega
)

bootega_reducedA <- run_bootega_selected(
  data          = emfra_items_reducedA_ega,
  selected_obj = reduced_selected_matrix_A_ega,
  model_name    = "Reduced Model A",
  iter          = 1000,
  seed          = 123,
  ncores        = 1,
  plot.EGA      = TRUE,
  verbose       = TRUE
)
```

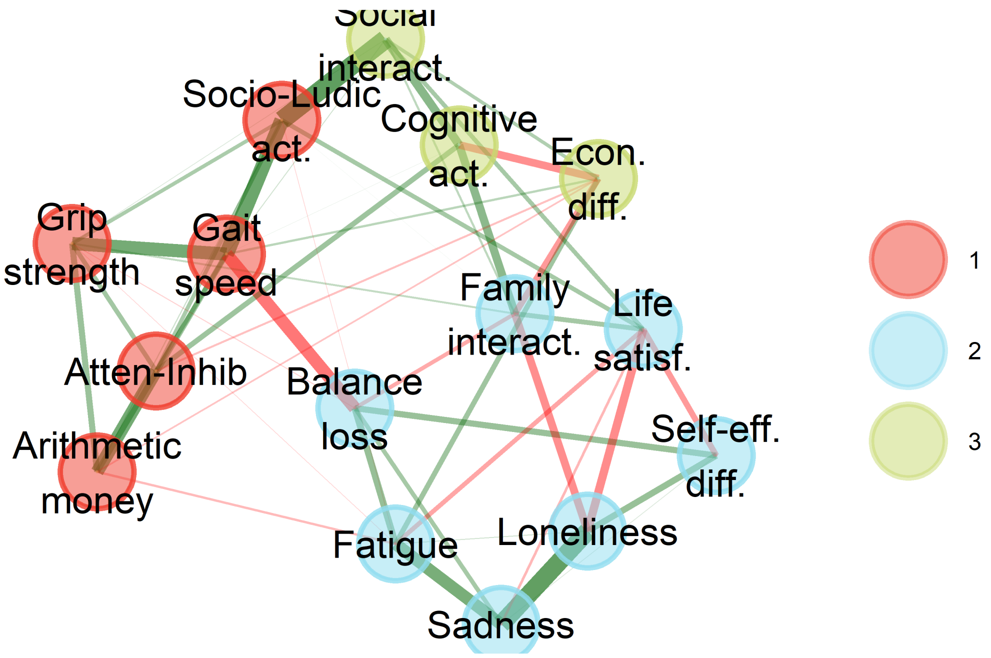

```
ndim_freq_full_rma <- build_ndim_frequency_table(
  boot_list = list(
    "Full Model"      = bootega_full$boot,
    "Reduced Model A" = bootega_reducedA$boot
  ),
  n_factors_range = 1:7
)

plot_ndim_frequency(
  freq_tbl = ndim_freq_full_rma,
  y_limit  = 70,
  facet    = TRUE
)
```

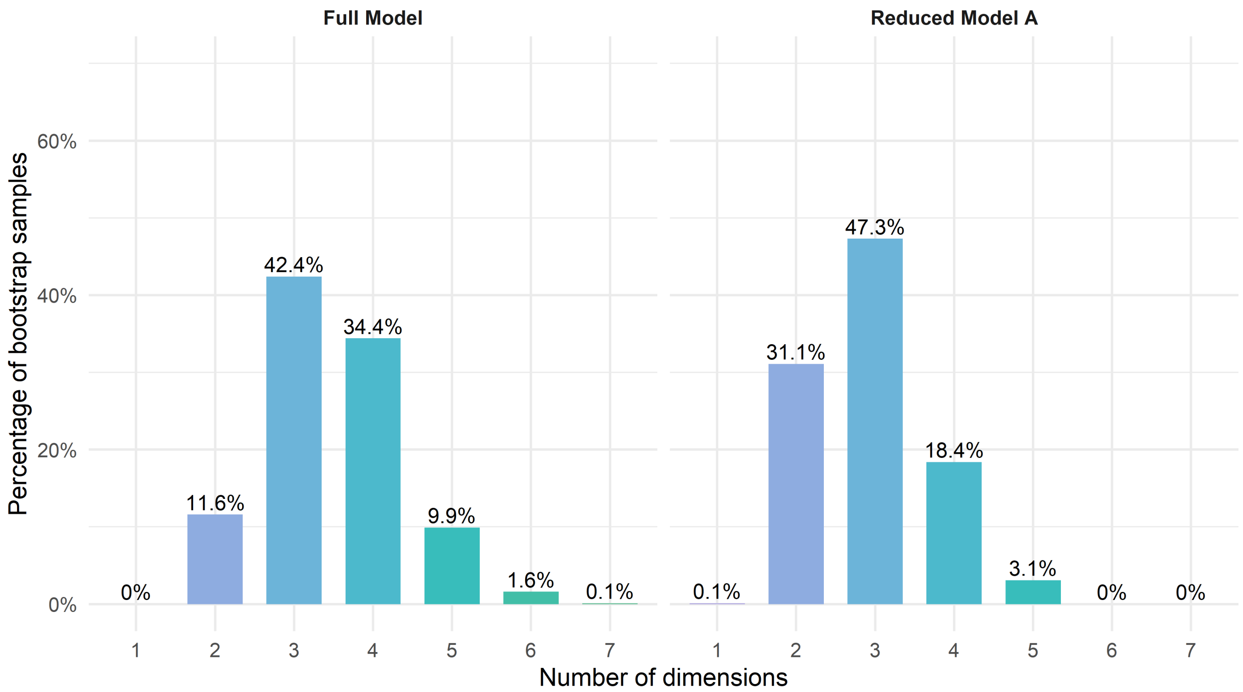

Figure 18: Frequency of the number of communities across bootstrap samples in the Full and Reduced Model A

```r
ndim_summary_reducedA <- summarise_ndim_frequency(
  freq_tbl   = ndim_freq_full_rma,
  model_name = "Reduced Model A",
  top_n      = 4,
  digits     = 1
)

ndim_summary_full_rma <- dplyr::bind_rows(
  ndim_summary_full,
  ndim_summary_reducedA
)
```

Table 25: Dimensional summary of EGA dimensions in the Full and Reduced A Models

| Model | Modal solution | Ranked dimensional solutions | Difference 1st-2nd | Difference 1st-3rd | Difference 1st-4th |
|---|---|---|---|---|---|
| Full Model | 3 dimensions (42.4%) | 3 dimensions: 42.4%; 4 dimensions: 34.4%; 2 dimensions: 11.6%; 5 dimensions: 9.9% | 8% | 30.8% | 32.5% |
| Reduced Model A | 3 dimensions (47.3%) | 3 dimensions: 47.3%; 2 dimensions: 31.1%; 4 dimensions: 18.4%; 5 dimensions: 3.1% | 16.2% | 28.9% | 44.2% |

```r
structural_consistency_reducedA <- extract_structural_consistency(
  boot = bootega_reducedA$boot,
  model = "Reduced Model A"
)

structural_consistency_full_rma <- dplyr::bind_rows(
  structural_consistency_full,
  structural_consistency_reducedA
)
```

Table 26: Structural consistency of EGA dimensions in the Full and Reduced A Models

| Model | Dim 1 | Dim 2 | Dim 3 |
|---|---|---|---|
| Full Model | 30.8% | 19.7% | 15.1% |
| Reduced Model A | 44.5% | 16.2% | 29.9% |

###### 4.7.4 Dimensional diagnosis - Model acceptance criteria

```r
dimensional_decision_reducedA <- build_dimensional_decision_table(
  model_name = "Reduced Model A",
  pa_obj = boot_pa_reducedA,
  fa_fit_table = fa_fit_reducedA,
  ndim_summary = ndim_summary_reducedA,
  structural_consistency = structural_consistency_reducedA,
  previous_model_comparison = "Non relevant improvement in Model and Error Fit in
  ↪  FA over previous model. Slight improvement in EGA dimensionality measures.",
  researcher_judgement = c(
    "Convergence between Bootstrap PA and modal Bootstrap EGA",
    "Poor model performance",
    "Poor model performance",
    "Similar performance",
    "Do not retain. Proceed with item reduction."
  )
)
```

Table 27: Dimensional acceptability criteria of the Reduced Model A

| Model | Criterion | Objective evidence | Researcher judgement |
|---|---|---|---|
| Reduced Model A | 1. Bootstrap PA-EGA dimensional convergence | Bootstrap PA suggested 3 factors; EGA modal solution was 3 dimensions (47.3%). | Convergence between Bootstrap PA and modal Bootstrap EGA |

| Model | Criterion | Objective evidence | Researcher judgement |
|---|---|---|---|
| Reduced Model A | 2. FA dimensional performance | Proportion Var. = MR1: 0.230; MR2: 0.161; MR3: 0.090; model fit = poor model fit; error fit = poor-to-marginal residual fit. | Poor model performance |
| Reduced Model A | 3. EGA dimensional stability | Modal solution = 3 dimensions (47.3%); differences: 1st-2nd = 16.2%, 1st-3rd = 28.9%, 1st-4th = 44.2%; structural consistency = Dim 1: 44.5%; Dim 2: 16.2%; Dim 3: 29.9%. | Poor model performance |
| Reduced Model A | 4. Improvement over previous model | Non relevant improvement in Model and Error Fit in FA over previous model. Slight improvement in EGA dimensionality measures. | Similar performance |
| Reduced Model A | 5. Model retention | Final model decision based on the previous dimensional criteria (1-4) | Do not retain. Proceed with item reduction. |

##### 4.7.5 Item diagnosis - PA and FA

```
fa_item_table_reducedA <- build_fa_item_table(
  fa_obj = fa_reducedA,
  model_name = "Reduced Model A",
  item_ids = emfra_item_ids_reducedA,
  item_labels = emfra_item_labels_reducedA,
  item_labels_short = emfra_item_labels_short_reducedA,
  loading_cutoff = 0.30,
  primary_loading_cutoff = 0.50,
  communality_cutoff = 0.30,
  digits = 3
)
```

Table 28: Factor analysis item diagnostics for the Reduced Model A

| Model | Item number | Item name | Communality | MR1 | MR2 | MR3 | Low loadings | High cross-loadings | Low communalities |
|---|---|---|---|---|---|---|---|---|---|
| Reduced Model A | Item 1 | Grip strength | 0.483 | 0.690 | -0.016 | 0.032 | No | No | No |
| Reduced Model A | Item 2 | Gait speed | 0.737 | 0.844 | -0.028 | 0.112 | No | No | No |
| Reduced Model A | Item 4 | Balance loss | 0.359 | -0.348 | 0.375 | 0.071 | No | No | No |
| Reduced Model A | Item 5 | Fatigue | 0.351 | -0.168 | 0.478 | 0.134 | No | No | No |
| Reduced Model A | Item 6 | Arithmetic money | 0.354 | 0.565 | 0.036 | -0.217 | No | No | No |
| Reduced Model A | Item 7 | Atten-Inhib | 0.416 | 0.632 | 0.126 | -0.246 | No | No | No |
| Reduced Model A | Item 10 | Cognitive act. | 0.280 | 0.425 | 0.063 | -0.345 | No | No | Yes |
| Reduced Model A | Item 11 | Loneliness | 0.641 | 0.071 | 0.812 | 0.034 | No | No | No |
| Reduced Model A | Item 12 | Sadness | 0.531 | 0.011 | 0.736 | -0.021 | No | No | No |

| Model | Item number | Item name | Communality | MR1 | MR2 | MR3 | Low loadings | High cross-loadings | Low communalities |
|---|---|---|---|---|---|---|---|---|---|
| Reduced Model A | Item 14 | Self-eff. diff. | 0.252 | 0.050 | 0.523 | -0.048 | No | No | Yes |
| Reduced Model A | Item 15 | Life satisf. | 0.411 | 0.173 | -0.564 | -0.011 | No | No | No |
| Reduced Model A | Item 16 | Econ. diff. | 1.016 | 0.049 | 0.038 | 0.999 | No | No | No |
| Reduced Model A | Item 17 | Family interact. | 0.289 | 0.221 | -0.346 | -0.210 | No | No | Yes |
| Reduced Model A | Item 18 | Social interact. | 0.398 | 0.609 | -0.050 | 0.092 | No | No | No |
| Reduced Model A | Item 19 | Socio-Ludic act. | 0.704 | 0.815 | -0.061 | 0.093 | No | No | No |

```
fa_candidate_items_reducedA <- build_fa_candidate_items_table(
  fa_item_table = fa_item_table_reducedA,
  digits = 3
)
```

Table 29: Candidate items for elimination in FA from Reduced Model A

| Model | Item number | Item name | Low factor loadings reason | High cross-loadings reason | Low communality reason |
|---|---|---|---|---|---|
| Reduced Model A | Item 10 | Cognitive act. | No | No | Communality = 0.280 |
| Reduced Model A | Item 14 | Self-eff. diff. | No | No | Communality = 0.252 |
| Reduced Model A | Item 17 | Family interact. | No | No | Communality = 0.289 |

##### 4.7.6 Item diagnosis - EGA

```
item_stability_reducedA <- extract_item_stability(
  boot = bootega_reducedA$boot,
  model = "Reduced Model A",
  item_ids = emfra_item_ids_reducedA,
  item_labels = emfra_item_labels_reducedA,
  item_labels_short = emfra_item_labels_short_reducedA,
  stability_cutoff = 0.75
)

item_stability_table_reducedA <- item_stability_reducedA$table
item_stability_plot_reducedA <- item_stability_reducedA$plot
```

```
item_stability_plot_reducedA
```

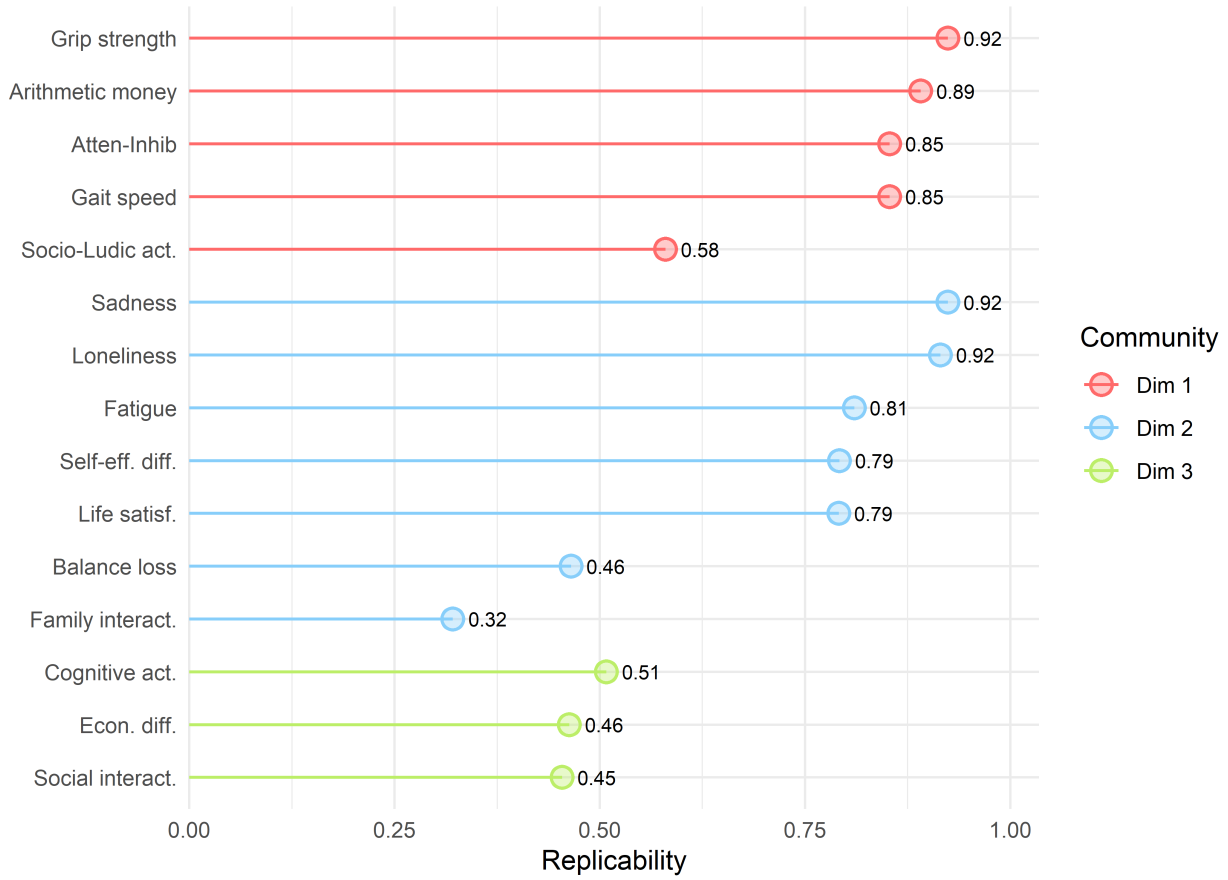

Figure 19: Item replicability from Reduced Model A

```
ega_redundancy_reducedA <- build_ega_redundancy_table(
  data = emfra_items_reducedA_ega,
  selected_obj = reduced_selected_matrix_A_ega,
  model_name = "Reduced Model A",
  uva.method = "MBR",
  cut.off = 0.25,
  reduce = FALSE,
  report_cutoff = 0.20
)

ega_redundancy_table_reducedA <- ega_redundancy_reducedA$table
```

Table 30: Item redundancy from Reduced Model A

| Model | Item 1 | Item 2 | wTO value | wTO cut-off | Redundancy level |
|---|---|---|---|---|---|
| Reduced Model A | Item 18. Social interaction | Item 19. Social-ludic activities | 0.297 | 0.25 | Moderate-to-large |
| Reduced Model A | Item 11. Loneliness | Item 12. Sadness | 0.292 | 0.25 | Moderate-to-large |
| Reduced Model A | Item 1. Hand-grip strength | Item 2. Gait speed | 0.246 | 0.2 | Small-to-moderate |
| Reduced Model A | Item 6. Arithmetics for money | Item 7. Attention-inhibition | 0.232 | 0.2 | Small-to-moderate |
| Reduced Model A | Item 2. Gait speed | Item 19. Social-ludic activities | 0.219 | 0.2 | Small-to-moderate |
| Reduced Model A | Item 5. Fatigue | Item 12. Sadness | 0.201 | 0.2 | Small-to-moderate |

```
ega_item_diagnostics_reducedA <- run_item_diagnostics_selected(
  data         = emfra_items_reducedA_ega,
  selected_obj = reduced_selected_matrix_A_ega,
  model_name   = "Reduced Model A",
  iter         = 1000,
  ncores       = 1,
  seed         = 123
)

ega_item_diagnostics_table_reducedA <- ega_item_diagnostics_reducedA$table
```

Table 31: Item diagnostics (Minor dimension, Two-item dimension, Multidimensional) from Reduced Model A

| Model | Item number | Item name | Diagnostic |
|---|---|---|---|

```
ega_loadings_reducedA <- extract_ega_community_loadings(
  boot = bootega_reducedA$boot,
  model = "Reduced Model A",
  item_ids = emfra_item_ids_reducedA,
  item_labels = emfra_item_labels_reducedA,
  item_labels_short = emfra_item_labels_short_reducedA,
  loading.method = "revised",
  scaling = 2,
  rotation = "oblimin",
  loading_cutoff = 0.30,
  primary_loading_cutoff = 0.50,
  digits = 3
```

```r
)

ega_loadings_table_reducedA <- ega_loadings_reducedA$table
```

Table 32: Item loadings in EGA from Reduced Model A

| Model | Item number | Item name | 1 | 2 | 3 | Low loadings | High cross-loadings |
|---|---|---|---|---|---|---|---|
| Reduced Model A | Item 2 | Gait speed | 0.601 | -0.077 | -0.059 | No | No |
| Reduced Model A | Item 1 | Grip strength | 0.435 | 0.014 | -0.008 | No | No |
| Reduced Model A | Item 7 | Atten-Inhib | 0.371 | 0.084 | 0.172 | No | No |
| Reduced Model A | Item 6 | Arithmetic money | 0.400 | 0.031 | 0.025 | No | No |
| Reduced Model A | Item 19 | Socio-Ludic act. | 0.284 | 0.048 | 0.449 | No | No |
| Reduced Model A | Item 11 | Loneliness | -0.003 | 0.600 | 0.008 | No | No |
| Reduced Model A | Item 12 | Sadness | -0.003 | 0.544 | 0.008 | No | No |
| Reduced Model A | Item 5 | Fatigue | -0.029 | 0.321 | -0.129 | No | No |
| Reduced Model A | Item 4 | Balance loss | -0.173 | 0.290 | 0.014 | Yes | No |
| Reduced Model A | Item 14 | Self-eff. diff. | -0.002 | 0.279 | 0.004 | Yes | No |
| Reduced Model A | Item 17 | Family interact. | -0.064 | -0.179 | 0.428 | No | No |
| Reduced Model A | Item 15 | Life satisf. | 0.036 | -0.355 | 0.107 | No | No |
| Reduced Model A | Item 10 | Cognitive act. | -0.058 | -0.003 | 0.624 | No | No |
| Reduced Model A | Item 18 | Social interact. | 0.215 | -0.020 | 0.185 | Yes | No |
| Reduced Model A | Item 16 | Econ. diff. | 0.013 | 0.117 | -0.158 | Yes | No |

```r
ega_candidate_items_reducedA <- build_ega_candidate_items_table(
  item_stability_table       = item_stability_table_reducedA,
  ega_redundancy_table       = ega_redundancy_table_reducedA,
  ega_loadings_table         = ega_loadings_table_reducedA,
  ega_item_diagnostics_table = ega_item_diagnostics_table_reducedA,
  stability_cutoff           = 0.75,
  redundancy_cutoff          = 0.25,
```

```
  loading_cutoff              = 0.30,
  primary_loading_cutoff      = 0.50,
  digits                      = 3
)
```

Table 33: Candidate items for elimination in EGA from Reduced Model A

| Model | Item number | Item name | EGA candidate criterion | Low item stability reason | Redundancy reason | Item diagnostics reason | Low community loading reason | High community cross-loading reason |
|---|---|---|---|---|---|---|---|---|
| Reduced Model A | Item 4 | Balance loss | Low replicability + Low community loading | Replicability = 46.5% (Dim 2) | No | No | Highest abs. community loading = 0.290 (Dim 2) | No |
| Reduced Model A | Item 10 | Cognitive act. | Low replicability | Replicability = 50.8% (Dim 3) | No | No | No | No |
| Reduced Model A | Item 11 | Loneliness | Redundancy | No | Redundant with Item 12. Sadness (wTO = 0.292; 0.25; Moderate-to-large) | No | No | No |
| Reduced Model A | Item 12 | Sadness | Redundancy | No | Redundant with Item 11. Loneliness (wTO = 0.292; 0.25; Moderate-to-large) | No | No | No |
| Reduced Model A | Item 14 | Self-eff. diff. | Low community loading | No | No | No | Highest abs. community loading = 0.279 (Dim 2) | No |
| Reduced Model A | Item 16 | Econ. diff. | Low replicability + Low community loading | Replicability = 46.3% (Dim 3) | No | No | Highest abs. community loading = 0.158 (Dim 3) | No |
| Reduced Model A | Item 17 | Family inter-act. | Low replicability | Replicability = 32.1% (Dim 2) | No | No | No | No |
| Reduced Model A | Item 18 | Social inter-act. | Low replicability + Redundancy + Low community loading | Replicability = 45.4% (Dim 3) | Redundant with Item 19. Social-ludic activities (wTO = 0.297; 0.25; Moderate-to-large) | No | Highest abs. community loading = 0.215 (Dim 1) | No |
| Reduced Model A | Item 19 | Socio-Ludic act. | Low replicability + Redundancy | Replicability = 58.0% (Dim 1) | Redundant with Item 18. Social interaction (wTO = 0.297; 0.25; Moderate-to-large) | No | No | No |

##### 4.7.7 Item diagnosis - Retention and elimination of items

```
item_reduction_summary_reducedA <- build_item_reduction_summary_table(
  model_name = "Reduced Model A",
  fa_candidate_items_table = fa_candidate_items_reducedA,
  ega_candidate_items_table = ega_candidate_items_reducedA,
  all_item_labels = emfra_item_labels_reducedA,
  all_item_labels_short = emfra_item_labels_short_reducedA
)
```

Table 34: Item summary diagnostics from FA and EGA in Reduced Model A

| Model | Item number | Item name | FA diagnostic summary | EGA diagnostic summary |
|---|---|---|---|---|
| Reduced Model A | Item 1 | Grip strength | No FA diagnostic issue detected. | No EGA diagnostic issue detected. |
| Reduced Model A | Item 2 | Gait speed | No FA diagnostic issue detected. | No EGA diagnostic issue detected. |
| Reduced Model A | Item 4 | Balance loss | No FA diagnostic issue detected. | Low item stability: 46.5% (Dim 2); Low community loading: 0.290 (Dim 2) |
| Reduced Model A | Item 5 | Fatigue | No FA diagnostic issue detected. | No EGA diagnostic issue detected. |
| Reduced Model A | Item 6 | Arithmetic money | No FA diagnostic issue detected. | No EGA diagnostic issue detected. |
| Reduced Model A | Item 7 | Atten-Inhib | No FA diagnostic issue detected. | No EGA diagnostic issue detected. |
| Reduced Model A | Item 10 | Cognitive act. | Low communality: h2 = 0.280 | Low item stability: 50.8% (Dim 3) |
| Reduced Model A | Item 11 | Loneliness | No FA diagnostic issue detected. | Redundancy with Item 12. Sadness (wTO = 0.292) |
| Reduced Model A | Item 12 | Sadness | No FA diagnostic issue detected. | Redundancy with Item 11. Loneliness (wTO = 0.292) |
| Reduced Model A | Item 14 | Self-eff. diff. | Low communality: h2 = 0.252 | Low community loading: 0.279 (Dim 2) |
| Reduced Model A | Item 15 | Life satisf. | No FA diagnostic issue detected. | No EGA diagnostic issue detected. |
| Reduced Model A | Item 16 | Econ. diff. | No FA diagnostic issue detected. | Low item stability: 46.3% (Dim 3); Low community loading: 0.158 (Dim 3) |
| Reduced Model A | Item 17 | Family interact. | Low communality: h2 = 0.289 | Low item stability: 32.1% (Dim 2) |
| Reduced Model A | Item 18 | Social interact. | No FA diagnostic issue detected. | Low item stability: 45.4% (Dim 3); Redundancy with Item 19. Social-ludic activities (wTO = 0.297); Low community loading: 0.215 (Dim 1) |
| Reduced Model A | Item 19 | Socio-Ludic act. | No FA diagnostic issue detected. | Low item stability: 58.0% (Dim 1); Redundancy with Item 18. Social interaction (wTO = 0.297) |

The following items were detected as conflictive through both FA and EGA, and they would be eliminated: Item 10 (Cognitive act.), Item 14 (Selff-efficacy difficulty), Item 17 (Family interaction). In addition, item 18 (Social interaction) would be eliminated due to its high redundancy with item 19 (social-ludic activities), with a relativelly high zero-order correlation (0.67) and moderate partial correlation (0.45), but presenting a lower replicability rate compared to its redundant pair (58% vs 45%).

#### 4.8 Reduced Model B

##### 4.8.1 Matrix diagnosis and selection

```
excluded_item_ids_rmb <- c(
  "EMFRA_item3_trait",   # Item 3. Gait assistance
  "EMFRA_item8_trait",   # Item 8. Memory difficulty
  "EMFRA_item9_trait",   # Item 9. Concentration difficulty.
  "EMFRA_item10_trait",  # Item 10. Cognitive activities
  "EMFRA_item13_trait",  # Item 13. Irritability
  "EMFRA_item14_trait",  # Item 14. Self-efficacy difficulty
  "EMFRA_item17_trait",  # Item 17. Family interaction
  "EMFRA_item18_trait",  # Item 18. Social interaction
  "EMFRA_item20_trait",  # Item 20. Communication difficulty
  "EMFRA_item21_trait"  # Item 21. Help availability
)

excluded_labels_rmb <- emfra_item_labels_short[
  match(excluded_item_ids_rmb, emfra_item_ids)
]

emfra_item_ids_reducedB <- emfra_item_ids[
  !emfra_item_ids %in% excluded_item_ids_rmb
]

emfra_item_labels_reducedB <- emfra_item_labels[
  match(emfra_item_ids_reducedB, emfra_item_ids)
]

emfra_item_labels_short_reducedB <- emfra_item_labels_short[
  match(emfra_item_ids_reducedB, emfra_item_ids)
]

label_map_reducedB <- setNames(
  emfra_item_labels_reducedB,
  emfra_item_labels_short_reducedB
)

emfra_items_reducedB <- data_emfra %>%
```

```r
  dplyr::select(all_of(emfra_item_ids_reducedB))

colnames(emfra_items_reducedB) <- emfra_item_labels_short_reducedB

reduced_selected_matrix_B <- select_polychoric_matrices(emfra_items_reducedB)

matrix_diagnostics_reducedB <- build_matrix_diagnostics_table(
  selected_obj = reduced_selected_matrix_B,
  model_name   = "Reduced Model B"
)
```

Table 35: Selected matrix for the Reduced Model B

| Model | Package | Correction | Selected for dim. analysis | Symmetric | PSD | Total eigenvalues (n) | Negative eigenvalues (n) | Minimum eigenvalue | Negative eigenvalues |
|---|---|---|---|---|---|---|---|---|---|
| Reduced Model B | psych | 0 | Yes | TRUE | TRUE | 11 | 0 | 0.173 | NA |
| Reduced Model B | EGAnet | 0 | Yes | TRUE | TRUE | 11 | 0 | 0.187 | NA |

##### 4.8.2 Dimensional diagnosis - Bootstrap PA and FA

```r
if (!exists("boot_pa_reducedB")) {

  boot_pa_reducedB <- run_parallel_bootstrap(
    selected_obj = reduced_selected_matrix_B,
    data         = emfra_items_reducedB,
    model_name   = "Reduced Model B"
  )

  dir.create("cache", showWarnings = FALSE)
  saveRDS(boot_pa_reducedB, ruta_cache_pa_reducedB)
}
```

Table 36: Bootstrap Parallel Analysis — Reduced Model B

| Factor | Eigenvalue | Null_mean | Null_P95 | Boot_mean | Boot_P5 | Boot_P95 | Retained - Horn's PA (P95) | Retained - Bootstrap PA (90% CI, one-sided) | Non-overlap with permuted eigenvalues (%) |
|---|---|---|---|---|---|---|---|---|---|
| 1 | 3.421 | 1.202 | 1.890 | 3.520 | 3.032 | 4.056 | Yes | Yes | 100.0 |
| 2 | 1.209 | 0.484 | 0.614 | 1.286 | 0.977 | 1.631 | Yes | Yes | 100.0 |
| 3 | 0.403 | 0.335 | 0.434 | 0.482 | 0.327 | 0.648 | No | No | 68.1 |
| 4 | 0.128 | 0.222 | 0.308 | 0.213 | 0.103 | 0.334 | No | No | 9.2 |
| 5 | -0.007 | 0.124 | 0.195 | 0.056 | -0.039 | 0.159 | No | No | 1.3 |
| 6 | -0.075 | 0.033 | 0.095 | -0.066 | -0.146 | 0.012 | No | No | 0.0 |
| 7 | -0.138 | -0.055 | 0.007 | -0.169 | -0.251 | -0.092 | No | No | 0.0 |
| 8 | -0.215 | -0.142 | -0.077 | -0.272 | -0.352 | -0.189 | No | No | 0.0 |
| 9 | -0.313 | -0.232 | -0.163 | -0.380 | -0.468 | -0.299 | No | No | 0.0 |
| 10 | -0.446 | -0.331 | -0.254 | -0.501 | -0.597 | -0.416 | No | No | 0.0 |
| 11 | -0.545 | -0.451 | -0.355 | -0.649 | -0.788 | -0.536 | No | No | 0.0 |

```r
plot_parallel_bootstrap_ggplot(boot_pa_reducedB)
```

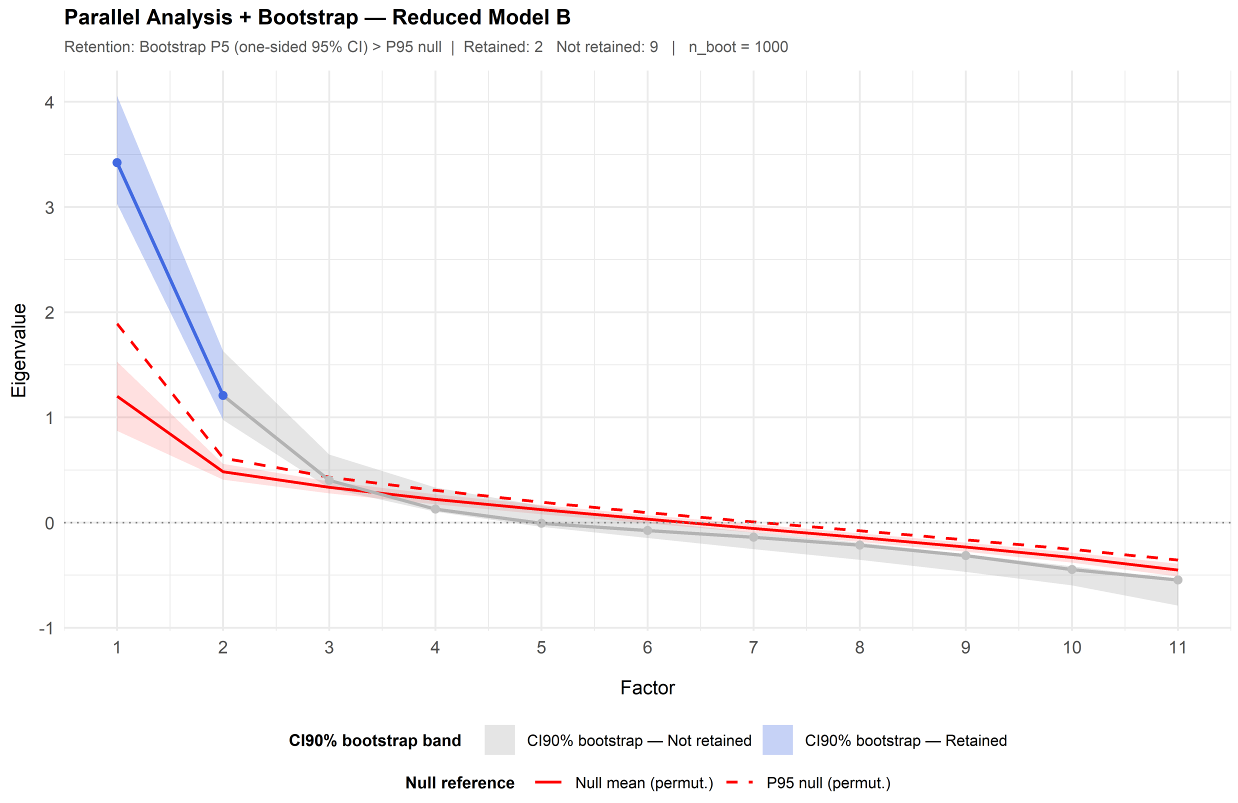

```r
fa_reducedB <- run_fa_psych_selected(
  selected_obj = reduced_selected_matrix_B,
  data = emfra_items_reducedB,
  model_name = "Reduced Model B",
  pa_obj = boot_pa_reducedB,
  fm = "minres",
  rotate = "oblimin",
  n.iter = 1000,
  SMC = TRUE,
  smooth = FALSE,
  warnings = FALSE,
  residuals = FALSE,
  scores = "tenBerge"
)

fa_fit_reducedB <- extract_fa_fit_table(
  fa_obj = fa_reducedB
)
```

```r
fa_fit_full_rmb <- dplyr::bind_rows(
  fa_fit_full,
  fa_fit_reducedA,
  fa_fit_reducedB
)
```

Table 37: Exploratory Factor Analysis of Reduced Model B

| Model | No. factors | Proportion Var | Chi-square | p-value | TLI | RMSEA | RMSR | Model fit | Error fit |
|---|---|---|---|---|---|---|---|---|---|
| Full Model | 3 | MR1: 0.199; MR2: 0.138; MR3: 0.075 | 992.864 | 0 | 0.446 | 0.167 | 0.074 | poor model fit | poor-to-marginal residual fit |
| Reduced Model A | 3 | MR1: 0.230; MR2: 0.161; MR3: 0.090 | 405.660 | 0 | 0.551 | 0.164 | 0.073 | poor model fit | poor-to-marginal residual fit |
| Reduced Model B | 2 | MR1: 0.260; MR2: 0.195 | 193.478 | 0 | 0.691 | 0.153 | 0.073 | poor model fit | poor-to-marginal residual fit |

##### 4.8.3 Dimensional diagnosis - EGA

```r
emfra_items_reducedB_ega <- emfra_items_reducedB
colnames(emfra_items_reducedB_ega) <- emfra_item_labels_short[
  !emfra_item_ids %in% excluded_item_ids_rmb
]

reduced_selected_matrix_B_ega <- select_polychoric_matrices(
  data = emfra_items_reducedB_ega
)

bootega_reducedB <- run_bootega_selected(
  data         = emfra_items_reducedB_ega,
  selected_obj = reduced_selected_matrix_B_ega,
  model_name   = "Reduced Model B",
  iter         = 1000,
  seed         = 123,
  ncores       = 1,
  plot.EGA     = TRUE,
  verbose      = TRUE
)
```

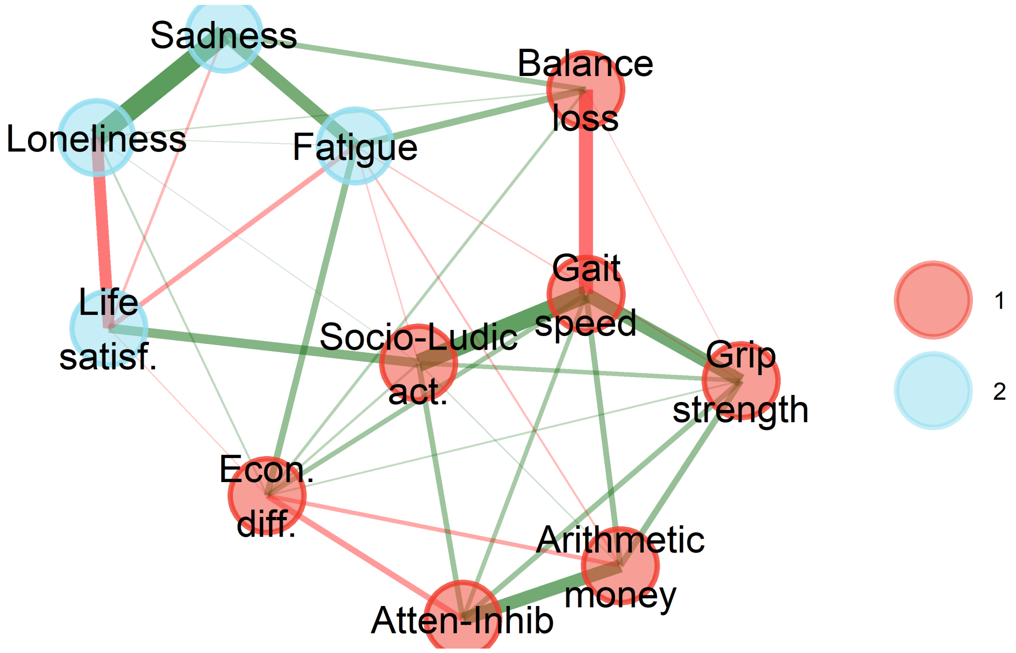

```
ndim_freq_full_rmb <- build_ndim_frequency_table(
  boot_list = list(
    "Full Model"      = bootega_full$boot,
    "Reduced Model A" = bootega_reducedA$boot,
    "Reduced Model B" = bootega_reducedB$boot
  ),
  n_factors_range = 1:7
)

plot_ndim_frequency(
  freq_tbl = ndim_freq_full_rmb,
  y_limit  = 80,
  facet    = TRUE
)
```

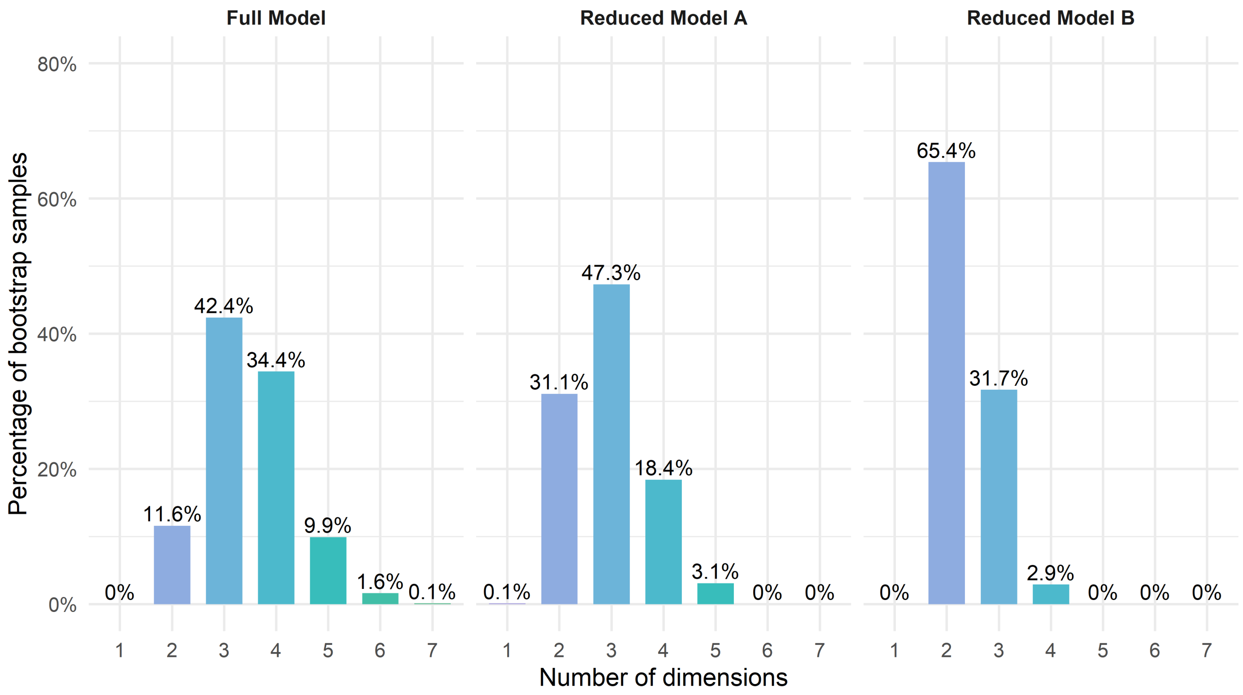

Figure 20: Frequency of the number of communities across bootstrap samples in the Full and Reduced Models A-B

```r
ndim_summary_reducedB <- summarise_ndim_frequency(
  freq_tbl   = ndim_freq_full_rmb,
  model_name = "Reduced Model B",
  top_n      = 4,
  digits     = 1
)

ndim_summary_full_rmb <- dplyr::bind_rows(
  ndim_summary_full,
  ndim_summary_reducedA,
  ndim_summary_reducedB
)
```

Table 38: Dimensional summary of EGA dimensions in the Full and Reduced A-B Models

| Model | Modal solution | Ranked dimensional solutions | Difference 1st-2nd | Difference 1st-3rd | Difference 1st-4th |
|---|---|---|---|---|---|
| Full Model | 3 dimensions (42.4%) | 3 dimensions: 42.4%; 4 dimensions: 34.4%; 2 dimensions: 11.6%; 5 dimensions: 9.9% | 8% | 30.8% | 32.5% |
| Reduced Model A | 3 dimensions (47.3%) | 3 dimensions: 47.3%; 2 dimensions: 31.1%; 4 dimensions: 18.4%; 5 dimensions: 3.1% | 16.2% | 28.9% | 44.2% |

| Model | Modal solution | Ranked dimensional solutions | Difference 1st-2nd | Difference 1st-3rd | Difference 1st-4th |
|---|---|---|---|---|---|
| Reduced Model B | 2 dimensions (65.4%) | 2 dimensions: 65.4%; 3 dimensions: 31.7%; 4 dimensions: 2.9% | 33.7% | 62.5% | NA |

```
structural_consistency_reducedB <- extract_structural_consistency(
  boot = bootega_reducedB$boot,
  model = "Reduced Model B"
)

structural_consistency_full_rmb <- dplyr::bind_rows(
  structural_consistency_full,
  structural_consistency_reducedA,
  structural_consistency_reducedB
)
```

Table 39: Structural consistency of EGA dimensions in the Full and Reduced A-B Models

| Model | Dim 1 | Dim 2 | Dim 3 |
|---|---|---|---|
| Full Model | 30.8% | 19.7% | 15.1% |
| Reduced Model A | 44.5% | 16.2% | 29.9% |
| Reduced Model B | 29.9% | 70.2% | NA |

##### 4.8.4 Dimensional diagnosis - Model acceptance criteria

```
dimensional_decision_reducedB <- build_dimensional_decision_table(
  model_name = "Reduced Model B",
  pa_obj = boot_pa_reducedB,
  fa_fit_table = fa_fit_reducedB,
  ndim_summary = ndim_summary_reducedB,
  structural_consistency = structural_consistency_reducedB,
  previous_model_comparison = "Greater reduction of Chi-square value, and increase
   ↪  in TLI, as slight reduction in RMSEA. Greater improvement in Bootstrap EGA
   ↪  modal solution, and its structural consistency.",
  researcher_judgement = c(
    "Convergence between Bootstrap PA and modal Bootstrap EGA",
    "Better error fit measures",
    "Good model performance",
    "Relevant improvement",
    "Proceed with further item reduction, and check model performance."
  )
)
```

Table 40: Dimensional acceptability criteria of the Reduced Model B

| Model | Criterion | Objective evidence | Researcher judgement |
|---|---|---|---|
| Reduced Model B | 1. Bootstrap PA-EGA dimensional convergence | Bootstrap PA suggested 2 factors; EGA modal solution was 2 dimensions (65.4%). | Convergence between Bootstrap PA and modal Bootstrap EGA |
| Reduced Model B | 2. FA dimensional performance | Proportion Var. = MR1: 0.260; MR2: 0.195; model fit = poor model fit; error fit = poor-to-marginal residual fit. | Better error fit measures |
| Reduced Model B | 3. EGA dimensional stability | Modal solution = 2 dimensions (65.4%); differences: 1st-2nd = 33.7%, 1st-3rd = 62.5%, 1st-4th = NA; structural consistency = Dim 1: 29.9%; Dim 2: 70.2%. | Good model performance |
| Reduced Model B | 4. Improvement over previous model | Greater reduction of Chi-square value, and increase in TLI, as slight reduction in RMSEA. Greater improvement in Bootstrap EGA modal solution, and its structural consistency. | Relevant improvement |
| Reduced Model B | 5. Model retention | Final model decision based on the previous dimensional criteria (1-4) | Proceed with further item reduction, and check model performance. |

##### 4.8.5 Item diagnosis - PA and FA

```
fa_item_table_reducedB <- build_fa_item_table(
  fa_obj = fa_reducedB,
  model_name = "Reduced Model B",
  item_ids = emfra_item_ids_reducedB,
  item_labels = emfra_item_labels_reducedB,
  item_labels_short = emfra_item_labels_short_reducedB,
  loading_cutoff = 0.30,
  primary_loading_cutoff = 0.50,
  communality_cutoff = 0.30,
  digits = 3
)
```

Table 41: Factor analysis item diagnostics for the Reduced Model B

| Model | Item number | Item name | Communality | MR1 | MR2 | Low loadings | High cross-loadings | Low communalities |
|---|---|---|---|---|---|---|---|---|
| Reduced Model B | Item 1 | Grip strength | 0.537 | 0.746 | 0.042 | No | No | No |
| Reduced Model B | Item 2 | Gait speed | 0.776 | 0.890 | 0.026 | No | No | No |
| Reduced Model B | Item 4 | Balance loss | 0.362 | -0.393 | 0.343 | No | No | No |
| Reduced Model B | Item 5 | Fatigue | 0.431 | -0.151 | 0.590 | No | No | No |
| Reduced Model B | Item 6 | Arithmetic money | 0.353 | 0.570 | -0.063 | No | No | No |

| Model | Item number | Item name | Communality | MR1 | MR2 | Low loadings | High cross-loadings | Low communalities |
|---|---|---|---|---|---|---|---|---|
| Reduced Model B | Item 7 | Atten-Inhib | 0.366 | 0.598 | -0.021 | No | No | No |
| Reduced Model B | Item 11 | Loneliness | 0.573 | 0.063 | 0.776 | No | No | No |
| Reduced Model B | Item 12 | Sadness | 0.619 | 0.039 | 0.799 | No | No | No |
| Reduced Model B | Item 15 | Life satisf. | 0.333 | 0.125 | -0.523 | No | No | No |
| Reduced Model B | Item 16 | Econ. diff. | 0.085 | 0.092 | 0.309 | No | No | Yes |
| Reduced Model B | Item 19 | Socio-Ludic act. | 0.566 | 0.745 | -0.022 | No | No | No |

```
fa_candidate_items_reducedB <- build_fa_candidate_items_table(
  fa_item_table = fa_item_table_reducedB,
  digits = 3
)
```

Table 42: Candidate items for elimination in FA from Reduced Model B

| Model | Item number | Item name | Low factor loadings reason | High cross-loadings reason | Low communality reason |
|---|---|---|---|---|---|
| Reduced Model B | Item 16 | Econ. diff. | No | No | Communality = 0.085 |

##### 4.8.6 Item diagnosis - EGA

```
item_stability_reducedB <- extract_item_stability(
  boot = bootega_reducedB$boot,
  model = "Reduced Model B",
  item_ids = emfra_item_ids_reducedB,
  item_labels = emfra_item_labels_reducedB,
  item_labels_short = emfra_item_labels_short_reducedB,
  stability_cutoff = 0.75
)

item_stability_table_reducedB <- item_stability_reducedB$table
item_stability_plot_reducedB <- item_stability_reducedB$plot
```

```
item_stability_plot_reducedB
```

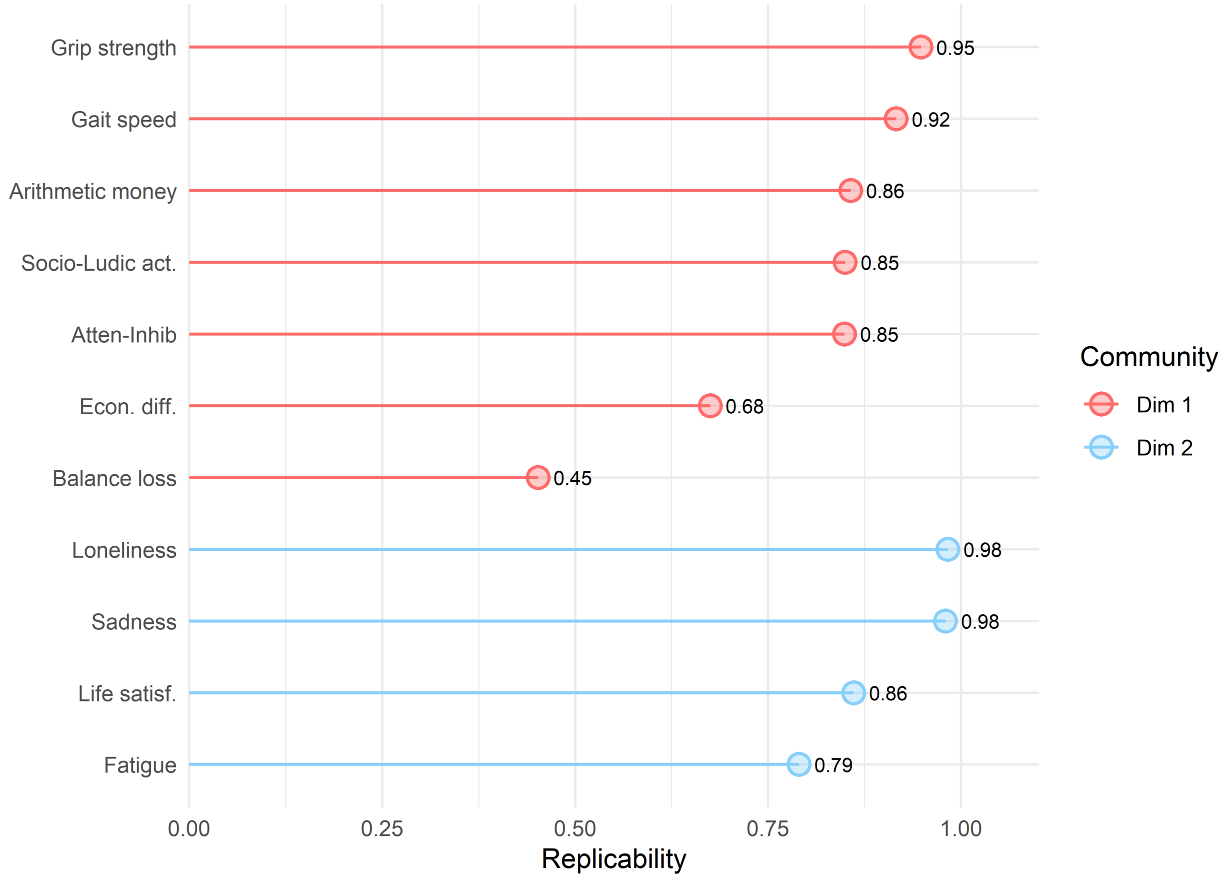

Figure 21: Item replicability from Reduced Model B

```
ega_redundancy_reducedB <- build_ega_redundancy_table(
  data = emfra_items_reducedB_ega,
  selected_obj = reduced_selected_matrix_B_ega,
  model_name = "Reduced Model B",
  uva.method = "MBR",
  cut.off = 0.25,
  reduce = FALSE,
  report_cutoff = 0.20
)

ega_redundancy_table_reducedB <- ega_redundancy_reducedB$table
```

Table 43: Item redundancy from Reduced Model B

| Model | Item 1 | Item 2 | wTO value | wTO cut-off | Redundancy level |
|---|---|---|---|---|---|
| Reduced Model B | Item 11. Loneliness | Item 12. Sadness | 0.338 | 0.3 | Large-to-very large |

| Model | Item 1 | Item 2 | wTO value | wTO cut-off | Redundancy level |
|---|---|---|---|---|---|
| Reduced Model B | Item 2. Gait speed | Item 19. Social-ludic activities | 0.286 | 0.25 | Moderate-to-large |
| Reduced Model B | Item 1. Hand-grip strength | Item 2. Gait speed | 0.259 | 0.25 | Moderate-to-large |
| Reduced Model B | Item 6. Arithmetics for money | Item 7. Attention-inhibition | 0.239 | 0.2 | Small-to-moderate |
| Reduced Model B | Item 2. Gait speed | Item 4. Balance loss | 0.221 | 0.2 | Small-to-moderate |
| Reduced Model B | Item 11. Loneliness | Item 15. Life satisfaction | 0.211 | 0.2 | Small-to-moderate |
| Reduced Model B | Item 5. Fatigue | Item 12. Sadness | 0.201 | 0.2 | Small-to-moderate |

```r
ega_item_diagnostics_reducedB <- run_item_diagnostics_selected(
  data         = emfra_items_reducedB_ega,
  selected_obj = reduced_selected_matrix_B_ega,
  model_name   = "Reduced Model B",
  iter         = 1000,
  ncores       = 1,
  seed         = 123
)

ega_item_diagnostics_table_reducedB <- ega_item_diagnostics_reducedB$table
```

```r
ega_loadings_reducedB <- extract_ega_community_loadings(
  boot = bootega_reducedB$boot,
  model = "Reduced Model B",
  item_ids = emfra_item_ids_reducedB,
  item_labels = emfra_item_labels_reducedB,
  item_labels_short = emfra_item_labels_short_reducedB,
  loading.method = "revised",
  scaling = 2,
  rotation = "oblimin",
  loading_cutoff = 0.30,
  primary_loading_cutoff = 0.50,
  digits = 3
)

ega_loadings_table_reducedB <- ega_loadings_reducedB$table
```

Table 44: Item loadings in EGA from Reduced Model B

| Model | Item number | Item name | 1 | 2 | Low loadings | High cross-loadings |
|---|---|---|---|---|---|---|
| Reduced Model B | Item 2 | Gait speed | 0.714 | 0.025 | No | No |
| Reduced Model B | Item 7 | Atten-Inhib | 0.501 | 0.018 | No | No |
| Reduced Model B | Item 6 | Arithmetic money | 0.449 | -0.016 | No | No |
| Reduced Model B | Item 1 | Grip strength | 0.411 | -0.009 | No | No |
| Reduced Model B | Item 19 | Socio-Ludic act. | 0.367 | -0.119 | No | No |
| Reduced Model B | Item 16 | Econ. diff. | -0.037 | 0.131 | Yes | No |
| Reduced Model B | Item 4 | Balance loss | -0.237 | 0.176 | Yes | No |
| Reduced Model B | Item 12 | Sadness | 0.011 | 0.608 | No | No |
| Reduced Model B | Item 11 | Loneliness | 0.041 | 0.571 | No | No |
| Reduced Model B | Item 5 | Fatigue | -0.196 | 0.314 | No | No |
| Reduced Model B | Item 15 | Life satisf. | 0.086 | -0.344 | No | No |

```r
ega_candidate_items_reducedB <- build_ega_candidate_items_table(
  item_stability_table       = item_stability_table_reducedB,
  ega_redundancy_table       = ega_redundancy_table_reducedB,
  ega_loadings_table         = ega_loadings_table_reducedB,
  ega_item_diagnostics_table = ega_item_diagnostics_table_reducedB,
  stability_cutoff           = 0.75,
  redundancy_cutoff          = 0.25,
  loading_cutoff             = 0.30,
  primary_loading_cutoff     = 0.50,
  digits                     = 3
)
```

Table 45: Candidate items for elimination in EGA from Reduced Model B

| Model | Item number | Item name | EGA candidate criterion | Low item stability reason | Redundancy reason | Item diagnostics reason | Low community loading reason | High community cross-loading reason |
|---|---|---|---|---|---|---|---|---|
| Reduced Model B | Item 1 | Grip strength | Redundancy | No | Redundant with Item 2. Gait speed (wTO = 0.259; 0.25; Moderate-to-large) | No | No | No |
| Reduced Model B | Item 2 | Gait speed | Redundancy | No | Redundant with Item 19. Social-ludic activities (wTO = 0.286; 0.25; Moderate-to-large); Redundant with Item 1. Hand-grip strength (wTO = 0.259; 0.25; Moderate-to-large) | No | No | No |
| Reduced Model B | Item 4 | Balance loss | Low replicability + Low community loading | Replicability = 45.2% (Dim 1) | No | No | Highest abs. community loading = 0.237 (Dim 1) | No |
| Reduced Model B | Item 11 | Loneliness | Redundancy | No | Redundant with Item 12. Sadness (wTO = 0.338; 0.3; Large-to-very large) | No | No | No |
| Reduced Model B | Item 12 | Sadness | Redundancy | No | Redundant with Item 11. Loneliness (wTO = 0.338; 0.3; Large-to-very large) | No | No | No |
| Reduced Model B | Item 16 | Econ. diff. | Low replicability + Low community loading | Replicability = 67.5% (Dim 1) | No | No | Highest abs. community loading = 0.131 (Dim 2) | No |
| Reduced Model B | Item 19 | Socio-Ludic act. | Redundancy | No | Redundant with Item 2. Gait speed (wTO = 0.286; 0.25; Moderate-to-large) | No | No | No |

##### 4.8.7 Item diagnosis - Retention and elimination of items

```
item_reduction_summary_reducedB <- build_item_reduction_summary_table(
  model_name = "Reduced Model B",
  fa_candidate_items_table = fa_candidate_items_reducedB,
  ega_candidate_items_table = ega_candidate_items_reducedB,
  all_item_labels = emfra_item_labels_reducedB,
  all_item_labels_short = emfra_item_labels_short_reducedB
)
```

Table 46: Item summary diagnostics from FA and EGA in Reduced Model B

| Model | Item number | Item name | FA diagnostic summary | EGA diagnostic summary |
|---|---|---|---|---|
| Reduced Model B | Item 1 | Grip strength | No FA diagnostic issue detected. | Redundancy with Item 2. Gait speed (wTO = 0.259) |
| Reduced Model B | Item 2 | Gait speed | No FA diagnostic issue detected. | Redundancy with Item 19. Social-ludic activities (wTO = 0.286); Redundancy with Item 1. Hand-grip strength (wTO = 0.259) |
| Reduced Model B | Item 4 | Balance loss | No FA diagnostic issue detected. | Low item stability: 45.2% (Dim 1); Low community loading: 0.237 (Dim 1) |
| Reduced Model B | Item 5 | Fatigue | No FA diagnostic issue detected. | No EGA diagnostic issue detected. |
| Reduced Model B | Item 6 | Arithmetic money | No FA diagnostic issue detected. | No EGA diagnostic issue detected. |
| Reduced Model B | Item 7 | Atten-Inhib | No FA diagnostic issue detected. | No EGA diagnostic issue detected. |
| Reduced Model B | Item 11 | Loneliness | No FA diagnostic issue detected. | Redundancy with Item 12. Sadness (wTO = 0.338) |
| Reduced Model B | Item 12 | Sadness | No FA diagnostic issue detected. | Redundancy with Item 11. Loneliness (wTO = 0.338) |
| Reduced Model B | Item 15 | Life satisf. | No FA diagnostic issue detected. | No EGA diagnostic issue detected. |
| Reduced Model B | Item 16 | Econ. diff. | Low communality: h2 = 0.085 | Low item stability: 67.5% (Dim 1); Low community loading: 0.131 (Dim 2) |
| Reduced Model B | Item 19 | Socio-Ludic act. | No FA diagnostic issue detected. | Redundancy with Item 2. Gait speed (wTO = 0.286) |

The item 16 (Economical difficulty) would be elimited due to its conflictive behaviour across FA and EGA. In addition, Item 4 (Balance loss) would be eliminated due to its low replicability rate in EGA.

#### 4.9 Reduced Model C

##### 4.9.1 Matrix diagnosis and selection

```
excluded_item_ids_rmc <- c(
  "EMFRA_item3_trait",   # Item 3. Gait assistance
  "EMFRA_item4_trait",   # Item 4. Balance loss
  "EMFRA_item8_trait",   # Item 8. Memory difficulty
  "EMFRA_item9_trait",   # Item 9. Concentration difficulty.
  "EMFRA_item10_trait",  # Item 10. Cognitive activities
  "EMFRA_item13_trait",  # Item 13. Irritability
  "EMFRA_item14_trait",  # Item 14. Self-efficacy difficulty
  "EMFRA_item16_trait",  # Item 16. Economical difficulty
  "EMFRA_item17_trait",  # Item 17. Family interaction
  "EMFRA_item18_trait",  # Item 18. Social interaction
  "EMFRA_item20_trait",  # Item 20. Communication difficulty
  "EMFRA_item21_trait"  # Item 21. Help availability
)

excluded_labels_rmc <- emfra_item_labels_short[
  match(excluded_item_ids_rmc, emfra_item_ids)
]

emfra_item_ids_reducedC <- emfra_item_ids[
  !emfra_item_ids %in% excluded_item_ids_rmc
]

emfra_item_labels_reducedC <- emfra_item_labels[
  match(emfra_item_ids_reducedC, emfra_item_ids)
]

emfra_item_labels_short_reducedC <- emfra_item_labels_short[
  match(emfra_item_ids_reducedC, emfra_item_ids)
]

label_map_reducedC <- setNames(
  emfra_item_labels_reducedC,
  emfra_item_labels_short_reducedC
)

emfra_items_reducedC <- data_emfra %>%
  dplyr::select(all_of(emfra_item_ids_reducedC))
```

```r
colnames(emfra_items_reducedC) <- emfra_item_labels_short_reducedC

reduced_selected_matrix_C <- select_polychoric_matrices(emfra_items_reducedC)

matrix_diagnostics_reducedC <- build_matrix_diagnostics_table(
  selected_obj = reduced_selected_matrix_C,
  model_name   = "Reduced Model C"
)
```

Table 47: Selected matrix for the Reduced Model C

| Model | Package | Correction | Selected for dim. analysis | Symmetric | PSD | Total eigenvalues (n) | Negative eigenvalues (n) | Minimum eigenvalue | Negative eigenvalues |
|---|---|---|---|---|---|---|---|---|---|
| Reduced Model C | psych | 0 | Yes | TRUE | TRUE | 9 | 0 | 0.205 | NA |
| Reduced Model C | EGAnet | 0 | Yes | TRUE | TRUE | 9 | 0 | 0.212 | NA |

##### 4.9.2 Dimensional diagnosis - Bootstrap PA and FA

```r
if (!exists("boot_pa_reducedC")) {

  boot_pa_reducedC <- run_parallel_bootstrap(
    selected_obj = reduced_selected_matrix_C,
    data         = emfra_items_reducedC,
    model_name   = "Reduced Model C"
  )

  dir.create("cache", showWarnings = FALSE)
  saveRDS(boot_pa_reducedC, ruta_cache_pa_reducedC)
}
```

Table 48: Bootstrap Parallel Analysis — Reduced Model C

| Factor | Eigenvalue | Null_mean | Null_P95 | Boot_mean | Boot_P5 | Boot_P95 | Retained - Horn's PA (P95) | Retained - Bootstrap PA (90% CI, one-sided) | Non-overlap with permuted eigenvalues (%) |
|---|---|---|---|---|---|---|---|---|---|
| 1 | 3.048 | 0.803 | 1.171 | 3.102 | 2.672 | 3.543 | Yes | Yes | 100.0 |
| 2 | 1.104 | 0.344 | 0.444 | 1.121 | 0.884 | 1.367 | Yes | Yes | 100.0 |
| 3 | 0.210 | 0.229 | 0.309 | 0.276 | 0.137 | 0.438 | No | No | 33.9 |
| 4 | -0.030 | 0.128 | 0.197 | 0.052 | -0.036 | 0.157 | No | No | 1.3 |
| 5 | -0.085 | 0.040 | 0.100 | -0.061 | -0.137 | 0.017 | No | No | 0.0 |
| 6 | -0.127 | -0.044 | 0.012 | -0.160 | -0.237 | -0.090 | No | No | 0.1 |
| 7 | -0.206 | -0.131 | -0.070 | -0.269 | -0.351 | -0.188 | No | No | 0.0 |
| 8 | -0.354 | -0.227 | -0.154 | -0.402 | -0.494 | -0.316 | No | No | 0.0 |
| 9 | -0.512 | -0.341 | -0.251 | -0.557 | -0.682 | -0.447 | No | No | 0.0 |

```r
plot_parallel_bootstrap_ggplot(boot_pa_reducedC)
```

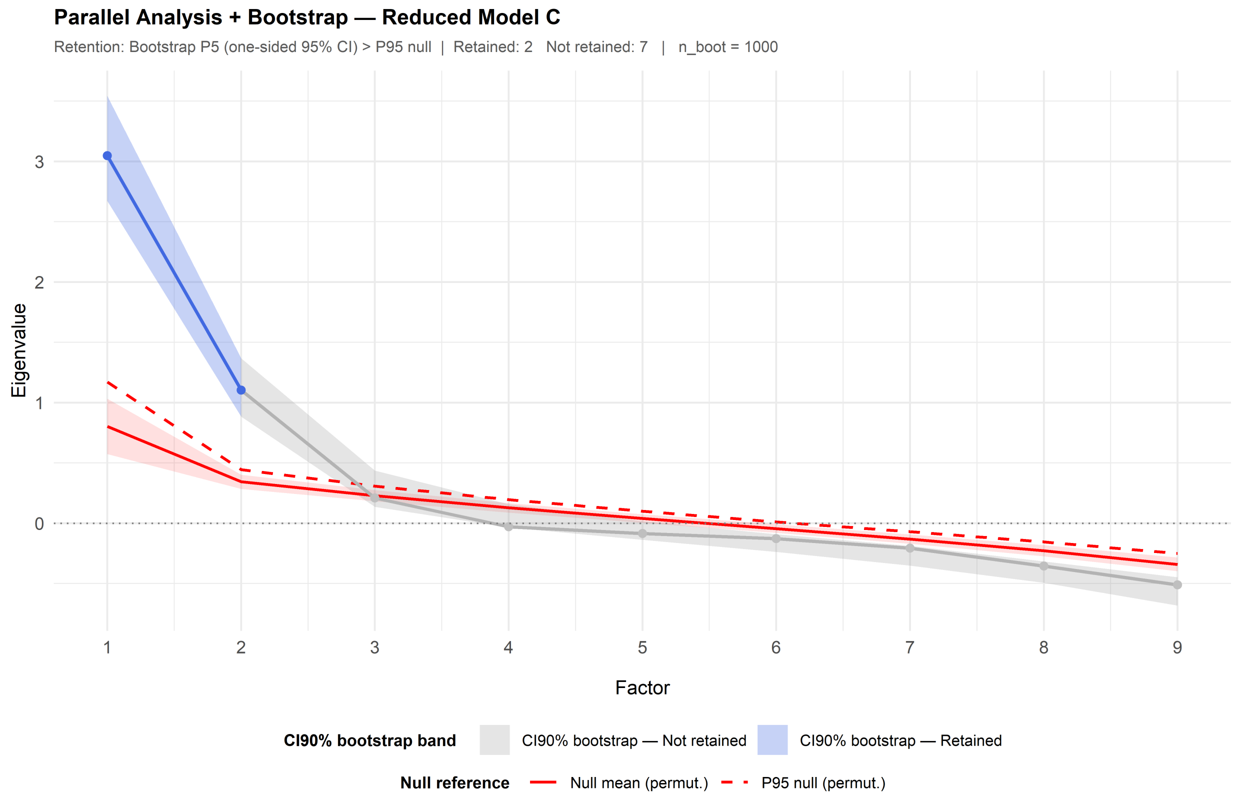

```r
fa_reducedC <- run_fa_psych_selected(
  selected_obj = reduced_selected_matrix_C,
  data = emfra_items_reducedC,
  model_name = "Reduced Model C",
  pa_obj = boot_pa_reducedC,
  fm = "minres",
  rotate = "oblimin",
  n.iter = 1000,
  SMC = TRUE,
  smooth = FALSE,
  warnings = FALSE,
  residuals = FALSE,
  scores = "tenBerge"
)

fa_fit_reducedC <- extract_fa_fit_table(
  fa_obj = fa_reducedC
)
```

```r
fa_fit_full_rmc <- dplyr::bind_rows(
  fa_fit_full,
  fa_fit_reducedA,
  fa_fit_reducedB,
  fa_fit_reducedC
 )
```

Table 49: Exploratory Factor Analysis of Reduced Model C

| Model | No. factors | Proportion Var | Chi-square | p-value | TLI | RMSEA | RMSR | Model fit | Error fit |
|---|---|---|---|---|---|---|---|---|---|
| Full Model | 3 | MR1: 0.199; MR2: 0.138; MR3: 0.075 | 992.864 | 0 | 0.446 | 0.167 | 0.074 | poor model fit | poor-to-marginal residual fit |
| Reduced Model A | 3 | MR1: 0.230; MR2: 0.161; MR3: 0.090 | 405.660 | 0 | 0.551 | 0.164 | 0.073 | poor model fit | poor-to-marginal residual fit |
| Reduced Model B | 2 | MR1: 0.260; MR2: 0.195 | 193.478 | 0 | 0.691 | 0.153 | 0.073 | poor model fit | poor-to-marginal residual fit |
| Reduced Model C | 2 | MR1: 0.297; MR2: 0.211 | 86.818 | 0 | 0.805 | 0.133 | 0.053 | poor model fit | poor-to-acceptable residual fit |

##### 4.9.3 Dimensional diagnosis - EGA

```r
emfra_items_reducedC_ega <- emfra_items_reducedC
colnames(emfra_items_reducedC_ega) <- emfra_item_labels_short[
  !emfra_item_ids %in% excluded_item_ids_rmc
]

reduced_selected_matrix_C_ega <- select_polychoric_matrices(
  data = emfra_items_reducedC_ega
)

bootega_reducedC <- run_bootega_selected(
  data         = emfra_items_reducedC_ega,
  selected_obj = reduced_selected_matrix_C_ega,
  model_name   = "Reduced Model C",
  iter         = 1000,
  seed         = 123,
  ncores       = 1,
  plot.EGA     = TRUE,
  verbose      = TRUE
)
```

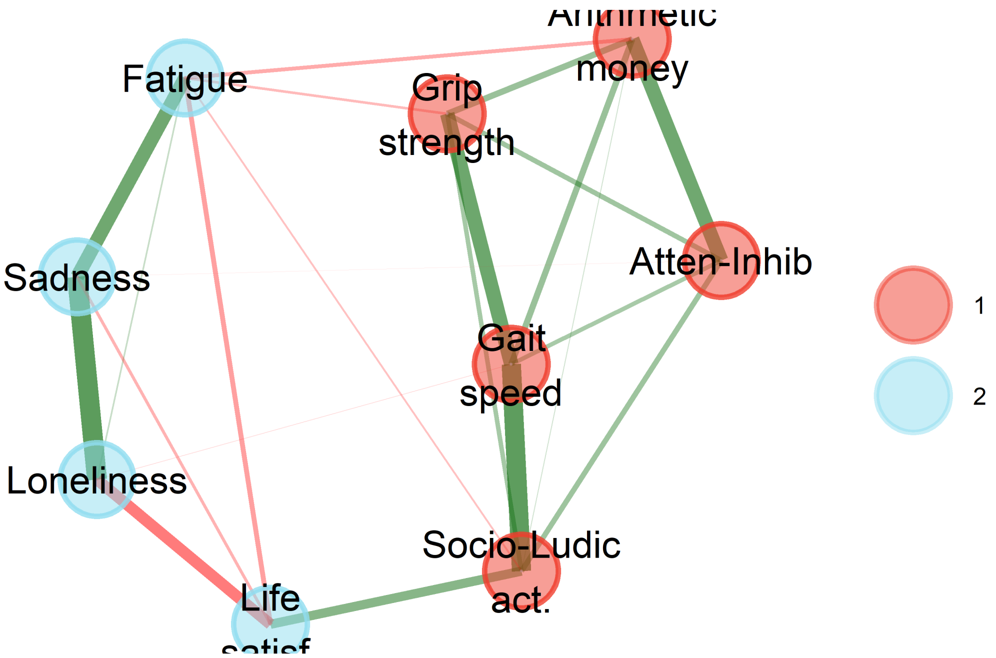

```r
ndim_freq_full_rmc <- build_ndim_frequency_table(
  boot_list = list(
    "Full Model"      = bootega_full$boot,
    "Reduced Model A" = bootega_reducedA$boot,
    "Reduced Model B" = bootega_reducedB$boot,
    "Reduced Model C" = bootega_reducedC$boot
  ),
  n_factors_range = 1:7
)

plot_ndim_frequency(
  freq_tbl  = ndim_freq_full_rmc,
  y_limit   = 100,
  facet     = TRUE
)
```

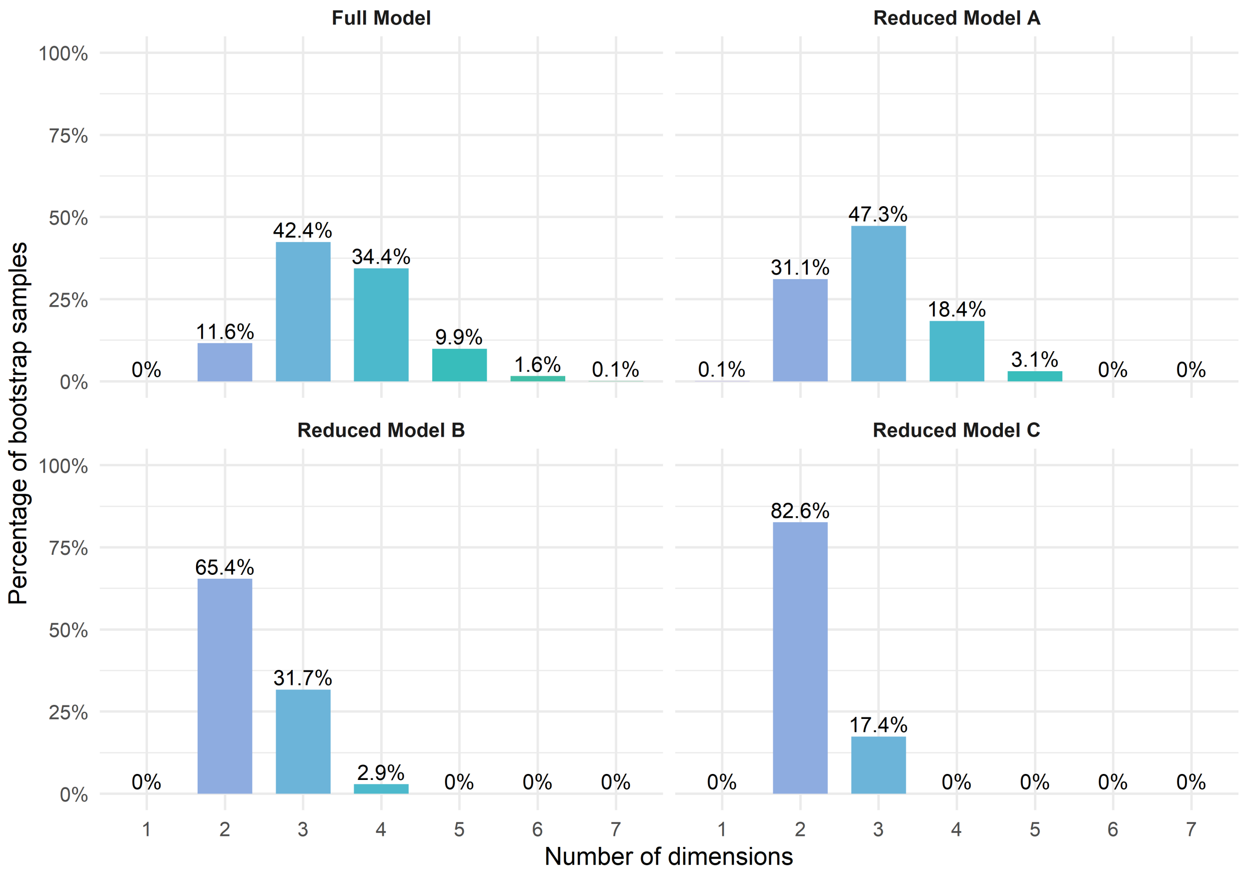

Figure 22: Frequency of the number of communities across bootstrap samples in the Full and Reduced Models A-C

```r
ndim_summary_reducedC <- summarise_ndim_frequency(
  freq_tbl    = ndim_freq_full_rmc,
  model_name = "Reduced Model C",
  top_n       = 4,
  digits      = 1
)

ndim_summary_full_rmc <- dplyr::bind_rows(
  ndim_summary_full,
  ndim_summary_reducedA,
  ndim_summary_reducedB,
  ndim_summary_reducedC
)
```

Table 50: Dimensional summary of EGA dimensions in the Full and Reduced A-C Models

| Model | Modal solution | Ranked dimensional solutions | Difference 1st-2nd | Difference 1st-3rd | Difference 1st-4th |
|---|---|---|---|---|---|
| Full Model | 3 dimensions (42.4%) | 3 dimensions: 42.4%; 4 dimensions: 34.4%; 2 dimensions: 11.6%; 5 dimensions: 9.9% | 8% | 30.8% | 32.5% |
| Reduced Model A | 3 dimensions (47.3%) | 3 dimensions: 47.3%; 2 dimensions: 31.1%; 4 dimensions: 18.4%; 5 dimensions: 3.1% | 16.2% | 28.9% | 44.2% |
| Reduced Model B | 2 dimensions (65.4%) | 2 dimensions: 65.4%; 3 dimensions: 31.7%; 4 dimensions: 2.9% | 33.7% | 62.5% | NA |
| Reduced Model C | 2 dimensions (82.6%) | 2 dimensions: 82.6%; 3 dimensions: 17.4% | 65.2% | NA | NA |

```
structural_consistency_reducedC <- extract_structural_consistency(
  boot = bootega_reducedC$boot,
  model = "Reduced Model C"
)

structural_consistency_full_rmc <- dplyr::bind_rows(
  structural_consistency_full,
  structural_consistency_reducedA,
  structural_consistency_reducedB,
  structural_consistency_reducedC
)
```

Table 51: Structural consistency of EGA dimensions in the Full and Reduced A-C Models

| Model | Dim 1 | Dim 2 | Dim 3 |
|---|---|---|---|
| Full Model | 30.8% | 19.7% | 15.1% |
| Reduced Model A | 44.5% | 16.2% | 29.9% |
| Reduced Model B | 29.9% | 70.2% | NA |
| Reduced Model C | 80.3% | 92.4% | NA |

###### 4.9.4 Dimensional diagnosis - Model acceptance criteria

```
dimensional_decision_reducedC <- build_dimensional_decision_table(
  model_name = "Reduced Model C",
  pa_obj = boot_pa_reducedC,
  fa_fit_table = fa_fit_reducedC,
  ndim_summary = ndim_summary_reducedC,
  structural_consistency = structural_consistency_reducedC,
  previous_model_comparison = "Convergence between Bootstrap PA and Bootstrap EGA
   ↪ modal solution. Best model fit and error measures compared to previous
   ↪ models. Best model performance in EGA",
```

```
  researcher_judgement = c(
    "Convergence between Bootstrap PA and Bootstrap EGA modal solution",
    "Still poor model performance, but better than previous",
    "Relevant improvement in model performance",
    "Better performance than Model B",
    "Possible retention, continue with item analyses to check the whole
    ↪  performance"
  )
)
```

Table 52: Dimensional acceptability criteria of the Reduced Model C

| Model | Criterion | Objective evidence | Researcher judgement |
|---|---|---|---|
| Reduced Model C | 1. Bootstrap PA-EGA dimensional convergence | Bootstrap PA suggested 2 factors; EGA modal solution was 2 dimensions (82.6%). | Convergence between Bootstrap PA and Bootstrap EGA modal solution |
| Reduced Model C | 2. FA dimensional performance | Proportion Var. = MR1: 0.297; MR2: 0.211; model fit = poor model fit; error fit = poor-to-acceptable residual fit. | Still poor model performance, but better than previous |
| Reduced Model C | 3. EGA dimensional stability | Modal solution = 2 dimensions (82.6%); differences: 1st-2nd = 65.2%, 1st-3rd = NA, 1st-4th = NA; structural consistency = Dim 1: 80.3%; Dim 2: 92.4%. | Relevant improvement in model performance |
| Reduced Model C | 4. Improvement over previous model | Convergence between Bootstrap PA and Bootstrap EGA modal solution. Best model fit and error measures compared to previous models. Best model performance in EGA | Better performance than Model B |
| Reduced Model C | 5. Model retention | Final model decision based on the previous dimensional criteria (1-4) | Possible retention, continue with item analyses to check the whole performance |

##### 4.9.5 Item diagnosis - PA and FA

```
fa_item_table_reducedC <- build_fa_item_table(
  fa_obj = fa_reducedC,
  model_name = "Reduced Model C",
  item_ids = emfra_item_ids_reducedC,
  item_labels = emfra_item_labels_reducedC,
  item_labels_short = emfra_item_labels_short_reducedC,
  loading_cutoff = 0.30,
  primary_loading_cutoff = 0.50,
  communality_cutoff = 0.30,
  digits = 3
)
```

Table 53: Factor analysis item diagnostics for the Reduced Model C

| Model | Item number | Item name | Communality | MR1 | MR2 | Low loadings | High cross-loadings | Low communalities |
|---|---|---|---|---|---|---|---|---|
| Reduced Model C | Item 1 | Grip strength | 0.530 | 0.739 | 0.037 | No | No | No |
| Reduced Model C | Item 2 | Gait speed | 0.691 | 0.832 | 0.001 | No | No | No |
| Reduced Model C | Item 5 | Fatigue | 0.373 | -0.181 | 0.528 | No | No | No |
| Reduced Model C | Item 6 | Arithmetic money | 0.395 | 0.620 | -0.025 | No | No | No |
| Reduced Model C | Item 7 | Atten-Inhib | 0.400 | 0.639 | 0.022 | No | No | No |
| Reduced Model C | Item 11 | Loneliness | 0.591 | 0.042 | 0.781 | No | No | No |
| Reduced Model C | Item 12 | Sadness | 0.675 | 0.039 | 0.833 | No | No | No |
| Reduced Model C | Item 15 | Life satisf. | 0.351 | 0.146 | -0.530 | No | No | No |
| Reduced Model C | Item 19 | Socio-Ludic act. | 0.565 | 0.738 | -0.041 | No | No | No |

```r
loading_cols_fa_rmc <- setdiff(
  colnames(fa_item_table_reducedC),
  c("Model", "Item number", "Item name", "Communality",
    "Low loadings", "High cross-loadings", "Low communalities")
)

plot_loadings_fa_rmc <- fa_item_table_reducedC %>%
  dplyr::select(`Item name`, dplyr::all_of(loading_cols_fa_rmc)) %>%
  tidyr::pivot_longer(
    cols = dplyr::all_of(loading_cols_fa_rmc),
    names_to = "Factor",
    values_to = "Loading"
  ) %>%
  dplyr::mutate(
    `Item name` = factor(`Item name`, levels = rev(fa_item_table_reducedC$`Item
     ↪  name`)),
    is_primary = abs(Loading) >= 0.50
  )

ggplot(plot_loadings_fa_rmc, aes(x = Factor, y = `Item name`, fill = Loading)) +
  geom_tile(color = "white", linewidth = 0.3) +
  geom_tile(
    data = dplyr::filter(plot_loadings_fa_rmc, is_primary),
    color = "black", linewidth = 0.4, fill = NA
  ) +
  geom_text(aes(label = sprintf("%.2f", Loading)), size = 2.5) +
  scale_fill_gradient2(
    low      = "blue",
```

```
    mid      = "white",
    high     = "red",
    midpoint = 0,
    limits   = c(-1, 1),
    name     = "Factor loadings"
  ) +
  labs(
    x     = "Factor",
    y     = NULL,
    title = "Factor Loadings - Reduced Model C"
  ) +
  theme_minimal(base_size = 10) +
  theme(
    panel.grid   = element_blank(),
    axis.text.y  = element_text(size = 8.5),
    plot.title   = element_text(hjust = 0.5, face = "bold", size = 10),
    legend.title = element_text(size = 9),
    legend.text  = element_text(size = 8),
  )
```

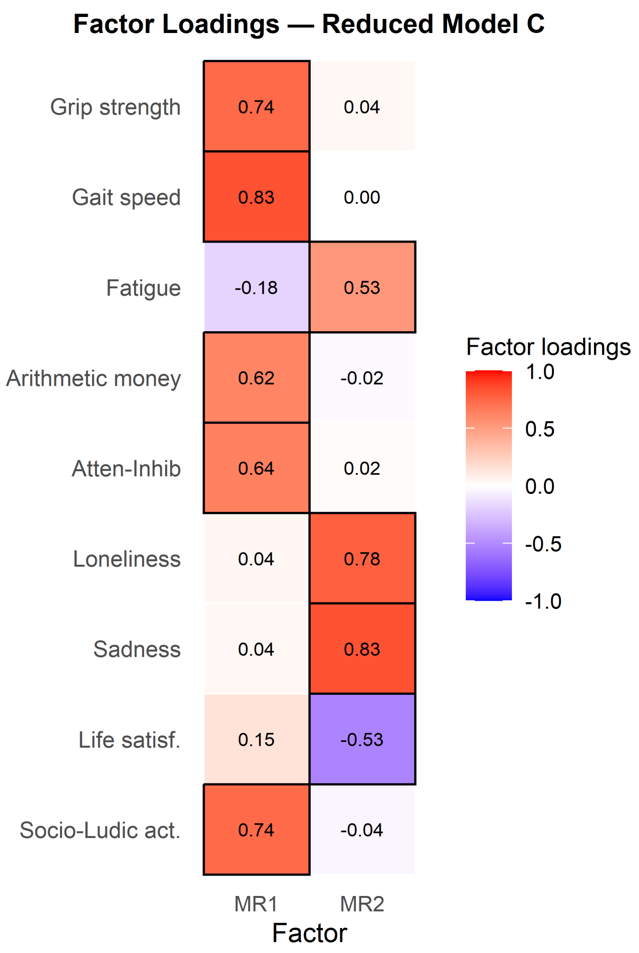

Figure 23: Factor loading heatmap for Reduced Model C. Black borders mark primary loadings ($|\lambda| \geq 0.50$).

```
fa_candidate_items_reducedC <- build_fa_candidate_items_table(
  fa_item_table = fa_item_table_reducedC,
  digits = 3
)
```

Table 54: Candidate items for elimination in FA from Reduced Model C

| Model | Item number | Item name | Low factor loadings reason | High cross-loadings reason | Low communality reason |
|---|---|---|---|---|---|

##### 4.9.6 Item diagnosis - EGA

```
item_stability_reducedC <- extract_item_stability(
  boot = bootega_reducedC$boot,
```

```r
  model = "Reduced Model C",
  item_ids = emfra_item_ids_reducedC,
  item_labels = emfra_item_labels_reducedC,
  item_labels_short = emfra_item_labels_short_reducedC,
  stability_cutoff = 0.75
)

item_stability_table_reducedC <- item_stability_reducedC$table
item_stability_plot_reducedC <- item_stability_reducedC$plot
```

```r
item_stability_plot_reducedC
```

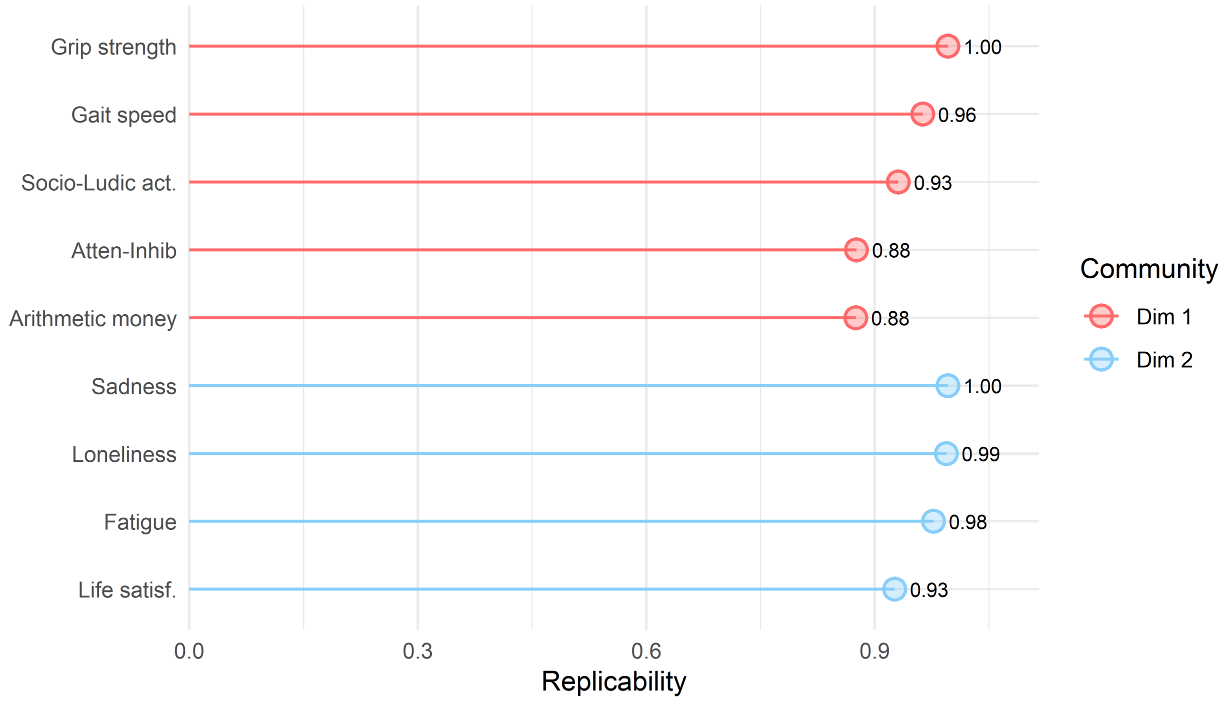

Figure 24: Item replicability from Reduced Model C

```r
ega_redundancy_reducedC <- build_ega_redundancy_table(
  data = emfra_items_reducedC_ega,
  selected_obj = reduced_selected_matrix_C_ega,
  model_name = "Reduced Model C",
  uva.method = "MBR",
  cut.off = 0.25,
  reduce = FALSE,
  report_cutoff = 0.20
)

ega_redundancy_table_reducedC <- ega_redundancy_reducedC$table
```

Table 55: Item redundancy from Reduced Model C

| Model | Item 1 | Item 2 | wTO value | wTO cut-off | Redundancy level |
|---|---|---|---|---|---|
| Reduced Model C | Item 11. Loneliness | Item 12. Sadness | 0.365 | 0.3 | Large-to-very large |
| Reduced Model C | Item 2. Gait speed | Item 19. Social-ludic activities | 0.318 | 0.3 | Large-to-very large |
| Reduced Model C | Item 1. Hand-grip strength | Item 2. Gait speed | 0.284 | 0.25 | Moderate-to-large |
| Reduced Model C | Item 6. Arithmetics for money | Item 7. Attention-inhibition | 0.259 | 0.25 | Moderate-to-large |
| Reduced Model C | Item 5. Fatigue | Item 12. Sadness | 0.251 | 0.25 | Moderate-to-large |
| Reduced Model C | Item 11. Loneliness | Item 15. Life satisfaction | 0.213 | 0.2 | Small-to-moderate |

```
ega_item_diagnostics_reducedC <- run_item_diagnostics_selected(
  data         = emfra_items_reducedC_ega,
  selected_obj = reduced_selected_matrix_C_ega,
  model_name   = "Reduced Model C",
  iter         = 1000,
  ncores       = 1,
  seed         = 123
)

ega_item_diagnostics_table_reducedC <- ega_item_diagnostics_reducedC$table
```

```
ega_loadings_reducedC <- extract_ega_community_loadings(
  boot = bootega_reducedC$boot,
  model = "Reduced Model C",
  item_ids = emfra_item_ids_reducedC,
  item_labels = emfra_item_labels_reducedC,
  item_labels_short = emfra_item_labels_short_reducedC,
  loading.method = "revised",
  scaling = 2,
  rotation = "oblimin",
  loading_cutoff = 0.30,
  primary_loading_cutoff = 0.50,
  digits = 3
)

ega_loadings_table_reducedC <- ega_loadings_reducedC$table
```

Table 56: Item loadings in EGA from Reduced Model C

| Model | Item number | Item name | 1 | 2 | Low loadings | High cross-loadings |
|---|---|---|---|---|---|---|
| Reduced Model C | Item 2 | Gait speed | 0.647 | 0.034 | No | No |
| Reduced Model C | Item 1 | Grip strength | 0.447 | -0.003 | No | No |
| Reduced Model C | Item 19 | Socio-Ludic act. | 0.437 | -0.111 | No | No |
| Reduced Model C | Item 7 | Atten-Inhib | 0.427 | 0.024 | No | No |
| Reduced Model C | Item 6 | Arithmetic money | 0.401 | -0.023 | No | No |
| Reduced Model C | Item 12 | Sadness | 0.020 | 0.627 | No | No |
| Reduced Model C | Item 11 | Loneliness | 0.012 | 0.558 | No | No |
| Reduced Model C | Item 5 | Fatigue | -0.088 | 0.345 | No | No |
| Reduced Model C | Item 15 | Life satisf. | 0.093 | -0.334 | No | No |

```r
loading_cols_ega_rmc <- setdiff(
  colnames(ega_loadings_table_reducedC),
  c("Model", "Item number", "Item name", "Low loadings", "High cross-loadings")
)

plot_loadings_ega_rmc <- ega_loadings_table_reducedC %>%
  dplyr::select(`Item name`, dplyr::all_of(loading_cols_ega_rmc)) %>%
  tidyr::pivot_longer(
    cols = dplyr::all_of(loading_cols_ega_rmc),
    names_to = "Community",
    values_to = "Loading"
  ) %>%
  dplyr::mutate(
    `Item name` = factor(`Item name`, levels =
     ↪ rev(ega_loadings_table_reducedC$`Item name`)),
    is_primary = abs(Loading) >= 0.50
  )

ggplot(plot_loadings_ega_rmc, aes(x = Community, y = `Item name`, fill = Loading))
 ↪ +
  geom_tile(color = "white", linewidth = 0.3) +
  geom_tile(
    data = dplyr::filter(plot_loadings_ega_rmc, is_primary),
    color = "black", linewidth = 0.4, fill = NA
```

```
) +
geom_text(aes(label = sprintf("%.2f", Loading)), size = 2.5) +
scale_fill_gradient2(
  low      = "blue",
  mid      = "white",
  high     = "red",
  midpoint = 0,
  limits   = c(-1, 1),
  name     = "Community loading"
) +
labs(
  x     = "Community",
  y     = NULL,
  title = "Community Loadings - Reduced Model C"
) +
theme_minimal(base_size = 10) +
theme(
  panel.grid   = element_blank(),
  axis.text.y  = element_text(size = 8.5),
  plot.title   = element_text(hjust = 0.5, face = "bold", size = 10),
  legend.title = element_text(size = 9),
  legend.text  = element_text(size = 8),
)
```

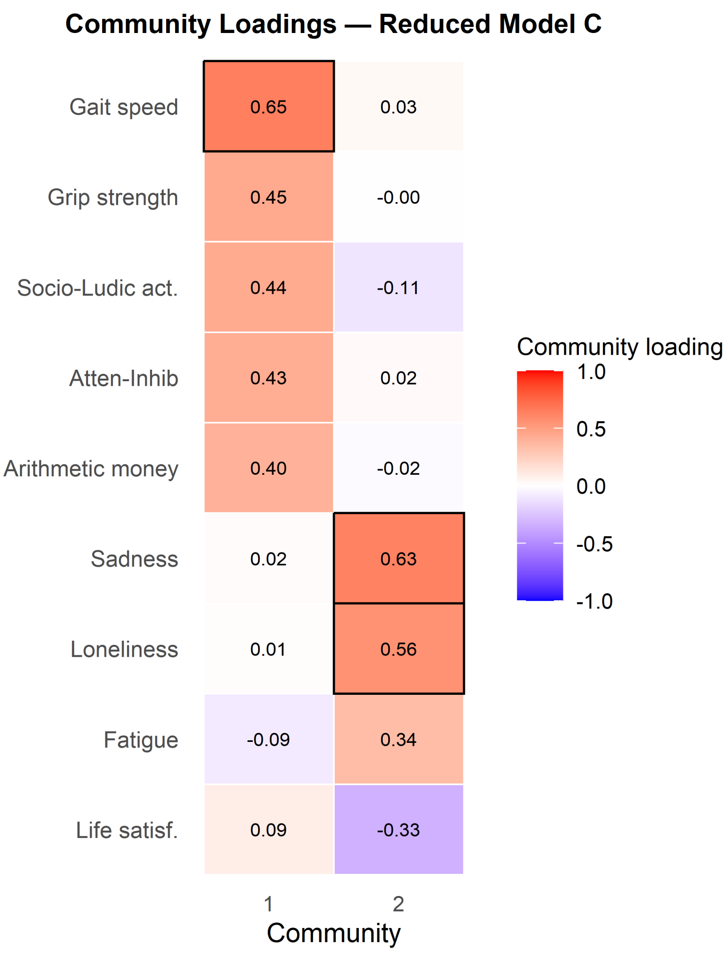

Figure 25: Community loading (CL) heatmap for Reduced Model C (EGA). Black borders mark primary loadings ($CL \geq 0.50$).

```
ega_candidate_items_reducedC <- build_ega_candidate_items_table(
  item_stability_table        = item_stability_table_reducedC,
  ega_redundancy_table        = ega_redundancy_table_reducedC,
  ega_loadings_table          = ega_loadings_table_reducedC,
  ega_item_diagnostics_table = ega_item_diagnostics_table_reducedC,
  stability_cutoff            = 0.75,
  redundancy_cutoff           = 0.25,
  loading_cutoff              = 0.30,
  primary_loading_cutoff      = 0.50,
  digits                      = 3
)
```

Table 57: Candidate items for elimination in EGA from Reduced Model C

| Model | Item number | Item name | EGA candidate criterion | Low item stability reason | Redundancy reason | Item diagnostics reason | Low community loading reason | High community cross-loading reason |
|---|---|---|---|---|---|---|---|---|
| Reduced Model C | Item 1 | Grip strength | Redundancy | No | Redundant with Item 2. Gait speed (wTO = 0.284; 0.25; Moderate-to-large) | No | No | No |
| Reduced Model C | Item 2 | Gait speed | Redundancy | No | Redundant with Item 19. Social-ludic activities (wTO = 0.318; 0.3; Large-to-very large); Redundant with Item 1. Hand-grip strength (wTO = 0.284; 0.25; Moderate-to-large) | No | No | No |
| Reduced Model C | Item 5 | Fatigue | Redundancy | No | Redundant with Item 12. Sadness (wTO = 0.251; 0.25; Moderate-to-large) | No | No | No |
| Reduced Model C | Item 6 | Arithmetic money | Redundancy | No | Redundant with Item 7. Attention-inhibition (wTO = 0.259; 0.25; Moderate-to-large) | No | No | No |
| Reduced Model C | Item 7 | Atten-Inhib | Redundancy | No | Redundant with Item 6. Arithmetics for money (wTO = 0.259; 0.25; Moderate-to-large) | No | No | No |
| Reduced Model C | Item 11 | Loneliness | Redundancy | No | Redundant with Item 12. Sadness (wTO = 0.365; 0.3; Large-to-very large) | No | No | No |
| Reduced Model C | Item 12 | Sadness | Redundancy | No | Redundant with Item 11. Loneliness (wTO = 0.365; 0.3; Large-to-very large); Redundant with Item 5. Fatigue (wTO = 0.251; 0.25; Moderate-to-large) | No | No | No |
| Reduced Model C | Item 19 | Socio-Ludic act. | Redundancy | No | Redundant with Item 2. Gait speed (wTO = 0.318; 0.3; Large-to-very large) | No | No | No |

##### 4.9.7 Item diagnosis - Retention and elimination of items

```
item_reduction_summary_reducedC <- build_item_reduction_summary_table(
  model_name = "Reduced Model C",
  fa_candidate_items_table = fa_candidate_items_reducedC,
  ega_candidate_items_table = ega_candidate_items_reducedC,
  all_item_labels = emfra_item_labels_reducedC,
  all_item_labels_short = emfra_item_labels_short_reducedC
)
```

Table 58: Item summary diagnostics from FA and EGA in Reduced Model C

| Model | Item number | Item name | FA diagnostic summary | EGA diagnostic summary |
|---|---|---|---|---|
| Reduced Model C | Item 1 | Grip strength | No FA diagnostic issue detected. | Redundancy with Item 2. Gait speed (wTO = 0.284) |
| Reduced Model C | Item 2 | Gait speed | No FA diagnostic issue detected. | Redundancy with Item 19. Social-ludic activities (wTO = 0.318); Redundancy with Item 1. Hand-grip strength (wTO = 0.284) |
| Reduced Model C | Item 5 | Fatigue | No FA diagnostic issue detected. | Redundancy with Item 12. Sadness (wTO = 0.251) |
| Reduced Model C | Item 6 | Arithmetic money | No FA diagnostic issue detected. | Redundancy with Item 7. Attention-inhibition (wTO = 0.259) |
| Reduced Model C | Item 7 | Atten-Inhib | No FA diagnostic issue detected. | Redundancy with Item 6. Arithmetics for money (wTO = 0.259) |
| Reduced Model C | Item 11 | Loneliness | No FA diagnostic issue detected. | Redundancy with Item 12. Sadness (wTO = 0.365) |
| Reduced Model C | Item 12 | Sadness | No FA diagnostic issue detected. | Redundancy with Item 11. Loneliness (wTO = 0.365); Redundancy with Item 5. Fatigue (wTO = 0.251) |
| Reduced Model C | Item 15 | Life satisf. | No FA diagnostic issue detected. | No EGA diagnostic issue detected. |
| Reduced Model C | Item 19 | Socio-Ludic act. | No FA diagnostic issue detected. | Redundancy with Item 2. Gait speed (wTO = 0.318) |

### 5 Summary FA features of the final model

#### 5.1 FA-based summary

```
fa_fit_rmc_obj <- if (!is.null(fa_reducedC$fa)) fa_reducedC$fa else fa_reducedC
phi_mat_rmc    <- fa_fit_rmc_obj$Phi

phi_text_rmc <- if (!is.null(phi_mat_rmc) && is.matrix(phi_mat_rmc) &&
↪  nrow(phi_mat_rmc) >= 2) {
  fn    <- colnames(phi_mat_rmc)
  if (is.null(fn)) fn <- paste0("F", seq_len(nrow(phi_mat_rmc)))
  pairs <- combn(nrow(phi_mat_rmc), 2)
  paste(
    apply(pairs, 2, function(idx) {
      paste0(fn[idx[1]], "-", fn[idx[2]], ": ",
             sprintf("%.3f", phi_mat_rmc[idx[1], idx[2]]))
    }),
    collapse = "; "
  )
} else {
  NA_character_
}

fa_fit_rmc_display <- fa_fit_reducedC %>%
  dplyr::mutate(`Interfactor correlation` = phi_text_rmc) %>%
  dplyr::relocate(`Interfactor correlation`, .after = `Proportion Var`)
```

Table 59: FA model fit indices and interfactor correlation for Reduced Model C (psych, minres, oblimin)

| Model | No. factors | Proportion Var | Interfactor correlation | Chi-square | p-value | TLI | RMSEA | RMSR | Model fit | Error fit |
|---|---|---|---|---|---|---|---|---|---|---|
| Reduced Model C | 2 | MR1: 0.297; MR2: 0.211 | MR1-MR2: -0.316 | 86.818 | 0 | 0.805 | 0.133 | 0.053 | poor model fit | poor-to-acceptable residual fit |

```
non_display_cols_rmc <- c("Model", "Low loadings", "High cross-loadings", "Low
↪  communalities")
loading_cols_fa_rmc_s   <- setdiff(
  colnames(fa_item_table_reducedC),
  c(non_display_cols_rmc, "Item number", "Item name", "Communality")
)

fa_summary_rmc <- fa_item_table_reducedC %>%
  dplyr::select(
    `Item number`, `Item name`,
```

```r
    dplyr::all_of(loading_cols_fa_rmc_s),
    Communality
  ) %>%
  dplyr::rename(h2 = Communality) %>%
  dplyr::mutate(u2 = round(1 - h2, 3))
```

Table 60: Factor loadings, communality (h2) and uniqueness (u2) for Reduced Model C

| Item number | Item name | MR1 | MR2 | h2 | u2 |
|---|---|---|---|---|---|
| Item 1 | Grip strength | 0.739 | 0.037 | 0.530 | 0.470 |
| Item 2 | Gait speed | 0.832 | 0.001 | 0.691 | 0.309 |
| Item 5 | Fatigue | -0.181 | 0.528 | 0.373 | 0.627 |
| Item 6 | Arithmetic money | 0.620 | -0.025 | 0.395 | 0.605 |
| Item 7 | Atten-Inhib | 0.639 | 0.022 | 0.400 | 0.600 |
| Item 11 | Loneliness | 0.042 | 0.781 | 0.591 | 0.409 |
| Item 12 | Sadness | 0.039 | 0.833 | 0.675 | 0.325 |
| Item 15 | Life satisf. | 0.146 | -0.530 | 0.351 | 0.649 |
| Item 19 | Socio-Ludic act. | 0.738 | -0.041 | 0.565 | 0.435 |

```r
plot_fa_loadings(fa_item_table_reducedC)
```

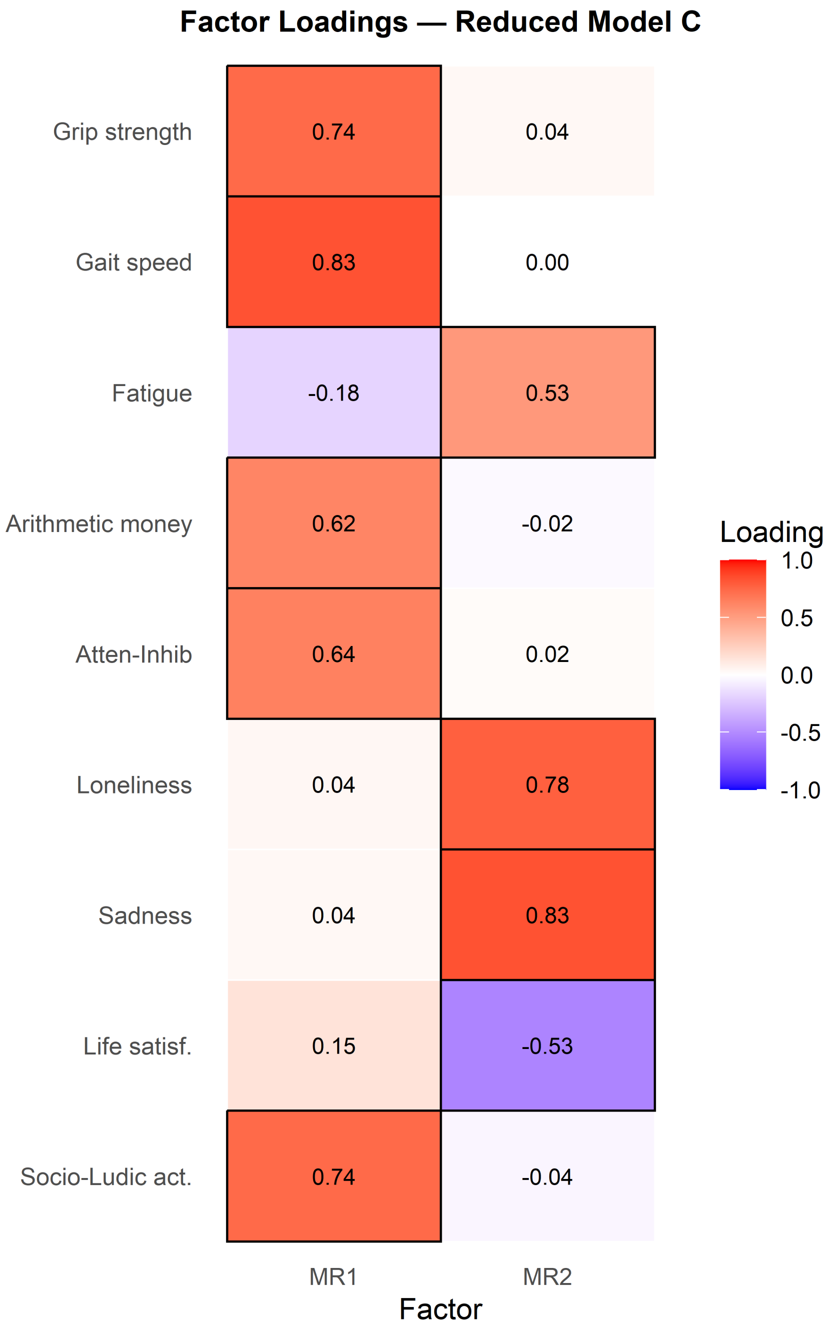

Figure 26: Factor loading heatmap for Reduced Model C (psych FA, oblimin). Black borders mark loadings ($|\lambda| \geq 0.50$).

#### 5.2 Internal consistency

```
fa_rmc_L   <- unclass(fa_reducedC$fa$loadings)
fa_rmc_Phi <- fa_reducedC$fa$Phi
```

```
omega_rmc <- psych::omegah(
  m         = fa_rmc_L,
  Phi       = fa_rmc_Phi,
  nfactors = fa_reducedC$nfactors,
  two.ok    = TRUE,
  plot      = FALSE
)
```

```
non_load_rmc <- c("Model", "Item number", "Item name", "Communality",
                  "Low loadings", "High cross-loadings", "Low communalities")
load_cols_fa <- setdiff(colnames(fa_item_table_reducedC), non_load_rmc)

item_assign_rmc <- fa_item_table_reducedC %>%
  dplyr::select(`Item name`, dplyr::all_of(load_cols_fa)) %>%
  dplyr::rowwise() %>%
  dplyr::mutate(
    .vals       = list(dplyr::c_across(dplyr::all_of(load_cols_fa))),
    .pri        = which.max(abs(unlist(.vals))),
    primary_fac = load_cols_fa[.pri],
    primary_sgn = sign(unlist(.vals)[.pri])
  ) %>%
  dplyr::ungroup() %>%
  dplyr::select(`Item name`, primary_fac, primary_sgn)

correction_rmc <- reduced_selected_matrix_C$correction_used

omega_sub_list <- purrr::map(load_cols_fa, function(f) {
  sub    <- dplyr::filter(item_assign_rmc, primary_fac == f)
  df_sub <- emfra_items_reducedC %>%
    dplyr::select(dplyr::all_of(sub$`Item name`))

  Rpoly_preferred <- compute_polychoric_psych(df_sub, correct = correction_rmc)
  diag_preferred  <- check_correlation_matrix(Rpoly_preferred)
  ok_preferred    <- isTRUE(diag_preferred$Symmetric) &&
 ↪  isTRUE(diag_preferred$PSD)

  Rpoly_sub <- if (ok_preferred) {
    Rpoly_preferred
  } else {
    warning(paste0(
      "Subscale ", f, ": correct = ", correction_rmc,
      " did not yield a symmetric/PSD matrix. Falling back to
       ↪  select_polychoric_matrices()."
    ))
    select_polychoric_matrices(df_sub)$psych$Rpoly
  }
```

```r
  om_sub <- psych::omega(
    Rpoly_sub,
    nfactors = 1,
    key      = sub$primary_sgn,
    n.obs    = nrow(df_sub),
    plot     = FALSE
  )
  tibble::tibble(
    Indicator = paste0("  subscale - ", f, " (", nrow(sub), " main loading items
     ↪  only)"),
    Value     = round(om_sub$omega.tot, 3)
  )
})

omega_subscale_rmc <- dplyr::bind_rows(omega_sub_list)
```

```r
omega_reliability_rmc <- dplyr::bind_rows(
  tibble::tibble(
    Indicator = c(
      " t - Omega total (full scale)",
      " h - Omega hierarchical",
      "ECV - Explained Common Variance"
    ),
    Value = round(c(
      omega_rmc$omega.tot,
      omega_rmc$omega_h,
      omega_rmc$ECV
    ), 3)
  ),
  omega_subscale_rmc
)
```

Table 61: Scale reliability and explained variance indices for Reduced Model C

| Indicator | Value |
|---|---|
| t — Omega total (full scale) | 0.864 |
| h — Omega hierarchical | 0.409 |
| ECV — Explained Common Variance | 0.327 |
| subscale — MR1 (5 main loading items only) | 0.840 |
| subscale — MR2 (4 main loading items only) | 0.785 |

```r
omega_group_rmc <- as.data.frame(round(omega_rmc$omega.group, 3)) %>%
  tibble::rownames_to_column("Component") %>%
  dplyr::rename(
    ` total`   = total,
```

```r
    `general` = general,
    `group`   = group
  )
```

Table 62: Omega decomposition by component for Reduced Model C. total: total reliability; general: variance due to g; group: variance due to the specific group factor.

| Component | total | general | group |
|---|---|---|---|
| g | 0.864 | 0.409 | 0.416 |
| F1* | 0.841 | 0.267 | 0.574 |
| F2* | 0.787 | 0.280 | 0.507 |

```r
sl_rmc <- as.data.frame(round(omega_rmc$schmid$sl, 3)) %>%
  tibble::rownames_to_column("Item") %>%
  dplyr::select(Item, g, dplyr::matches("^F"), h2, u2, p2)
```

Table 63: Schmid-Leiman orthogonalised solution for Reduced Model C. g: general factor loading; F1/*F2*: residual group factor loadings after removing g; h2: communality; u2: uniqueness; p2: proportion of item variance due to g.

| Item | g | F1* | F2* | h2 | u2 | p2 |
|---|---|---|---|---|---|---|
| Grip strength | 0.395 | 0.611 | 0.030 | 0.530 | 0.470 | 0.294 |
| Gait speed | 0.467 | 0.688 | 0.001 | 0.691 | 0.309 | 0.316 |
| Fatigue- | 0.399 | 0.150 | -0.437 | 0.373 | 0.627 | 0.427 |
| Arithmetic money | 0.363 | 0.513 | -0.021 | 0.395 | 0.605 | 0.333 |
| Atten-Inhib | 0.347 | 0.529 | 0.018 | 0.400 | 0.600 | 0.301 |
| Loneliness- | 0.415 | -0.035 | -0.646 | 0.591 | 0.409 | 0.292 |
| Sadness- | 0.447 | -0.032 | -0.689 | 0.675 | 0.325 | 0.296 |
| Life satisf. | 0.380 | 0.121 | -0.438 | 0.351 | 0.649 | 0.412 |
| Socio-Ludic act. | 0.438 | 0.610 | -0.034 | 0.565 | 0.435 | 0.340 |

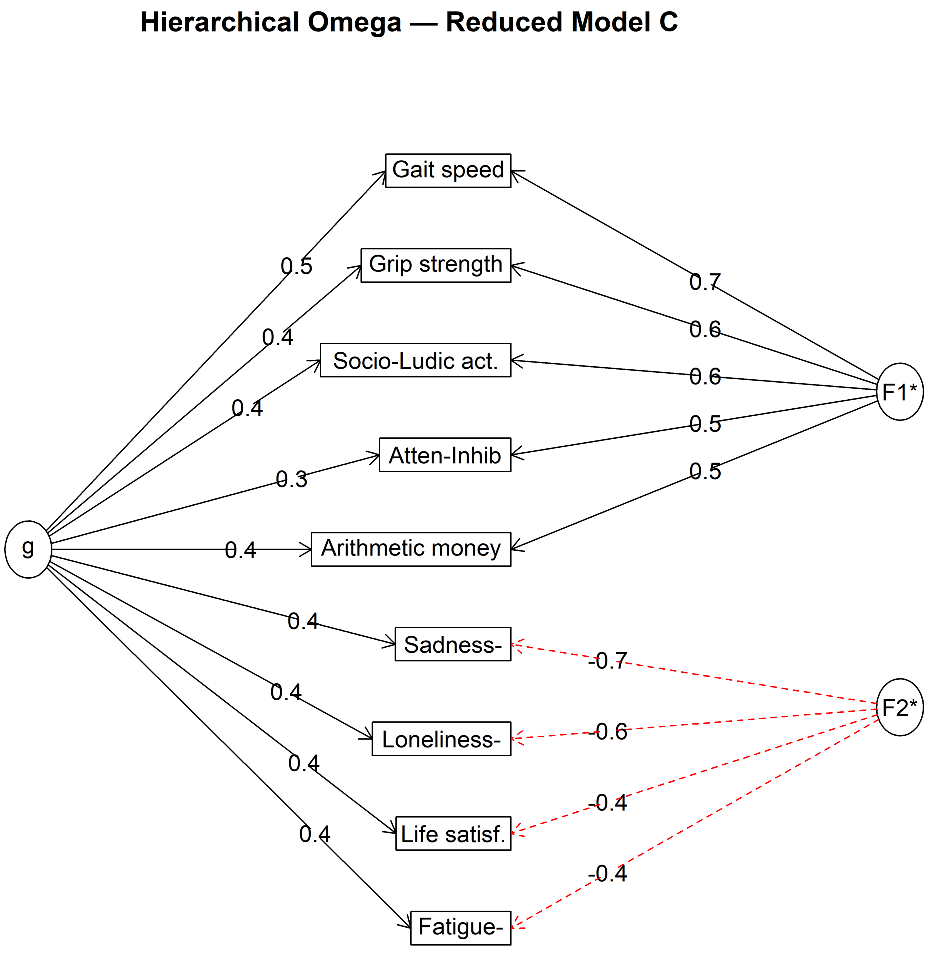

Figure 27: Hierarchical omega path diagram for Reduced Model C (Schmid-Leiman solution). g: general factor shared by all items; F1, *F2*: orthogonalised group factors after extracting g. Arrow widths are proportional to loadings.

#### 5.3 Factor score weights

```
fa_weights_rmc <- as.data.frame(round(fa_reducedC$fa$weights, 3)) %>%
  tibble::rownames_to_column("Item name")
```

Table 64: Factor score regression weights for Reduced Model C (tenBerge method).

| Item name | MR1 | MR2 |
|---|---|---|
| Grip strength | 0.233 | -0.011 |
| Gait speed | 0.405 | 0.034 |
| Fatigue | -0.054 | 0.157 |
| Arithmetic money | 0.172 | -0.009 |
| Atten-Inhib | 0.179 | 0.032 |

Table 64: Factor score regression weights for Reduced Model C (tenBerge method).

| Item name | MR1 | MR2 |
|---|---|---|
| Loneliness | 0.024 | 0.388 |
| Sadness | 0.009 | 0.510 |
| Life satisf. | 0.038 | -0.157 |
| Socio-Ludic act. | 0.234 | -0.068 |

```
acronym_rmc <- c(
  "Gait speed"        = "GS",
  "Grip strength"     = "HGS",
  "Socio-Ludic act." = "SLA",
  "Atten-Inhib"       = "AttIn",
  "Arithmetic money" = "AM",
  "Sadness"           = "SAD",
  "Loneliness"        = "LON",
  "Life satisf."      = "LS",
  "Fatigue"           = "FAT"
)

item_order_rmc <- names(acronym_rmc)

fa_weights_rmc_formula <- fa_weights_rmc %>%
  dplyr::filter(`Item name` %in% item_order_rmc) %>%
  dplyr::slice(match(item_order_rmc, `Item name`))

acronyms_rmc <- unname(acronym_rmc[fa_weights_rmc_formula$`Item name`])
factor_cols  <- setdiff(colnames(fa_weights_rmc_formula), "Item name")

reverse_by_factor <- lapply(factor_cols, function(f) {
  fa_weights_rmc_formula$`Item name`[fa_weights_rmc_formula[[f]] < 0]
})
names(reverse_by_factor) <- factor_cols

formulas_rmc <- lapply(seq_along(factor_cols), function(f_idx) {
  f_col     <- factor_cols[f_idx]
  coefs     <- round(abs(fa_weights_rmc_formula[[f_col]]), 3)
  items     <- fa_weights_rmc_formula$`Item name`
  rev_items <- reverse_by_factor[[f_col]]

  terms <- vapply(seq_along(coefs), function(i) {
    var_text <- if (items[i] %in% rev_items) {
      paste0(acronyms_rmc[i], "^{*}")
    } else {
      acronyms_rmc[i]
    }
    if (i == 1) sprintf("%.3f\\,%s", coefs[i], var_text)
```

```
    else         sprintf("\\; +\\; %.3f\\,%s", coefs[i], var_text)
  }, character(1))

  max_terms_per_line <- 4
  term_groups    <- split(terms, ceiling(seq_along(terms) / max_terms_per_line))
  line_strings   <- vapply(term_groups, paste, character(1), collapse = "")
  formula_body   <- paste(line_strings, collapse = " \\\\\n")

  paste0("\\begin{multline*}\n\\hat{F}_{", f_idx, "} = ", formula_body,
   ↪  "\n\\end{multline*}")
})

legend_lines <- vapply(seq_along(acronyms_rmc), function(i) {
  sprintf("%s = %s", acronyms_rmc[i], fa_weights_rmc_formula$`Item name`[i])
}, character(1))

legend_text <- paste0(
  "\n\n*Where:* ", paste(legend_lines, collapse = "; "), ".\n\n",
  "Indicators marked with $^{*}$ denote reverse-coded scores computed as ",
  "$X_i^{*} = 2 - X_i$ prior to factor score estimation."
)

knitr::asis_output(paste0(paste(formulas_rmc, collapse = "\n\n"), legend_text))
```

$$\hat{F}_1 = 0.405\,GS \;+\; 0.233\,HGS \;+\; 0.234\,SLA \;+\; 0.179\,AttIn \\ +\; 0.172\,AM \;+\; 0.009\,SAD \;+\; 0.024\,LON \;+\; 0.038\,LS \\ +\; 0.054\,FAT^{*}$$

$$\hat{F}_2 = 0.034\,GS \;+\; 0.011\,HGS^{*} \;+\; 0.068\,SLA^{*} \;+\; 0.032\,AttIn \\ +\; 0.009\,AM^{*} \;+\; 0.510\,SAD \;+\; 0.388\,LON \;+\; 0.157\,LS^{*} \\ +\; 0.157\,FAT$$

*Where:* GS = Gait speed; HGS = Grip strength; SLA = Socio-Ludic act.; AttIn = Atten-Inhib; AM = Arithmetic money; SAD = Sadness; LON = Loneliness; LS = Life satisf.; FAT = Fatigue.

Indicators marked with $^{*}$ denote reverse-coded scores computed as $X_i^{*} = 2 - X_i$ prior to factor score estimation.

#### 5.4 Factor naming

Based on the EGA structure of Reduced Model 5, and the respective correlation (loadings) between the items and their respective factor, we consider the following names for the latent factors:

*Factor 1: Autonomy.* Based on the contribution of physical functions (hand-grip strength, and gait speed), along with arithmetic for money, a process that represents both working memory performance, a part of instrumented daily activity performance, and outdoor social tasks. In addition, there is a contribution of the frequency of socio-ludic activities. In overall, we consider that all those components are indirect mesaures of a latent variable referred as "autonomy".

*Factor 2: Psychological distress.* The composition of this factor through the detected items aligns with the defining attributes identified in by Rinder 2004 concept analysis. The strongest loading corresponds to negatively weighted Life Satisfaction, indicating a state of discomfort, a core component of "psychological distress" (Rinder 2004). In addition, Loneliness and Irritability reflect emotional change and psychological discomfort, two core attributes described in the literature (Rinder 2004). The item Self-Efficacy Difficulty directly corresponds to the defining attribute of perceived inability to cope effectively, a central component of psychological distress (Rinder 2004). Although Concentration Difficulty represents a cognitive complaint, it is frequently reported as a manifestation of distress-related interference and therefore remains conceptually coherent within this domain.

### 6 Factor descriptives

Factor scores for Reduced Model C are computed separately for each factor using the absolute values of the tenBerge regression weights. Items whose weight is negative in a given factor are **reverse-coded** prior to scoring using the transformation $X_i^* = 2 - X_i$, so that higher values consistently reflect greater intensity of the latent construct. The reversal set is factor-specific: an item may be used directly for one factor and reversed for the other. After reversal, all weights enter the scoring formula as positive values:

$$\hat{F}_j = \sum_{i \in \mathcal{P}_j} |w_{ij}| \, X_i \; + \sum_{i \in \mathcal{N}_j} |w_{ij}| \, X_i^*$$

where $\mathcal{P}_j$ denotes items with positive weight and $\mathcal{N}_j$ denotes items with negative weight for factor $j$, and $X_i^* = 2 - X_i$. Under this parameterisation the minimum possible score is always 0 (all items at their minimum) and the maximum is $2\sum_i |w_{ij}|$ (all items at their maximum):

$$\hat{F}_{j,\min} = 0, \qquad \hat{F}_{j,\max} = 2\sum_{i=1}^{k} |w_{ij}|$$

```
factor_cols_w    <- setdiff(colnames(fa_weights_rmc_formula), "Item name")
factor_label_map <- setNames(
  c("Autonomy", "Psychological distress"),
  factor_cols_w
)

fa_minmax_rmc <- purrr::map_dfr(factor_cols_w, function(f) {
  w <- fa_weights_rmc_formula[[f]]
  tibble::tibble(
    Factor                  = f,
    `Factor name`           = factor_label_map[[f]],
```

```
    `Minimum possible score` = 0,
    `Maximum possible score` = round(2 * sum(abs(w)), 3)
  )
})
```

Table 65: Theoretical minimum and maximum factor scores for Reduced Model C. Scores are anchored at 0 and computed using absolute tenBerge weights after factor-specific item reversal.

| Factor | Factor name | Minimum possible score | Maximum possible score |
|---|---|---|---|
| MR1 | Autonomy | 0 | 2.696 |
| MR2 | Psychological distress | 0 | 2.732 |

The following descriptive statistics are indicated for both Factors.

```
item_order_rmc <- fa_weights_rmc$`Item name`

item_col_map_emfra <- setNames(
  emfra_item_ids,
  emfra_item_labels_short
)[item_order_rmc]

emfra_weights_f1 <- setNames(
  lapply(item_order_rmc, function(lbl) {
    list(
      weight = round(abs(fa_weights_rmc$MR1[fa_weights_rmc$`Item name` == lbl]),
       ↪  3)
    )
  }),
  item_order_rmc
)

emfra_weights_f2 <- setNames(
  lapply(item_order_rmc, function(lbl) {
    list(
      weight = round(abs(fa_weights_rmc$MR2[fa_weights_rmc$`Item name` == lbl]),
       ↪  3)
    )
  }),
  item_order_rmc
)

data_emfra <- data_emfra %>%
  mutate(
    Factor_1_Autonomy =
```

```r
      emfra_weights_f1$`Grip strength`$weight    * EMFRA_item1_trait         +   #
↪  HGS  - Grip strength
      emfra_weights_f1$`Gait speed`$weight       * EMFRA_item2_trait         +   #
↪  GS   - Gait speed
      emfra_weights_f1$Fatigue$weight            * EMFRA_item5_trait_reverse +   #
↪  FAT  - Fatigue (negative → reverse)
      emfra_weights_f1$`Arithmetic money`$weight * EMFRA_item6_trait         +   #
↪  AM   - Arithmetic money
      emfra_weights_f1$`Atten-Inhib`$weight      * EMFRA_item7_trait         +   #
↪  AttIn - Atten-Inhib
      emfra_weights_f1$Loneliness$weight         * EMFRA_item11_trait        +   #
↪  LON  - Loneliness
      emfra_weights_f1$Sadness$weight            * EMFRA_item12_trait        +   #
↪  SAD  - Sadness
      emfra_weights_f1$`Life satisf.`$weight     * EMFRA_item15_trait        +   #
↪  LS   - Life satisf.
      emfra_weights_f1$`Socio-Ludic act.`$weight * EMFRA_item19_trait            #
↪  SLA  - Socio-Ludic act.
  ) %>%
  mutate(
    Factor_2_Psych_Distress =
      emfra_weights_f2$`Grip strength`$weight    * EMFRA_item1_trait_reverse +   #
↪  HGS  - Grip strength (negative → reverse)
      emfra_weights_f2$`Gait speed`$weight       * EMFRA_item2_trait         +   #
↪  GS   - Gait speed
      emfra_weights_f2$Fatigue$weight            * EMFRA_item5_trait         +   #
↪  FAT  - Fatigue
      emfra_weights_f2$`Arithmetic money`$weight * EMFRA_item6_trait_reverse +   #
↪  AM   - Arithmetic money (negative → reverse)
      emfra_weights_f2$`Atten-Inhib`$weight      * EMFRA_item7_trait         +   #
↪  AttIn - Atten-Inhib
      emfra_weights_f2$Loneliness$weight         * EMFRA_item11_trait        +   #
↪  LON  - Loneliness
      emfra_weights_f2$Sadness$weight            * EMFRA_item12_trait        +   #
↪  SAD  - Sadness
      emfra_weights_f2$`Life satisf.`$weight     * EMFRA_item15_trait_reverse +  #
↪  LS   - Life satisf. (negative → reverse)
      emfra_weights_f2$`Socio-Ludic act.`$weight * EMFRA_item19_trait_reverse    #
↪  SLA  - Socio-Ludic act. (negative → reverse)
  )

factor_vars <- c(
  "Factor_1_Autonomy",
  "Factor_2_Psych_Distress"
)

factor_labels <- c(
  "Factor_1_Autonomy"       = "Factor 1: Autonomy",
```

```
  "Factor_2_Psych_Distress" = "Factor 2: Psychological Distress"
)

factor_range_max <- c(
  "Factor 1: Autonomy"               = fa_minmax_rmc$`Maximum possible
   ↪  score`[fa_minmax_rmc$Factor == "MR1"],
  "Factor 2: Psychological Distress" = fa_minmax_rmc$`Maximum possible
   ↪  score`[fa_minmax_rmc$Factor == "MR2"]
)

fmt_pct_range <- function(x, max_score, digits = 2) {
  pct <- round(100 * x / max_score, 1)
  paste0(round(x, digits), " (", pct, "%)")
}

factor_descriptives_table <- format_continuous_table(
  get_continuous_stats(data_emfra, factor_vars),
  labels = factor_labels,
  digits = 2
) %>%
  mutate(
    .max   = factor_range_max[Variable],
    Min    = fmt_pct_range(Min,    .max),
    Q1     = fmt_pct_range(Q1,     .max),
    Median = fmt_pct_range(Median, .max),
    Q3     = fmt_pct_range(Q3,     .max),
    Max    = fmt_pct_range(Max,    .max)
  ) %>%
  select(-.max) %>%
  rename(
    `Min (% of max scale value)`    = Min,
    `Q1 (% of max scale value)`     = Q1,
    `Median (% of max scale value)` = Median,
    `Q3 (% of max scale value)`     = Q3,
    `Max (% of max scale value)`    = Max
  )
```

Table 66: Descriptive statistics of Reduced Model 5 Factors

| Variable | Mean ± SD | Min (% of max scale value) | Q1 (% of max scale value) | Median (% of max scale value) | Q3 (% of max scale value) | Max (% of max scale value) | Skewness | Kurtosis |
|---|---|---|---|---|---|---|---|---|
| Factor 1: Autonomy | 1.75 ± 0.62 | 0.36 (13.4%) | 1.2 (44.5%) | 1.86 (69%) | 2.33 (86.4%) | 2.69 (99.8%) | -0.32 | -1.13 |
| Factor 2: Psychological Distress | 0.97 ± 0.57 | 0.09 (3.3%) | 0.52 (19%) | 0.93 (34%) | 1.32 (48.3%) | 2.69 (98.5%) | 0.59 | 0.43 |

```r
factor_plot_order <- c(
  "Factor_1_Autonomy",
  "Factor_2_Psych_Distress"
)

factor_plot_labels <- c(
  "Factor_1_Autonomy"       = "Factor 1: Autonomy (Score)",
  "Factor_2_Psych_Distress" = "Factor 2: Psychological Distress (Score)"
)

color_map_factor_desc <- list(
  Factor_1_Autonomy = list(
    fill_density  = "orangered",
    color_density = "orangered",
    fill_hist     = "sienna"
  ),
  Factor_2_Psych_Distress = list(
    fill_density  = "steelblue1",
    color_density = "steelblue1",
    fill_hist     = "slateblue"
  )
)

plots_factors <- lapply(factor_plot_order, function(v) {
  cols <- color_map_factor_desc[[v]]
  plot_distribution(
    data                = data_emfra,
    var_raw             = v,
    var_label           = unname(factor_plot_labels[v]),
    fill_density        = cols$fill_density,
    color_density       = cols$color_density,
    fill_hist           = cols$fill_hist,
    density_top_shrink = 0.9,
    binwidth            = 0.08,
    xmin                = 0,
    xmax                = 3.0,
    ymax                = 25,
    show_percentiles    = FALSE
  )
})

gridExtra::grid.arrange(grobs = plots_factors, ncol = 2)
```

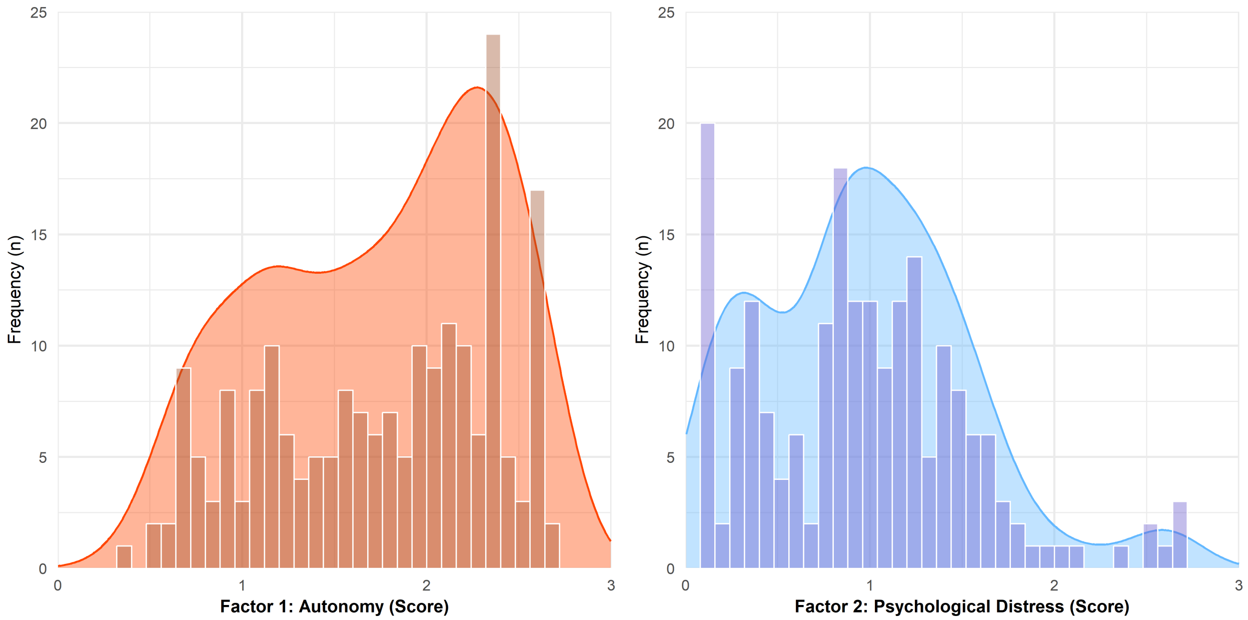

Figure 28: Distribution of EMFRA factor scores

### 7 Factor-level test-retest reliability, SEM, and MDC.

```
compute_reliability <- function(data, pairs, labels = NULL, max_scores = NULL) {

  compute_icc_sem_mdc <- function(test_col, retest_col, data) {
    data_pair <- data[, c(test_col, retest_col)] %>%
      as.data.frame()
    data_pair[] <- lapply(data_pair, as.numeric)
    data_pair   <- data_pair[complete.cases(data_pair), ]
    N  <- nrow(data_pair)
    df <- N - 1

    if (N < 2) {
      return(data.frame(
        Variable = test_col, N = N,
        ICC = NA_real_, CI_low = NA_real_, CI_high = NA_real_,
        p_value = NA_real_, SEM = NA_real_, MDC95 = NA_real_,
        stringsAsFactors = FALSE
      ))
    }

    icc_res  <- psych::ICC(data_pair, lmer = FALSE)
    icc_row  <- icc_res$results["Single_random_raters", , drop = FALSE]
    icc_val  <- as.numeric(icc_row[1, "ICC"])
```

```r
  SD_pooled <- sqrt((sd(data_pair[[test_col]],   na.rm = TRUE)^2 +
                      sd(data_pair[[retest_col]], na.rm = TRUE)^2) / 2)
  SEM   <- SD_pooled * sqrt(1 - icc_val)
  MDC95 <- SEM * sqrt(2) * qt(0.975, df = df)

  data.frame(
    Variable = test_col,
    N        = N,
    ICC      = icc_val,
    CI_low   = as.numeric(icc_row[1, "lower bound"]),
    CI_high  = as.numeric(icc_row[1, "upper bound"]),
    p_value  = as.numeric(icc_row[1, "p"]),
    SEM      = SEM,
    MDC95    = MDC95,
    stringsAsFactors = FALSE
  )
}

fmt_pct_scale <- function(x, max_score) {
  pct <- round(100 * x / max_score, 1)
  paste0(round(x, 3), " (", pct, "%)")
}

result <- purrr::map_dfr(pairs, function(p) {
  compute_icc_sem_mdc(p[1], p[2], data)
}) %>%
  mutate(
    Variable    = if (!is.null(labels)) dplyr::recode(Variable, !!!labels) else
     ↪  Variable,
    `ICC (2,1)` = round(ICC, 3),
    `95% CI`    = paste0("(", round(CI_low, 3), ", ", round(CI_high, 3), ")"),
    `p-value`   = format.pval(p_value, digits = 3, eps = .001)
  )

if (!is.null(max_scores)) {
  result <- result %>%
    mutate(
      .max           = max_scores[Variable],
      `SEM (agree.)` = mapply(fmt_pct_scale, SEM,   .max),
      `MDC95`        = mapply(fmt_pct_scale, MDC95, .max)
    ) %>%
    select(-.max)
} else {
  result <- result %>%
    mutate(
      `SEM (agree.)` = round(SEM,   3),
      `MDC95`        = round(MDC95, 3)
    )
```

```
  }

  result %>%
    select(Variable, N, `ICC (2,1)`, `95% CI`, `p-value`, `SEM (agree.)`, `MDC95`)
}

data_emfra <- data_emfra %>%
  mutate(
    Factor_1_Autonomy_retest =
      emfra_weights_f1$`Grip strength`$weight    * EMFRA_item1_trait_retest
↪   +
      emfra_weights_f1$`Gait speed`$weight       * EMFRA_item2_trait_retest
↪   +
      emfra_weights_f1$Fatigue$weight            *
↪   EMFRA_item5_trait_reverse_retest +
      emfra_weights_f1$`Arithmetic money`$weight * EMFRA_item6_trait_retest
↪   +
      emfra_weights_f1$`Atten-Inhib`$weight      * EMFRA_item7_trait_retest
↪   +
      emfra_weights_f1$Loneliness$weight         * EMFRA_item11_trait_retest
↪   +
      emfra_weights_f1$Sadness$weight            * EMFRA_item12_trait_retest
↪   +
      emfra_weights_f1$`Life satisf.`$weight     * EMFRA_item15_trait_retest
↪   +
      emfra_weights_f1$`Socio-Ludic act.`$weight * EMFRA_item19_trait_retest
  ) %>%
  mutate(
    Factor_2_Psych_Distress_retest =
      emfra_weights_f2$`Grip strength`$weight    *
↪   EMFRA_item1_trait_reverse_retest +
      emfra_weights_f2$`Gait speed`$weight       * EMFRA_item2_trait_retest
↪   +
      emfra_weights_f2$Fatigue$weight            * EMFRA_item5_trait_retest
↪   +
      emfra_weights_f2$`Arithmetic money`$weight *
↪   EMFRA_item6_trait_reverse_retest +
      emfra_weights_f2$`Atten-Inhib`$weight      * EMFRA_item7_trait_retest
↪   +
      emfra_weights_f2$Loneliness$weight         * EMFRA_item11_trait_retest
↪   +
      emfra_weights_f2$Sadness$weight            * EMFRA_item12_trait_retest
↪   +
      emfra_weights_f2$`Life satisf.`$weight     *
↪   EMFRA_item15_trait_reverse_retest +
      emfra_weights_f2$`Socio-Ludic act.`$weight *
↪   EMFRA_item19_trait_reverse_retest
```

```r
  )

Dataset_retest_factors <- data_emfra %>%
  filter(Assessed_retest == "Yes") %>%
  select(
    Factor_1_Autonomy,       Factor_1_Autonomy_retest,
    Factor_2_Psych_Distress, Factor_2_Psych_Distress_retest
  )

factor_reliability_labels <- c(
  "Factor_1_Autonomy"       = "Factor 1: Autonomy",
  "Factor_2_Psych_Distress" = "Factor 2: Psychological Distress"
)

factor_reliability_max <- setNames(
  fa_minmax_rmc$`Maximum possible score`,
  factor_reliability_labels
)

factor_reliability_pairs <- list(
  c("Factor_1_Autonomy",       "Factor_1_Autonomy_retest"),
  c("Factor_2_Psych_Distress", "Factor_2_Psych_Distress_retest")
)

Reliability_ICC_SEM_MDC <- compute_reliability(
  data       = Dataset_retest_factors,
  pairs      = factor_reliability_pairs,
  labels     = factor_reliability_labels,
  max_scores = factor_reliability_max
)

reliability_footnote <- paste(
  "ICC: Intraclass Correlation Coefficient (model 2,1; single measures, absolute
   ↪ agreement).",
  "95% CI: 95% confidence interval of the ICC.",
  "SEM: Standard Error of Measurement, computed as SD_pooled * sqrt(1 - ICC).",
  "MDC95: Minimal Detectable Change at 95% confidence, computed as SEM * sqrt(2) *
   ↪ t(0.975, df = N-1).",
  "Values in parentheses for SEM and MDC95 indicate the corresponding percentage",
  "of the maximum possible scale score (% of max scale value),",
  "providing an indication of measurement error and detectable change relative to
   ↪ the total score range."
)
```

Table 67: Factor Test-retest Reliability

| Variable | N | ICC (2,1) | 95% CI | p-value | SEM (agree.) | MDC95 |
|---|---|---|---|---|---|---|
| Factor 1: Autonomy | 22 | 0.82 | (0.613, 0.923) | <0.001 | 0.187 (7%) | 0.551 (20.5%) |
| Factor 2: Psychological Distress | 22 | 0.78 | (0.532, 0.901) | <0.001 | 0.297 (10.9%) | 0.872 (31.9%) |

*Note.* ICC: Intraclass Correlation Coefficient (model 2,1; single measures, absolute agreement). 95% CI: 95% confidence interval of the ICC. SEM: Standard Error of Measurement, computed as SD_pooled * sqrt(1 - ICC). MDC95: Minimal Detectable Change at 95% confidence, computed as SEM * sqrt(2) * t(0.975, df = N-1). Values in parentheses for SEM and MDC95 indicate the corresponding percentage of the maximum possible scale score (% of max scale value), providing an indication of measurement error and detectable change relative to the total score range.

### 8 Factor-level floor-ceiling effects

```
compute_floor_ceiling <- function(var, data, min_scale, max_scale,
                                  primary_items_min = NULL,
                                  primary_items_max = NULL,
                                  weights_f1 = NULL,
                                  weights_f2 = NULL) {
  x <- data[[var]]
  N <- sum(!is.na(x))

  fmt <- function(n, range_low = NULL, range_high = NULL) {
    base <- paste0(n, " (", round(100 * n / N, 1), "%)")
    if (!is.null(range_low) && !is.null(range_high) &&
        !is.na(range_low)   && !is.na(range_high)) {
      pct_low  <- round(100 * range_low  / max_scale, 1)
      pct_high <- round(100 * range_high / max_scale, 1)
      range_str <- paste0(
        " [", round(range_low, 3), "\u2013", round(range_high, 3),
        " (", pct_low, "%\u2013", pct_high, "%)]"
      )
      base <- paste0(base, range_str)
    }
    base
  }

  n_floor   <- sum(x == min_scale, na.rm = TRUE)
  n_ceiling <- sum(x == max_scale, na.rm = TRUE)

  if (!is.null(primary_items_min) && !is.null(primary_items_max)) {
```

```
  weights <- if (grepl("Factor_1", var)) weights_f1 else weights_f2

  primary_names  <- if (grepl("Factor_1", var)) primary_items_min else
↪ primary_items_max
  minority_names <- setdiff(names(weights), primary_names)

  primary_weights  <- weights[primary_names]
  minority_weights <- weights[minority_names]

  primary_weights  <- primary_weights[!is.na(primary_weights)]
  minority_weights <- minority_weights[!is.na(minority_weights)]

  primary_floor_low    <- 0
  primary_floor_high   <- sum(2 * abs(minority_weights))
  primary_ceiling_low  <- sum(2 * abs(primary_weights))
  primary_ceiling_high <- max_scale

  primary_data <- data %>% select(all_of(primary_names))

  n_primary_floor <- sum(
    apply(primary_data, 1, function(r) all(r == 0, na.rm = TRUE)),
    na.rm = TRUE
  )
  n_primary_ceiling <- sum(
    apply(primary_data, 1, function(r) all(r == 2, na.rm = TRUE)),
    na.rm = TRUE
  )
}

range_10pct      <- 0.10 * max_scale
near_floor_low   <- min_scale
near_floor_high  <- min_scale + range_10pct
near_ceil_low    <- max_scale - range_10pct
near_ceil_high   <- max_scale

n_near_floor   <- sum(x <= near_floor_high, na.rm = TRUE)
n_near_ceiling <- sum(x >= near_ceil_low,   na.rm = TRUE)

data.frame(
  Factor                                 = var,
  N                                      = N,
  `Minimum observed`                     = round(min(x, na.rm = TRUE), 3),
  `Maximum observed`                     = round(max(x, na.rm = TRUE), 3),
  `Floor - exact (n, %)`                 = fmt(n_floor),
  `Floor - primary items with 0 points (n, %)`    = fmt(n_primary_floor,
                                          primary_floor_low,
                                          primary_floor_high),
```

```
    `Floor - within 10% range (n, %)`   = fmt(n_near_floor,
                                                near_floor_low,
                                                near_floor_high),
    `Ceiling - exact (n, %)`            = fmt(n_ceiling),
    `Ceiling - primary items with 2 points (n, %)`  = fmt(n_primary_ceiling,
                                                primary_ceiling_low,
                                                primary_ceiling_high),
    `Ceiling - within 10% range (n, %)` = fmt(n_near_ceiling,
                                                near_ceil_low,
                                                near_ceil_high),
    check.names = FALSE
  )
}

weights_f1 <- c(
  EMFRA_item1_trait         = emfra_weights_f1$`Grip strength`$weight,
  EMFRA_item2_trait         = emfra_weights_f1$`Gait speed`$weight,
  EMFRA_item5_trait_reverse = emfra_weights_f1$Fatigue$weight,
  EMFRA_item6_trait         = emfra_weights_f1$`Arithmetic money`$weight,
  EMFRA_item7_trait         = emfra_weights_f1$`Atten-Inhib`$weight,
  EMFRA_item11_trait        = emfra_weights_f1$Loneliness$weight,
  EMFRA_item12_trait        = emfra_weights_f1$Sadness$weight,
  EMFRA_item15_trait        = emfra_weights_f1$`Life satisf.`$weight,
  EMFRA_item19_trait        = emfra_weights_f1$`Socio-Ludic act.`$weight
)

weights_f2 <- c(
  EMFRA_item1_trait_reverse  = emfra_weights_f2$`Grip strength`$weight,
  EMFRA_item2_trait          = emfra_weights_f2$`Gait speed`$weight,
  EMFRA_item5_trait          = emfra_weights_f2$Fatigue$weight,
  EMFRA_item6_trait_reverse  = emfra_weights_f2$`Arithmetic money`$weight,
  EMFRA_item7_trait          = emfra_weights_f2$`Atten-Inhib`$weight,
  EMFRA_item11_trait         = emfra_weights_f2$Loneliness$weight,
  EMFRA_item12_trait         = emfra_weights_f2$Sadness$weight,
  EMFRA_item15_trait_reverse = emfra_weights_f2$`Life satisf.`$weight,
  EMFRA_item19_trait_reverse = emfra_weights_f2$`Socio-Ludic act.`$weight
)

primary_items_F1 <- c(
  "EMFRA_item1_trait",    # HGS
  "EMFRA_item2_trait",    # GS
  "EMFRA_item6_trait",    # AM
  "EMFRA_item7_trait",    # AttIn
  "EMFRA_item19_trait"    # SLA
)

primary_items_F2 <- c(
  "EMFRA_item5_trait",         # FAT
```

```r
  "EMFRA_item11_trait",         # LON
  "EMFRA_item12_trait",         # SAD
  "EMFRA_item15_trait_reverse" # LS
)

Floor_Ceiling_EMFRA <- dplyr::bind_rows(
  compute_floor_ceiling(
    var               = "Factor_1_Autonomy",
    data              = data_emfra,
    min_scale         = 0,
    max_scale         = fa_minmax_rmc$`Maximum possible
     ↪  score`[fa_minmax_rmc$Factor == "MR1"],
    primary_items_min = primary_items_F1,
    primary_items_max = primary_items_F1,
    weights_f1        = weights_f1,
    weights_f2        = weights_f2
  ),
  compute_floor_ceiling(
    var               = "Factor_2_Psych_Distress",
    data              = data_emfra,
    min_scale         = 0,
    max_scale         = fa_minmax_rmc$`Maximum possible
     ↪  score`[fa_minmax_rmc$Factor == "MR2"],
    primary_items_min = primary_items_F2,
    primary_items_max = primary_items_F2,
    weights_f1        = weights_f1,
    weights_f2        = weights_f2
  )
) %>%
  dplyr::mutate(
    Factor = dplyr::recode(
      Factor,
      "Factor_1_Autonomy"       = "Factor 1: Autonomy",
      "Factor_2_Psych_Distress" = "Factor 2: Psychological Distress"
    )
  ) %>%
  dplyr::select(
    Factor,
    N,
    `Minimum observed`,
    `Maximum observed`,
    `Floor - exact (n, %)`,
    `Ceiling - exact (n, %)`,
    `Floor - primary items with 0 points (n, %)`,
    `Ceiling - primary items with 2 points (n, %)`,
    `Floor - within 10% range (n, %)`,
    `Ceiling - within 10% range (n, %)`
  )
```

```
floor_ceiling_footnote <- paste(
  "Values indicate the number of subjects (N) and the percentaje of the total
   ↪ analysed (%).",
  "Values in square brackets indicate the absolute score range [lower\u2013upper]
   ↪ covered by each criterion,",
  "followed by the corresponding percentage range relative to the maximum possible
   ↪ scale score (% of max scale value).",
  "Floor and ceiling effects are evaluated under three independent and
   ↪ non-mutually exclusive criteria:",
  "(1) Exact: subjects whose observed factor score equals exactly the minimum (0)
   ↪ or maximum possible scale value.",
  "(2) Primary items: subjects scoring 0 (floor) or 2 (ceiling) on all items with
   ↪ the highest factor loadings",
  "(items 1, 2, 6, 7, and 19 for Autonomy; items 5, 11, 12, and 15 for
   ↪ Psychological Distress).",
  "This criterion operates at the item level, not on the factor score directly.",
  "The score range shown reflects the minimum and maximum factor score achievable
   ↪ under each response pattern:",
  "for the floor, primary items score 0 while non-primary items may range from 0
   ↪ to 2;",
  "for the ceiling, primary items score 2 while non-primary items may range from 0
   ↪ to 2.",
  "(3) Within 10% range: subjects whose observed factor score falls within the
   ↪ lowest or highest 10% of the maximum possible scale value.",
  "Unlike criterion (2), this criterion operates directly on the observed factor
   ↪ score.",
  "Because criteria (2) and (3) operate at different levels (item-level vs. factor
   ↪ score-level),",
  "they may capture different numbers of subjects even when their score ranges
   ↪ overlap."
)
```

Table 68: Floor and Ceiling effects

| Factor | N | Minimum observed | Maximum observed | Floor — exact (n, %) | Ceiling — exact (n, %) | Floor — primary items with 0 points (n, %) | Ceiling — primary items with 2 points (n, %) | Floor — within 10% range (n, %) | Ceiling — within 10% range (n, %) |
|---|---|---|---|---|---|---|---|---|---|
| Factor 1: Autonomy | 201 | 0.36 | 2.69 | 0 (0%) | 0 (0%) | 0 (0%) [0–0.25 (0%–9.3%)] | 22 (10.9%) [2.446–2.696 (90.7%–100%)] | 0 (0%) [0–0.27 (0%–10%)] | 23 (11.4%) [2.426–2.696 (90%–100%)] |
| Factor 2: Psychological Distress | 201 | 0.09 | 2.69 | 0 (0%) | 0 (0%) | 26 (12.9%) [0–0.308 (0%–11.3%)] | 4 (2%) [2.424–2.732 (88.7%–100%)] | 25 (12.4%) [0–0.273 (0%–10%)] | 6 (3%) [2.459–2.732 (90%–100%)] |

*Note.* Values indicate the number of subjects (N) and the percentaje of the total analysed (%). Values in square brackets indicate the absolute score range [lower–upper] covered by each criterion, followed by the corresponding percentage range relative to the maximum possible scale score (% of max scale value). Floor and ceiling effects are evaluated under three independent and non-mutually exclusive criteria: (1) Exact: subjects whose observed factor score equals exactly the minimum (0) or maximum possible scale value. (2) Primary items: subjects scoring 0 (floor) or 2 (ceiling) on all items with the highest factor loadings (items 1, 2, 6, 7, and 19 for Autonomy; items 5, 11, 12, and 15 for Psychological Distress). This criterion operates at the item level, not on the factor score directly. The score range shown reflects the minimum and maximum factor score achievable under each response pattern: for the floor, primary items score 0 while non-primary items may range from 0 to 2; for the ceiling, primary items score 2 while non-primary items may range from 0 to 2. (3) Within 10% range: subjects whose observed factor score falls within the lowest or highest 10% of the maximum possible scale value. Unlike criterion (2), this criterion operates directly on the observed factor score. Because criteria (2) and (3) operate at different levels (item-level vs. factor score-level), they may capture different numbers of subjects even when their score ranges overlap.

### 9 Concurrent validity

```r
dd_concurrent <- datadist(data_emfra)
options(datadist = "dd_concurrent")

run_orm_boot <- function(data, y, x, B = 2000, seed = 123){

  data_reg <- data %>%
    select(all_of(c(y, x))) %>%
    filter(!is.na(.data[[y]]), !is.na(.data[[x]])) %>%
    mutate(
      !!y := as.numeric(.data[[y]]),
      !!x := as.numeric(.data[[x]])
    )

  model_formula <- as.formula(paste(y, "~", x))

  fit <- orm(
    model_formula,
    data = data_reg,
    family = "logistic",
    r2 = 0,
    x = TRUE,
    y = TRUE
  )

  set.seed(seed)

  fit_boot <- bootcov(
    fit,
    B = B,
    coef.reps = TRUE
  )
```

```r
  r2 <- as.numeric(fit$stats["R2"])

  coef_val <- coef(fit)[[x]]

  boot_coefs <- fit_boot$boot.Coef[, x]

  ci_low <- quantile(boot_coefs, 0.025, na.rm = TRUE)
  ci_high <- quantile(boot_coefs, 0.975, na.rm = TRUE)

  pearson_r <- cor(
    data_reg[[y]],
    data_reg[[x]],
    method = "pearson",
    use = "complete.obs"
    )

  spearman_rho <- cor(
    data_reg[[y]],
    data_reg[[x]],
    method = "spearman",
    use = "complete.obs"
  )

  result <- data.frame(
    Outcome = y,
    Predictor = x,
    N = nrow(data_reg),
    `Nagelkerke pseudo-R2` = r2,
    `Pearson (r)` = round(pearson_r, 3),
    `Spearman rho` = round(spearman_rho, 3),
    `Regression coefficient` = round(coef_val, 3),
    `Boot 95%CI Percentile` = paste0(
      "(", round(ci_low, 3), ", ",
      round(ci_high, 3), ")"
    ),
    check.names = FALSE
  )

  return(list(
  table = result,
  model = fit_boot,
  fit = fit,
  data = data_reg
))
}

predictors_conc <- c(
  "SPPB_points",
```

```r
  "MoCA_without_studies",
  "HADS_anxiety_total",
  "HADS_depression_total",
  "HADS_total",
  "Barthel_total"
)

var_labels_conc <- c(
  Factor_1_Autonomy = "Factor 1 - Autonomy",
  Factor_2_Psych_Distress = "Factor 2 - Psychological distress",
  SPPB_points = "SPPB score",
  MoCA_without_studies = "MoCA score",
  HADS_anxiety_total = "HADS Anxiety",
  HADS_depression_total = "HADS Depression",
  HADS_total = "HADS Total",
  Barthel_total = "Barthel Index"
)
```

```r
results_factor1 <- lapply(predictors_conc, function(p){
  run_orm_boot(data_emfra, "Factor_1_Autonomy", p)
})

results_factor2 <- lapply(predictors_conc, function(p){
  run_orm_boot(data_emfra, "Factor_2_Psych_Distress", p)
})

results_factor1_table <- do.call(rbind, lapply(results_factor1, function(x)
 ↪  x$table))
results_factor2_table <- do.call(rbind, lapply(results_factor2, function(x)
 ↪  x$table))

results_orm <- rbind(results_factor1_table, results_factor2_table)
```

Table 69: Concurrent validity between EMFRA factors and clinical variables

| Outcome | Predictor | N | Nagelkerke pseudo-R2 | Pearson (r) | Spearman rho | Regression coefficient | Boot 95%CI Percentile |
|---|---|---|---|---|---|---|---|
| Factor 1 - Autonomy | SPPB score | 201 | 0.529 | 0.751 | 0.773 | 0.528 | (0.447, 0.64) |
| Factor 1 - Autonomy | MoCA score | 201 | 0.466 | 0.697 | 0.697 | 0.281 | (0.234, 0.341) |
| Factor 1 - Autonomy | HADS Anxiety | 201 | 0.017 | -0.122 | -0.077 | -0.058 | (-0.12, 0.004) |
| Factor 1 - Autonomy | HADS Depression | 201 | 0.176 | -0.414 | -0.394 | -0.240 | (-0.318, -0.171) |
| Factor 1 - Autonomy | HADS Total | 201 | 0.086 | -0.288 | -0.257 | -0.082 | (-0.124, -0.046) |
| Factor 1 - Autonomy | Barthel Index | 201 | 0.322 | 0.560 | 0.625 | 0.074 | (0.054, 0.105) |
| Factor 2 - Psychological distress | SPPB score | 201 | 0.078 | -0.273 | -0.282 | -0.145 | (-0.229, -0.066) |
| Factor 2 - Psychological distress | MoCA score | 201 | 0.074 | -0.291 | -0.263 | -0.083 | (-0.124, -0.042) |
| Factor 2 - Psychological distress | HADS Anxiety | 201 | 0.277 | 0.566 | 0.511 | 0.289 | (0.218, 0.373) |
| Factor 2 - Psychological distress | HADS Depression | 201 | 0.280 | 0.550 | 0.559 | 0.339 | (0.246, 0.459) |
| Factor 2 - Psychological distress | HADS Total | 201 | 0.353 | 0.625 | 0.590 | 0.208 | (0.16, 0.266) |
| Factor 2 - Psychological distress | Barthel Index | 201 | 0.094 | -0.306 | -0.337 | -0.033 | (-0.052, -0.017) |

```r
plot_rms_scatter <- function(model, data, x, y, x_range = NULL){

  if(is.null(x_range)){
    x_range <- range(data[[x]], na.rm = TRUE)
  }

  if(y == "Factor_1_Autonomy"){
    scatter_col <- "coral4"
    line_col <- "orangered2"
    ribbon_col <- "orangered2"
  } else if(y == "Factor_2_Psych_Distress"){
    scatter_col <- "slategray4"
    line_col <- "slateblue1"
    ribbon_col <- "slateblue1"
  } else {
    scatter_col <- "grey60"
    line_col <- "black"
    ribbon_col <- "black"
  }

  pred <- do.call(
    rms::Predict,
    c(
      list(
        model,
        fun = Mean(model),
        conf.int = 0.95
      ),
      setNames(list(seq(x_range[1], x_range[2], length = 200)), x)
    )
  )

  pred_df <- as.data.frame(pred)

  ggplot(data, aes(x = .data[[x]], y = .data[[y]])) +

    geom_point(
      color = scatter_col,
      alpha = 0.4
    ) +

    geom_ribbon(
      data = pred_df,
      aes(
        x = .data[[x]],
        ymin = lower,
        ymax = upper
      ),
```

```r
      fill = ribbon_col,
      alpha = 0.2,
      inherit.aes = FALSE
    ) +

    geom_line(
      data = pred_df,
      aes(
        x = .data[[x]],
        y = yhat
      ),
      color = line_col,
      linewidth = 1,
      inherit.aes = FALSE
    ) +

    theme_minimal()
}

plot_rms_grid <- function(results_list, predictors_conc, outcome, ncol = 3){
  library(patchwork)

  plots <- lapply(seq_along(results_list), function(i){
    plot_rms_scatter(
      model = results_list[[i]]$fit,
      data  = results_list[[i]]$data,
      x     = predictors_conc[i],
      y     = outcome
    ) +
      labs(
        x = var_labels_conc[predictors_conc[i]],
        y = var_labels_conc[outcome]
      )
  })

  wrap_plots(plots, ncol = ncol)
}
```

```r
plot_rms_grid(
  results_factor1,
  predictors_conc,
  "Factor_1_Autonomy",
  ncol = 3
)
```

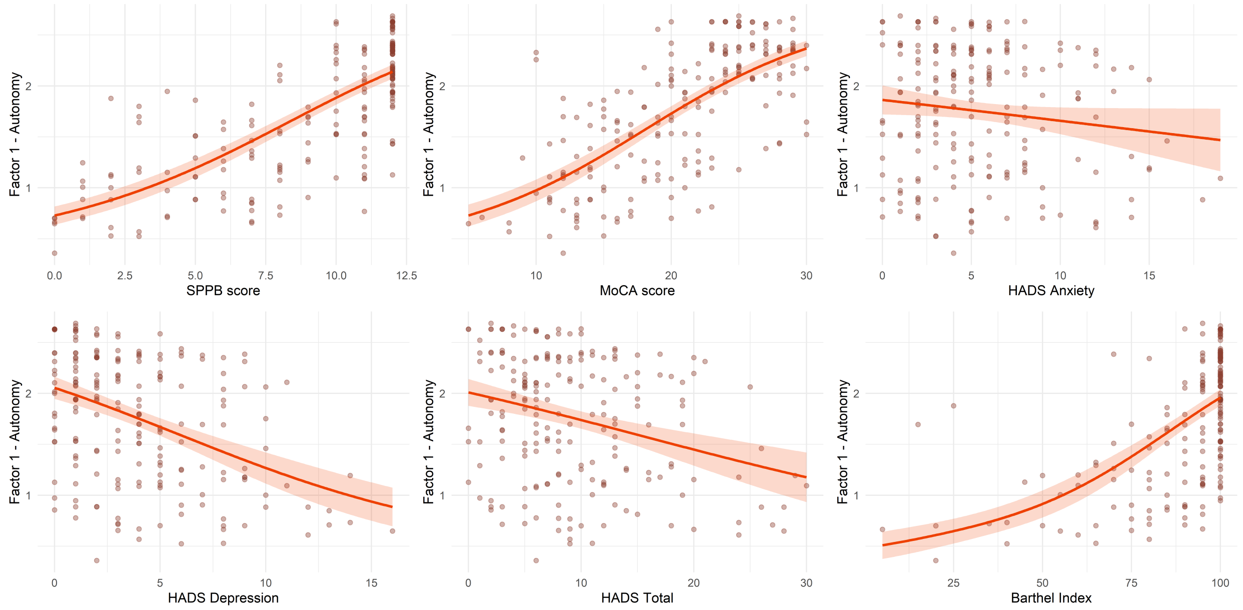

Figure 29: Concurrent validity between clinical outcomes and Autonomy factor

```
plot_rms_grid(
  results_factor2,
  predictors_conc,
  "Factor_2_Psych_Distress",
  ncol = 3
)
```

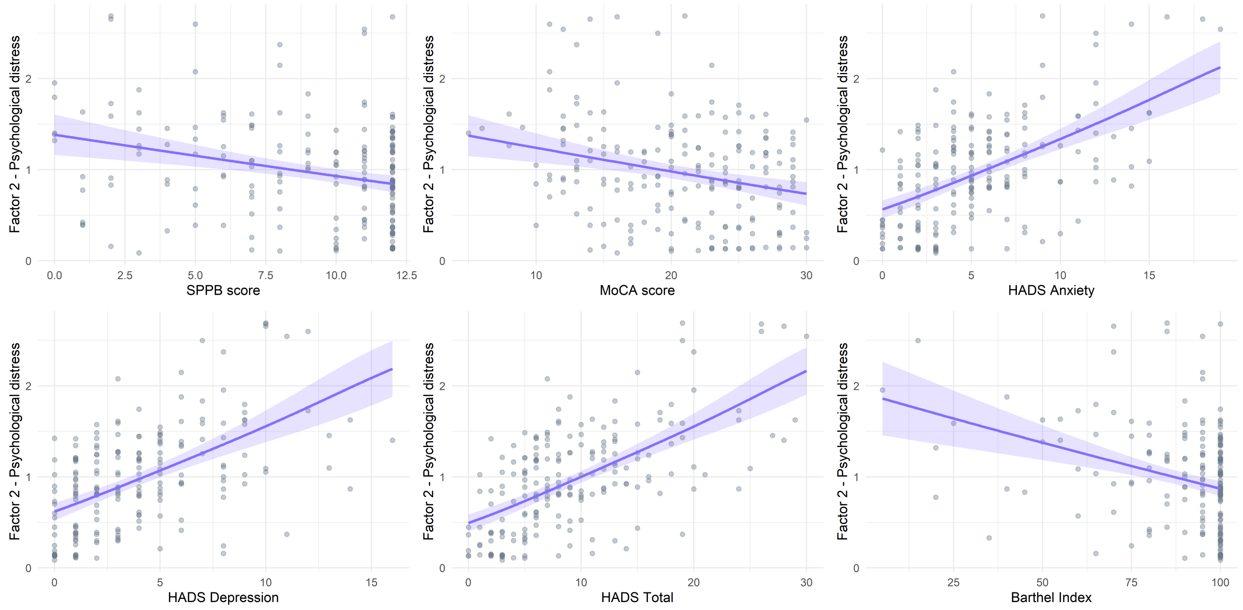

Figure 30: Concurrent validity between clinical outcomes and Psychological Distress factor
